# Genome-wide association study of body fat distribution traits in Hispanics/Latinos from the HCHS/SOL Study

**DOI:** 10.1101/2021.02.23.21251958

**Authors:** Anne E. Justice, Kristin Young, Stephanie M. Gogarten, Tamar Sofer, Misa Graff, Shelly Ann Love, Yujie Wang, Yann C. Klimentidis, Miguel Cruz, Xiuqing Guo, Fernando Hartwig, Lauren Petty, Jie Yao, Matthew A. Allison, Jennifer E. Below, Thomas A. Buchanan, Yii-Der Ida Chen, Mark O. Goodarzi, Craig Hanis, Heather M. Highland, Willa A. Hsueh, Eli Ipp, Esteban Parra, Walter Palmas, Leslie J. Raffel, Jerome I. Rotter, Jingyi Tan, Kent D. Taylor, Adan Valladares, Anny H. Xiang, Lisa Sánchez-Johnsen, Carmen R. Isasi, Kari E. North

## Abstract

Central obesity is a leading health concern with a great burden carried by ethnic minority populations, and especially Hispanics/Latinos. Genetic factors contribute to the obesity burden overall and to inter-population differences. We aim to: 1) identify novel loci associated with central adiposity measured as waist-to-hip ratio (WHR), waist circumference (WC), and hip circumference (HIP), all adjusted for body mass index (adjBMI), using the Hispanic Community Health Study/Study of Latinos (HCHS/SOL); 2) determine if differences in genetic associations differ by background group within HCHS/SOL; 3) determine whether previously reported association regions generalize to HCHS/SOL. Our analyses included 7,472 women and 5,200 men of mainland (Mexican, Central and South American) and Caribbean (Puerto Rican, Cuban, and Dominican) background residing in the US, with genome-wide array data imputed to the 1000 genomes Phase I multiethnic reference panel. We analyzed associations stratified by sex in addition to sexes combined using linear mixed-model regression. We identified 16 variants for WHRadjBMI, 22 for WCadjBMI, and 28 for HIPadjBMI that reached suggestive significance (P<1×10^−6^). Many of the loci exhibited differences in strength of associations by ethnic background and sex. We brought a total of 66 variants forward for validation in nine cohort studies (N=34,161) with participants of Hispanic/Latino, African and European descent. We confirmed four novel loci (ancestry-specific P<0.05 in replication, consistent direction of effect with HCHS/SOL, and P<5×10^−8^ after meta-analysis with HCHS/SOL), including rs13301996 in the sexes-combined analysis, and rs79478137 for women-only for WHRadjBMI; rs28692724 in women-only for HIPadjBMI; and rs3168072 in the sexes combined analysis for WCadjBMI. Also, a total of eight previously reported WHRadjBMI association regions, 12 for HIPadjBMI, and 10 for WCadjBMI generalized to HCHS/SOL. Our study findings highlight the importance of large-scale genomic studies in ancestrally diverse Hispanic/Latino populations for identifying and characterizing central obesity-susceptibility that may be ancestry-specific.

## Introduction

Obesity, and especially central obesity, is a leading risk factor for metabolic and cardiovascular diseases, with the greatest burden carried by minority populations (1–4), particularly Hispanic/Latino Americans and African Americans (5). Emerging evidence suggests that genetic factors may contribute not only to the obesity burden overall, explaining 40% to 70% of the inter-individual variation (6), but also to population-specific differences in obesity susceptibility (7–12). For example, although a majority of the >1000 genome-wide association study (GWAS)-identified obesity (body mass index (BMI), waist-to-hip ratio (WHR), waist circumference (WC), hip circumference (HIP), and body fat percentage) loci generalize across populations (13–20), recent studies in populations of Asian (19, 20) and African (16, 21) ancestry have revealed a number of novel and population-specific loci. These observations highlight the importance of large-scale genomic studies in ancestrally diverse populations, including Hispanic/Latinos, to identify obesity-susceptibility loci, and more specifically, alleles that are ancestry-specific and may thus partly explain disparities. However, no large-scale GWAS for any obesity-related traits has been performed in Hispanic/Latino populations, despite their increased prevalence of obesity.

While obesity is commonly assessed by BMI, measures of central adiposity such as WHR and WC are predictors of increased cardiometabolic risk independent of BMI (22–25). Here we consider three measures of central obesity: WHR, WC, and HIP after accounting for overall body size, measured as body mass index (BMI) (WHRadjBMI, WCadjBMI, HIPadjBMI). Larger WHR indicates increased abdominal fat and is associated with increased risk for type 2 diabetes (T2D) and cardiovascular disease (CVD) (26–28), while smaller WHR indicates a proportionately greater fat accumulation around the hips, and is associated with lower risk for T2D, hypertension and dyslipidemia (29). Previous GWAS have identified WHR, WC, and HIP loci that are enriched for association with other cardiometabolic traits and suggested that different fat distribution patterns can have distinct genetic underpinnings (30–32). Identifying genetic risk variants across these traits in Hispanic/Latinos may provide insights into these mechanisms and highlight population-specific variants that increase susceptibility to obesity in specific groups.

We aimed to: 1) identify novel genetic loci associated with central obesity, measured here as WHRadjBMI, WCadjBMI, and HIPadjBMI, in Hispanics/Latinos; 2) determine if differences in genetic associations by background group (mainland or Caribbean) and sex exist in HCHS/SOL; and 3) assess generalization of central adiposity associated loci discovered in European, African and multi-ethnic studies to Hispanics/Latinos.

## Results

### Discovery

We identified 16 loci for WHRadjBMI, 22 for WCadjBMI, and 28 for HIPadjBMI that exhibited suggestive evidence of association in the HCHS/SOL in at least one stratum (**Table 1, Figures 1–3, Supplementary Tables 2-4, Supplementary Figures 1-12**). For WHRadjBMI, we identified four loci that reach suggestive significance (*P*<1×10^−6^) in the combined sexes, including rs12435790 near *KIAA0391*, which is within a previously reported WHRadjBMI locus (+/ࢤ 500 Kb from tag SNP) (33). We also identified five loci for men only, including one reaching genome-wide significance (GWS, *P*<5×10^−8^). A total of eight suggestive loci were identified in the women-only analyses, including one, rs115981023 in *TAOK3*, which also reached suggestive significance in the combined sexes analysis, and rs79478137 in *SLC22A18AS* near a previously implicated WHRadjBMI locus(34). For WCadjBMI, we identified nine loci, including one GWS locus in the combined sexes; 11 for men only, including two SNPs that reach GWS; and two for women only. Of the WCadjBMI loci identified, two were nearby previously-reported WCadjBMI loci, rs6809759 near *PROK2* (men-only) (15, 17, 35) and rs77319470 near *ADAMTS3* (sexes-combined) (15, 17, 36). For HIPadjBMI, we identified eight loci that reach p<1×10^−6^ for the combined sexes; nine for men only, including one in a locus that reached suggestive significance for the combined sexes as well (near *ANO10*); and 12 for women only, including one SNP in a locus that reached suggestive significance for the combined sexes as well (near *LPPR4*). Of the WCadjBMI loci, rs10818474 near *MEGF9* was within 500 Kb of a recently reported WHRadjBMI association in women (35).

**Table 1.**
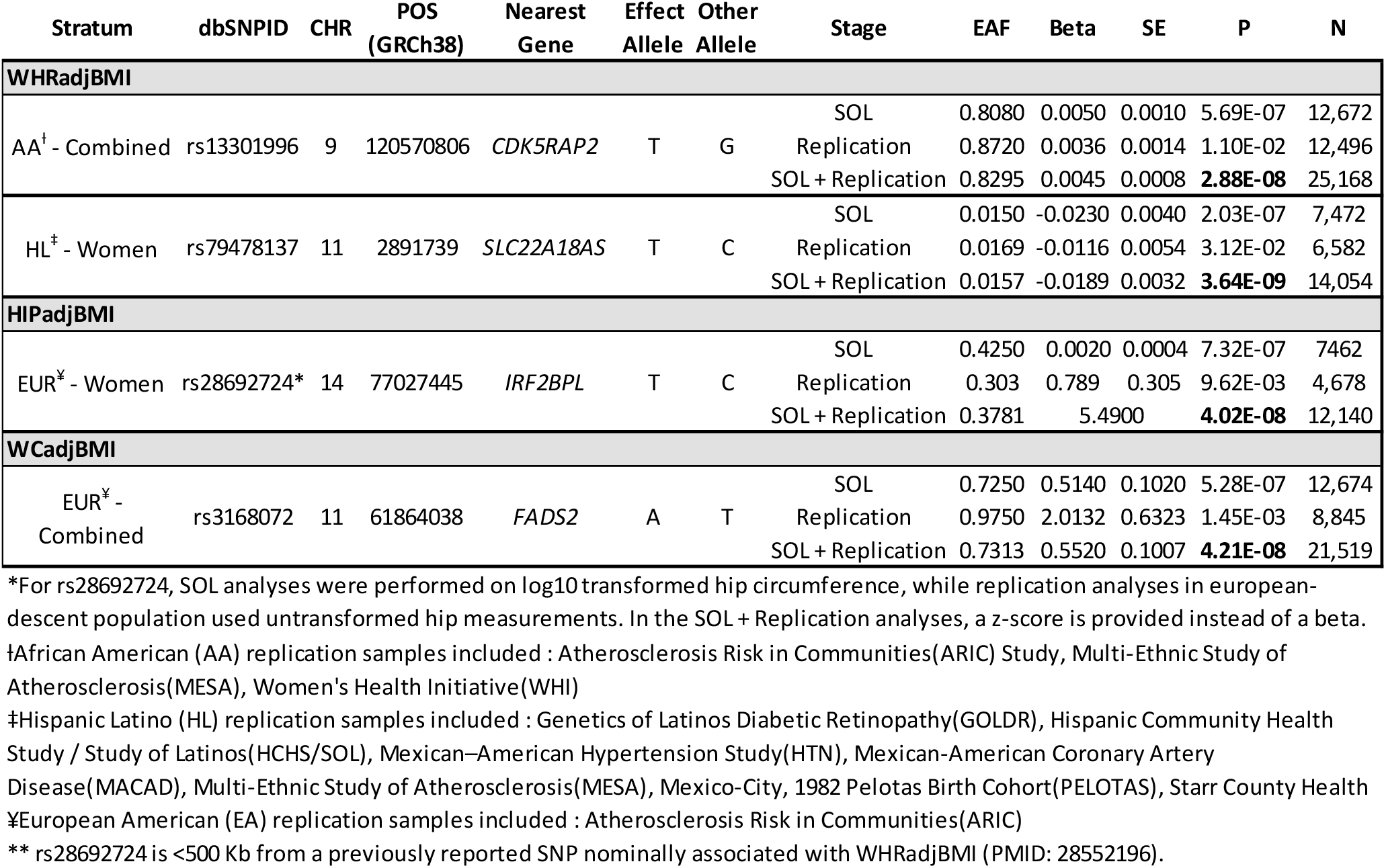
Summary of association results for all loci that passed replication criteria. EAF-estimated allele frequency, chr-chromosome, pos-position, SE-standard error, ISQ-I squared heterogeneity.

**Figure 1.**
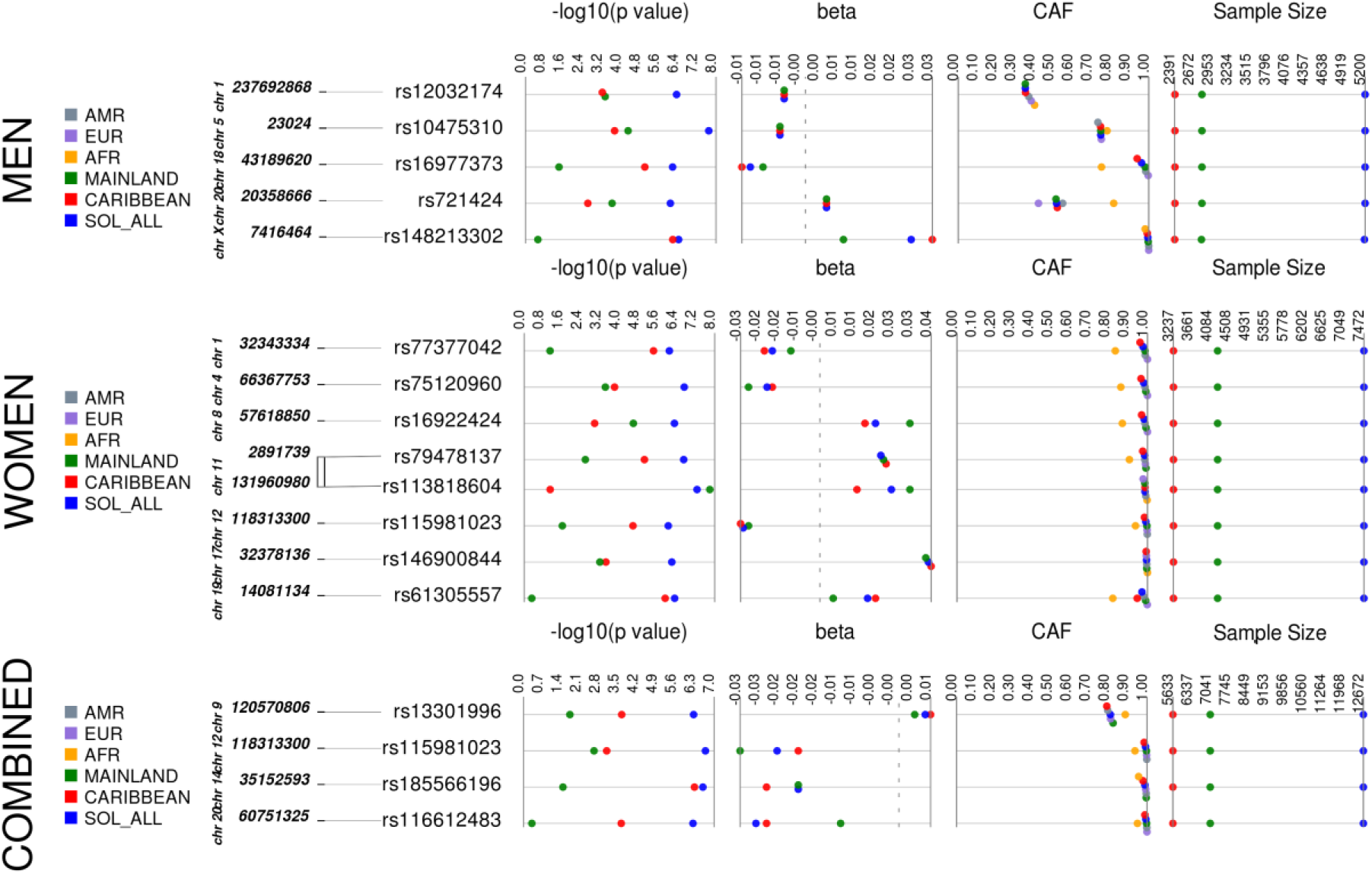
WCadjBMI Synthesis View plot that shows −log10 *P*-values, beta (effect estimate), effect/coded allele frequency (CAF), and sample size across analysis samples for all loci that reached suggestive significance in one or more of our discovery strata. This chart also shows the coded allele frequency (CAF) of each of our top loci by background group and by 1000 genomes reference panel (European - EUR, Latin American - AMR, and African - YRI).

**Figure 2.**
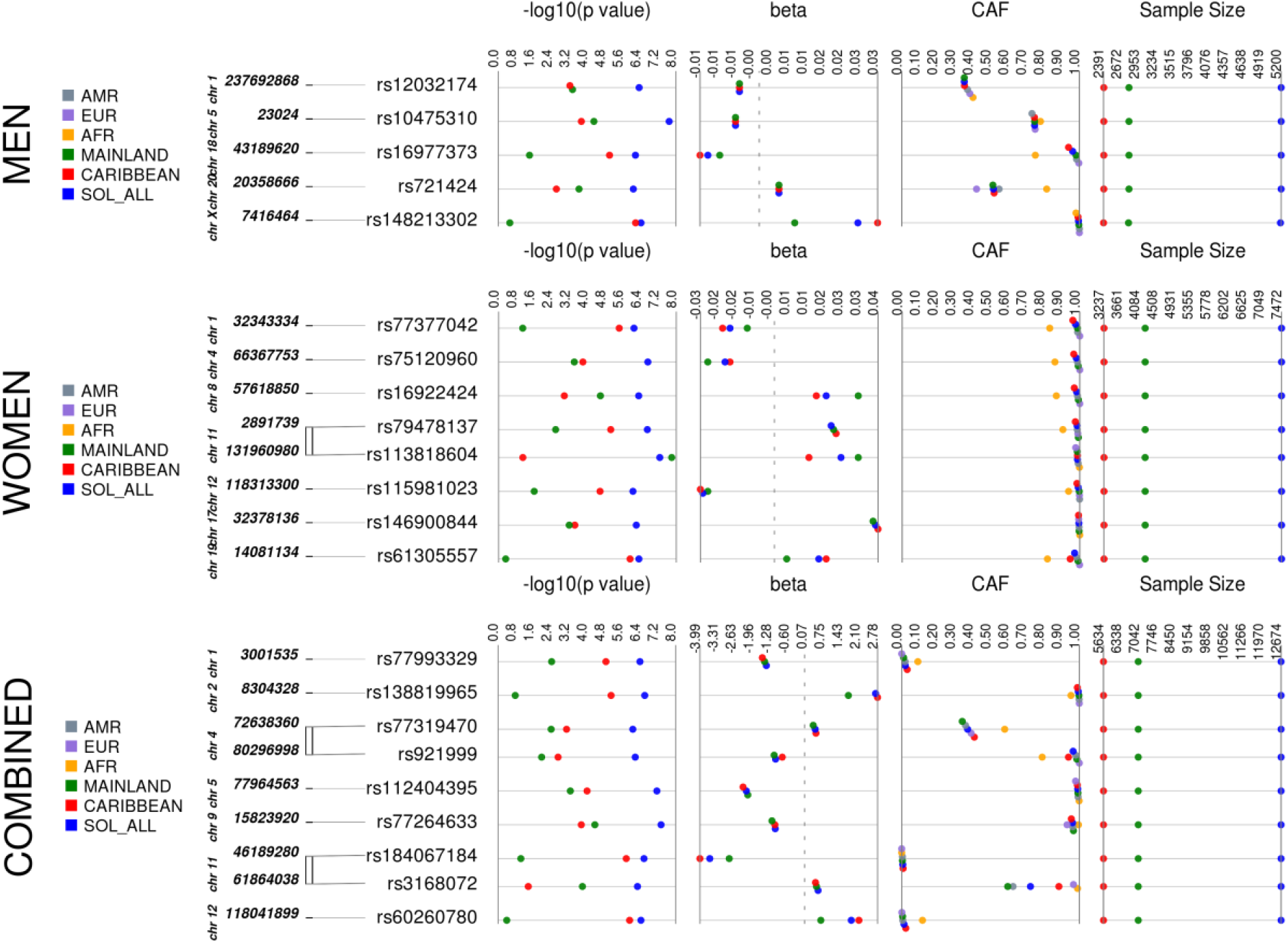
WHRadjBMI Synthesis View plot that shows −log10 *P*-values, beta (effect estimate), effect/coded allele frequency (CAF), and sample size across analysis samples for all loci that reached suggestive significance in one or more of our discovery strata. This chart also shows the coded allele frequency (CAF) of each of our top loci by background group and by 1000 genomes reference panel (European - EUR, Latin American - AMR, and African - YRI).

**Figure 3.**
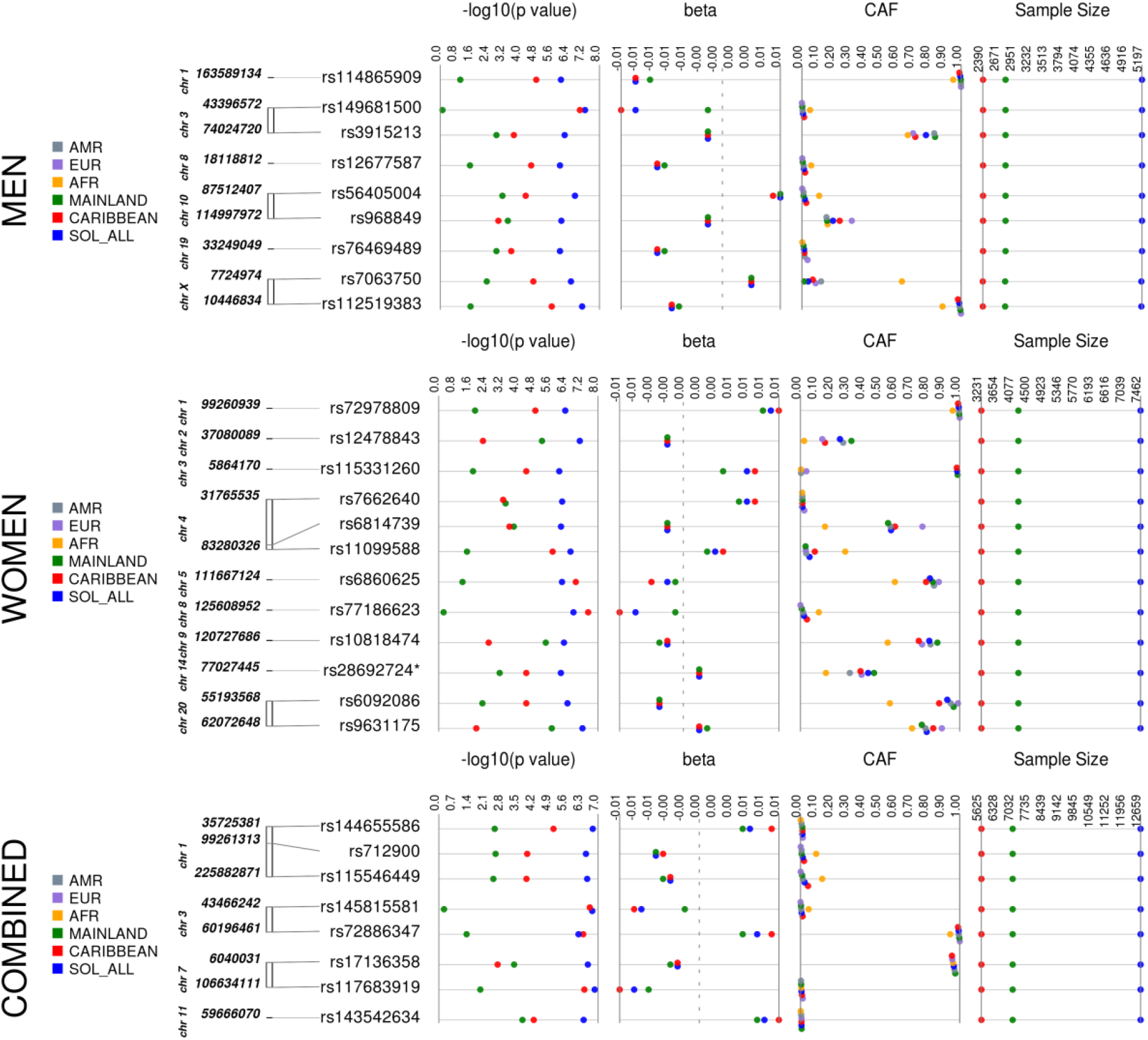
HIPadjBMI Synthesis View plot that shows the −log10 *P*-values, beta (effect estimate), effect/coded allele frequency (CAF), and sample size across analysis samples for all loci that reached suggestive significance in one or more of our discovery strata. This chart also shows the coded allele frequency (CAF) of each of our top loci by background group and by 1000 genomes reference panel (European - EUR, Latin American - AMR, and African - YRI).

### Association Differences by Genetic Ancestry

All of the top loci were directionally consistent in each background group, yet many of the loci exhibited effect heterogeneity by background group (**Table 2, Figures 1–3, Supplementary Tables 5-7**), as exhibited by moderate to high I^2^ values (ISQ>65%) and/or significant interaction across subgroups (P_diff_<0.05). For example, rs113818604 (β=0.0269, *P*= 5.47 x 10^−8^), I^2^=78.5%, P_diff_=0.38) in *NTM* is significantly associated with WHRadjBMI in women from the mainland background groups (N=4220, MAF=0.014, β=0.0343, *P*=1.63×10^−8^), but not in women from Caribbean background groups (N=3238, MAF=0.013, β=0.0144, *P*=0.08) **(Supplementary Table 5**). Also, for the women-only primary analysis, the rs77186623 in *LOC105375745* locus associated with HIPadjBMI (β=-0.006, P=1.74×10^−7^) exhibited nominally significant interaction by background group (I^2^=55.3%, P_diff_= 0.042), and was GWS in the Caribbean group (N=3231, MAF=0.041, β=-0.0078, *P*=3.05×10^−8^), but not significant in the mainland group (N=4216, MAF=0.008, β=-0.0015, *P*=0.567, **Supplementary Table 7**). Additional examples that cannot be explained due to power (i.e. MAF and sample size are similar) for WHRadjBMI include rs77377042 near *MARCKSL1* and rs61305557 in *C19orf67* for women, and rs16977373 near *RIT2* for men; for WCadjBMI in women-only include rs76842062 in *MAP4K4* and rs76941364 near *COBL;* and for HIPadjBMI rs6860625 near *NREP* for women and rs145815581 in *ANO10* for the combined sexes.

**Table 2.**
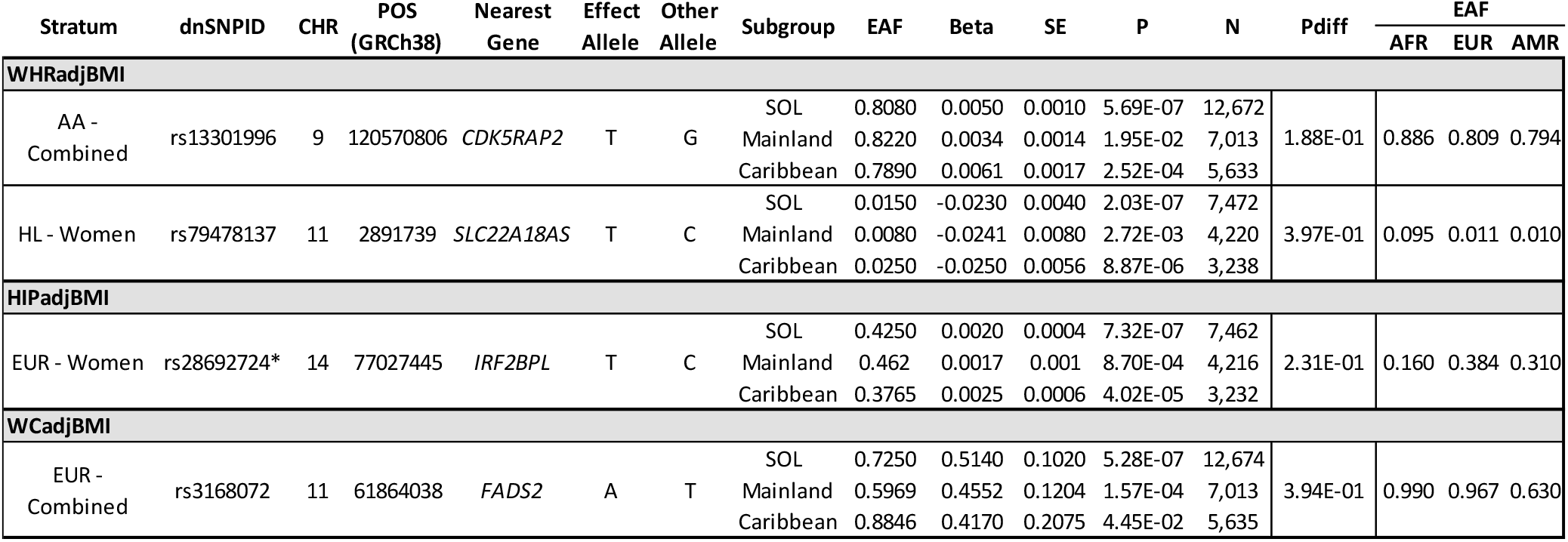
Summary of top association results in SOL subgroup analyses. EAF-effect allele frequency, CHR-chromosome, POS-position, SE- standard error, ISQ-I squared heterogeneity. *EAF for reference population obtained from ExAC; all other estimated are from 1000 Genomes Project Phase 3.

For other loci, allele frequency and linkage disequilibrium (LD) differences across Hispanic/Latino populations likely contributed to observed differences in magnitude of effect and significance levels (**Supplementary Table 8**). For example, while the magnitude of effect for the rs115981023 *TAOK3* association with WHRadjBMI in women (β=-0.029, P=8.88×10^−7^, I^2^=0, P_diff_=0.391) was similar across background groups, the P-value was far more significant in the Caribbean background group (MAF=0.016, β=-0.030, P=2.72×10^−5^) compared to the mainland (MAF=0.003, β=-0.027, P=0.025), likely due to the higher MAF in the Caribbean group. Of note, the minor allele at this SNP is more common in the 1000 Genomes AFR compared to the EUR and AMR reference samples (**Supplementary Table 5**) and the local ancestry of participants at this locus indicate that those with African ancestry exhibit the highest MAF (**Supplementary Table 8**). Additional loci where the significance level differences between Caribbean and mainland background groups appear to be driven by increased MAF due to African ancestry in Caribbean populations include the *SLC22A18AS* and *CDH4* loci for WHRadjBMI; *LOC102723448, FZD7, WSB2*, and *ACTRT2* loci for WCadjBMI; and *COQ2, LPPR4, TMEM63A*, and *FHIT* loci for HIPadjBMI (**Supplementary Tables 5-8**). Rs12478843 in *HEATR5B* (β=-0.002, P=8.2×10^−8^, I^2^=1.7%, P_diff_=0.385) is more significantly associated with HIPadjBMI in mainland (MAF=0.320, β=-0.002, *P*=6.50×10^−6^) women compared to Caribbean (MAF=0.154, β=-0.002, *P*=6.03×10^−3^), likely reflecting the higher MAF among those from mainland Latin America with greater Native American ancestry (**Supplementary Table 8**). Similarly, differences in effect magnitude between mainland and Caribbean background groups for the *TAF4* (HIPadjBMI in women) and the *ESRRG* (WCadjBMI in men) loci may also be due to higher MAF in the mainland group because of a greater proportion of Native American ancestry (**Supplementary Tables 6-8**).

### Replication

We brought 66 variants forward for replication in nine cohort studies (N up to 34,161) with participants of Hispanic/Latino, African and European descent, and for further examination of replication by ancestral background (**Table 1, Supplementary Tables 1-4, Supplementary Figures 10-12**). Our criteria for replication included both nominal evidence of an association (*P*<0.05), consistent direction of effect between the replication results and the HCHS/SOL discovery results for any ancestry/sex stratum, and genome-wide significance (*P*<5×10^−8^) when meta-analyzed together with HCHS/SOL. Based on these criteria, we were able to replicate four novel loci (**Table 1**) after combining our HCHS/SOL discovery sample with specific ancestry results. For WHRadjBMI in men and women combined, rs13301996 was significant after meta-analyzing HCHS/SOL with the African American replication sample (*P*=2.88×10^−8^). For WHRadjBMI in women only, rs79478137 was GWS after combining HCHS/SOL with the Hispanic/Latino replication sample (*P*=3.64×10^−9^). For HIPadjBMI in women only, rs28692724 was significant after meta-analyzing HCHS/SOL with the European American replication sample (*P*=4.02×10^−8^). For WCadjBMI in men and women combined, rs3168072 was significant after combining HCHS/SOL with the European American replication sample (*P*=4.21×10^−8^).

Of note, for rs13301996, which only replicated in African Americans, we see a larger effect size in the Caribbean background group compared to the mainland, although this is not a significant difference (**Table 2, Supplementary Table 5 Supplementary Figure 10**). This finding may provide insight into why the variants were more successful upon replication with a particular ancestry. For the remaining loci there is little difference in effect magnitude between the Caribbean and the mainland background groups that could explain differences in replication by ancestral group.

### Generalization of Previous Loci

We examined previously reported association regions from the GIANT Consortium (35) to assess generalization to the HCHS/SOL (**Supplementary Tables 9-11, Supplementary Figures 13-21**). To account for differences in LD between GIANT (primarily European descent populations) and HCHS/SOL (highly admixed Hispanic/Latino populations), we report generalization results based on the lead generalized SNP (the SNP with lowest r-value in the region of the previously reported variant in GIANT). In sex combined analyses, there were a total of 12 association regions across the genome that generalized to HCHS/SOL for WHRadjBMI (r<0.05), including three for both women-only and sexes combined, three for women-only, and six for the sexes-combined analysis (**Supplementary Table 9**). A total of 15 association regions generalized to HCHS/SOL for WCadjBMI, including seven sex-specific loci (two for men, five for women, **Supplementary Table 10**), one for the sexes-combined only, and seven for more than one stratum. Of note, we identified rs6809759 near *PROK2*, which was significantly associated with WCadjBMI in HCHS/SOL for men-only and sexes-combined and was within 500 kb (+/−) of rs12330322, identified in Shungin et al (35). However, this previously-identified locus did not generalize to HCHS/SOL (r>0.05); and may represent an independent association signal in a known region (i.e. all GIANT variants at this locus with *P*<1×10^−6^ exhibit r>0.05 in HCHS/SOL and rs6809759 had a *P*>1×10^−6^ in Shungin et al. (35) [*P*=1.4×10^−1^]). A total of 40 regions generalized to HCHS/SOL for HIPadjBMI, including 29 for sexes-combined, three of which were significant for both women-only and sexes combined analyses (**Supplementary Table 11**).

Because some of the SNPs previously reported by GIANT may not have generalized due to lack of power in HCHS/SOL, we calculated individual-level genetic scores based on trait-increasing alleles for each central adiposity phenotype (**Supplementary Table 12**) and sex stratum (three association tests per phenotype). For genetic scores based on SNPs with p-value<1×10^−7^ in GIANT, all association tests were significant (p-value<0.05). For genetic scores calculated from GIANT SNPs with 1×10^−7^<*P*<1×10^−6^, six of the nine association tests were significant. Given that only three out of 27 analyses had a p-value>0.05, there is considerable overlap in the association results of Hispanics/Latinos to those previously reported in the GIANT multiethnic analysis.

### Biological Curation

We examined the four SNPs (i.e. rs13301996, rs79478137, rs28692724, and rs3168072) in novel loci identified in the replication analyses (**Table 1**) for association with other phenotypes, gene expression, and metabolites in publicly available data using Phenoscanner (37, 38), and assessed the potential regulatory role of these variants and those in LD using publicly available databases, including RegulomeDB (39), Haploreg (40), UCSC GenomeBrowser (41), and GTeX (42). Known associations with these variants meeting Bonferroni-corrected significance after correcting for number of reported associations in Phenoscanner for the four variants within each category (P<0.05/7631=6.55×10^−5^ for GWAS; P<0.05/88=5.68×10^−4^ for gene expression; P<0.05/488=P<1.02×10^−4^ for metabolites) are provided in **Supplementary Tables 13-15**.

WHRadjBMI-associated variant, rs13301996, which is intronic to *CDK5RAP2* (CDK5 [cyclin-dependent kinase 5] Regulatory Subunit Associated Protein 2), was significantly associated with expression of 15 genes and one lncRNA across 17 tissue types (**Supplementary Table 14**). The most significant of these associations was with *MEGF9* (Multiple Epidermal Growth Factor-Like Domains 9) in whole blood (*P*=1.8×10^−149^), a gene that rests 30 Kb upstream of rs1330996. This SNP is also significantly associated with expression of *MEGF9* in subcutaneous adipose tissue, sun-exposed skin, and T-cells.

Additionally, our lead variant in *CDK5RAP2* is associated with expression of *MEGF9* in whole blood and the testis; and with expression of *PSMD5* (proteasome [prosome, macropain] 26S subunit, non-ATPase, 5) and/or *PSMD5-AS1* in several relevant tissues, including whole blood, tibial artery, tibial nerve, lung, thyroid, esophagus muscle, skeletal muscle, liver, cerebellum, and subcutaneous adipose tissues, among others. Although rs13301996 is associated with gene expression for several genes, there is additional support for a regulatory role of this SNP and those with which it is in high LD (r^2^>0.8). For example, our lead SNP lies just outside of a DNase hypersentivitiy cluster, lies within a region with evidence of histone modification in nine tissues including brain, skin, muscle, and heart; and likely falls in a transcription factor binding site active in skeletal and lung tissue; etc. (**Supplementary Table 16**) (39–41). While there are multiple lines of evidence for a regulatory role of this variant and multiple genes, rs13301996 has RegulomeDB Score of 6, indicating little evidence of binding.

WHRadjBMI-associated SNP, rs79478137, is a low frequency variant (MAF=1.6%) intronic in *SLC22A18AS* (Solute Carrier Family 22 (Organic Cation Transporter), Member 18 Antisense). This region is subject to genomic imprinting (43) which has been linked with Beckwith-Wiedemann syndrome, a disease caused by increased rate of growth in children (44–46). Our lead variant is associated with two Electronic Health Record (EHR)-derived phenotypes (cause of death: multisystem degeneration; and cause of death: tongue, unspecified) (**Supplementary Table 13**) in Phenoscanner. There is limited evidence of a regulatory role for our lead SNP (RegulomeDB Score = 4), but rs79478137 is in perfect LD with several variants with evidence of regulation (histone modification, open chromatin, DNAse hypersensitivity, transcription factor binding) in more than 50 tissues, including blood, pancreas, liver, and skeletal muscle, and hippocampal tissues, etc. (**Supplementary Table 16)** (39–41).

Our lead SNP associated with HIPadjBMI in women, rs28692724 (NC_000014.9:g.77027445C>T), is a synonymous variant exonic to *IRF2BPL* (interferon regulatory factor 2 binding protein like) that is significantly associated with expression of the same gene in whole blood (**Supplementary Table 14**).

Additionally, this variant lies in a known CTCF binding site (RegulomDB Score = 2b), among other transcription factors, and a DNAse Hypersentivity cluster (**Supplementary Table 16)** (39–41).

WCadjBMI-associated SNP, rs3168072, was significantly associated with existing GWAS traits present in Phenoscanner, including “cause of death: other specified respiratory disorders” (**Supplementary Table 13**). Additionally, rs3168072 is significantly associated with expression of several genes in whole blood, but most significantly associated with expression of *TMEM258* (Transmembrane Protein 258) (**Supplementary Table 14**). Rs3168072 is ~95 Kb upstream of *TMEM258*. Our lead variant is likely to play a role in gene expression regulation (RegulomeDB score= 2b, “likely to affect binding”) (39). Additionally, our lead variant and those in high LD (R^2^>0.8) lie within known DNase hypersentivitiy regions and within active areas of histone modification, open chromatin, and likely gene enhancer regions (**Supplementary Table 16)** (39–41). Our lead SNP associated with WCadjBMI, rs3168072, is significantly associated with five lipid-related metabolites **(Supplementary Table 15**), including “Other polyunsaturated fatty acids than 18:2,” “CH2 groups in fatty acids,” “Ratio of bis allylic bonds to double bonds in lipids,” “CH2 groups to double bonds ratio,” and “Ratio of bis allylic bonds to total fatty acids in lipids.”

## Discussion

We performed the first large-scale GWAS of 3 central adiposity traits (i.e., WHRadjBMI, WCadjBMI, and HIPadjBMI) in a sample of approximately 12,672 Hispanic/Latino individuals. We identified 16 variants that were suggestively associated (*P*<1×10^−6^) with WHRadjBMI, 22 for WCadjBMI, and 28 for HIPadjBMI. Of these 66 variants that were suggestively associated with the three central adiposity traits, four novel loci replicated after meta-analysis with replication samples. Additionally, we demonstrated that eight previously identified GWAS loci generalized to Hispanic/Latino study participants for WHRadjBMI, 10 for WCadjBMI, and 12 for HIPadjBMI in HCHS/SOL.

### Discovery of Four Novel Loci

Given the large number of published GWAS on central adiposity measures, it may seem surprising that four novel loci (rs13301996, rs79478137, rs28692724, and rs3168072) were mapped. We ascribe this to (1) previous GWAS were primarily conducted in European populations. Indeed, all four novel SNPs were absent from previous GIANT HapMap imputed analyses (35), and one (rs28692724) of the four absent from a more recent GWAS that included Europeans from the UK Biobank (33); (2) the consideration of a broad spectrum of ancestrally diverse Hispanic/Latino populations, including not just those of Mexican ancestry, but also those with ancestry from the Caribbean, Central, and/or South America (47); (3) the use of the entire 1,000 Genomes Phase I Reference panel, including populations with Native American ancestry: MXL (Mexico), CLM (Colombia), and PUR (Puerto Rico); (4) demonstrated differences in the patterning of body composition by ancestry (48, 49). More specifically, African ancestry populations have lower body fat percentages than men and women of non-Hispanic European, Native American, and East Asian ancestry at the same BMI. Additionally, non-Hispanic African ancestry men and women have greater skeletal and muscle mass than their non-Hispanic European ancestry counterparts, who in turn have greater skeletal and muscle mass than men and women of East Asian origin (48, 50–52)

Recent GWAS for coding variation of waist circumference traits identified the importance of central adiposity genes in lipid regulation, storage, and homeostasis (53). Similarly, we found a novel association of a variant in *FADS2* (rs3168072) with WCadjBMI following meta-analysis of HCHS/SOL results with results from an independent sample of European descent individuals, which further implies a role of this locus in central adiposity and lipid homeostasis. Genetic variations in the *FADS2* gene has been associated with several traits related to obesity and cardiometabolic health, including fatty acid metabolism and adipose tissue inflammation, leading to an interaction between weight loss and *FADS2* genes in the regulation of adipose tissue inflammation (54). A nearby variant, rs174546 (R^2^=0.3523,

D’=0.916 in AMR), in *FADS1* has previously been associated with four lipid traits (55). The A allele (MAF=38%) is associated with greater waist circumference in our samples, and is nearly fixed among sub-Saharan African populations (99% in 1000 Genomes AFR), at very high frequency in European populations (97% in EUR), and at a lower frequency in East Asian (75% in EAS) and Native American populations (63% in AMR). Rs3168072 is intronic to *FADS2* - a member of the fatty acid desaturase (FADS) gene family and is involved in the endogenous conversion of short-chain polyunsaturated fatty acids to long chain fatty acids. The *FADS* cluster of genes appears to have been under strong selection in several human populations, which likely explains the large differences in allele frequencies across global populations (56–59), and why previous GWAS of waist traits primarily focused on European descent populations did not detect an association signal in this region.

We identified a novel association for WHRadjBMI with rs13301996 following meta-analysis with an independent sample of African descent individuals. Rs13301996 is intronic to *CDK5RAP2*, which encodes a regulator of CDK5 (cyclin-dependent kinase 5) activity (60), interacts with CDK5R1 and pericentrin (PCNT) (60), plays a role in centriole engagement and microtubule nucleation (61), and has been linked to primary microcephaly and Alzheimer’s disease (62, 63). In addition, we identified a novel association for WHRadjBMI with rs79478137 (p-value= 3.64E^−9^) in Hispanic/Latino women. Rs79478137 is intronic to the antisense *SLC22A18AS* gene, which is highly expressed in the liver and kidney, as well as the gastrointestinal tract and placenta. Very little is known of the biological role of this gene (64), and *SLC22A18AS* has no counterpart in mice or other rodents (65). Thus, although its genomic organization is known, the regulation and function of this gene is not understood (66).

Lastly, we identified a novel association for HIPadjBMI at rs28692724 following meta-analysis with an independent sample of European women. Rs28692724 is a synonymous variant in *IRF2BPL*, which encodes a transcription factor that, acting within the neuroendocrine system, plays a role in regulating female reproductive function (67).

### Differences in Association by Background Group

Many of the loci mapped in this study displayed effect heterogeneity by background group. For example, the *NTM* locus associated with WHRadjBMI in women, displayed nearly threefold the effect size in the mainland background group compared to the Caribbean background group. Also, for the women-only primary analysis, rs77186623 in the *LOC105375745* locus displayed a fourfold greater effect in the Caribbean background group compared to the mainland group. These and other loci displaying heterogeneity by background group (i.e. *MARCKSL1, C19orf67, RIT2, MAP4K4, COBL, NREP*, and *ANO10*) were not validated in replication analyses, possibly due in part to heterogeneity by background group.

### Limitations

A limitation of this study was the small sample size within each HCHS/SOL background group. However, the use of genetic-analysis groups in our analyses accounted for heterogeneity of genetic effects among ethnic groups often ignored in GWAS studies. Compared to self-identified background groups, genetic-analysis groups are more genetically homogeneous and lack principal component outliers in stratified analysis, which may hinder detection of and adjustment for important population structure when ignored (68). In addition, genetic-analysis groups allow all individuals to be classified in a specific group, whereas many individuals in HCHS/SOL have a missing or non-specific self-identified background (68). Therefore, by using genetic-analysis groups in our analysis rather than self-identified background groups, we have increased our study’s power to detect novel and previously documented associations with central adiposity traits (68). Due to the diverse background of our discovery population, another limitation was the lack of an ideal replication study. We attempted to overcome this limitation by focusing on both multiethnic meta-analyses, which would validate those associations that generalize across ancestries, and meta-analyses stratified by ancestry, which may allow for validation of more population-specific associations. However, it is possible that the limited Native American ancestry present across our replication cohorts may have hindered replication, and further analyses in more diverse Hispanic/Latino populations are needed to confirm the relevance of promising central adiposity associated loci identified in our study.

## Conclusion

We identified 4 novel loci for central adiposity traits in a large population of Hispanic/Latino Americans. We also found that several previously identified central adiposity loci discovered in European American populations generalized to Hispanic / Latino Americans. Many of the loci interrogated exhibit subgroup-specific effects, likely due to population history (admixture, natural selection), that have resulted in changes in LD, or allele frequency differences, or due to variation in etiology. These observations highlight the importance of large-scale genomic studies in ancestrally diverse populations for identifying obesity-susceptibility loci that generalize and those that are ancestry-specific.

## Materials and Methods

### Study Sample

Details on the study and sampling design of the Hispanic Community Health Study /Study of Latinos (HCHS/SOL) have been previously described (69). Briefly, HCHS/SOL is a community based prospective cohort study of 16,415 self-identified Hispanic/Latino adults aged 18-74 years at screening from randomly selected households in four US field centers (Chicago, IL; Miami, FL; Bronx, NY; San Diego, CA) with baseline examination (2008 to 2011) and yearly telephone follow-up assessment for at least three years. The HCHS/SOL cohort includes participants who self-identified as being Central American (n=1,732), Cuban (n=2,348), Dominican (n=1,473), Mexican (n=6,472), Puerto-Rican (n=2,728), and South American (n=1,072). The goals of the HCHS/SOL are to describe the prevalence of risk and protective factors for chronic conditions (e.g. cardiovascular disease (CVD), diabetes and pulmonary disease), and to quantify all-cause mortality, fatal and non-fatal CVD and pulmonary disease, and pulmonary disease exacerbation over time. The baseline clinical examination (70) included comprehensive biological (e.g., anthropometrics, blood draw, oral glucose tolerance test, ankle brachial pressure index, electrocardiogram), behavioral (e.g. dietary intake assessed with two 24-hour recalls, physical activity assessment by accelerometer and self-report, overnight sleep exam for apneic events, tobacco and alcohol assessed by self-report), and socio-demographic (e.g., socioeconomic status, migration history) assessments. This study was approved by the institutional review boards at each field center, where all subjects gave written informed consent.

Participants in HCHS/SOL self-identified their background as Mexican, Central American, South American (mainland), Puerto Rican, Cuban, or Dominican (Caribbean). Some participants chose “more than one,” “other,” or chose not to self-identify. We addressed the missing or inconsistent data in self-identified background groups by defining “genetic analysis groups,” described in Conomos et al (68). To increase power in this analysis, we chose to stratify by the broader mainland or Caribbean categories rather than more specific groups. In this paper, we will use the term “background group” to refer to a super-group of genetic analysis groups by geographic region, mainland or Caribbean. Hispanics/Latinos have admixed ancestry from three continents: Africa, America, and Europe. In general, participants from the mainland group have higher proportions of American ancestry and lower African ancestry, while participants in the Caribbean group have higher proportions of African ancestry (68).

### Phenotypes

All measurements were taken from the baseline visit. Participants were dressed in scrub suits or light non-constricting clothing and shoes were removed for weight and height measurements. WC and HIP were measured using Gulick II 150 and 250 cm anthropometric tape and rounded to the nearest centimeter (cm). Height was measured using a wall mounted stadiometer and rounded to the nearest cm, and weight measured with a Tanita Body Composition Analyzer, TBF-300Ato the nearest tenth of a kilogram (kg). Height and weight were used to calculate BMI (kg/m^2^). We applied a log10 transformation on HIP, due to its non-normal trait distribution.

### Genotyping

Our analyses included 7,472 women and 5,200 men of mainland (Mexican, Central and South American) or Caribbean (Puerto Rican, Cuban, and Dominican) ancestry residing in the U.S. All participants were genotyped on the Illumina SOL Omni2.5M custom content array, which was subsequently used to impute millions of additional variants, based on the entire 1,000 Genomes Phase I Reference panel, including populations with Native American ancestry: MXL (Mexico), CLM (Colombia), and PUR (Puerto Rico). Pre-phasing was performed using SHAPEIT, followed by imputation with IMPUTE2 (71, 72).

### Discovery Analyses

Due to known differences in genetic effects on waist and hip traits between men and women (35, 73, 74), we analyzed associations stratified by sex for each trait, in addition to the entire sample. We used linear mixed-model regression, assuming an additive genetic model adjusted for age, age^2^, study center, sample weights, genetic analysis subgroup (68, 75), principal components to account for ancestry, population structure using kinship coefficients and sample eigenvectors, household, census block group, and sex in the combined analysis. Kinship, household, and block group were treated as random effects in each model. Sample weights were incorporated in our models as a fixed effect to account for oversampling of the communities in the 45-74 age group (n=9,714, 59.2%) which was intended to facilitate the examination of HCHS/SOL target outcomes. HCHS/SOL sampling weights are the product of a “base weight” (reciprocal of the probability of selection) and three adjustments: 1) non-response adjustments made relative to the sampling frame, 2) trimming to handle extreme values (to avoid a few weights with extreme values being overly influential in the analyses), and 3) calibration of weights to the 2010 U.S. Census according to age, sex, and Hispanic background. We used genetic-analysis groups in our analyses accounted for heterogeneity of genetic effects among ethnic groups.

Compared to self-identified background groups, genetic-analysis groups are more genetically homogeneous and lack principal component outliers in stratified analysis, which may hinder detection of and adjustment for important population structure when ignored (68). In addition, genetic-analysis groups allow all individuals to be classified in a specific group, whereas many individuals in HCHS/SOL have a missing or non-specific self-identified background (68). Also, we conducted stratified analyses by region (mainland vs. Caribbean) to identify potential heterogeneity in effect by background group. We examined heterogeneity across background group using I^2^ statistics calculated using METAL (76) and tested for significant interaction (P_diff_<0.05) by background group using EasyStrata (77).

To decrease the number of spurious associations, we filtered all results on minor allele frequency (MAF) < 0.5%, Hardy-Weinberg Equilibrium (HWE) *P* < 1×10^−7^, minor allele count (MAC [effective N]) < 30 (68). Additionally, we categorized suggestive loci as those with variants reaching *P*<1×10^−6^ and with at least one additional variant within 500 kb+/− with a *P*<1×10^−5^. We used regional association plots produced in LocusZoom to visualize association regions using 1000 Genomes Admixed American (AMR) reference population for LD (http://locuszoom.sph.umich.edu/).

### Local Ancestry Estimation

We estimated local ancestry (African, Native American, and European) using RFMix (78), which applies a conditional-random-field-based approach for estimation, to inform differences by background group. We used 236,456 genotyped SNPs available in both HCHS/SOL and reference-panel datasets in the Human Genome Diversity Project (HGDP) (79), HapMap 3 (80), and 1000 Genomes phase 1 for detecting African, Native American, and European ancestry. We used BEAGLE (v.4) to phase and impute sporadic missing genotypes in the HCHS/SOL and reference-panel datasets (81).

### Replication and Meta-Analyses

An aim of our study was to identify genetic variants that contribute to central adiposity which may vary by ancestry; therefore, we sought to replicate our association findings using 1000 Genomes imputed GWAS data available in independent cohorts including eight studies with Hispanics/Latinos (HL: N up to 12,341), three studies with African-Americans (AA; N up to 12,496), and one study with European-Americans (EUR: N up to 8,845). Study design and descriptive statistics for each replication study are provided in **Supplementary Table 1**. Each replication study excluded individuals that were pregnant or exhibited extreme values for waist or hip measures (outside of ±4 SD from the mean). Each study used measures from a single visit with the greatest sample size. We used linear regression (or linear mixed effects models if the study had related individuals) association analyses on the trait residuals after adjustment for age, age^2^, principal components to account for ancestry, BMI, other study specific factors (e.g. study center), and sex in the sex combined analysis, stratified by race/ethnicity where applicable for each SNP that reached suggestive significance (*P*<1×10^−6^) in the discovery analysis.

We employed a fixed-effects meta-analysis using the inverse variance-weighted method for WHRadjBMI and WCadjBMI. For HIPadjBMI, due to trait transformations, we used sample size weighted meta-analysis. All meta-analyses were implemented in METAL (82). We conducted meta-analyses stratified by race/ethnicity group and combined across groups. We included SNPs with a study and stratum specific imputation quality (Rsq) greater than 0.4, Hardy-Weinberg Equilibrium P-value greater than 1×10^−7^, and a minor allele count (MAC) greater than five. To declare statistical significance for replicated loci, we required in each replication sample a trait and stratum-specific *P*<0.05 with a consistent direction of effect with discovery, and genome-wide significance (*P*<5×10^−8^) when meta-analyzed together with HCHS/SOL.

### Generalization

To examine whether previously reported association regions generalized to the HCHS/SOL, we downloaded the publicly available multi-ethnic (European, Asian, and African ancestry) GWAS results from the Genetic Investigation of Anthropometric Traits (GIANT) consortium (14) for HIPadjBMI, WHRadjBMI, and WCadjBMI (https://portals.broadinstitute.org/collaboration/giant/index.php/GIANT_consortium_data_files#GIANT_consortium_2012-2015_GWAS_Metadata_is_Available_Here_for_Download), in men, women, and sexes combined, and applied the framework of Sofer et al. (2016) for generalization testing (83). We took all variant associations with *P*<1×10^−6^ in GIANT and identified the matching association test in HCHS/SOL. For each such association, we calculated a directional FDR r-value, by combining the *P*-values from GIANT and HCHS/SOL, while accounting for multiple testing and for the direction of estimated associations in each of the studies. Then, an association was declared as generalized, while controlling for the False Discovery Rate (FDR) at the 0.05 level, if its r-value was smaller than 0.05. Note that multiple SNPs from the same region were tested. Therefore, in an iterative procedure, we pruned the results list by identifying the SNP with the lowest r-value in an analysis, then finding all SNPs in a 1MB region around it and removing them from the list. Thus, the number of generalized regions is the number of generalized SNPs in the pruned list.

We also hypothesized that some regions did not generalize due to lack of power (the HCHS/SOL sample size is much smaller than the GIANT sample size). To test this, we took all tested SNPs from the non-generalized regions and considered the GIANT multi-ethnic GWAS results. In an iterative procedure, we pruned the list by first identifying the SNP with lowest GIANT *P*-value in the analysis, then found all SNPs in a 1MB region around it and removed them from the list. We repeated until no SNPs remained. All the SNPs in the pruned list were selected solely based on their GIANT *P*-values. Since there were many such variants, we further grouped them according to their *P*-values. Groups were formed by trait, sex (men, women, combined), and GIANT P-value (between 10^−6^ to 10^−7^, between 10^−7^ to 10^−8^, and smaller than 10^−8^). For each such group of SNPs, we created a genetic risk score (GRS) in HCHS/SOL. For each sex stratum and each group of SNPs, the value of the GRS was the sum of all trait increasing alleles in that group. We tested the GRS in the appropriate analysis group (men, women, combined). A low *P*-value implies that some of the SNPs in the group are likely associated with the trait in HCHS/SOL.

### Biological Curation

To gain further insight into the possible functional role of the identified variants and to assess the relevance of our identified variants with other phenotypes, we conducted lookups of our replicated variants in multiple publicly available databases, including PhenoScanner (37), RegulomeDB (39), Haploreg (40), and UCSC GenomeBrowser (41). Additionally, we conducted lookups of nearby genes in GTeX (42). The R package HaploR was used to query HaploReg and RegulomeDB (https://cran.r-project.org/web/packages/haploR/vignettes/haplor-vignette.html).

## Data Availability

Raw genetic data used in the discovery analysis for this project is available through request on dbGAP (dbGaP Study Accession: phs000810.v1.p1). Additionally, the full GWAS summary results for common variants with a MAF<1% will be made available upon request to the author and on the NHGRI-GWAS Catalog (upon publication).

## Author contributions

Conceived of Study Design: AEJ, KY, KEN; Drafted Manuscript: AEJ, KY, KEN, MG, SAL; Acquisition of Data: AEJ, KEN, CL, CI; Data Preparation: SMG, TS; Performed Statistical Analyses: SMG, TS, AEJ, LEP, YW; Prepared Figures and Table: AEJ, KL, SAL, MG, SMG, TS; Performed Look-ups: AEJ; Supervised the work: KEN, CL, CI. All authors revised and approved the manuscript, assisted in interpretation of results, and agree to be held accountable for the content.

## Acknowledgements

AEJ was supported in part by National Institute of Health (NIH), National Heart, Lung, and Blood Institutes (NHLBI) grant K99/R00 HL 130580 and American Heart Association (AHA) grant 13POST16500011. YCK is supported by the National Heart, Lung, and Blood Institutes (R01-HL136528). HMH was supported in part by NIH NHLBI grant T32 HL007055.

## HCHS/SOL

The authors thank the staff and participants of HCHS/SOL for their important contributions. (Investigators website - http://www.cscc.unc.edu/hchs/). The Hispanic Community Health Study/Study of Latinos is a collaborative study supported by contracts from the National Heart, Lung, and Blood Institute (NHLBI) to the University of North Carolina (HHSN268201300001I / N01-HC-65233), Universit y o f Miami (HHSN268201300004I / N01-HC-65234), Albert Einstein College of Medicine (HHSN268201300002I / N01-HC-65235), University of Illinois at Chicago – HHSN268201300003I / N01-HC-65236 Northwestern Univ), and San Diego State University (HHSN268201300005I / N01-HC-65237). The following Institutes/Centers/Offices have contributed to the HCHS/SOL through a transfer of funds to the NHLBI: National Institute on Minority Health and Health Disparities, National Institute on Deafness and Other Communication Disorders, National Institute of Dental and Craniofacial Research, National Institute of Diabetes and Digestive and Kidney Diseases, National Institute of Neurological Disorders and Stroke, NIH Institution-Office of Dietary Supplements. The Genetic Analysis Center at the University of Washington was supported by NHLBI and NIDCR contracts (HHSN268201300005C AM03 and MOD03).

## ARIC

The Atherosclerosis Risk in Communities Study has been funded in whole or in part with Federal funds from the National Heart, Lung, and Blood Institute, National Institutes of Health, Department of Health and Human Services (contract numbers HHSN268201700001I, HHSN268201700002I, HHSN268201700003I, HHSN268201700004I and HHSN268201700005I), R01HL087641, R01HL059367 and R01HL086694; National Human Genome Research Institute contract U01HG004402; and National Institutes of Health contract HHSN268200625226C. Infrastructure was partly supported by Grant Number UL1RR025005, a component of the National Institutes of Health and NIH Roadmap for Medical Research. The authors thank the staff and participants of the ARIC study for their important contributions.

## GUARDIAN, MACAD, HTN-IR, GOLDR

This research was supported by the GUARDIAN (Genetics Underlying Diabetes in Hispanics) Study, National Institute of Diabetes and Digestive and Kidney Diseases (NIDDK) contract DK085175; (Mexican-American Coronary Artery Disease (MACAD) National Heart, Lung, and Blood Institute, contracts R01-HL088457, R01-HL-60030; National Heart, Lung, and Blood Institute, Hypertension and Insulin Resistance (HTN-IR) contracts R01-HL067974, R01-HL-55005, R01-HL 067974; National Institutes of Health, Genetics of Latino Diabetic Retinopathy (GOLDR) contract EY014684.

## MESA

MESA and the MESA SHARe projects are conducted and supported by the National Heart, Lung, and Blood Institute (NHLBI) in collaboration with MESA investigators. Support for MESA is provided by contracts 75N92020D00001, HHSN268201500003I, N01-HC-95159, 75N92020D00005, N01-HC-95160, 75N92020D00002, N01-HC-95161, 75N92020D00003, N01-HC-95162, 75N92020D00006, N01-HC-95163, 75N92020D00004, N01-HC-95164, 75N92020D00007, N01-HC-95165, N01-HC-95166, N01-HC-95167, N01-HC-95168, N01-HC-95169, UL1-TR-000040, UL1-TR-001079, and UL1-TR-001420. Also supported by the National Center for Advancing Translational Sciences, CTSI grant UL1TR001881, and the National Institute of Diabetes and Digestive and Kidney Disease Diabetes Research Center (DRC) grant DK063491 to the Southern California Diabetes Endocrinology Research Center.

## The Mexico City 1 and Mexico City 2

studies were supported in Mexico by the Fondo Sectorial de Investigación en Salud y Seguridad Social (SSA/IMSS/ISSSTECONACYT, project 150352), Temas Prioritarios de Salud Instituto Mexicano del Seguro Social (2014-FIS/IMSS/PROT/PRIO/14/34), and the Fundación IMSS. We thank Miguel Alexander Vazquez Moreno, Daniel Locia and Araceli Méndez Padrón for technical support in Mexico. In Canada, this research was enabled in part by two CIHR Operating grants to EJP, a CIHR New Investigator Award to EJP and by support provided by Compute Ontario (www.computeontario.ca), and Compute Canada (www.compute.canada.ca).

## WHI

Funding support for the “Epidemiology of putative genetic variants: The Women’s Health Initiative” study is provided through the NHGRI grants HG006292 and HL129132. The WHI program is funded by the National Heart, Lung, and Blood Institute, National Institutes of Health, U.S. Department of Health and Human Services through contracts HHSN268201100046C, HHSN268201100001C, HHSN268201100002C, HHSN268201100003C, HHSN268201100004C, and HHSC271201100004C. The authors thank the WHI investigators and staff for their dedication, and the study participants for making the program possible. A full listing of WHI investigators can be found at: http://www.whiscience.org/publications/WHI_investigators_shortlist.pdf.

## Data Availability Statement

Raw genetic data used in the discovery analysis for this project is available through request on dbGAP (dbGaP Study Accession: phs000810.v1.p1). Additionally, the full GWAS summary results for common variants with a MAF>1% will be made available upon request to the author and on the NHGRI-GWAS Catalog (upon publication).

