## Supplement for "Genome-wide association study of body fat distribution traits in Hispanics/Latinos from the HCHS/SOL Study"

### Supplementary Materials

#### Table of Contents


| Supplementary Figures | | Pages 3-100 |
|---|---|---|
| Supplementary Figure 1 | Manhattan Plot: WHRadjBMI, Sexes-Combined | 3 |
| Supplementary Figure 2 | Locus Zoom Plots: WHRadjBMI, Sexes-Combined | 4-8 |
| Supplementary Figure 3 | Manhattan Plot: WHRadjBMI, Women | 9 |
| Supplementary Figure 4 | Locus Zoom Plots: WHRadjBMI, Women | 10-18 |
| Supplementary Figure 5 | Manhattan Plot: WHRadjBMI, Men | 19 |
| Supplementary Figure 6 | Locus Zoom Plots: WHRadjBMI, Men | 20-25 |
| Supplementary Figure 7 | QQ Plots: WHRadjBMI | 26 |
| Supplementary Figure 8 | Manhattan Plot: WCadjBMI, Sexes-Combined | 27 |
| Supplementary Figure 9 | Locus Zoom Plots: WCadjBMI, Sexes-Combined | 28-37 |
| Supplementary Figure 10 | Manhattan Plot: WCadjBMI, Women | 38 |
| Supplementary Figure 11 | Locus Zoom Plots: WCadjBMI, Women | 39-41 |
| Supplementary Figure 12 | Manhattan Plot: WCadjBMI, Men | 42 |
| Supplementary Figure 13 | Locus Zoom Plots: WCadjBMI, Men | 43-54 |
| Supplementary Figure 14 | QQ Plots: WCadjBMI | 55 |
| Supplementary Figure 15 | Manhattan Plot: HIPadjBMI, Sexes-Combined | 56 |
| Supplementary Figure 16 | Locus Zoom Plots: HIPadjBMI, Sexes-Combined | 57-65 |
| Supplementary Figure 17 | Manhattan Plot: HIPadjBMI, Women | 66 |
| Supplementary Figure 18 | Locus Zoom Plots: HIPadjBMI, Women | 67-79 |
| Supplementary Figure 19 | Manhattan Plot: HIPadjBMI, Men | 80 |
| Supplementary Figure 20 | Locus Zoom Plots: HIPadjBMI, Men | 81-90 |
| Supplementary Figure 21 | QQ Plots: HIPadjBMI | 91 |
| Supplementary Figure 22 | Generalization of Known Loci: WHRadjBMI, Sexes-Combined | 92 |
| Supplementary Figure 23 | Generalization of Known Loci: WHRadjBMI, Women | 93 |
| Supplementary Figure 24 | Generalization of Known Loci: WHRadjBMI, Men | 94 |
| Supplementary Figure 25 | Generalization of Known Loci: WCadjBMI, Sexes-Combined | 95 |
| Supplementary Figure 26 | Generalization of Known Loci: WCadjBMI, Women | 96 |
| Supplementary Figure 27 | Generalization of Known Loci: WCadjBMI, Men | 97 |
| Supplementary Figure 28 | Generalization of Known Loci: HIPadjBMI, Sexes-Combined | 98 |
| Supplementary Figure 29 | Generalization of Known Loci: HIPadjBMI, Women | 99 |
| Supplementary Figure 30 | Generalization of Known Loci: HIPadjBMI, Men | 100 |
| **Supplementary Tables** | | **Pages 101-121** |
| Supplementary Table 1 | Study Descriptives and Genotyping Info | 101 |
| Supplementary Table 2 | Discovery and Replication Results: WHRadjBMI | 102 |
| Supplementary Table 3 | Discovery and Replication Results: WCadjBMI | 103-104 |
| Supplementary Table 4 | Discovery and Replication Results: HIPadjBMI | 105-106 |
| Supplementary Table 5 | Association by Background Group: WHRadjBMI | 107 |
| Supplementary Table 6 | Association by Background Group: WCadjBMI | 108 |
| Supplementary Table 7 | Association by Background Group: HIPadjBMI | 109 |
| Supplementary Table 8 | Local Ancestry | 110-111 |
| Supplementary Table 9 | Top Variant Generalization: WHRadjBMI | 112 |
| Supplementary Table 10 | Top Variant Generalization: WCadjBMI | 113 |

| | | |
|---|---|---|
| Supplementary Table 11 | Top Variant Generalization: HIPadjBMI | 114 |
| Supplementary Table 12 | Genetic Risk Score Generalization | 115 |
| Supplementary Table 13 | Phenoscanner Results: GWAS | 116 |
| Supplementary Table 14 | Phenoscanner Results: eQTL | 117-118 |
| Supplementary Table 15 | Phenoscanner Results: mQTL | 119 |
| Supplementary Table 16 | HaploReg Results (Note: Full results in separate file) | 120-121 |

**Supplementary Figure 1. Manhattan plot.** Manhattan plot of the sexes-combined analysis for WHRadjBMI. All suggestively significant (*P*<1x10$^{-6}$) variants are highlighted in orange if they are >500 Kb from any previously-reported WHRadjBMI associated variants. Previously reported loci (+/- 500 Kb) are highlighted in blue if any variant in the locus reached suggestive significance. All suggestively significant loci that meet our criteria for replication are annotated with the closest gene. †Replicated in African American meta-analysis. ‡ Replicated in Hispanic/Latino meta-analysis. ¥ Replicated in European American meta-analysis.

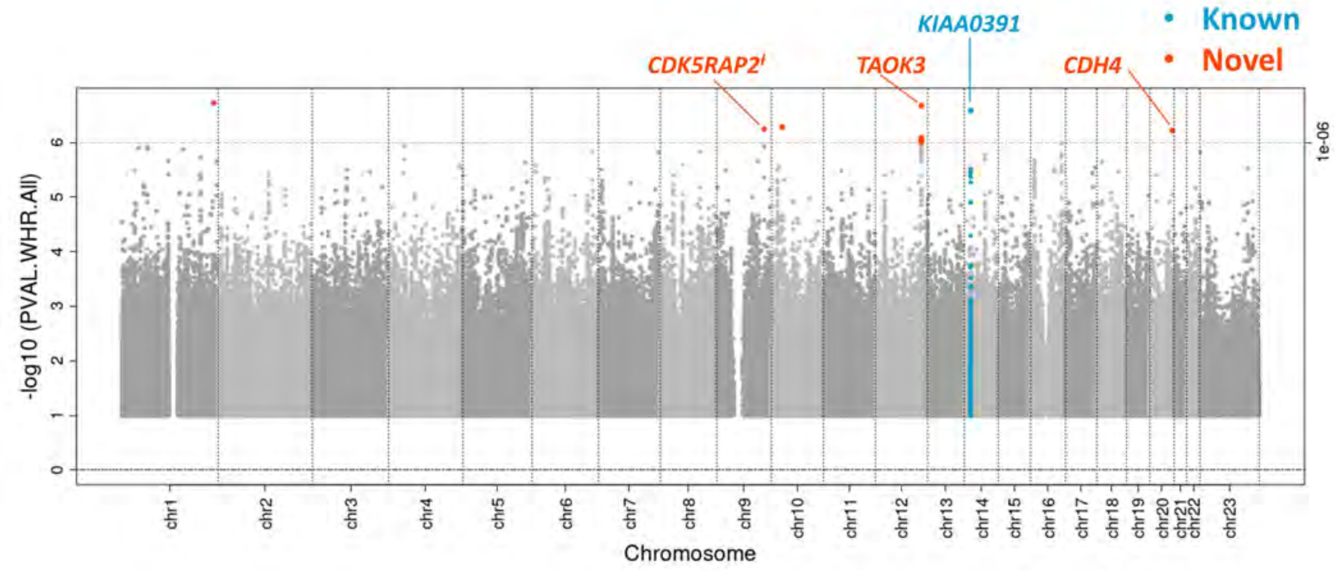

**Supplementary Figure 2. Locus Zoom Plots.** Regional association plots for suggestively significant loci in the HCHS/SOL WHRadjBMI sexes-combined analysis. The plots appear in chromosome:position order. Dot color reflects $R^2$ calculated from the 1000 Genomes AMR reference dataset. Point symbols represent variant functional classifications: a) rs13301996, *CDK5RAP2*; b) rs115981023, *TAOK3*; c) rs185566196, *KIAA0391*; d) rs116612483, *CDH4*.

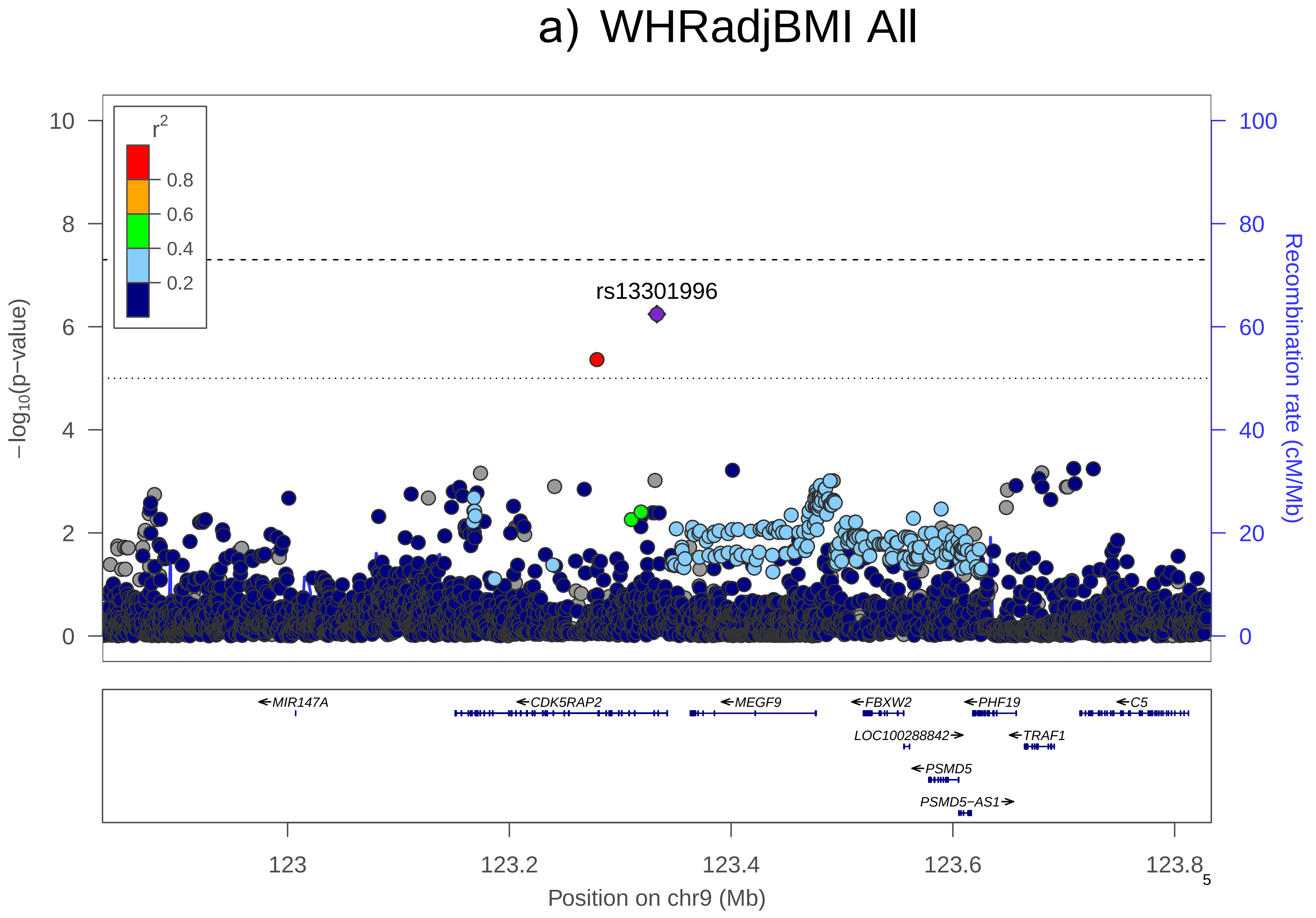
a) WHRadjBMI All
r2
0.8
0.6
0.4
0.2
rs13301996
−log10(p−value)
Recombination rate (cM/Mb)
10
8
6
4
2
0
100
80
60
40
20
0
MIR147A
CDK5RAP2
MEGF9
FBXW2
PHF19
C5
LOC100288842
TRAF1
PSMD5
PSMD5−AS1
123
123.2
123.4
123.6
123.8
Position on chr9 (Mb)

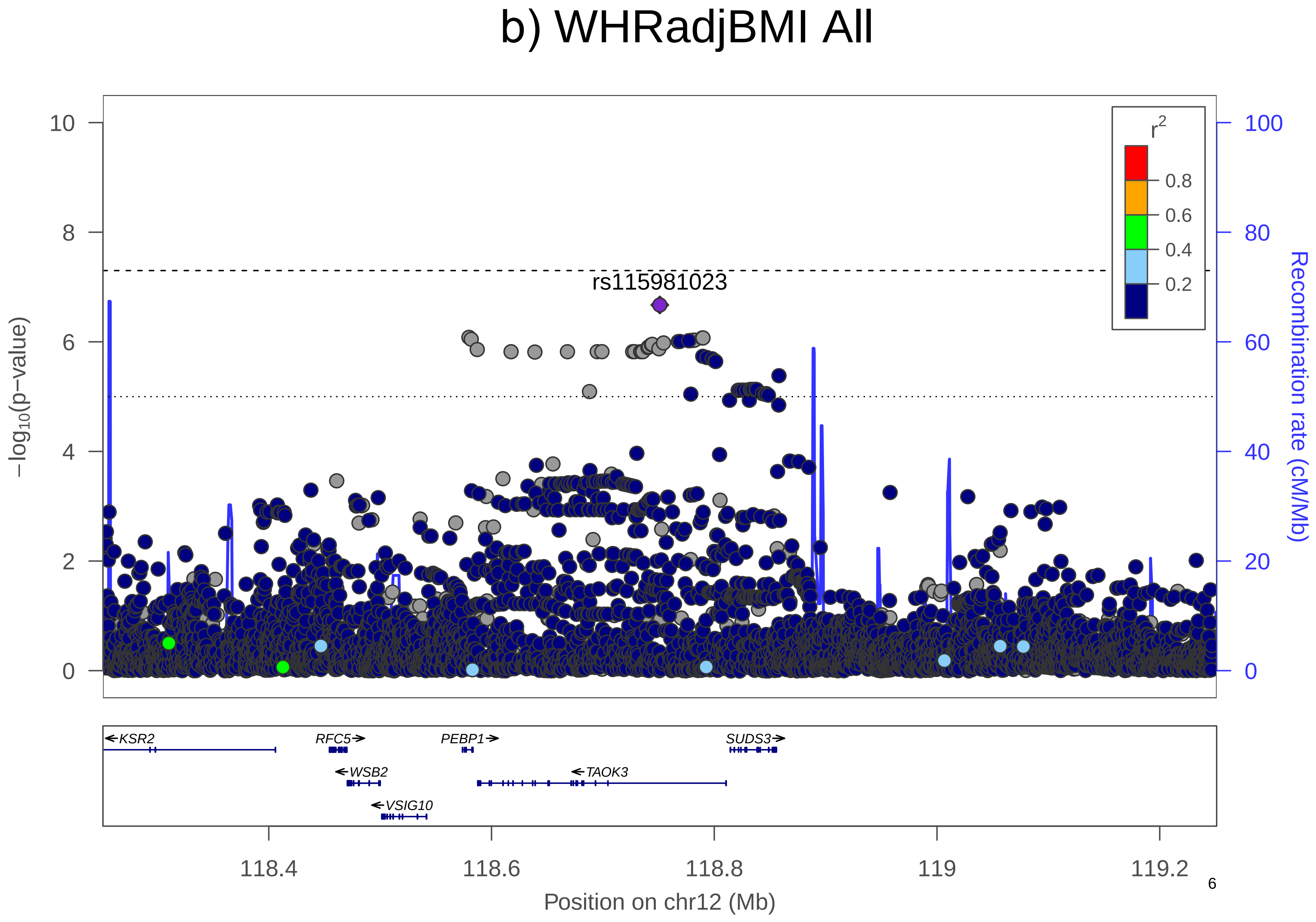
b) WHRadjBMI All
rs115981023
r2
0.8
0.6
0.4
0.2
10
8
6
4
2
0
100
80
60
40
20
0
−log10(p−value)
Recombination rate (cM/Mb)
KSR2
RFC5
PEBP1
SUDS3
WSB2
TAOK3
VSIG10
118.4
118.6
118.8
119
119.2
Position on chr12 (Mb)

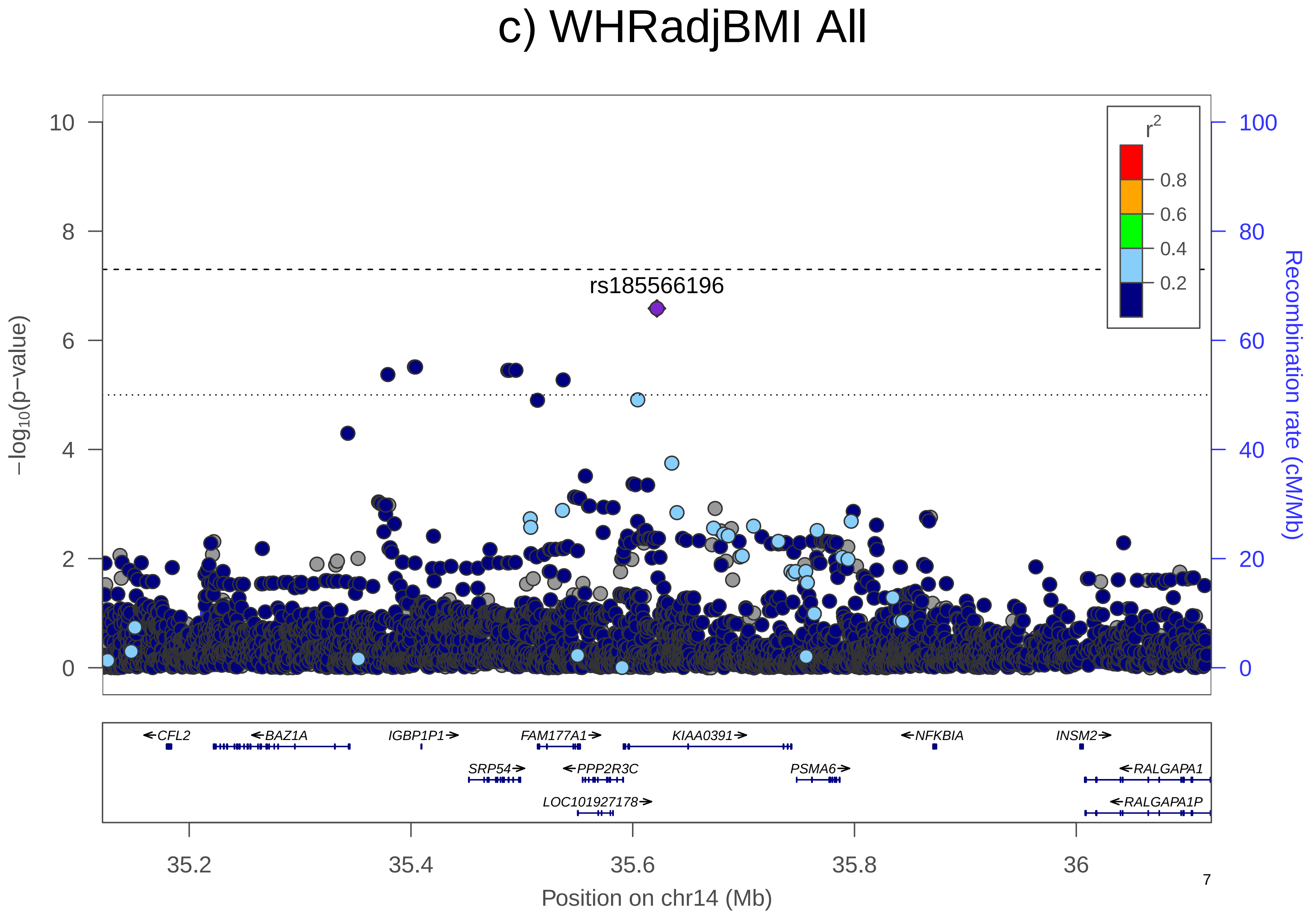
c) WHRadjBMI All
rs185566196
r2
0.8
0.6
0.4
0.2
−log10(p−value)
Recombination rate (cM/Mb)
CFL2
BAZ1A
IGBP1P1
FAM177A1
KIAA0391
NFKBIA
INSM2
SRP54
PPP2R3C
PSMA6
RALGAPA1
LOC101927178
RALGAPA1P
35.2
35.4
35.6
35.8
36
Position on chr14 (Mb)

### d) WHRadjBMI All

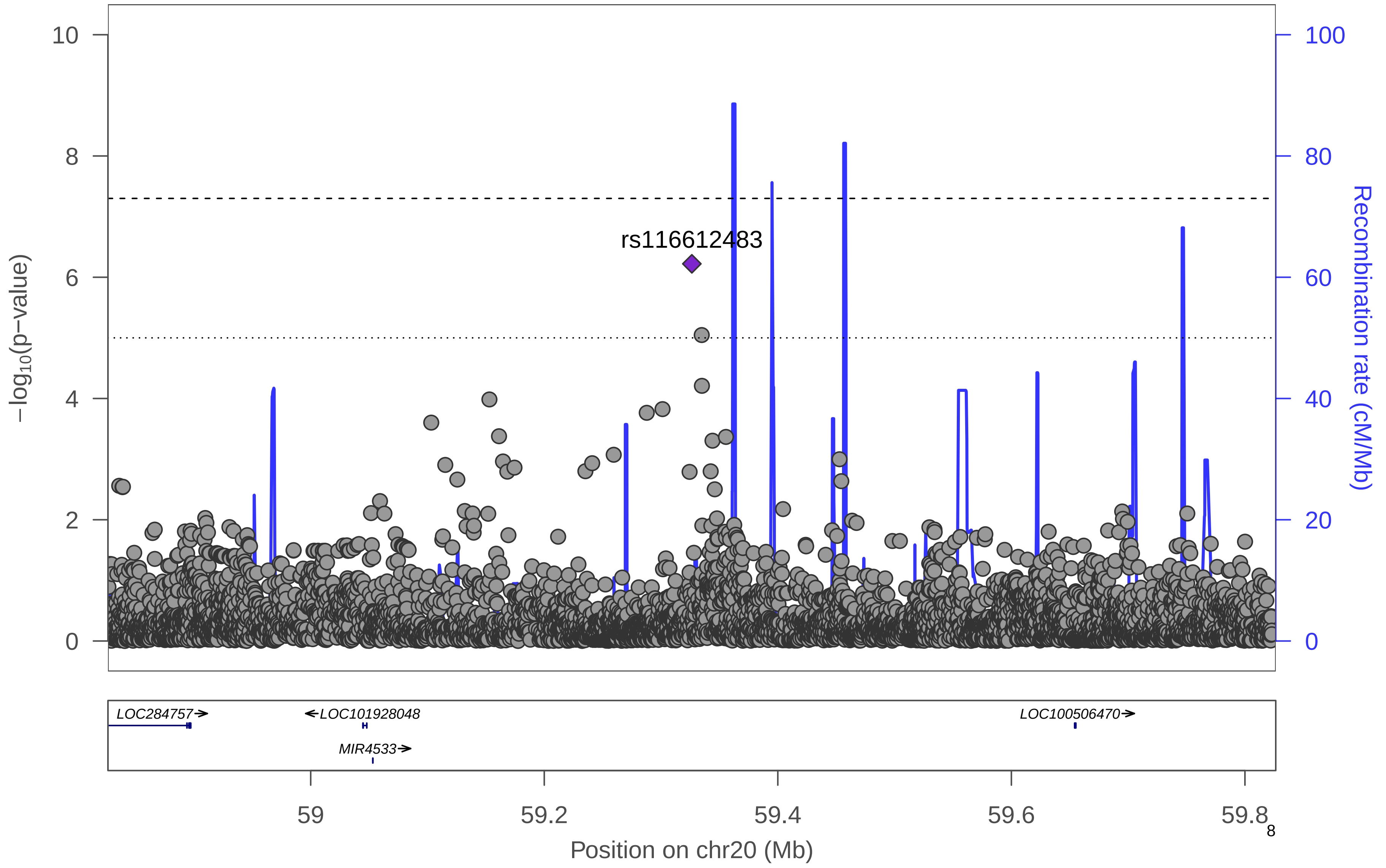

**Supplementary Figure 3. Manhattan plot.** Manhattan plot of the women-only analysis for WHRadjBMI. All suggestively significant (*P*<1x10$^{-6}$) variants are highlighted in orange if they are >500 Kb from any previously-reported WHRadjBMI associated variants. Previously reported loci (+/- 500 Kb) are highlighted in blue if any variant in the locus reached suggestive significance. All suggestively significant loci that meet our criteria for replication are annotated with the closest gene. †Replicated in African American meta-analysis. ‡ Replicated in Hispanic/Latino meta-analysis. ¥ Replicated in European American meta-analysis.

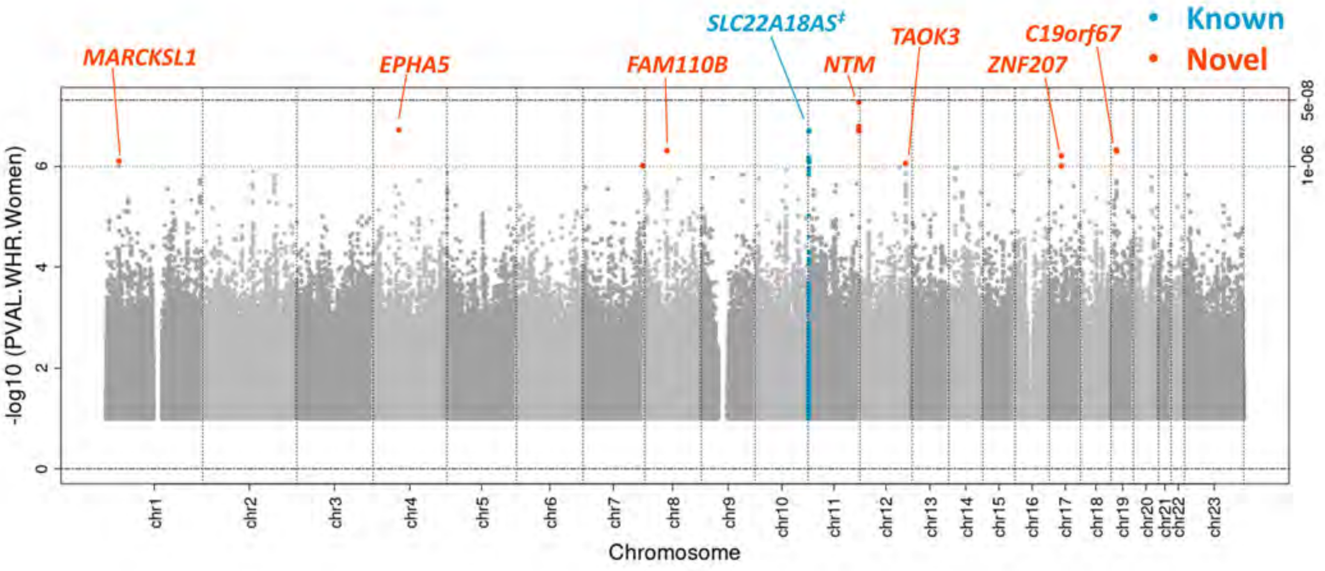

**Supplementary Figure 4. Locus Zoom Plots.** Regional association plots for suggestively significant loci in the HCHS/SOL WHRadjBMI women-only analysis. The plots appear in chromosome:position order. Dot color reflects $R^2$ calculated from the 1000 Genomes AMR reference dataset. Point symbols represent variant functional classifications: a) rs77377042, *MARCKSL1*; b) rs75120960, *EPHA5*; c) rs16922424, *FAM110B*; d) rs79478137, *SLC22A18AS*; e) rs113818604, *NTM*; f) rs115981023, *TAOK3*; g) rs146900844, *ZNF207*; h) rs61305557, *C19orf67*.

a) WHRadjBMI Women

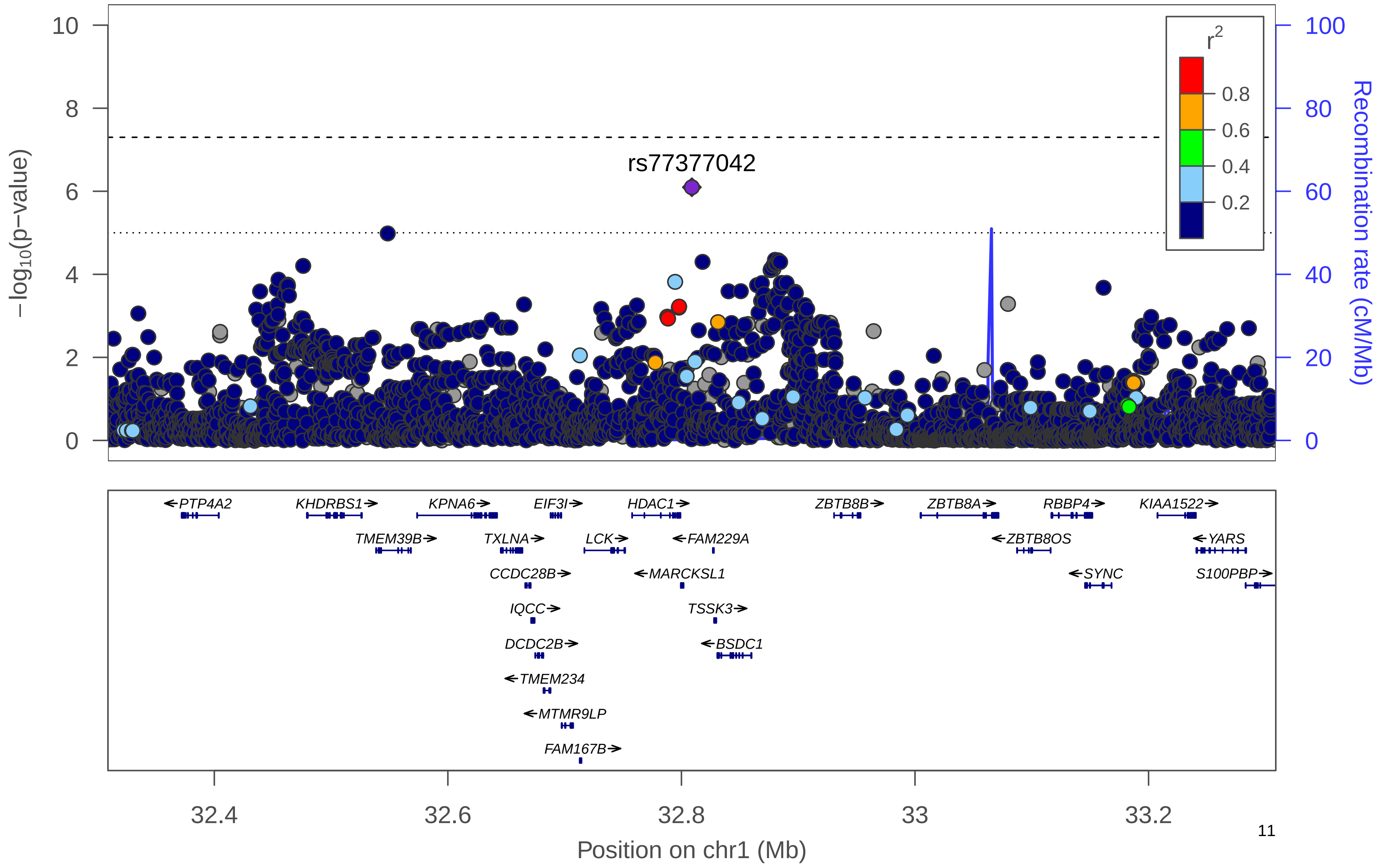

### b) WHRadjBMI Women

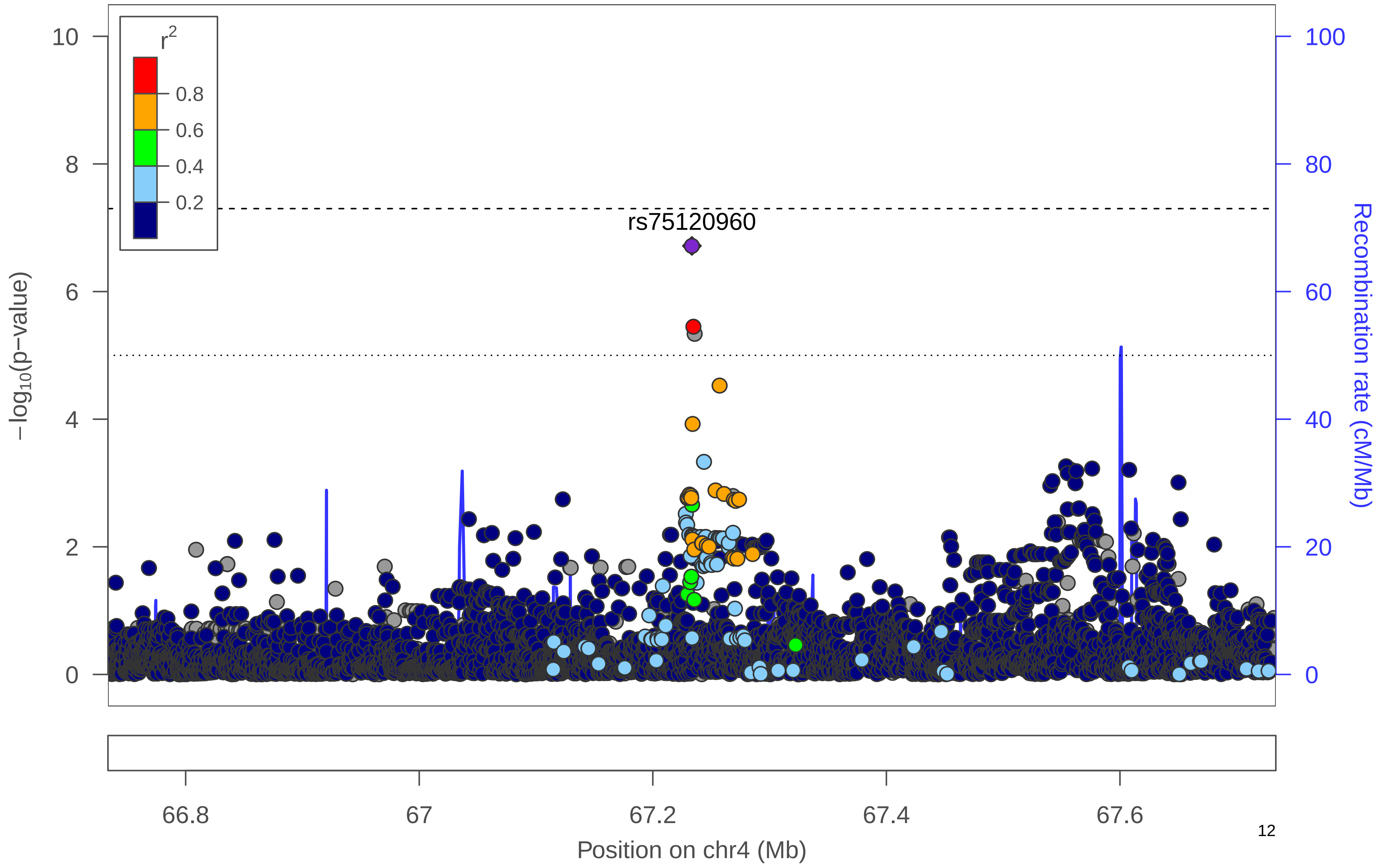

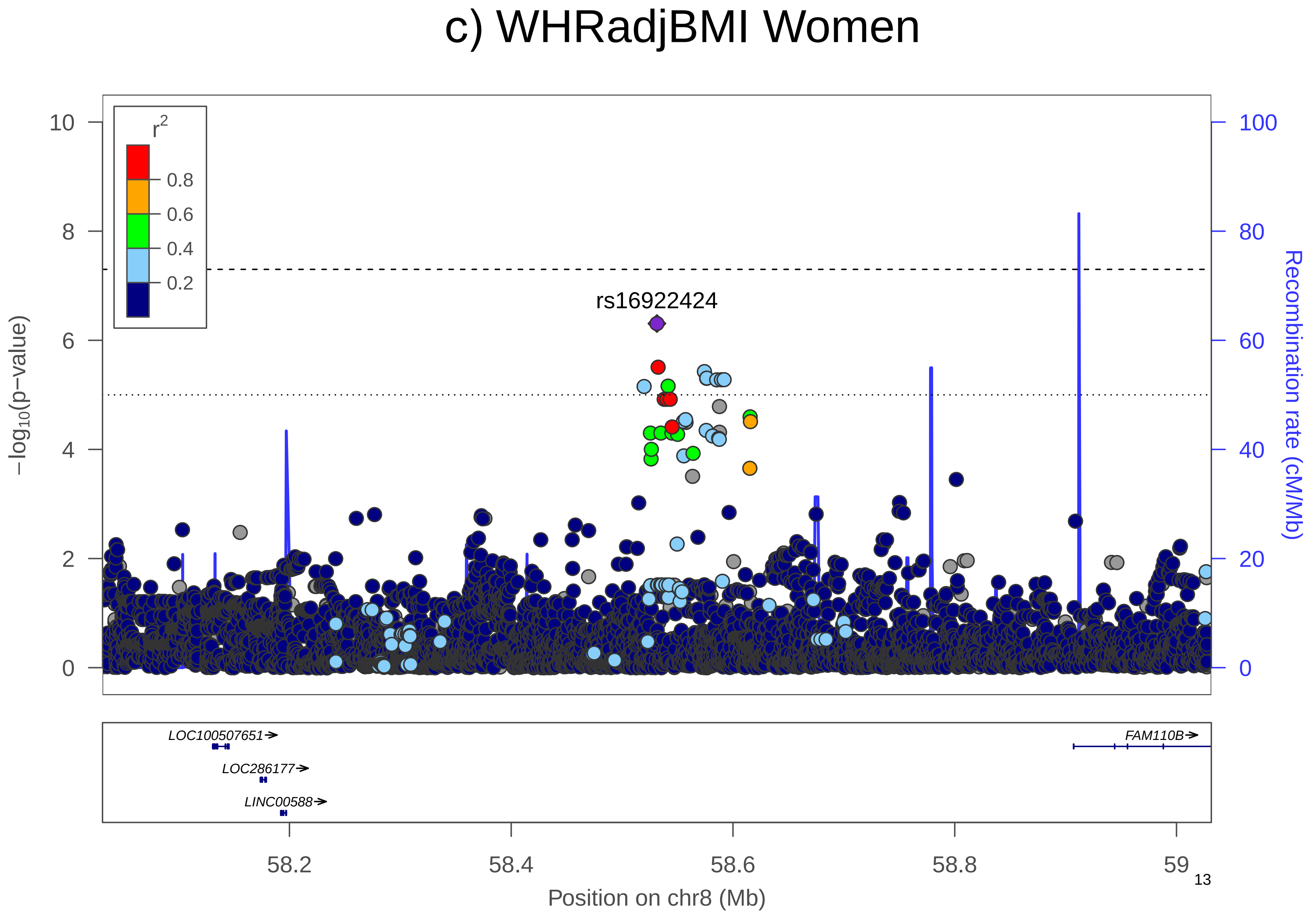

c) WHRadjBMI Women
r2
0.8
0.6
0.4
0.2
rs16922424
−log10(p−value)
Recombination rate (cM/Mb)
10
8
6
4
2
0
100
80
60
40
20
0
LOC100507651→
LOC286177→
LINC00588→
FAM110B→
58.2
58.4
58.6
58.8
59
Position on chr8 (Mb)

d) WHRadjBMI Women

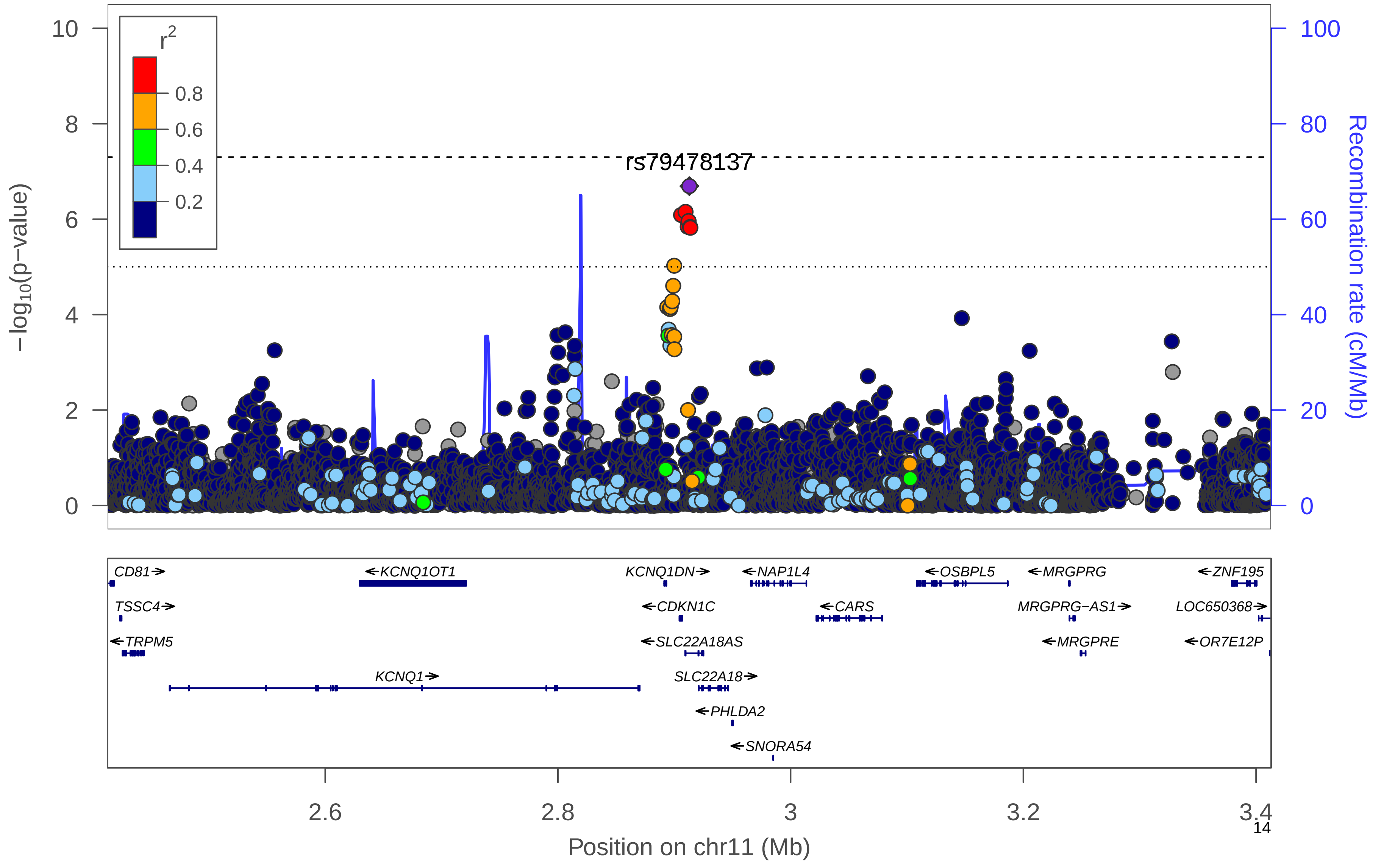

### e) WHRadjBMI Women

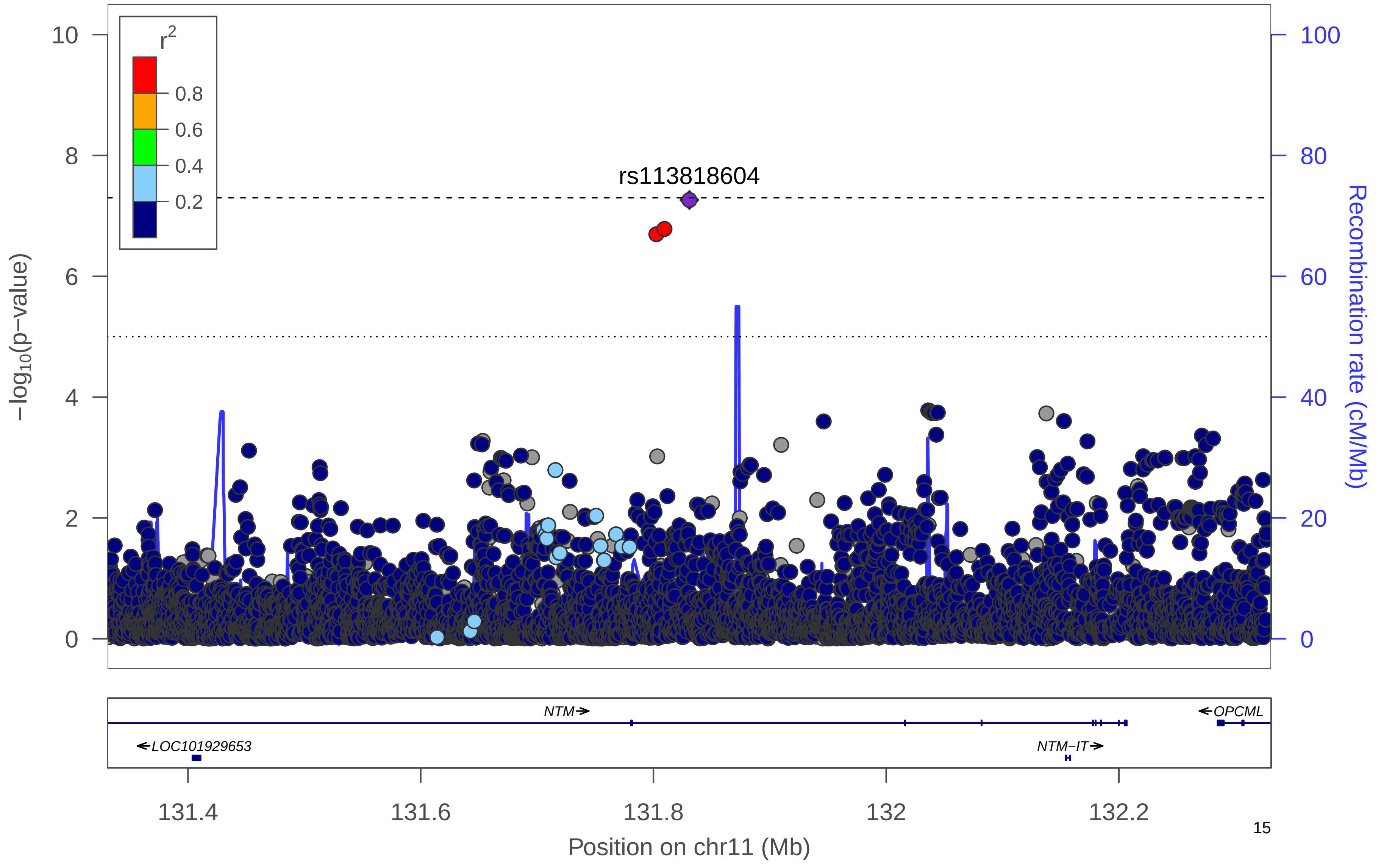

### f) WHRadjBMI Women

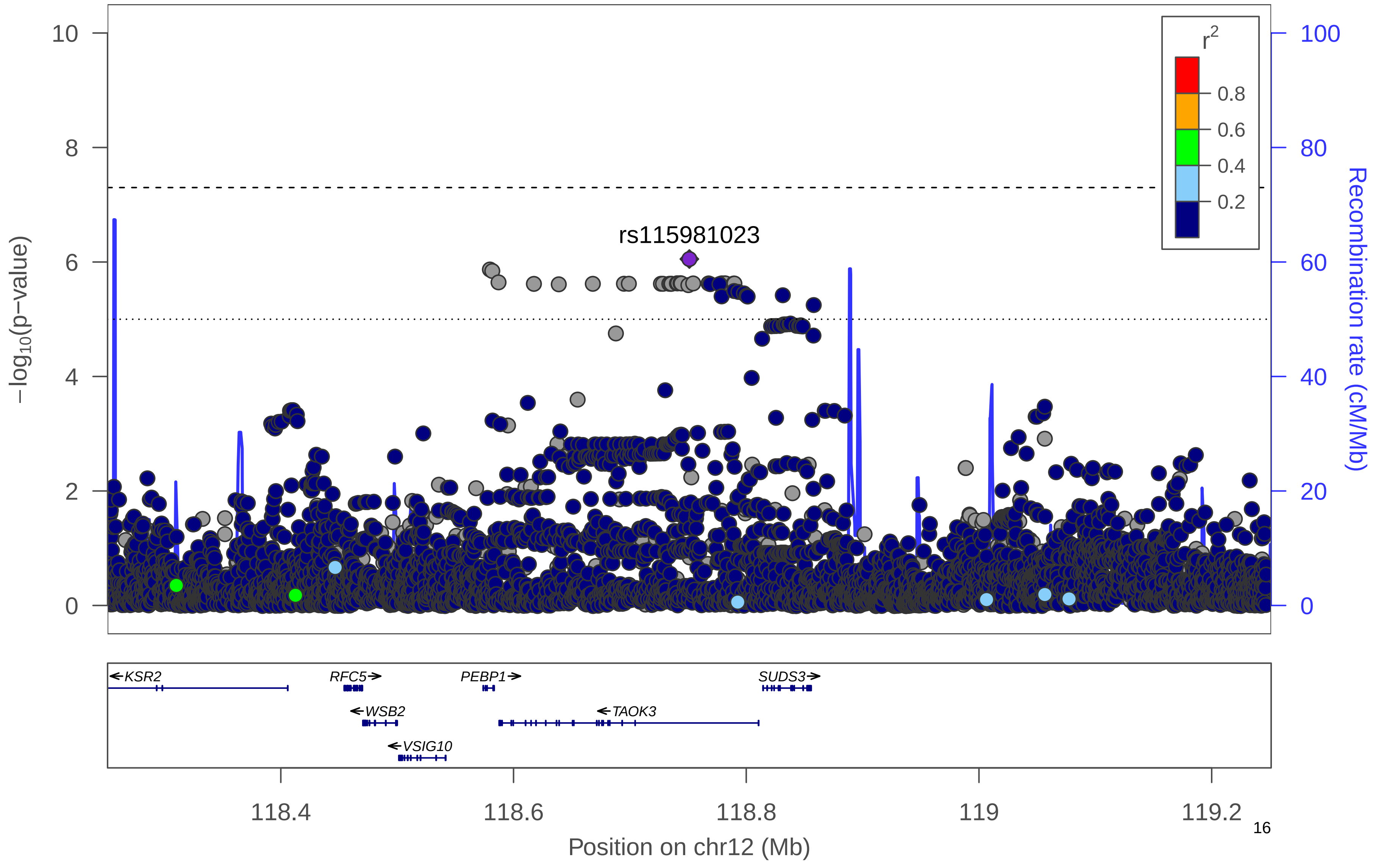

### g) WHRadjBMI Women

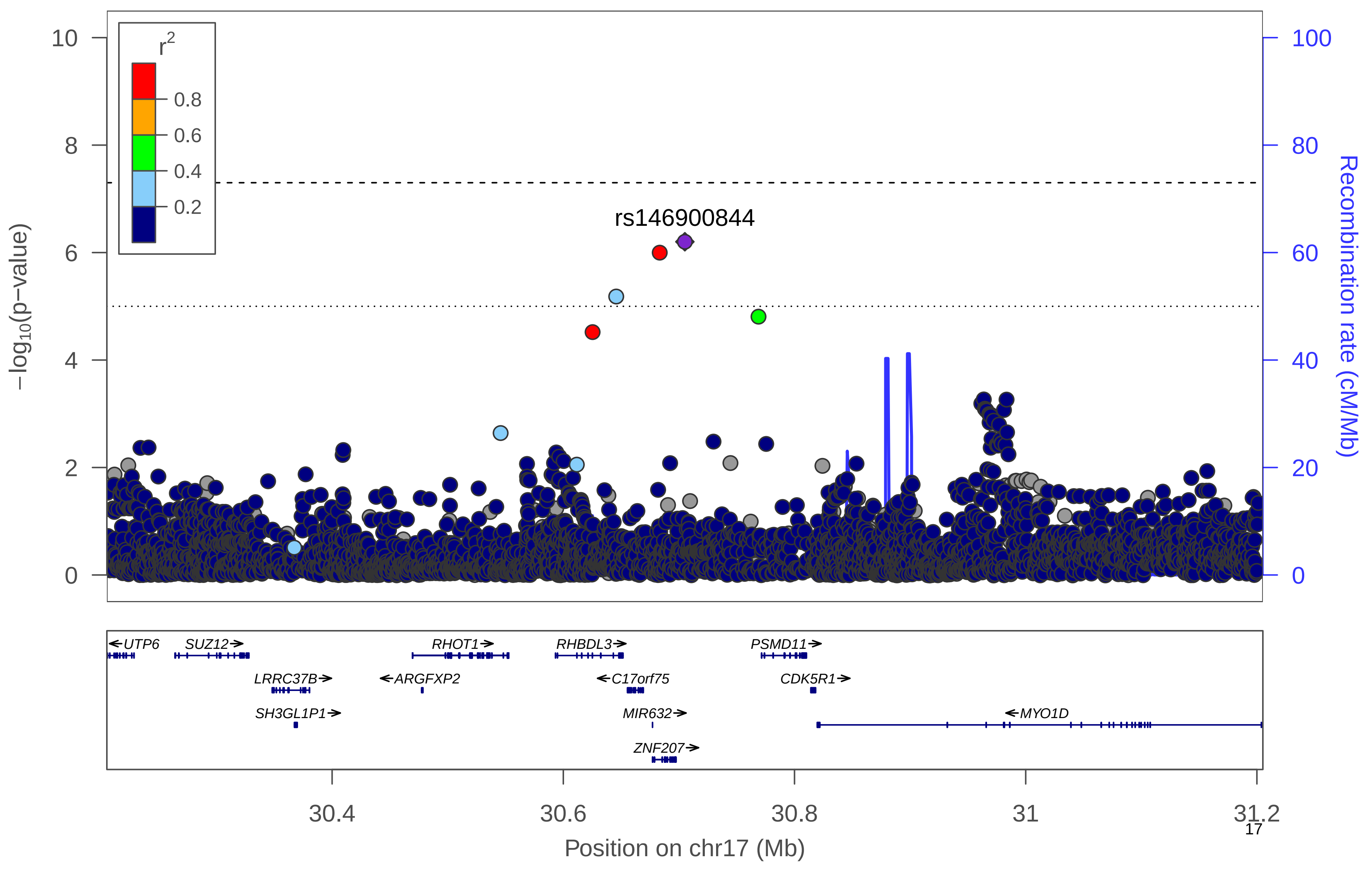

### h) WHRadjBMI Women

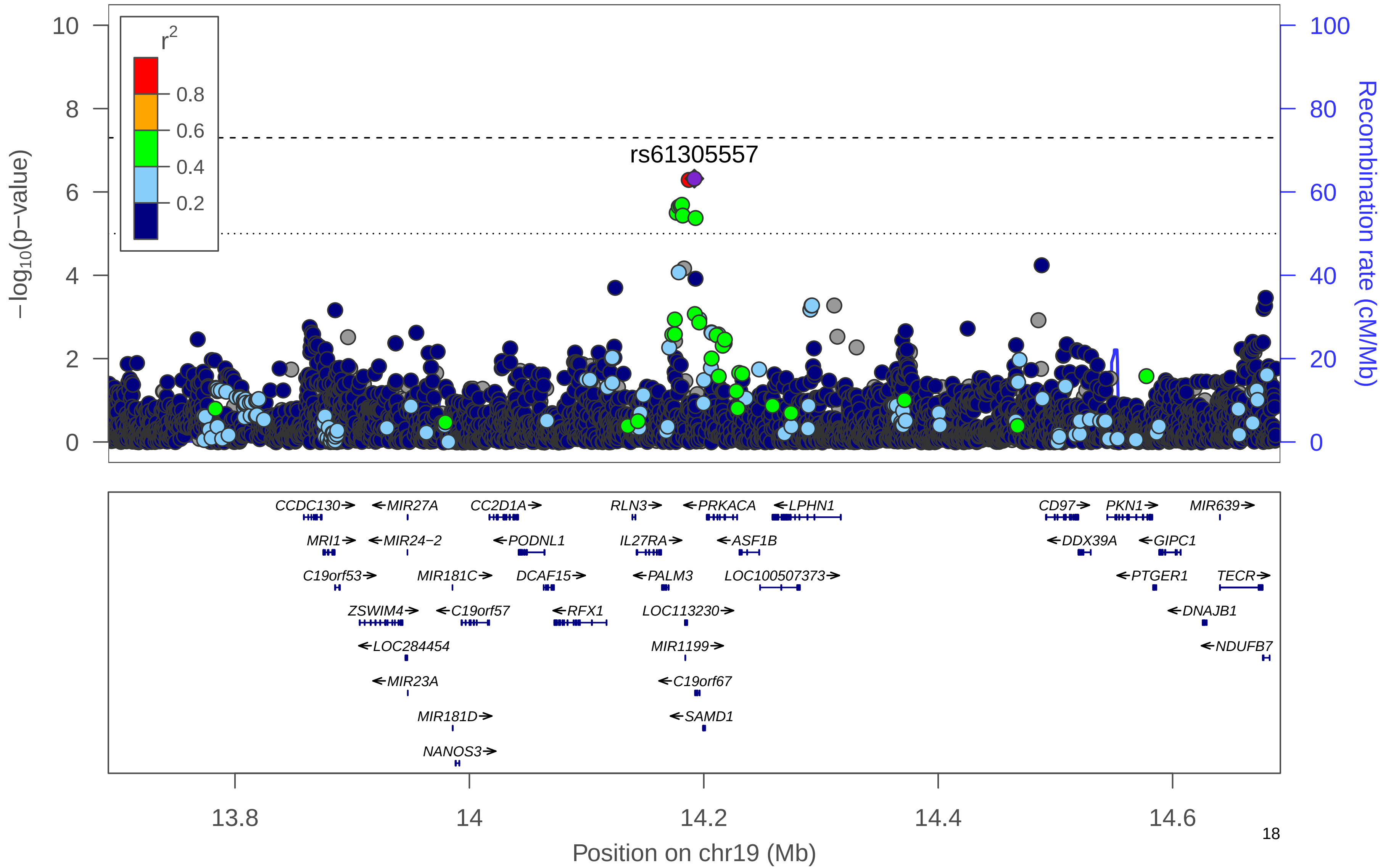

**Supplementary Figure 5. Manhattan plot.** Manhattan plot of the men-only analysis for WHRadjBMI. All suggestively significant (*P*<1x10$^{-6}$) variants are highlighted in orange if they are >500 Kb from any previously-reported WHRadjBMI associated variants. Previously reported loci (+/- 500 Kb) are highlighted in blue if any variant in the locus reached suggestive significance. All suggestively significant loci that meet our criteria for replication are annotated with the closest gene. †Replicated in African American meta-analysis. ‡ Replicated in Hispanic/Latino meta-analysis. ¥ Replicated in European American meta-analysis.

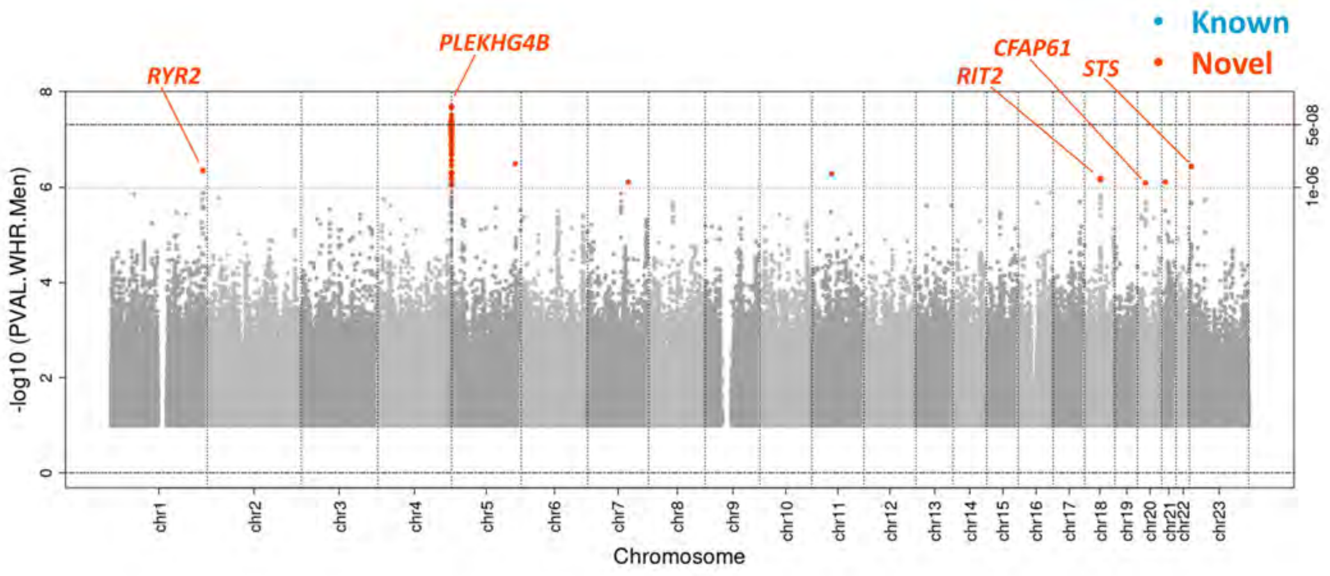

**Supplementary Figure 6. Locus Zoom Plots.** Regional association plots for suggestively significant loci in the HCHS/SOL WHRadjBMI men-only analysis. The plots appear in chromosome:position order. Dot color reflects $R^2$ calculated from the 1000 Genomes AMR reference dataset. Point symbols represent variant functional classifications: a) rs12032174, *RYR2*; b) rs10475310, *PLEKHG4B*; c) rs16977373, *RIT2*; d) rs721424, *CFAP61*; e) rs148213302, *STS*.

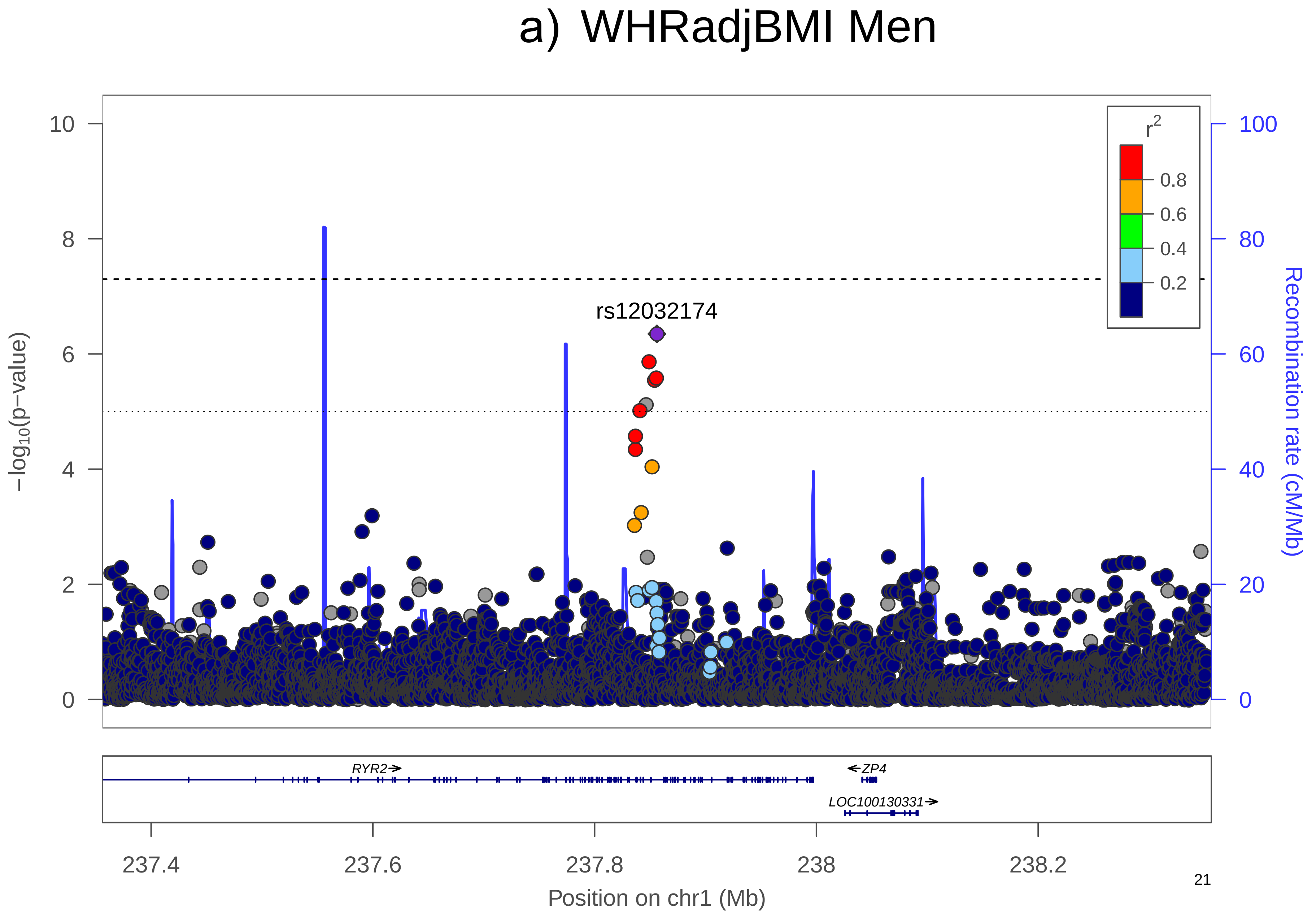
a) WHRadjBMI Men
−log10(p−value)
Recombination rate (cM/Mb)
rs12032174
r2
0.8
0.6
0.4
0.2
RYR2
ZP4
LOC100130331
237.4
237.6
237.8
238
238.2
Position on chr1 (Mb)

### b) WHRadjBMI Men

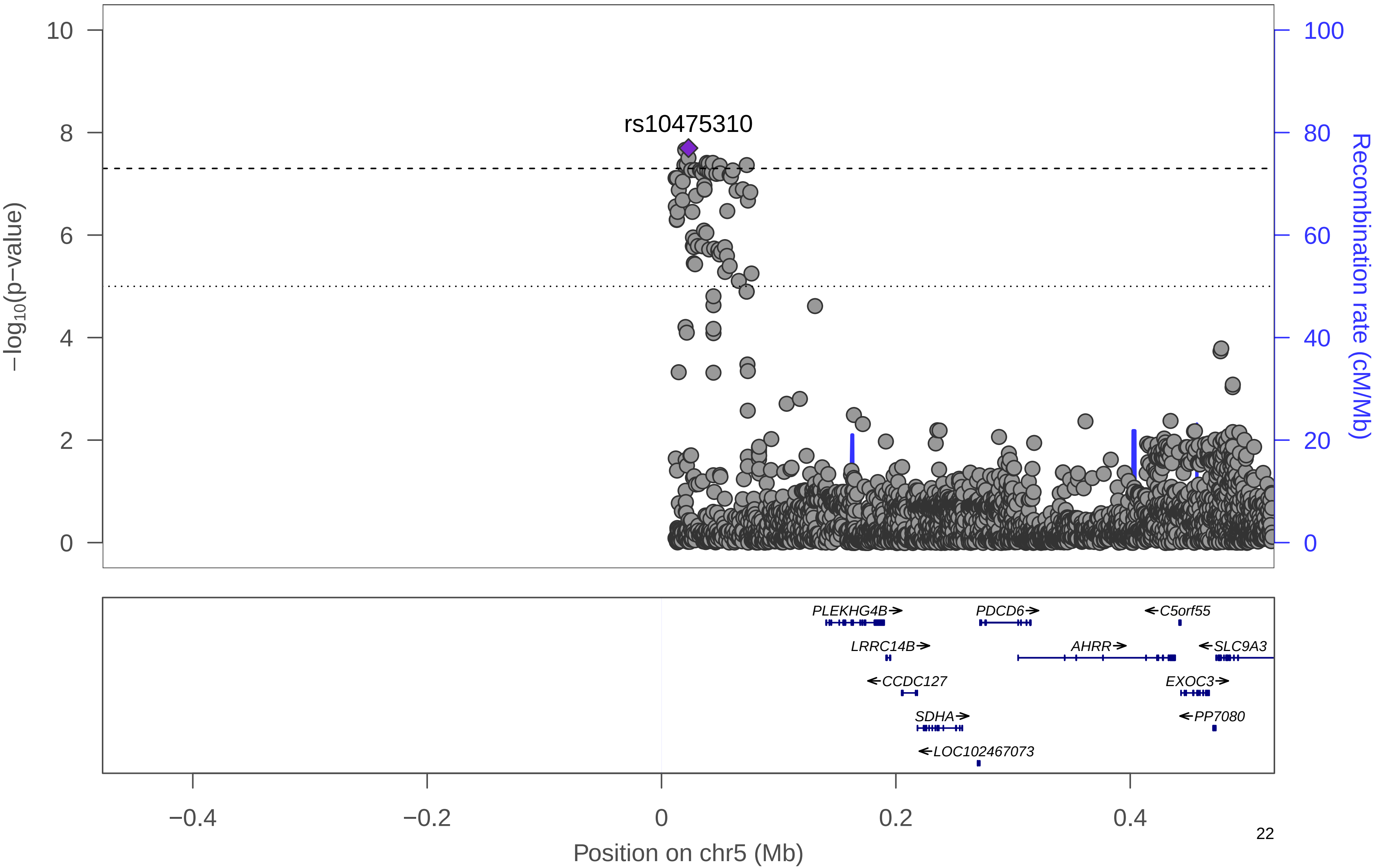

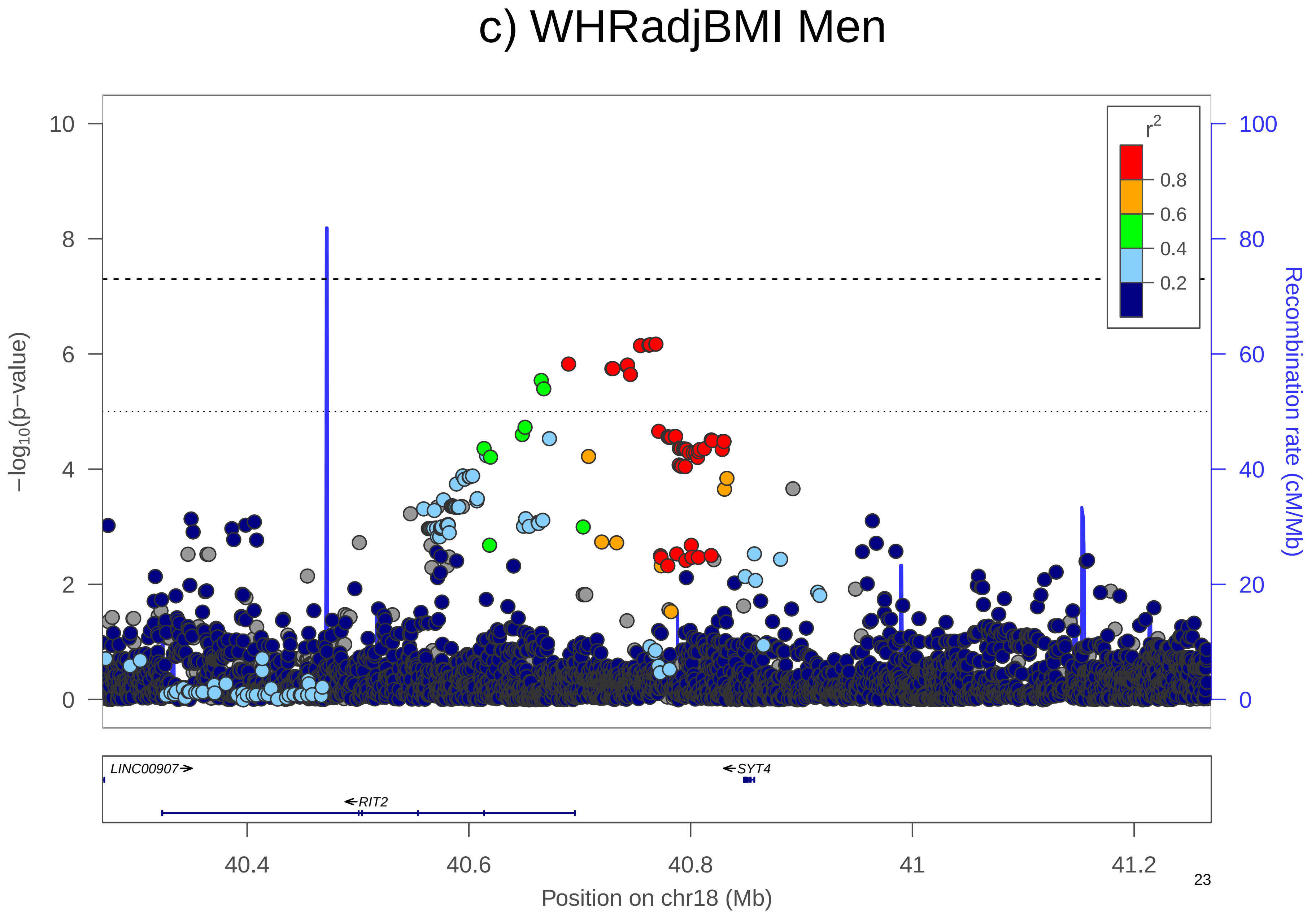

c) WHRadjBMI Men
−log10(p−value)
Recombination rate (cM/Mb)
r2
0.8
0.6
0.4
0.2
LINC00907
SYT4
RIT2
40.4
40.6
40.8
41
41.2
Position on chr18 (Mb)

### d) WHRadjBMI Men

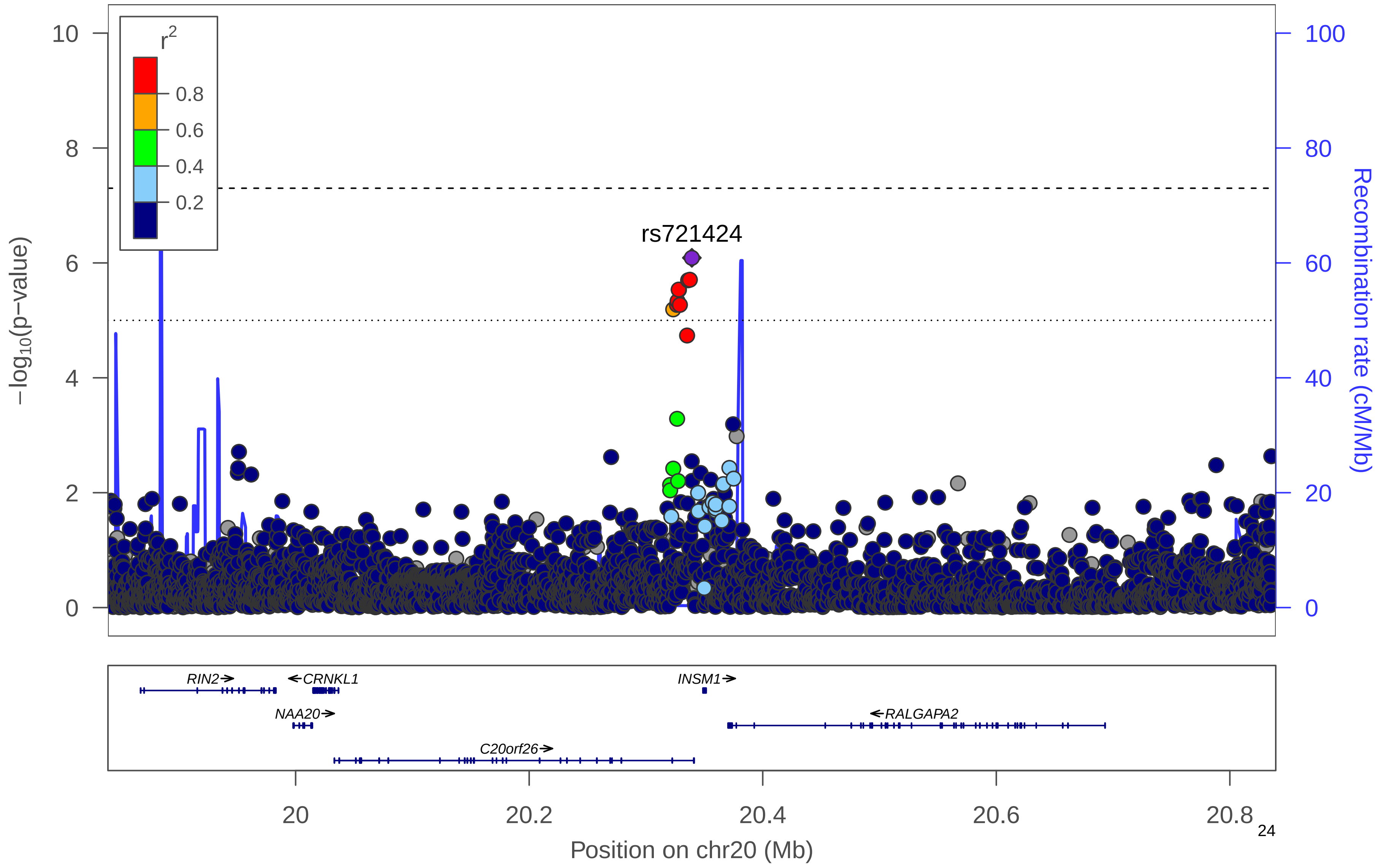

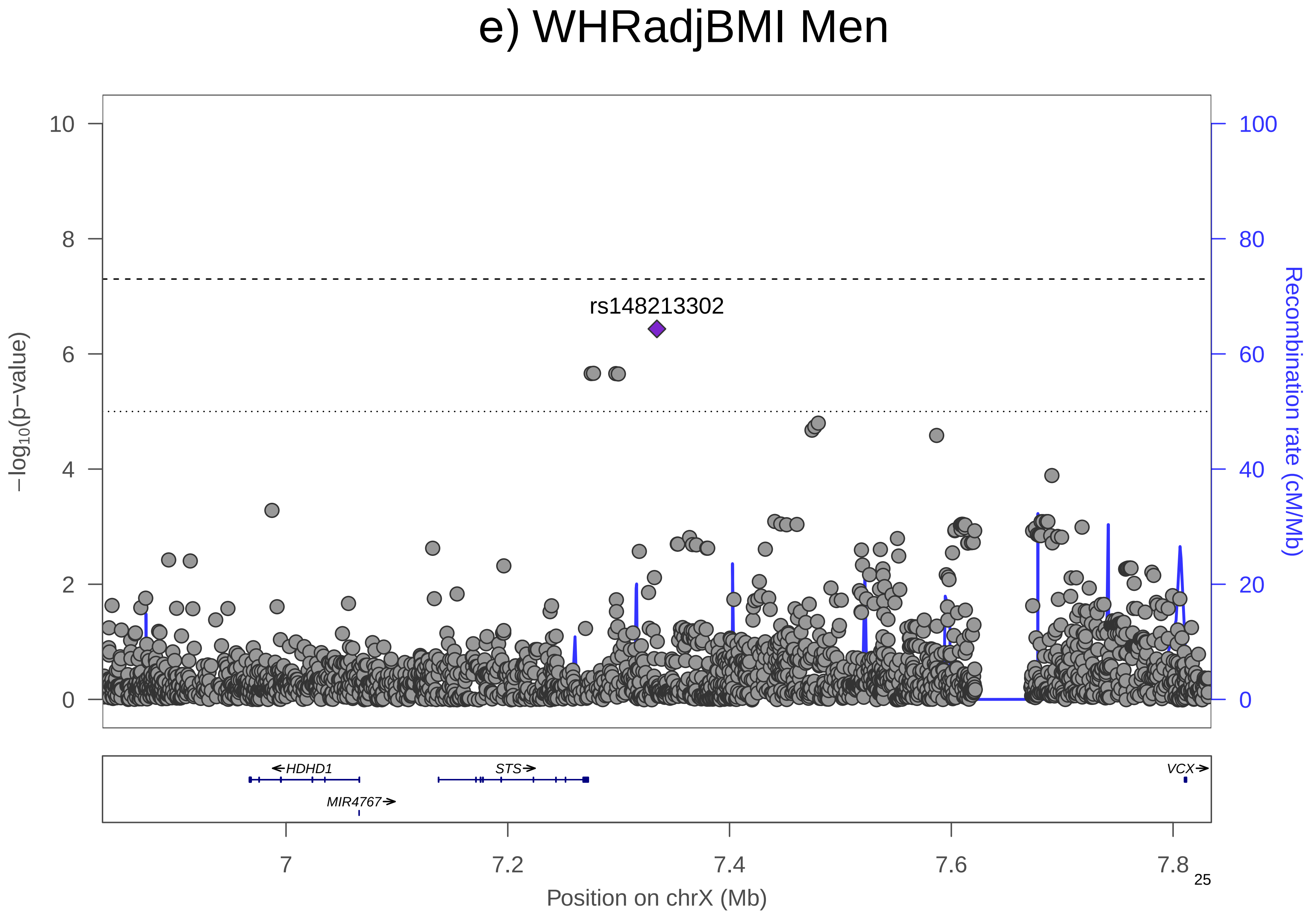

e) WHRadjBMI Men
rs148213302
−log10(p−value)
Recombination rate (cM/Mb)
0
2
4
6
8
10
0
20
40
60
80
100
HDHD1
STS
VCX
MIR4767
7
7.2
7.4
7.6
7.8
Position on chrX (Mb)

**Supplementary Figure 7. QQ Plots**. QQ Plots for WHRadjBMI, including sexes-combined (black), women-only (orange), men-only (blue).

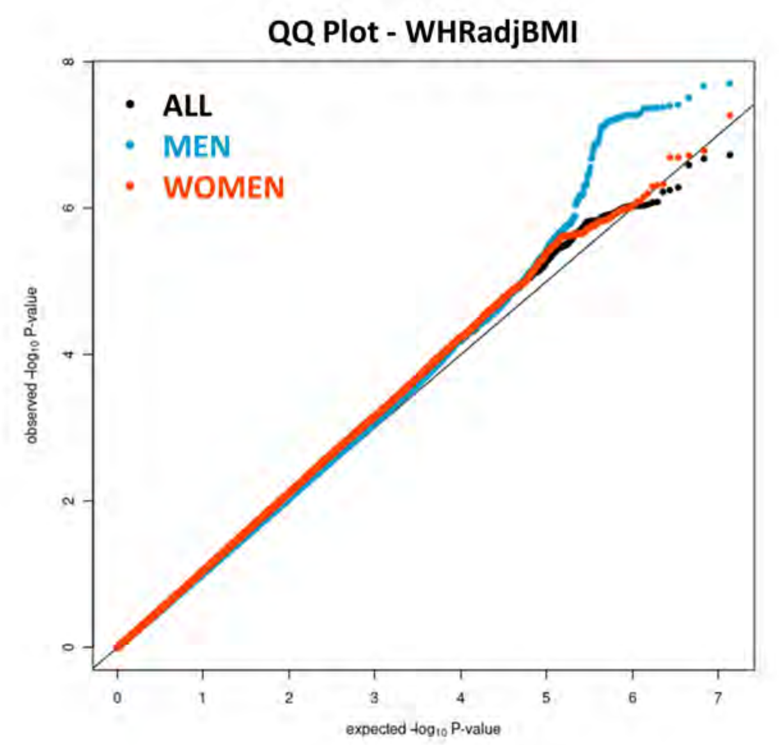

**Supplementary Figure 8. Manhattan plot.** Manhattan plot of the sexes-combined analysis for WCadjBMI. All suggestively significant ($P<1x10^{-6}$) variants are highlighted in orange if they are >500 Kb from any previously-reported WCadjBMI associated variants. Previously reported loci (+/- 500 Kb) are highlighted in blue if any variant in the locus reached suggestive significance. All suggestively significant loci that meet our criteria for replication are annotated with the closest gene. †Replicated in African American meta-analysis. ‡ Replicated in Hispanic/Latino meta-analysis. ¥ Replicated in European American meta-analysis.

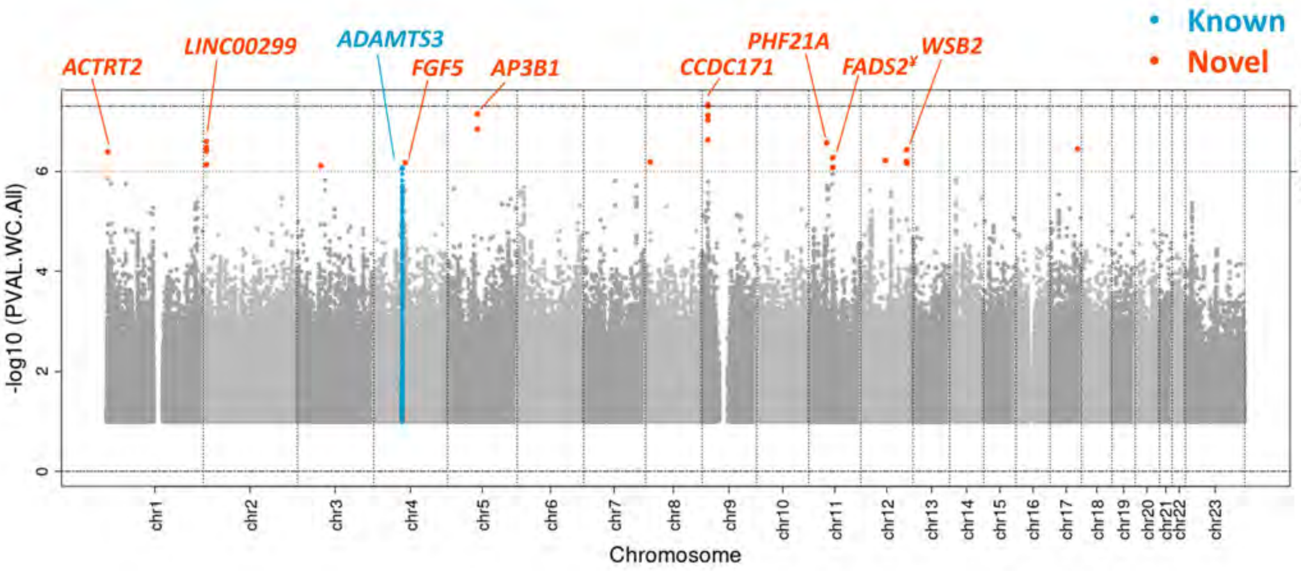

**Supplementary Figure 9. Locus Zoom Plots.** Regional association plots for suggestively significant loci in the HCHS/SOL WCadjBMI sexes-combined analysis. The plots appear in chromosome:position order. Dot color reflects $R^2$ calculated from the 1000 Genomes AMR reference dataset. Point symbols represent variant functional classifications: a) rs77993329, *ACTRT2*; b) rs138819965, *LINC00299*; c) rs77319470, *ADAMTS3*; d) rs921999, *FGF5*; e) rs112404395, *AP3B1*; f) rs77264633, *CCDC171*; g) rs184067184, *PHF21A*; h) rs3168072, *FADS2*; i) rs60260780, *WSB2*.

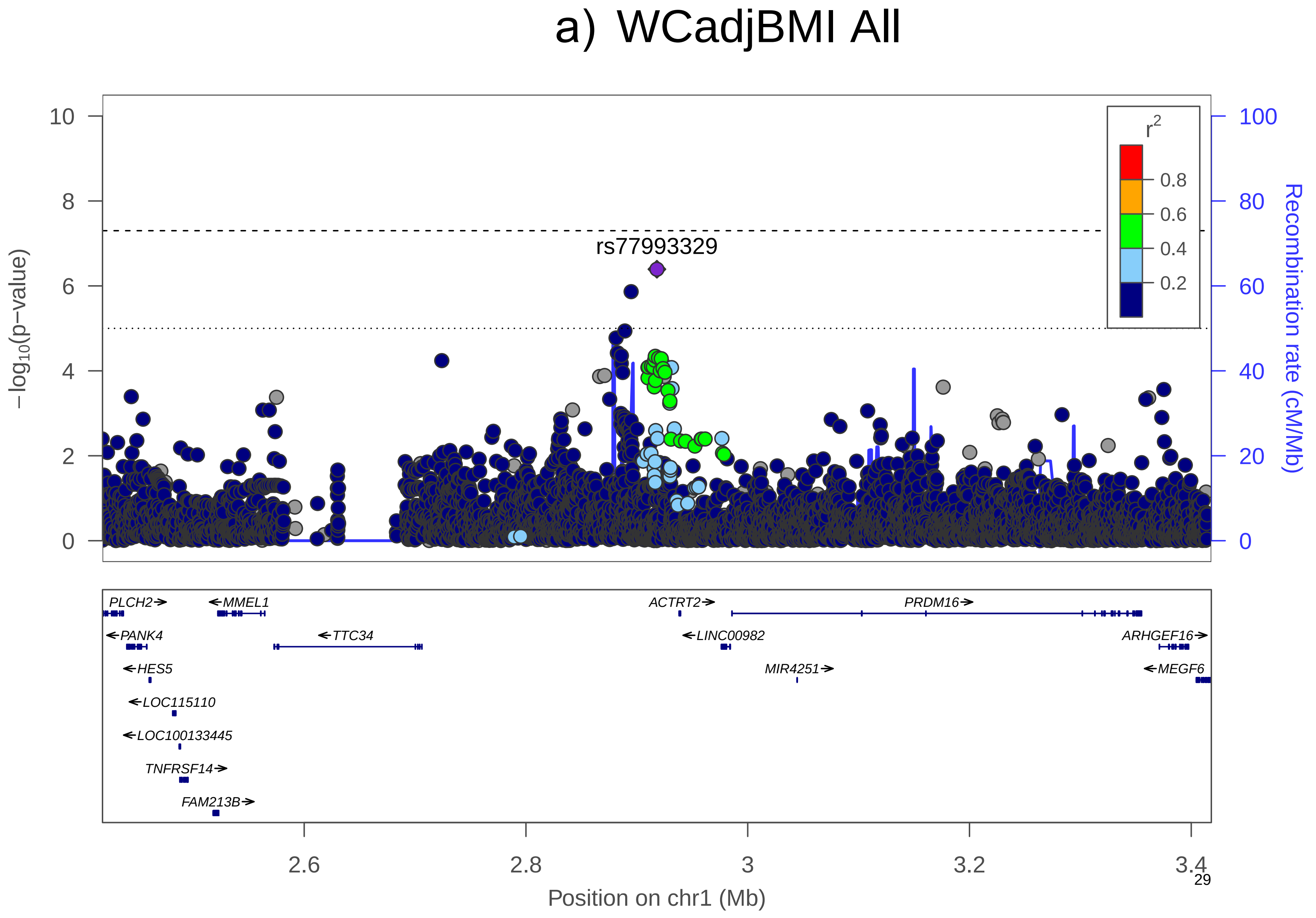
a) WCadjBMI All
rs77993329
−log10(p−value)
Recombination rate (cM/Mb)
r2
0.8
0.6
0.4
0.2
0
2
4
6
8
10
0
20
40
60
80
100
PLCH2
MMEL1
ACTRT2
PRDM16
PANK4
TTC34
LINC00982
ARHGEF16
HES5
MIR4251
MEGF6
LOC115110
LOC100133445
TNFRSF14
FAM213B
2.6
2.8
3
3.2
3.4
Position on chr1 (Mb)

b) WCadjBMI All

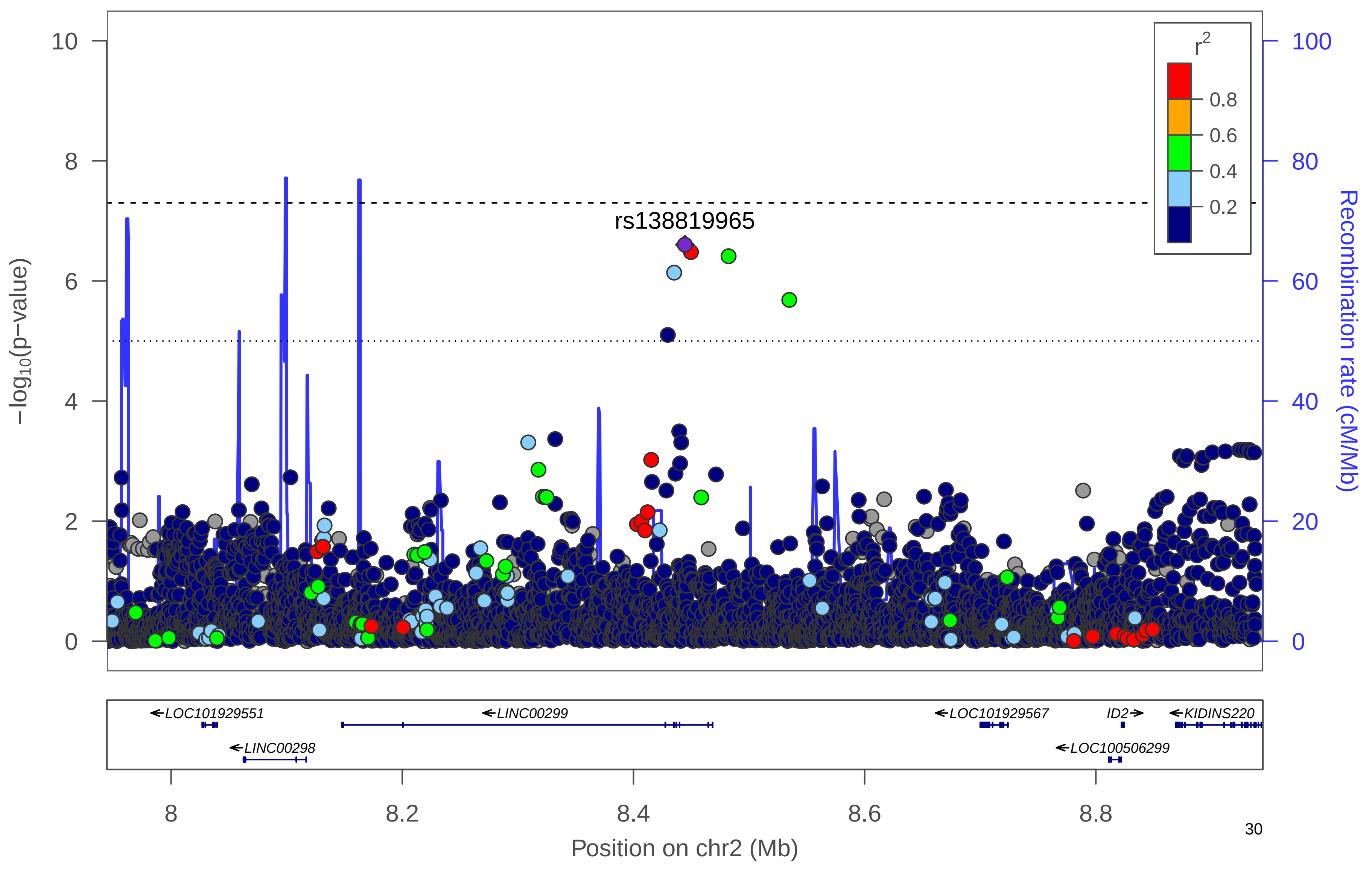

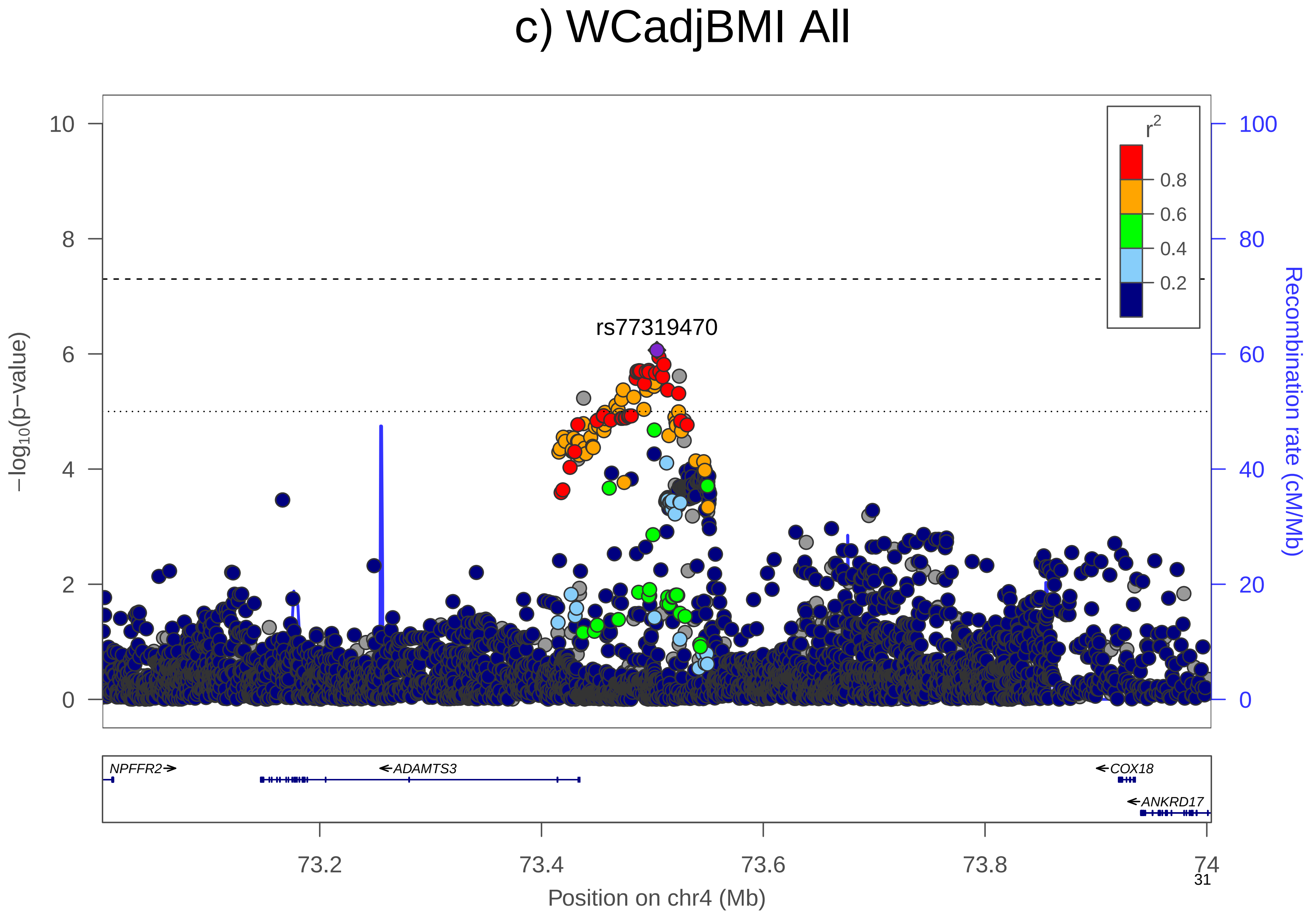

c) WCadjBMI All
rs77319470
r2
0.8
0.6
0.4
0.2
−log10(p−value)
Recombination rate (cM/Mb)
0
2
4
6
8
10
0
20
40
60
80
100
NPFFR2
ADAMTS3
COX18
ANKRD17
73.2
73.4
73.6
73.8
74
Position on chr4 (Mb)

d) WCadjBMI All

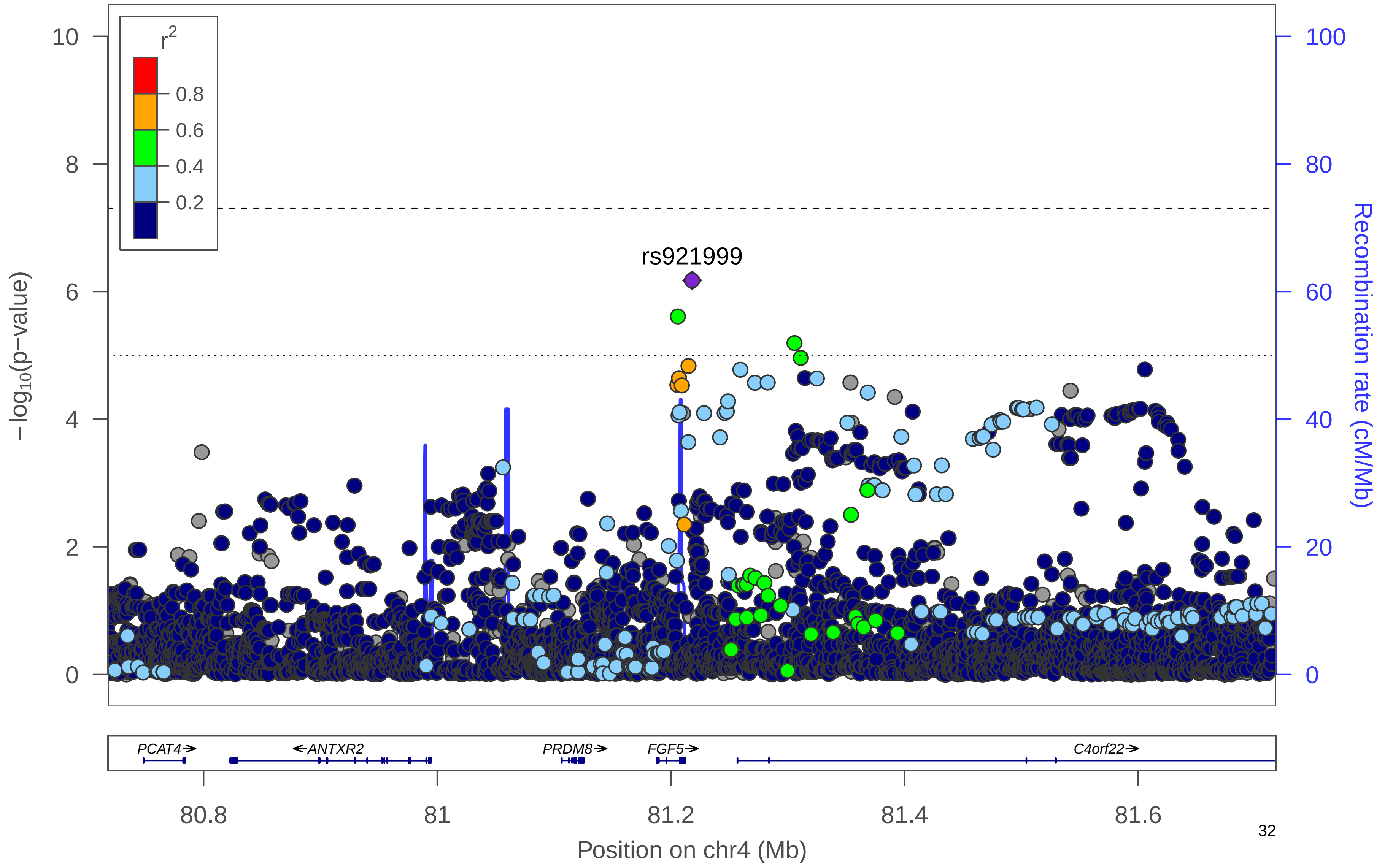

### e) WCadjBMI All

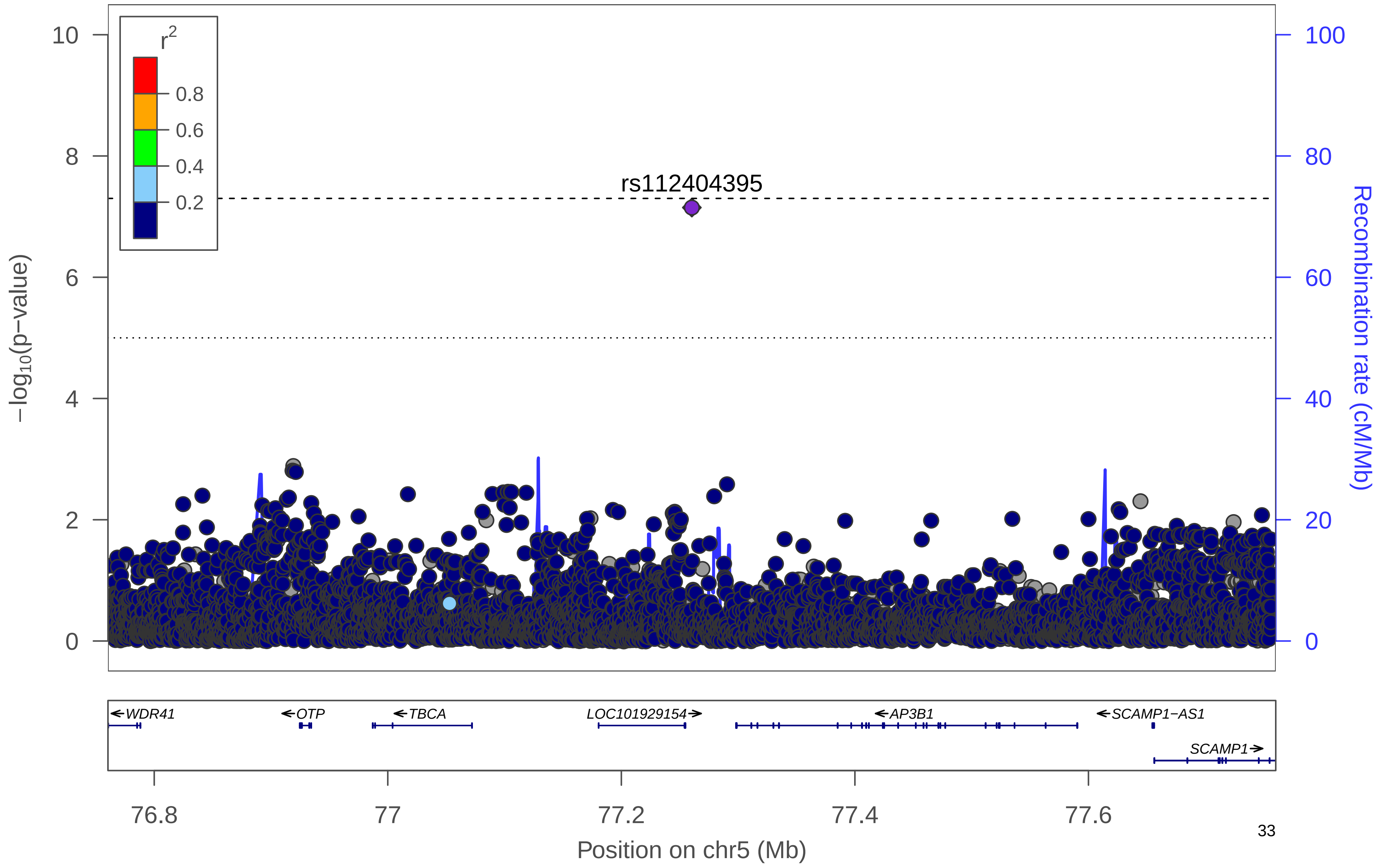

### f) WCadjBMI All

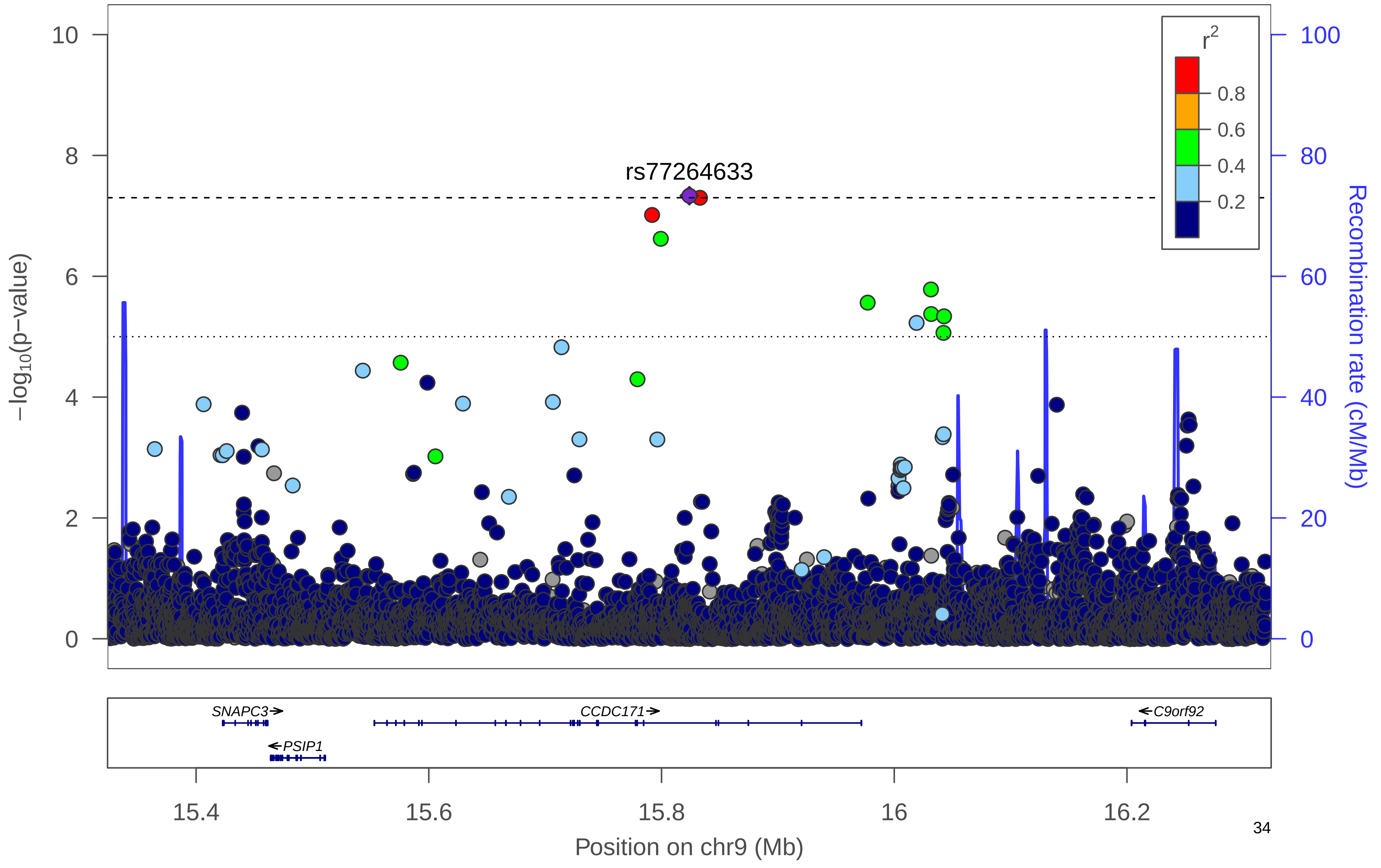

### g) WCadjBMI All

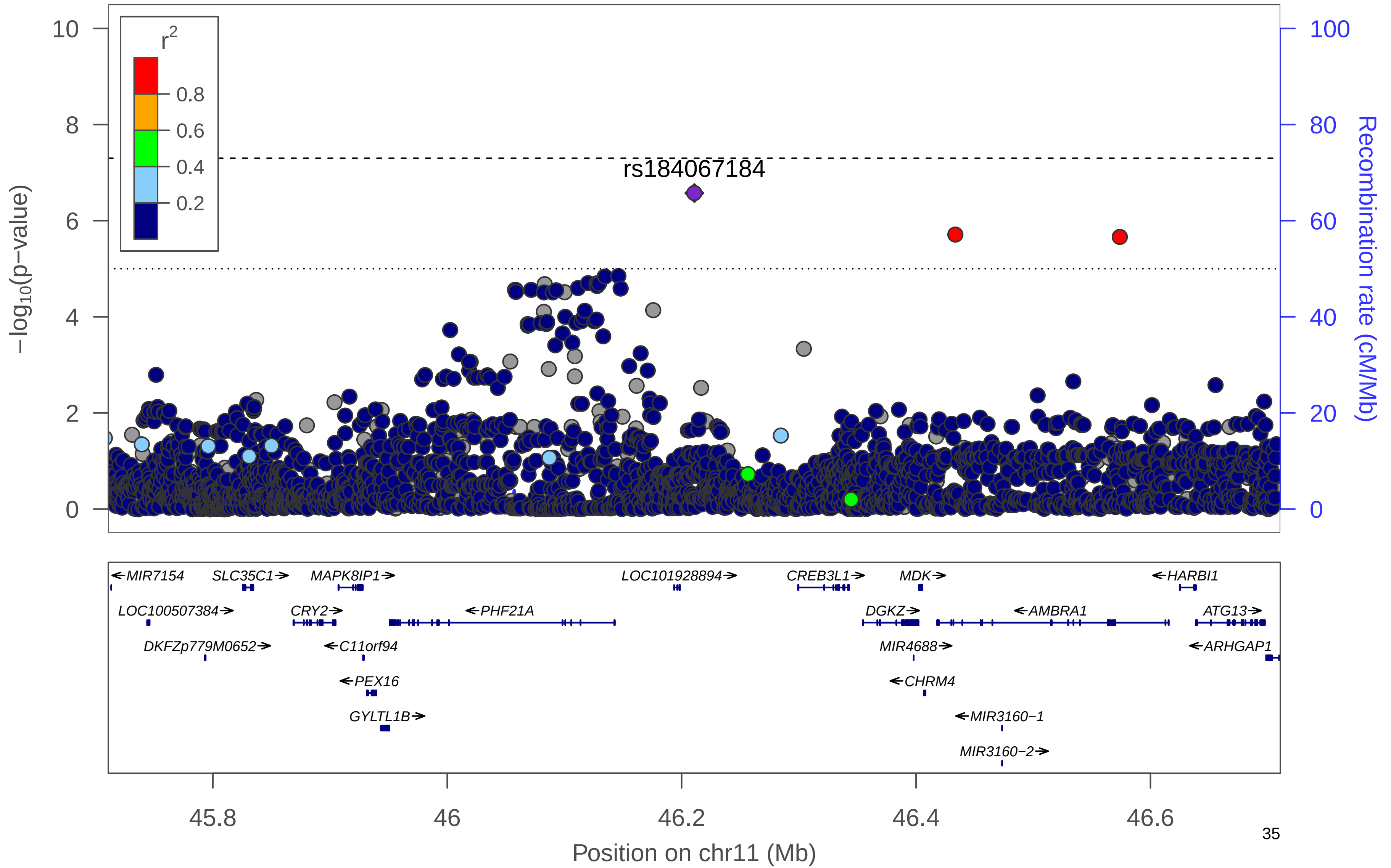

### h) WCadjBMI All

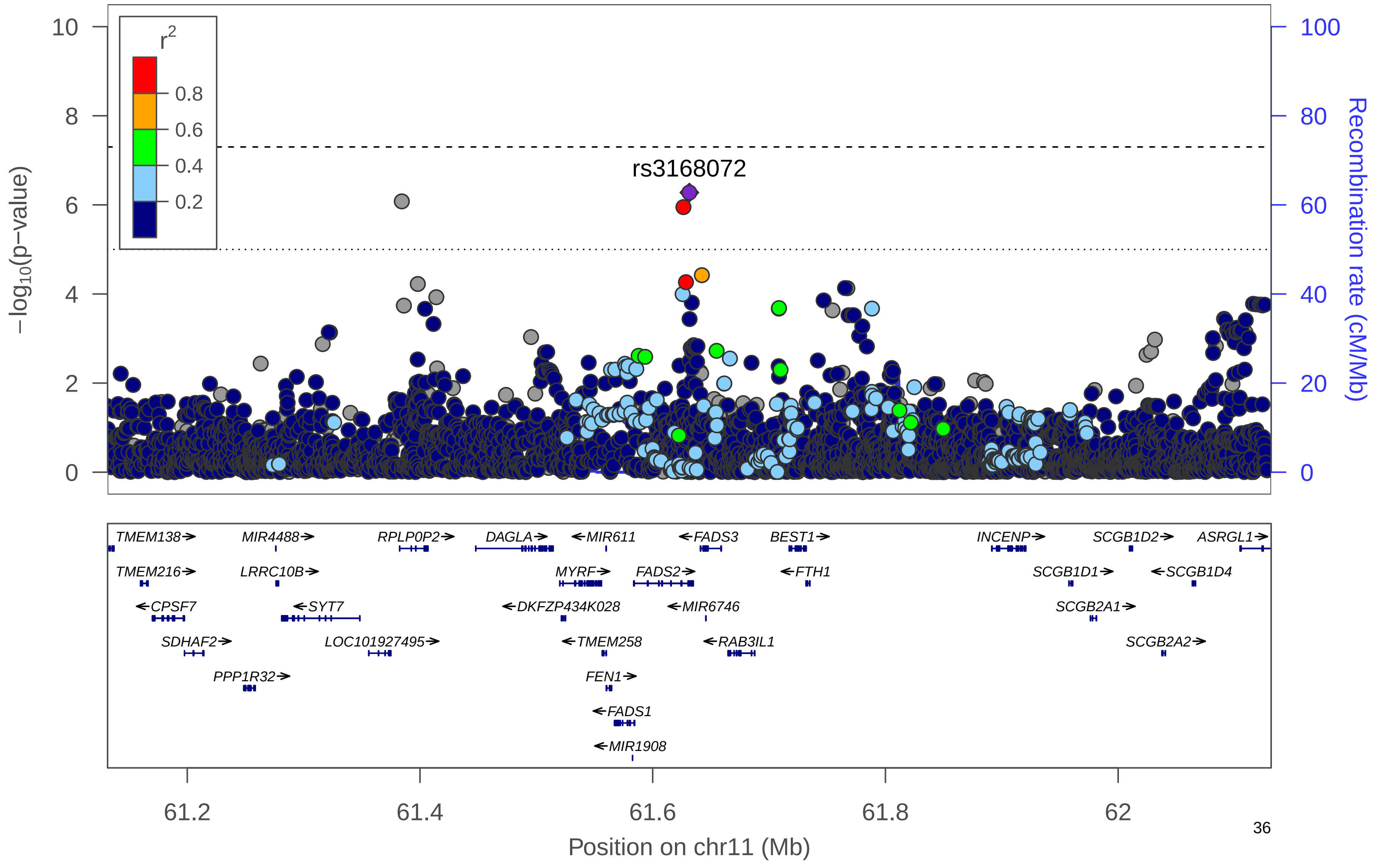

### i) WCadjBMI All

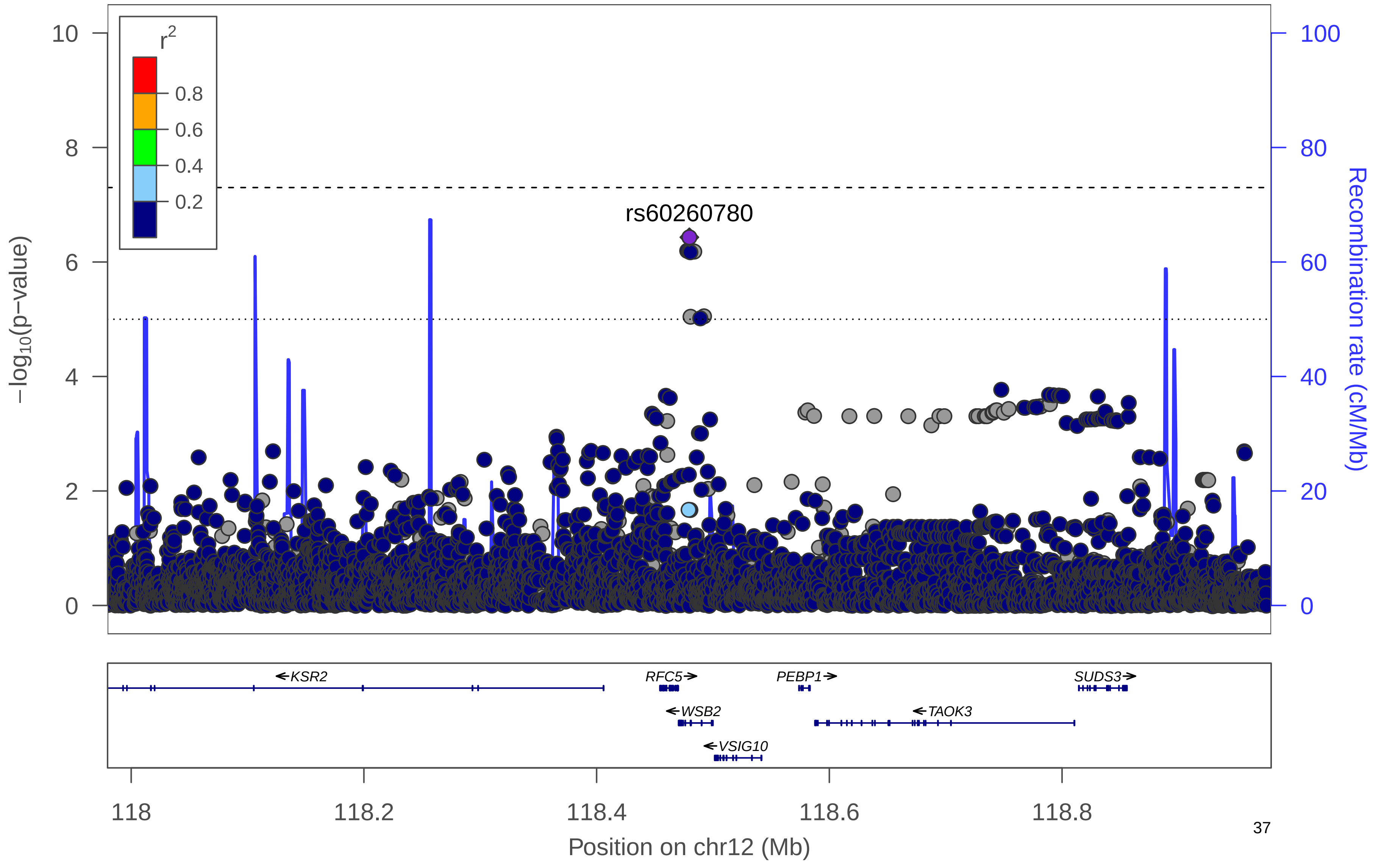

**Supplementary Figure 10. Manhattan plot.** Manhattan plot of the women-only analysis for WCadjBMI. All suggestively significant ($P<1x10^{-6}$) variants are highlighted in orange if they are >500 Kb from any previously-reported WCadjBMI associated variants. Previously reported loci (+/- 500 Kb) are highlighted in blue if any variant in the locus reached suggestive significance. All suggestively significant loci that meet our criteria for replication are annotated with the closest gene. †Replicated in African American meta-analysis. ‡ Replicated in Hispanic/Latino meta-analysis. ¥ Replicated in European American meta-analysis.

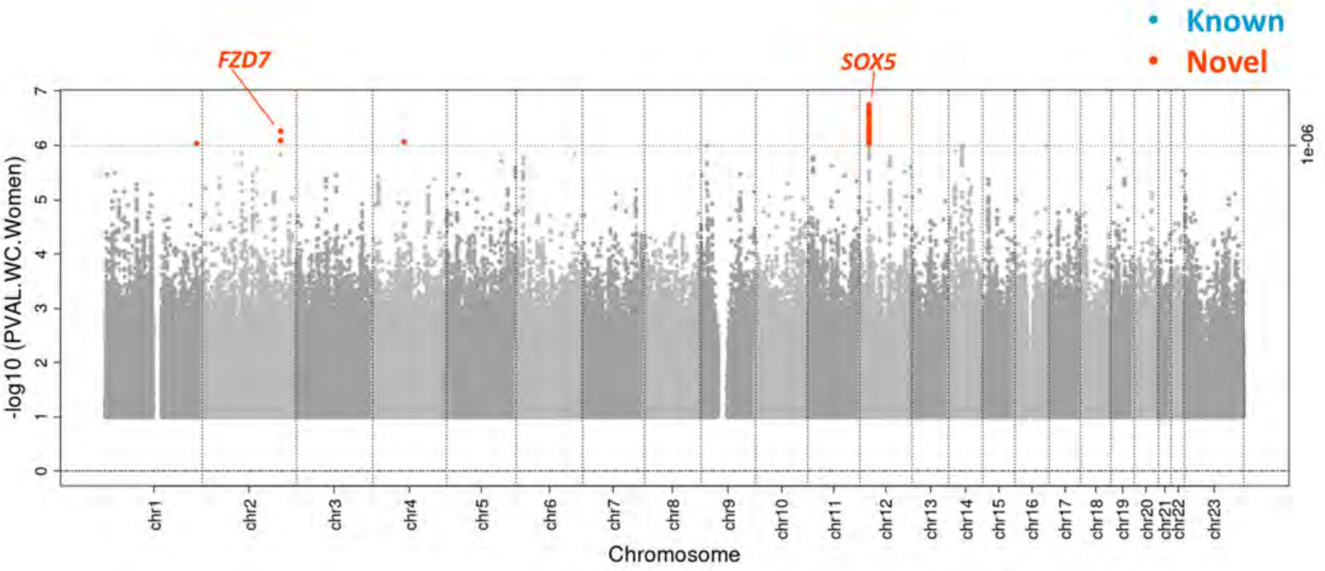

**Supplementary Figure 11. Locus Zoom Plots.** Regional association plots for suggestively significant loci in the HCHS/SOL WCadjBMI women-only analysis. The plots appear in chromosome:position order. Dot color reflects $R^2$ calculated from the 1000 Genomes AMR reference dataset. Point symbols represent variant functional classifications: a) rs112469617, *FZD7*; b) rs17385466, *SOX5*.

### a) WCadjBMI Women

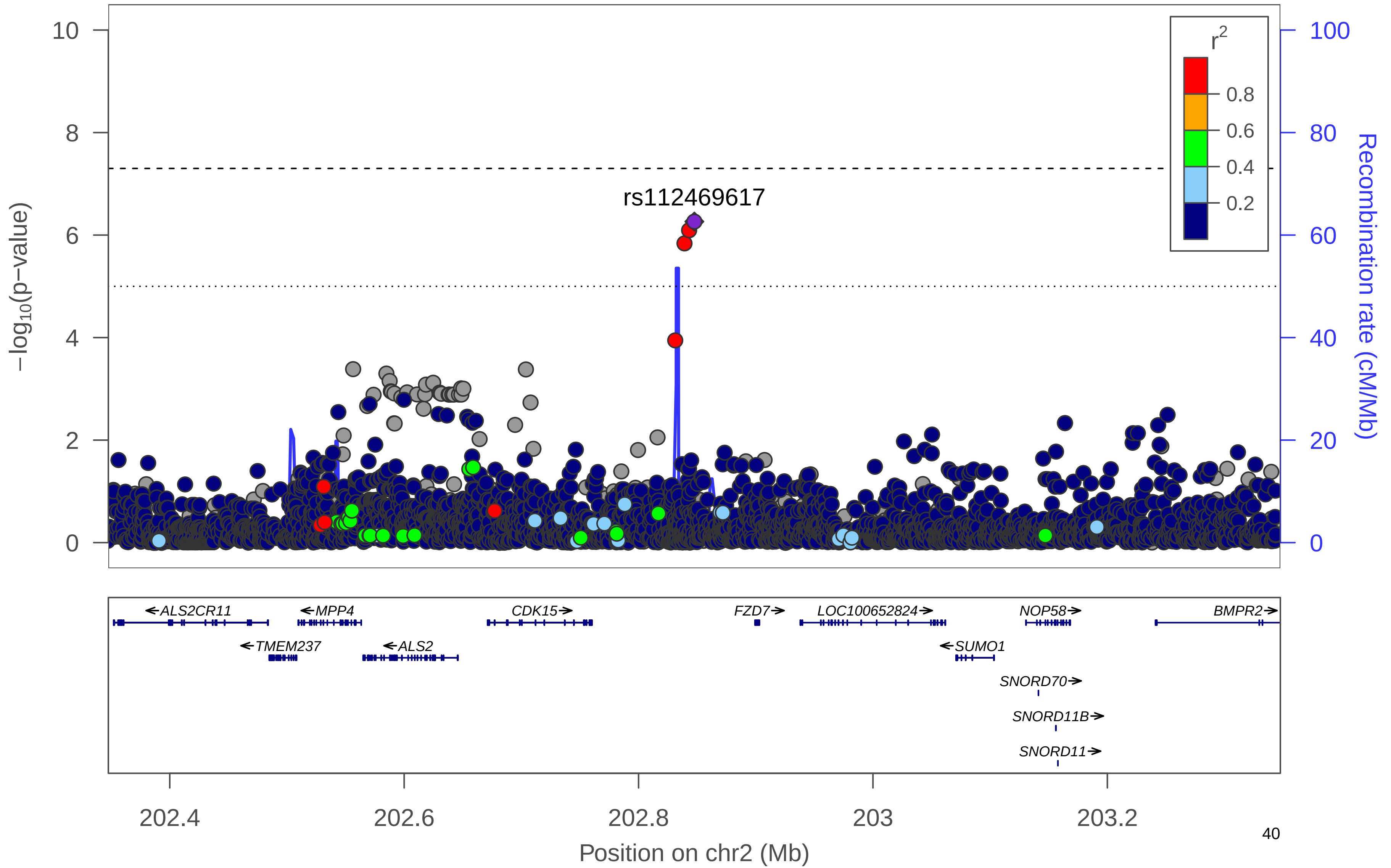

### b) WCadjBMI Women

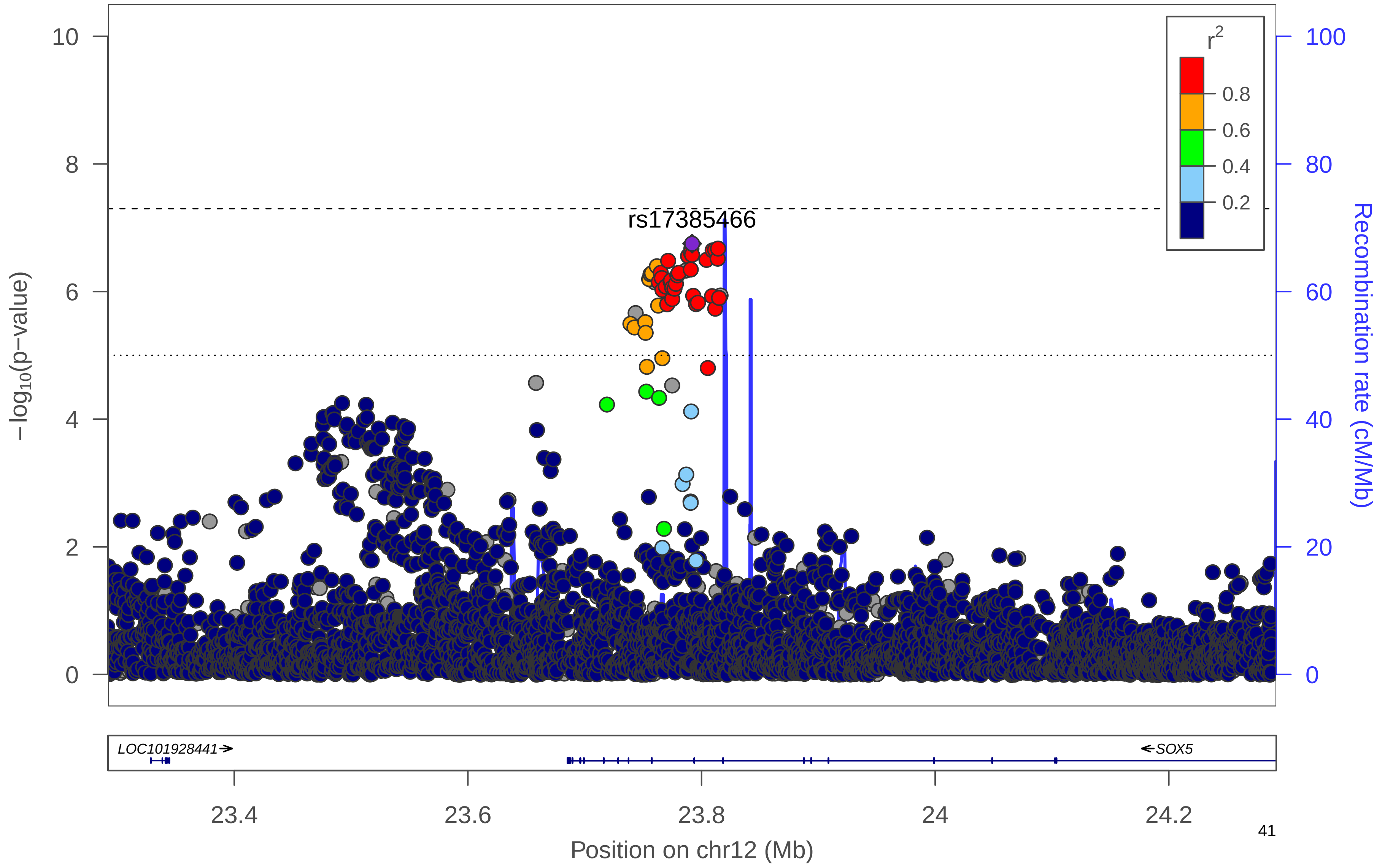

**Supplementary Figure 12. Manhattan plot.** Manhattan plot of the men-only analysis for WCadjBMI. All suggestively significant (*P*<1x10$^{-6}$) variants are highlighted in orange if they are >500 Kb from any previously-reported WCadjBMI associated variants. Previously reported loci (+/- 500 Kb) are highlighted in blue if any variant in the locus reached suggestive significance. All suggestively significant loci that meet our criteria for replication are annotated with the closest gene. †Replicated in African American meta-analysis. ‡ Replicated in Hispanic/Latino meta-analysis. ¥ Replicated in European American meta-analysis.

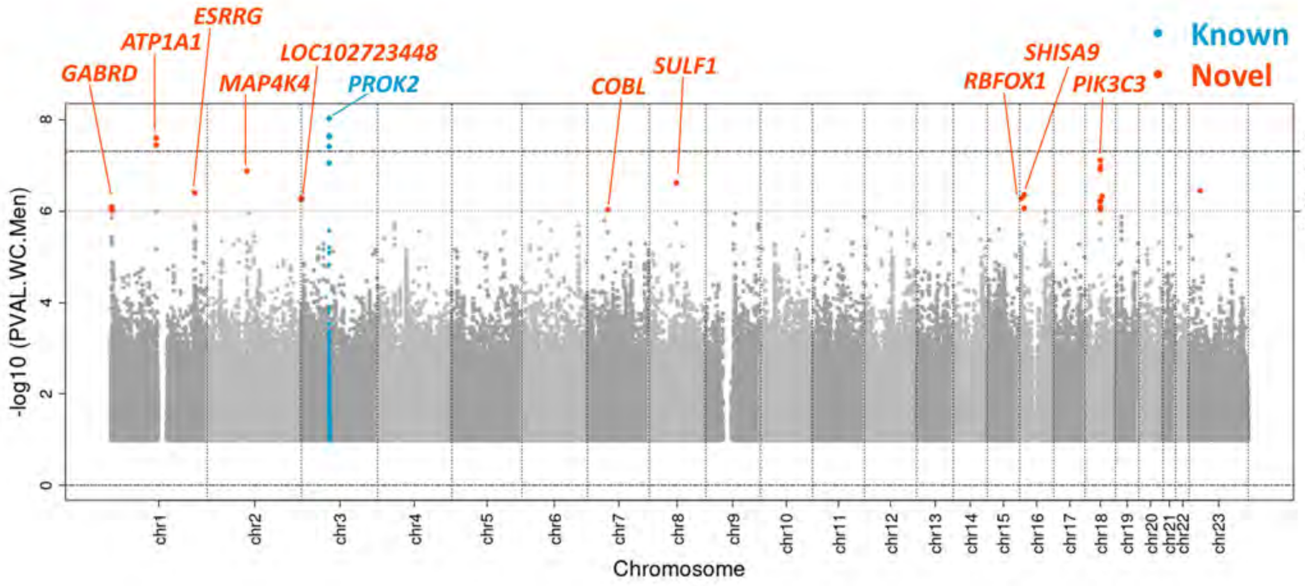

**Supplementary Figure 13. Locus Zoom Plots.** Regional association plots for suggestively significant loci in the HCHS/SOL WCadjBMI men-only analysis. The plots appear in chromosome:position order. Dot color reflects $R^2$ calculated from the 1000 Genomes AMR reference dataset. Point symbols represent variant functional classifications: a) rs74346221, *GABRD*; b) rs72693785, *ATP1A1*; c) rs11583298, *ESRRG*; d) rs76842062, *MAP4K4*; e) rs141365360, *LOC102723448*; f) rs6809759, *PROK2*; g) rs76941364, *COBL*; h) rs139139519, *SULF1*; i) rs35569658, *RBFOX1*; j) rs148280037, *SHISA9*; k) rs143565319, *PIK3C3*.

### a) WCadjBMI Men

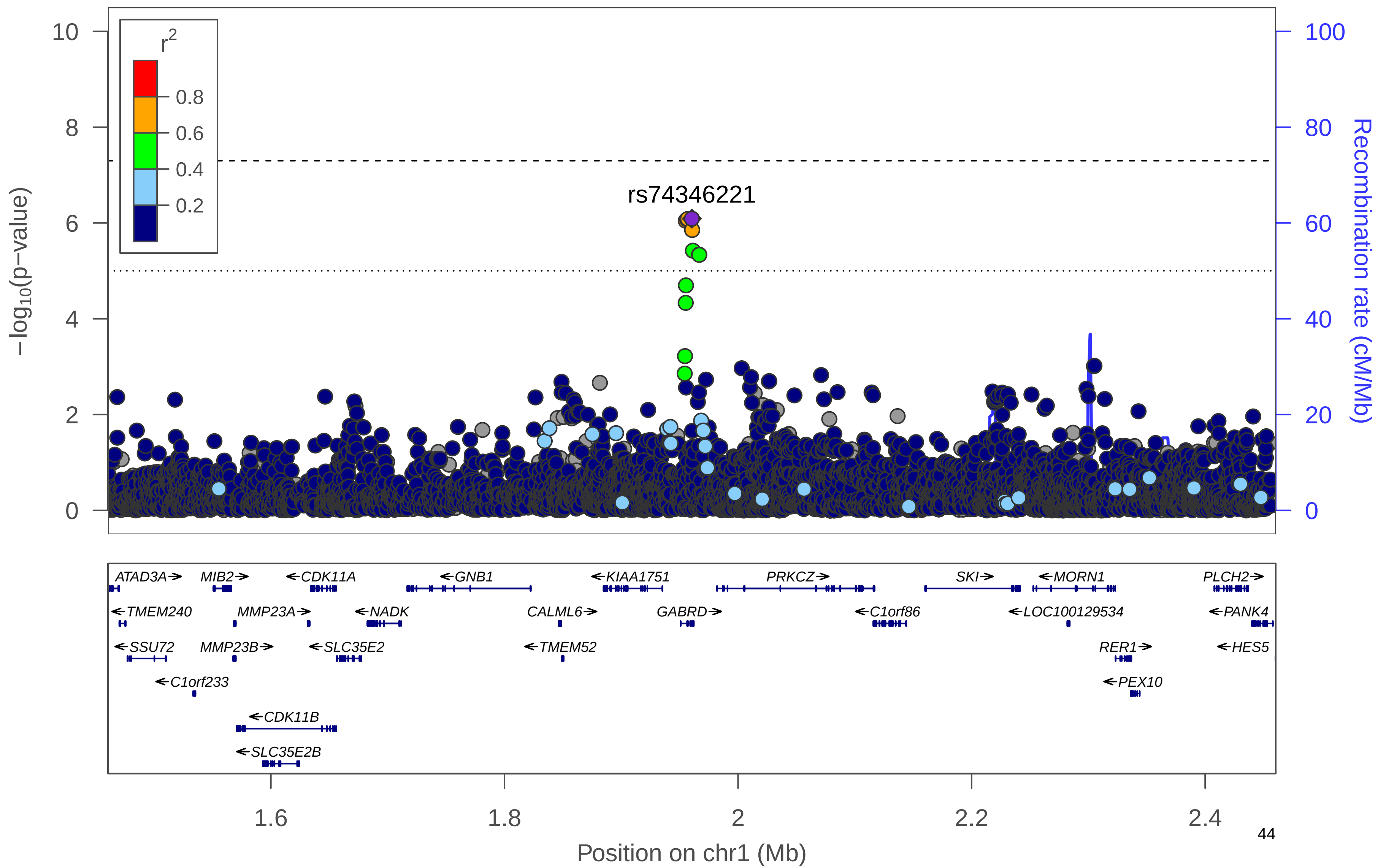

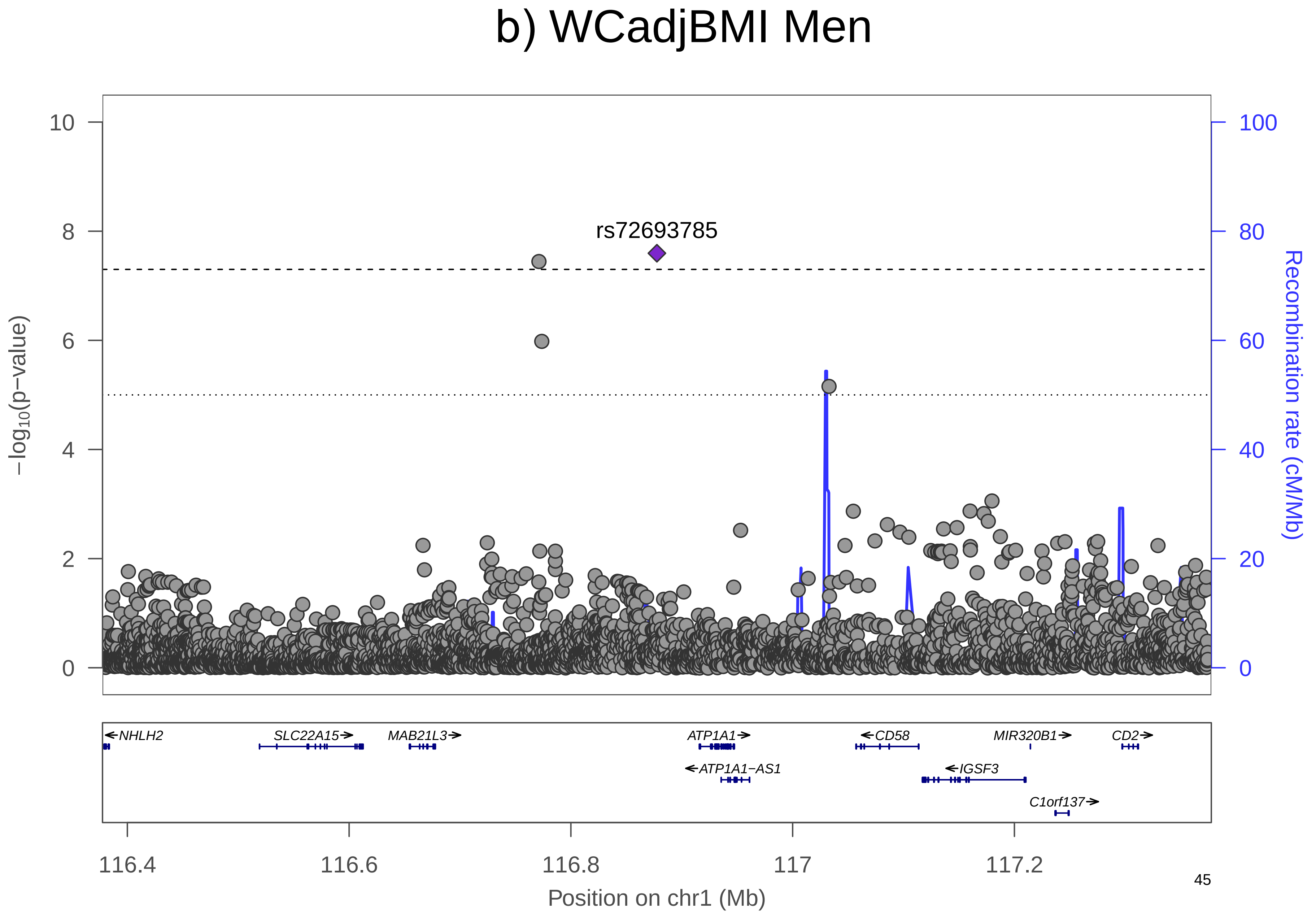
b) WCadjBMI Men
rs72693785
−log10(p−value)
Recombination rate (cM/Mb)
0
2
4
6
8
10
0
20
40
60
80
100
NHLH2
SLC22A15
MAB21L3
ATP1A1
ATP1A1−AS1
CD58
IGSF3
MIR320B1
C1orf137
CD2
116.4
116.6
116.8
117
117.2
Position on chr1 (Mb)

### c) WCadjBMI Men

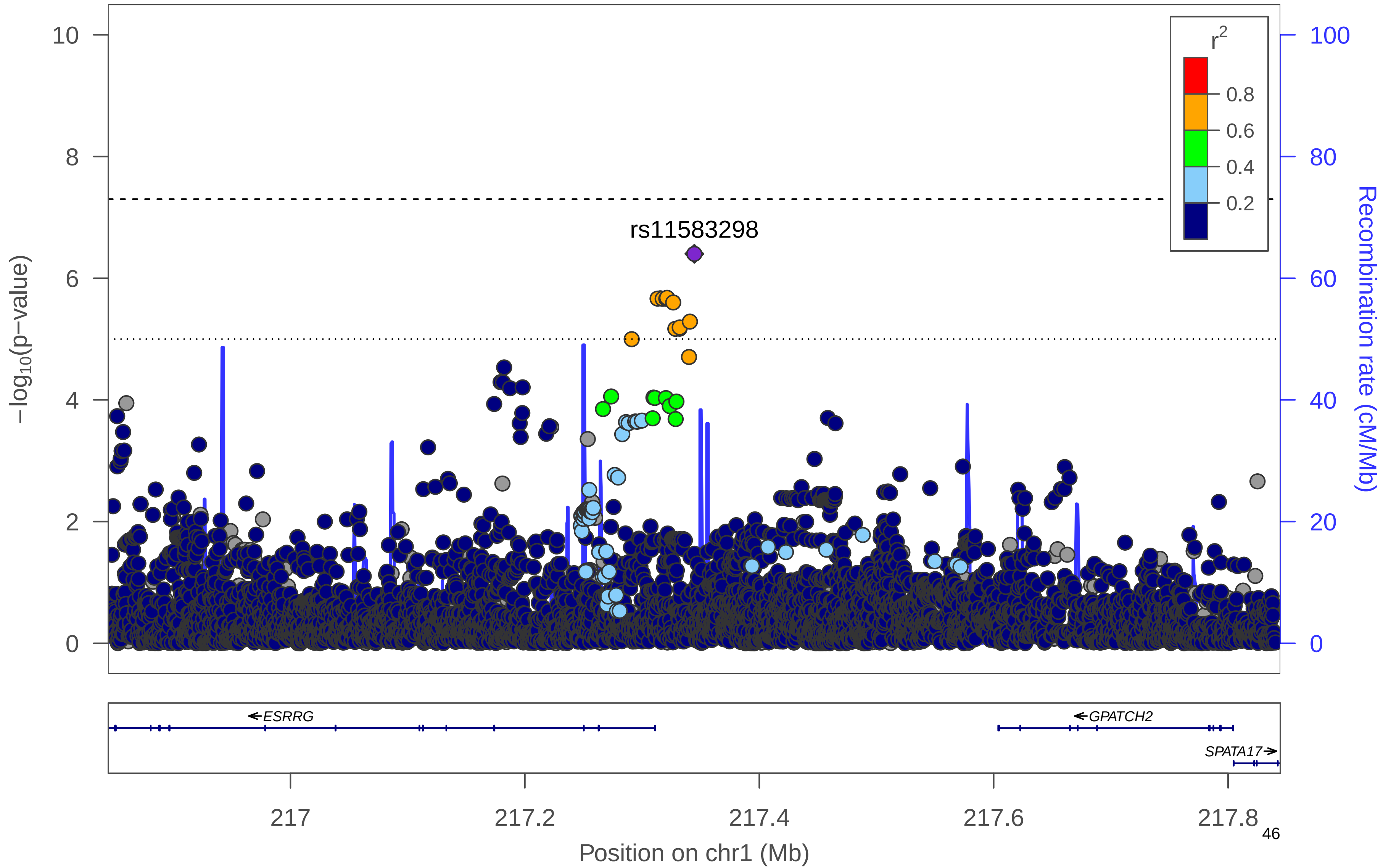

d) WCadjBMI Men

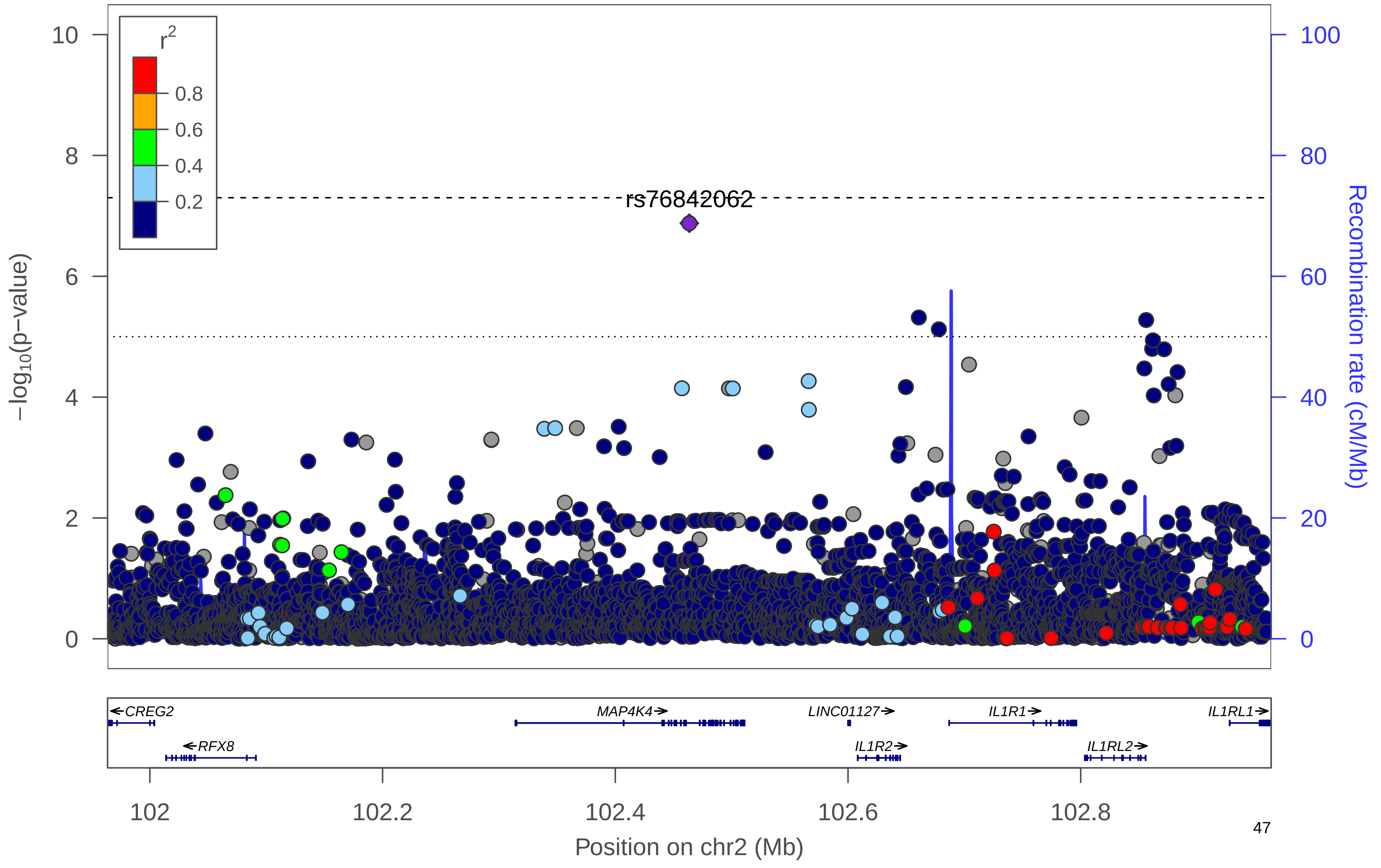

### e) WCadjBMI Men

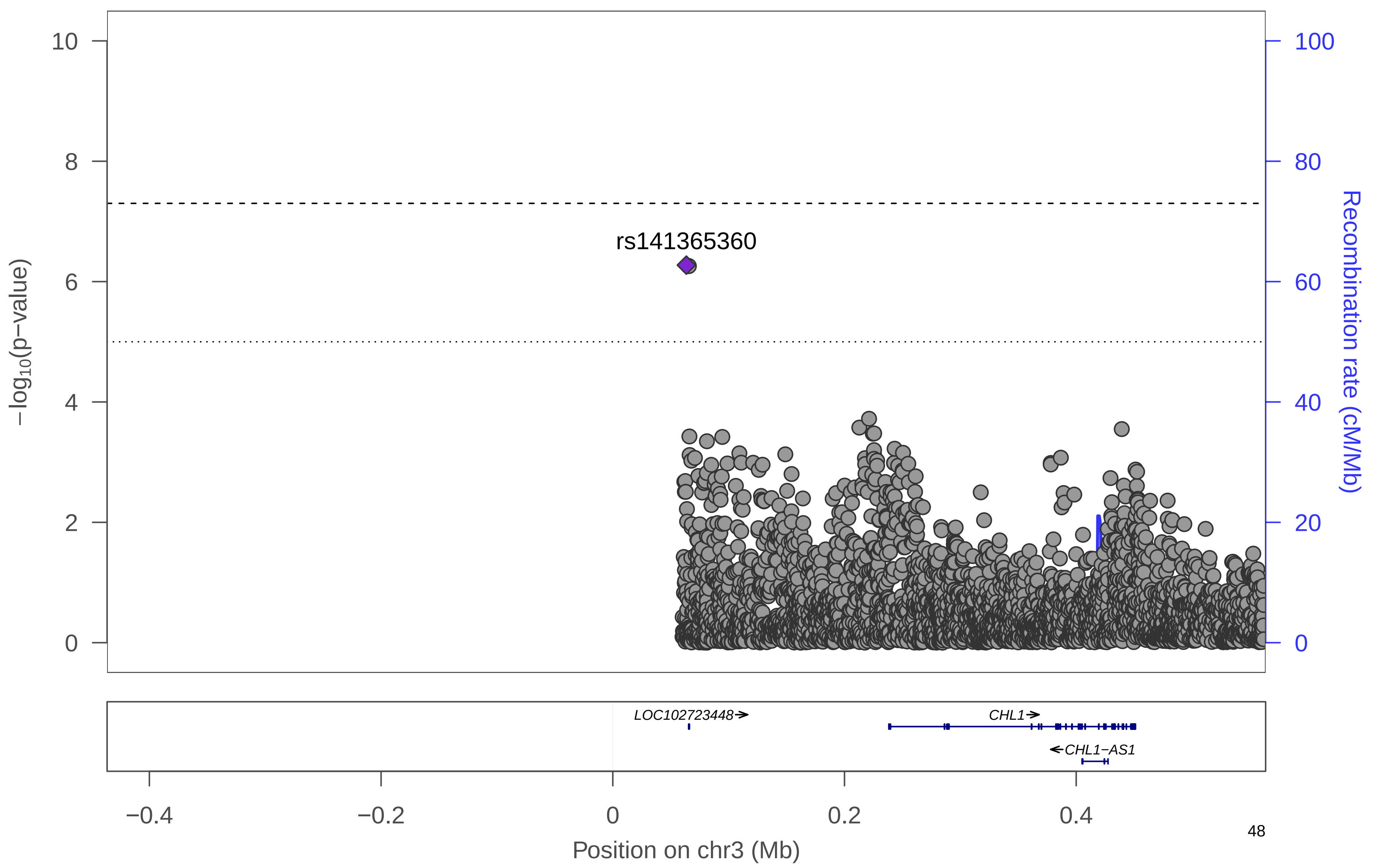

### f) WCadjBMI Men

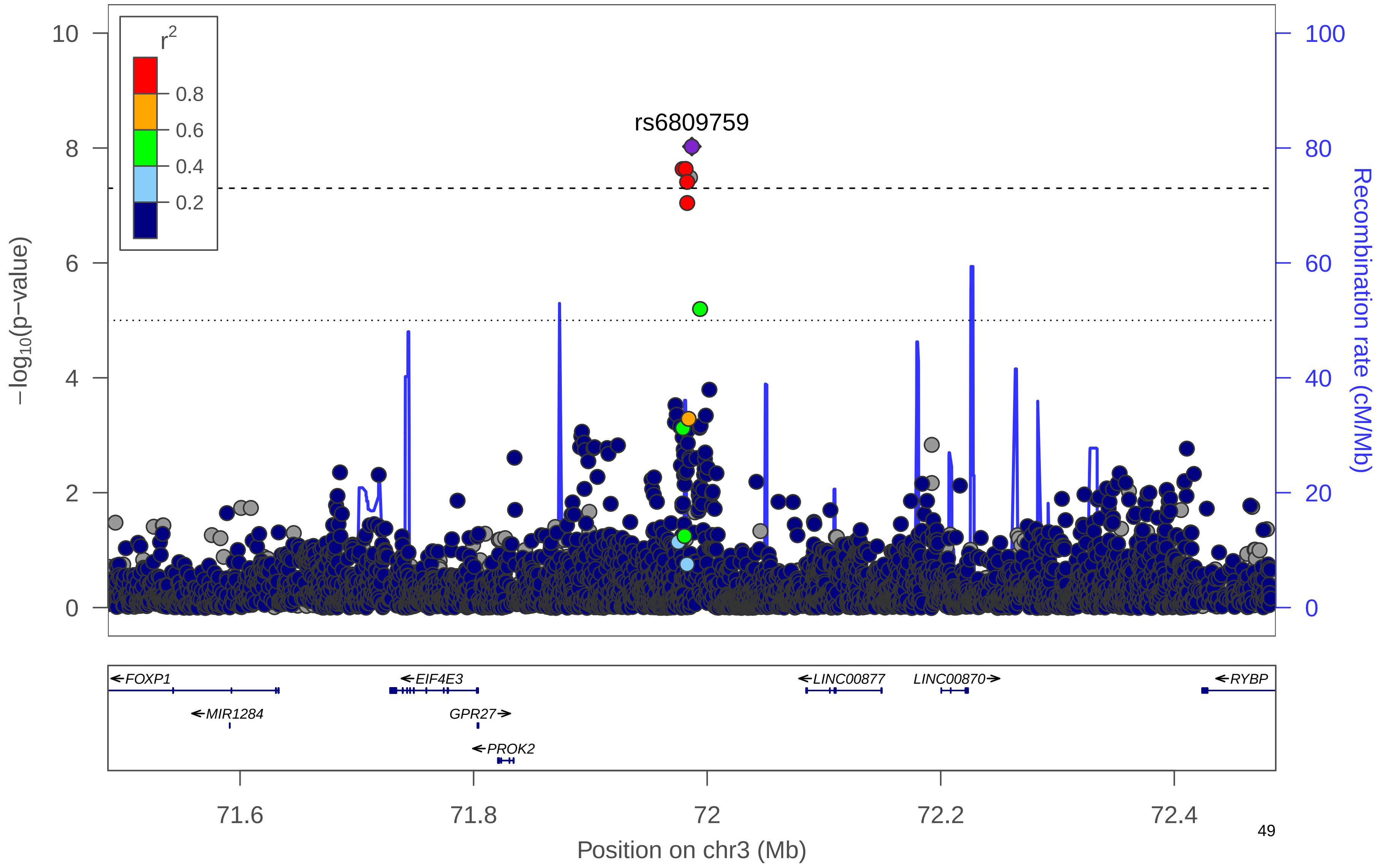

### g) WCadjBMI Men

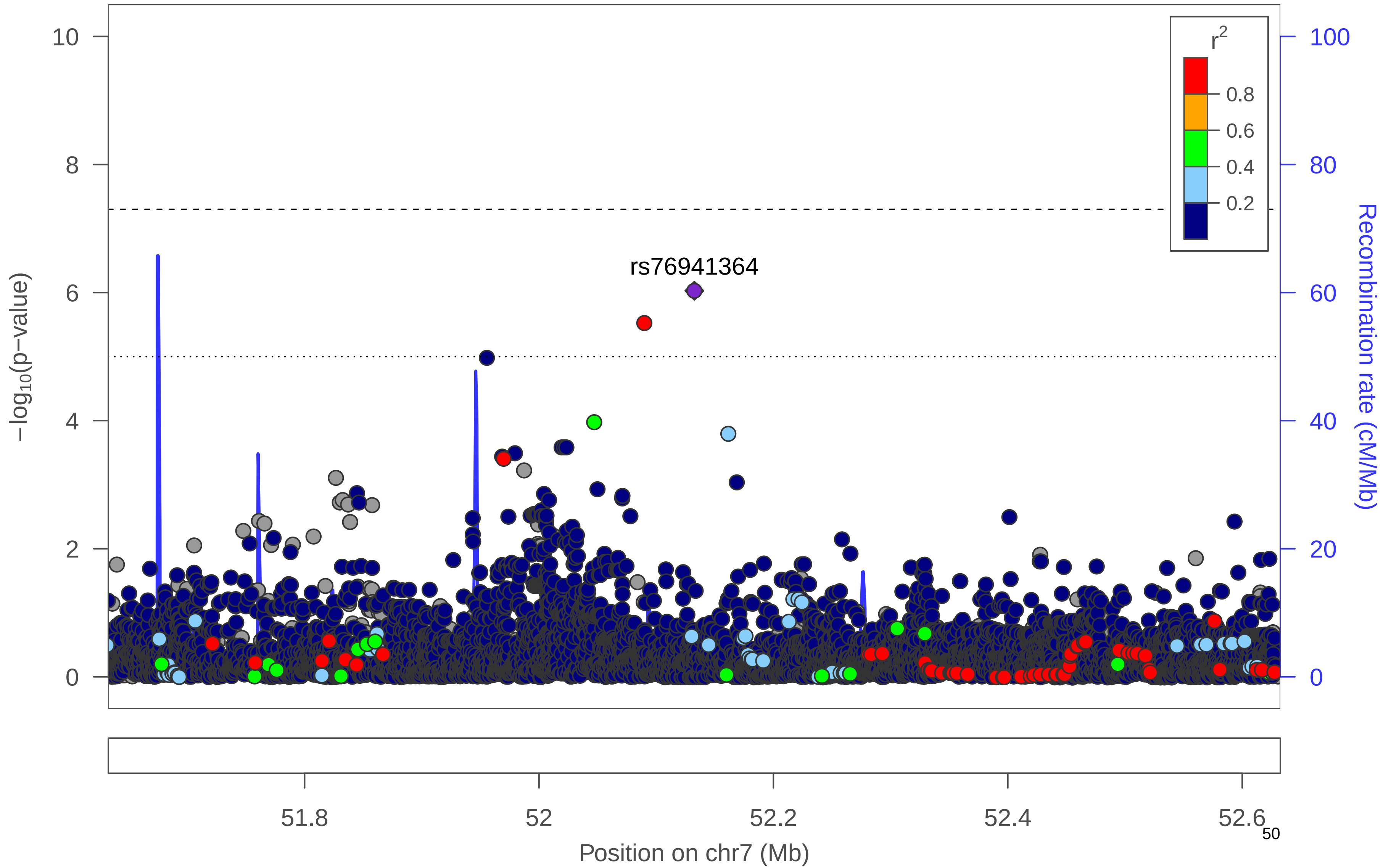

### h) WCadjBMI Men

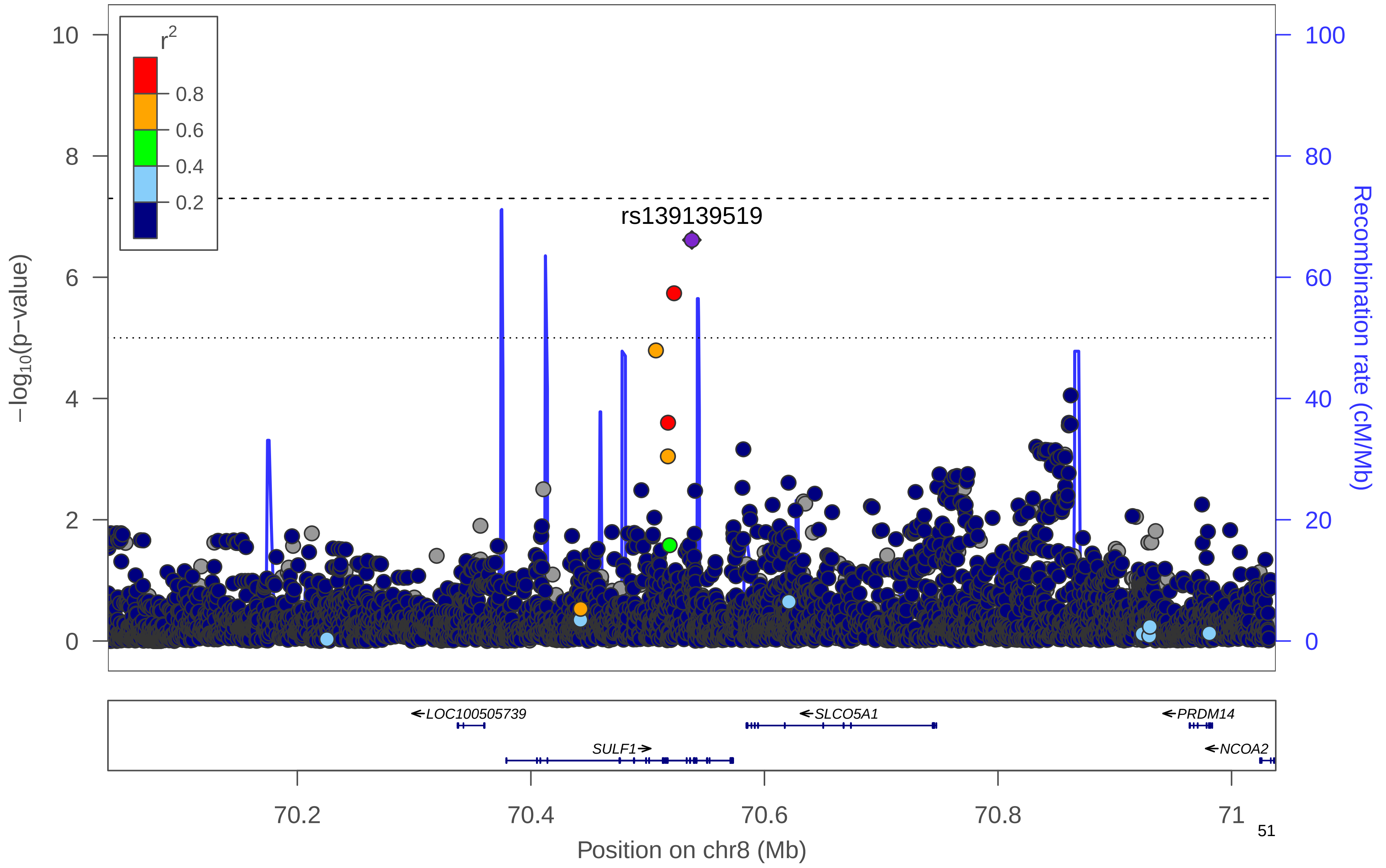

### i) WCadjBMI Men

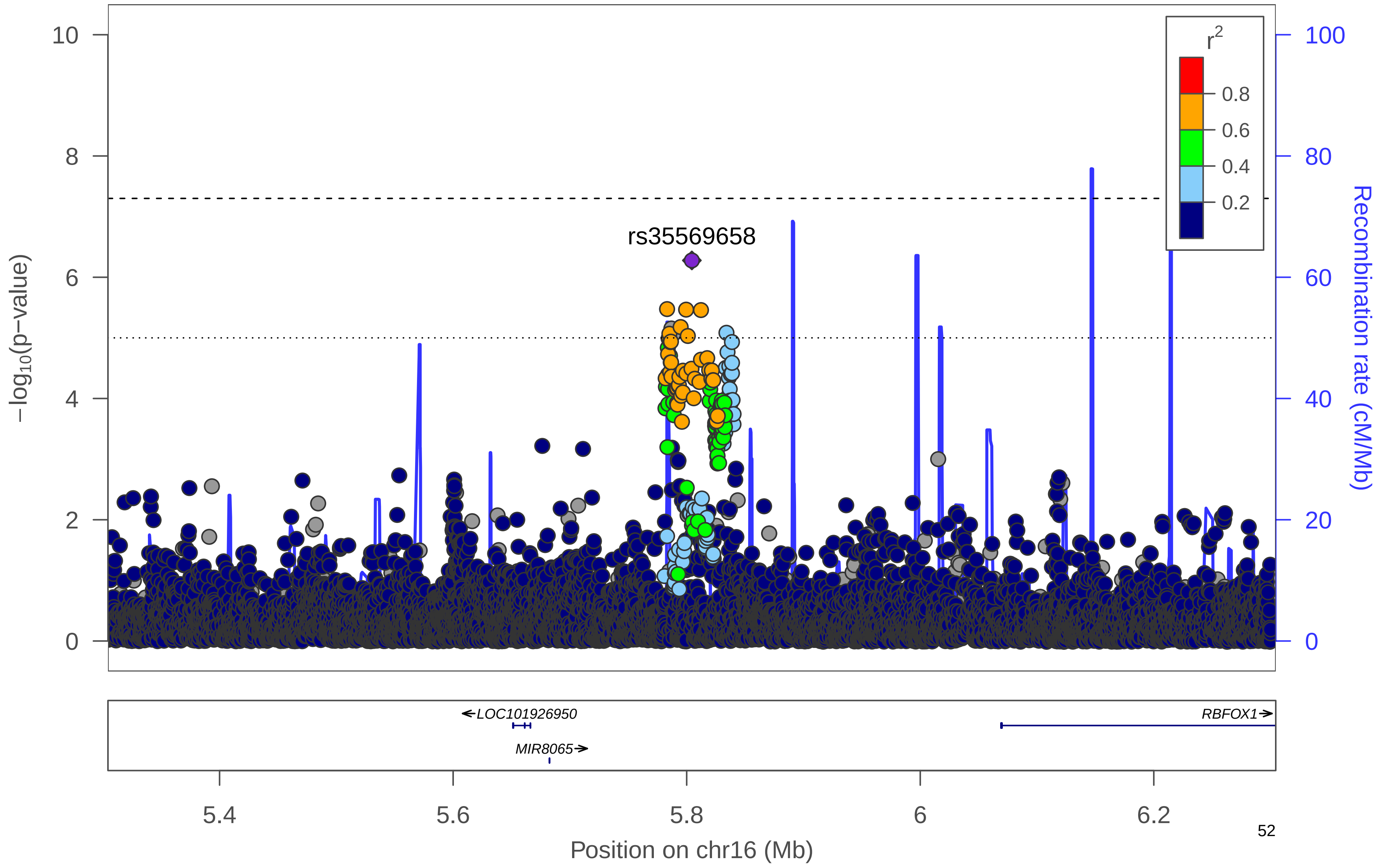

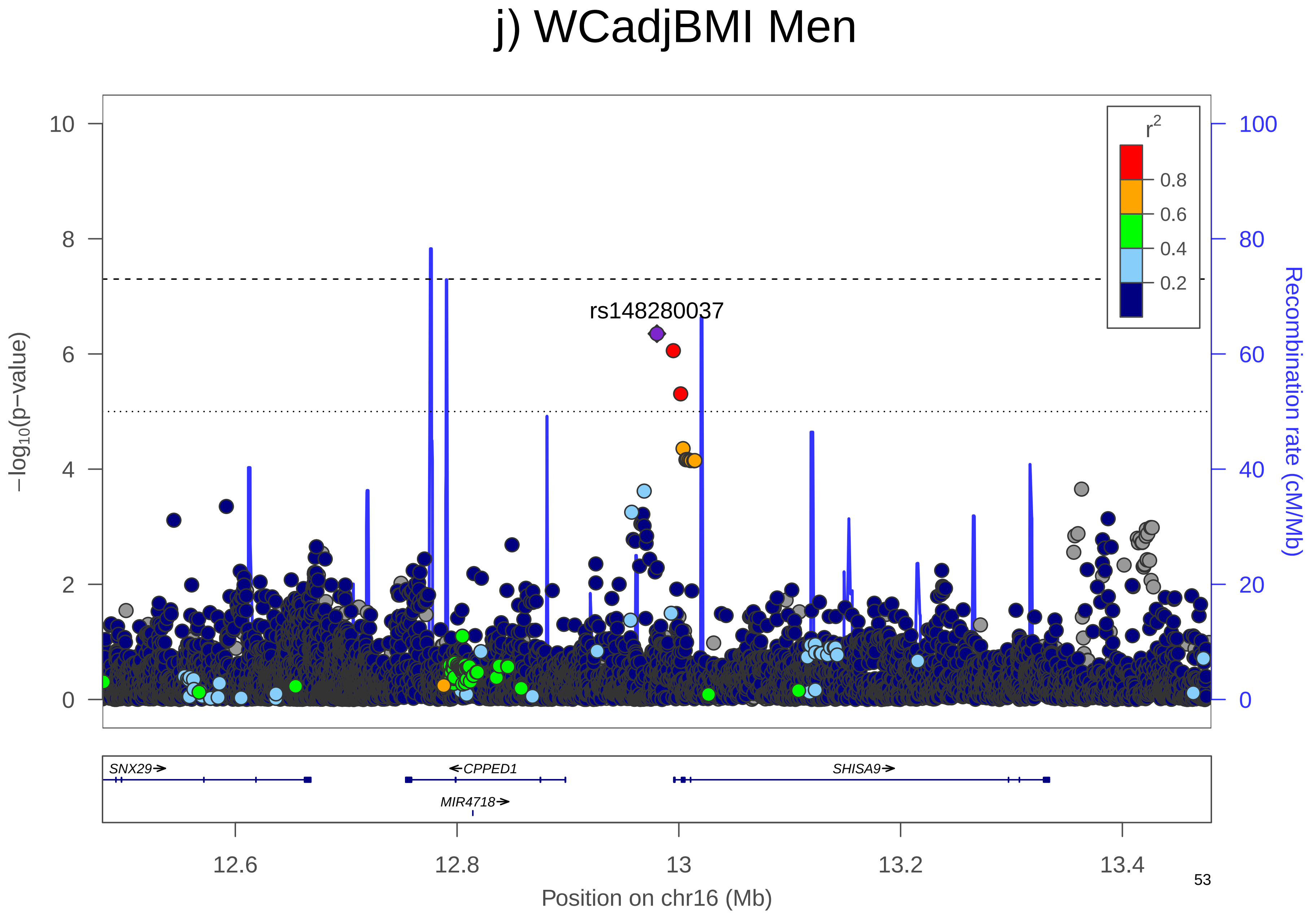

j) WCadjBMI Men
rs148280037
r2
0.8
0.6
0.4
0.2
10
8
6
4
2
0
100
80
60
40
20
0
−log10(p−value)
Recombination rate (cM/Mb)
SNX29
CPPED1
SHISA9
MIR4718
12.6
12.8
13
13.2
13.4
Position on chr16 (Mb)

### k) WCadjBMI Men

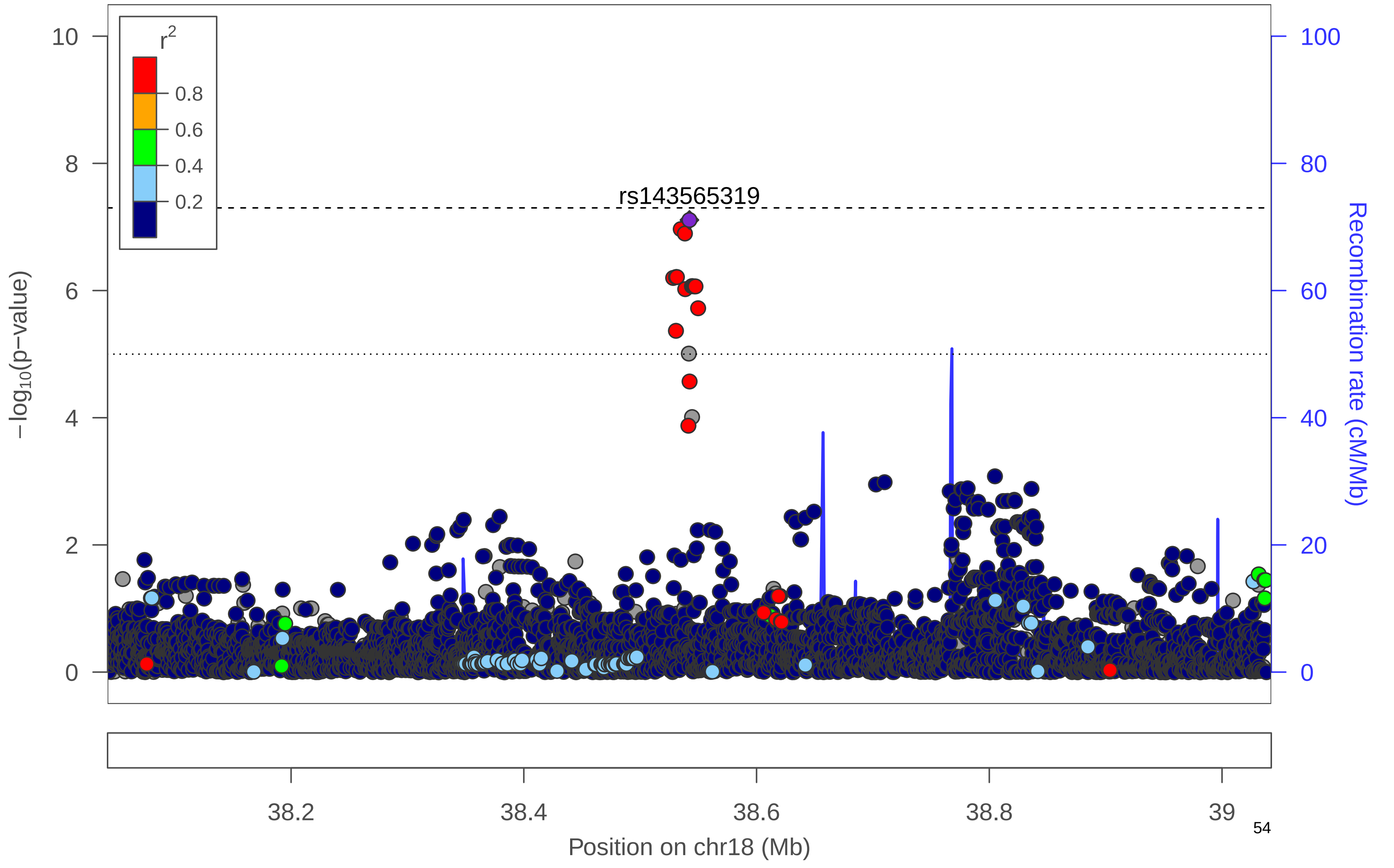

**Supplementary Figure 14. QQ Plots**. QQ Plots for WCadjBMI, including sexes-combined (black), women-only (orange), men-only (blue).

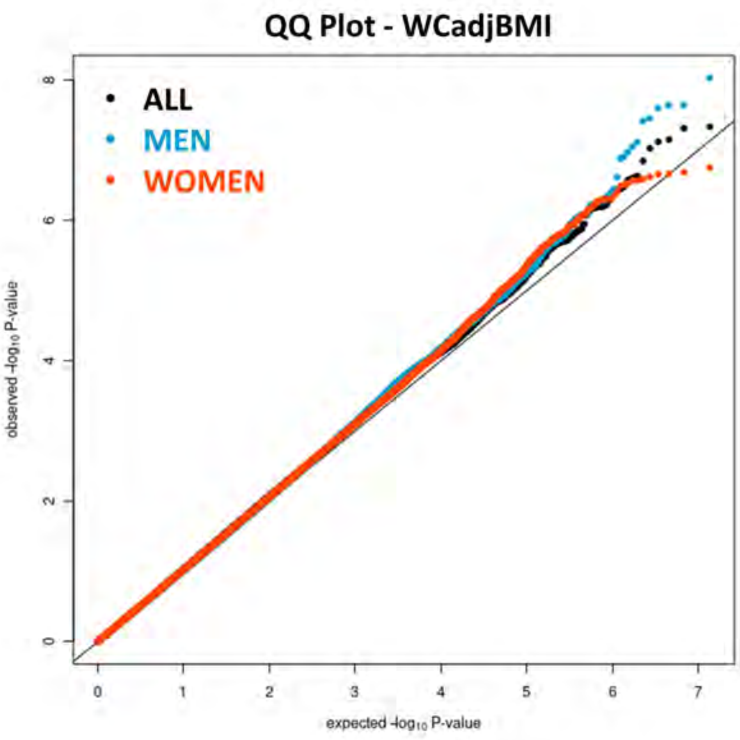

**Supplementary Figure 15. Manhattan plot.** Manhattan plot of the sexes-combined analysis for HIPadjBMI. All suggestively significant (*P*<1x10$^{-6}$) variants are highlighted in orange if they are >500 Kb from any previously-reported HIPadjBMI associated variants. Previously reported loci (+/- 500 Kb) are highlighted in blue if any variant in the locus reached suggestive significance. All suggestively significant loci that meet our criteria for replication are annotated with the closest gene. †Replicated in African American meta-analysis. ‡ Replicated in Hispanic/Latino meta-analysis. ¥ Replicated in European American meta-analysis.

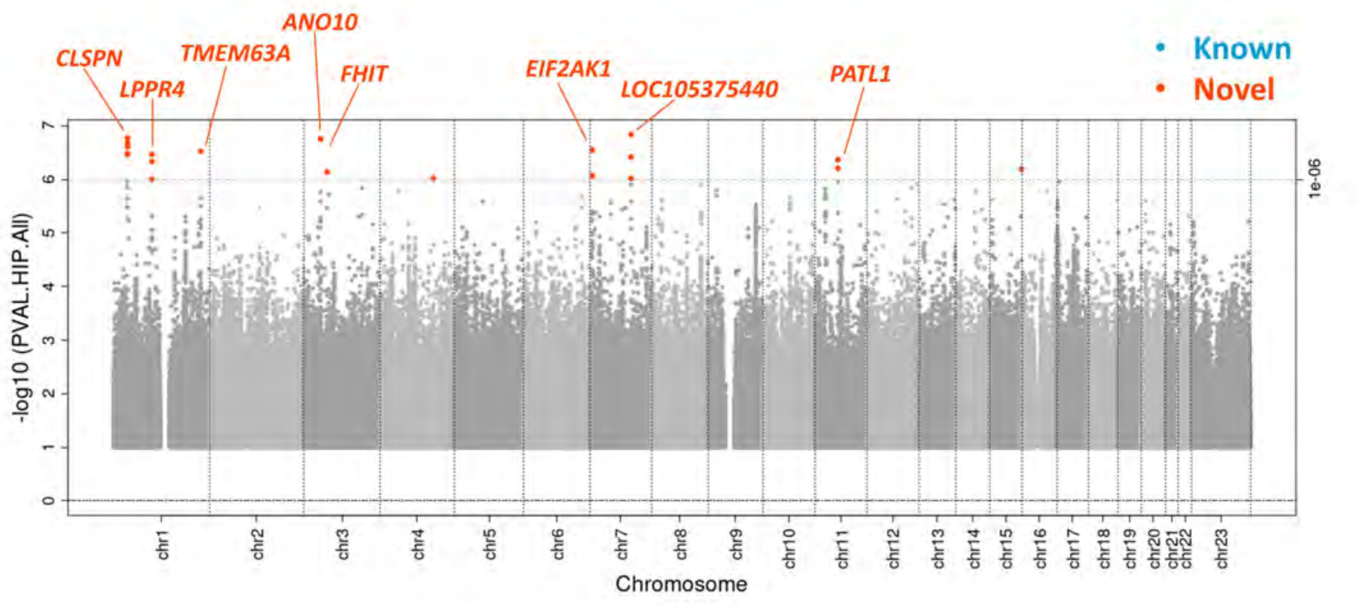

**Supplementary Figure 16. Locus Zoom Plots: HIPadjBMI.** Regional association plots for suggestively significant loci in the HCHS/SOL HIPadjBMI sexes-combined analysis. The plots appear in chromosome:position order. Dot color reflects $R^2$ calculated from the 1000 Genomes AMR reference dataset. Point symbols represent variant functional classifications: a) rs144655586, *CLSPN*; b) rs712900, *LPPR4*; c) rs115546449, *TMEM63A*; d) rs145815581, *ANO10*; e) rs72886347, *FHIT*; f) rs17136358, *EIF2AK1*; g) rs117683919, *LOC105375440*; h) rs143542634, *PATL1*.

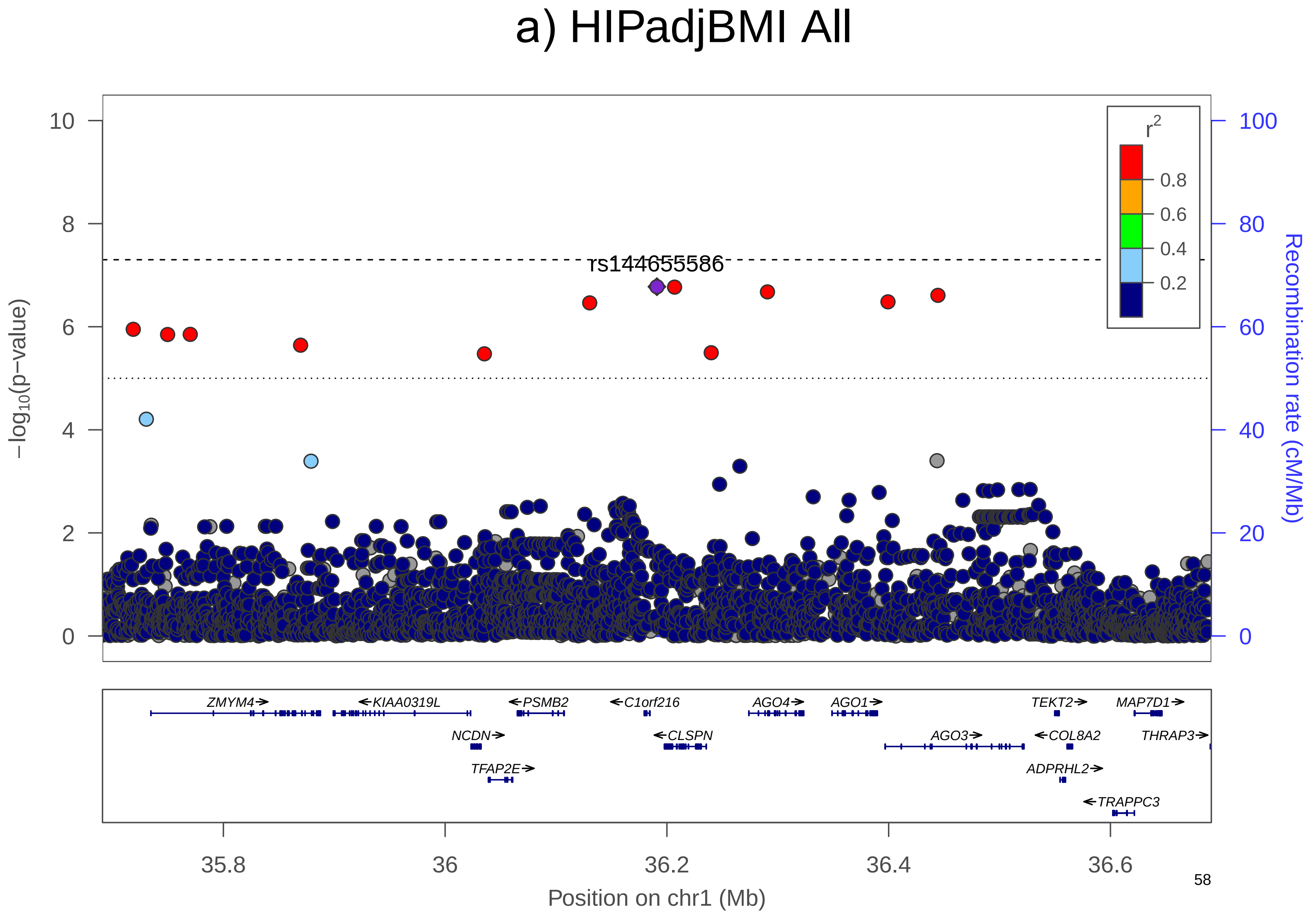
a) HIPadjBMI All
rs144655586
−log10(p−value)
Recombination rate (cM/Mb)
r2
0.8
0.6
0.4
0.2
ZMYM4
KIAA0319L
PSMB2
C1orf216
AGO4
AGO1
TEKT2
MAP7D1
NCDN
CLSPN
AGO3
COL8A2
THRAP3
TFAP2E
ADPRHL2
TRAPPC3
35.8
36
36.2
36.4
36.6
Position on chr1 (Mb)

### b) HIPadjBMI All

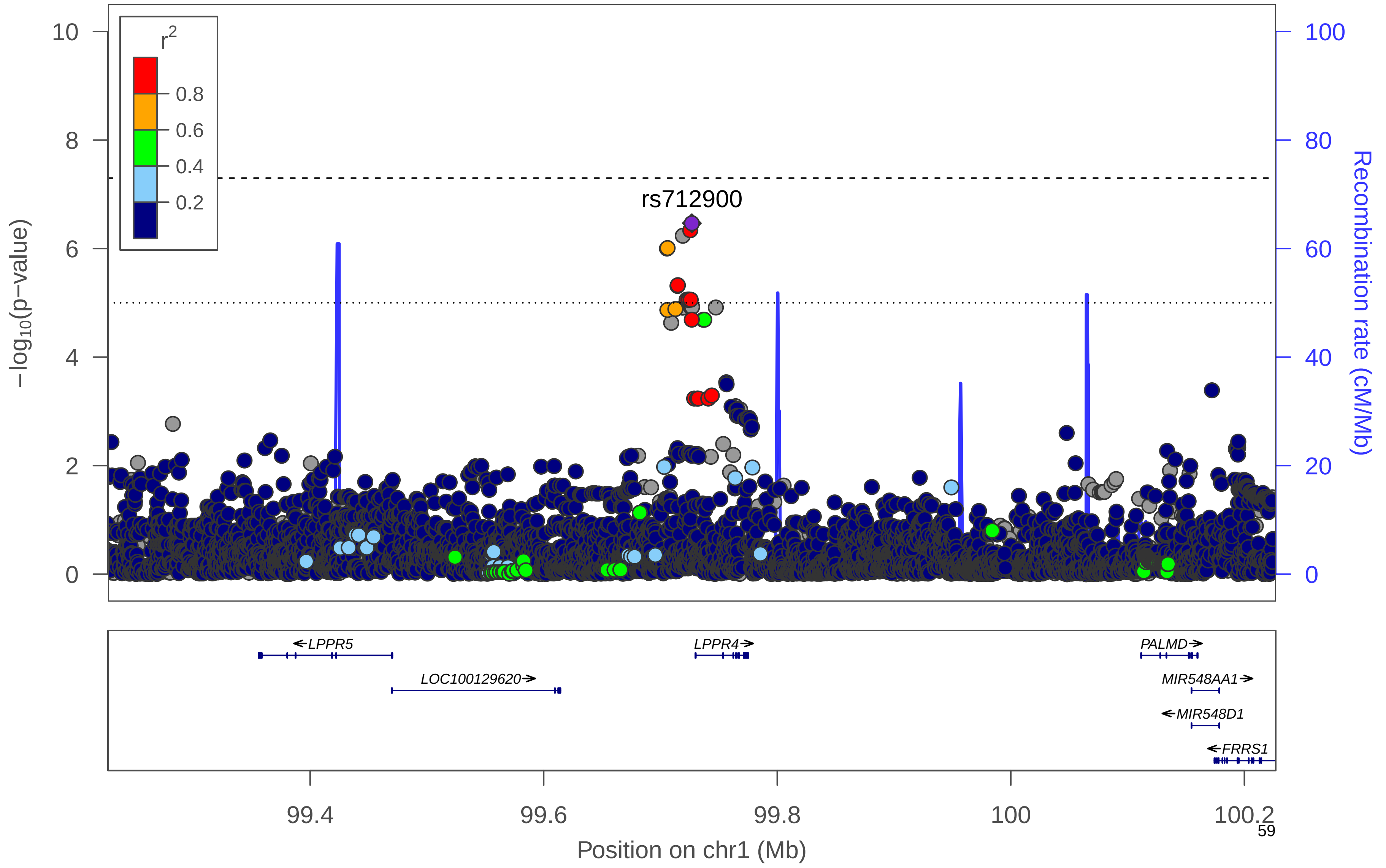

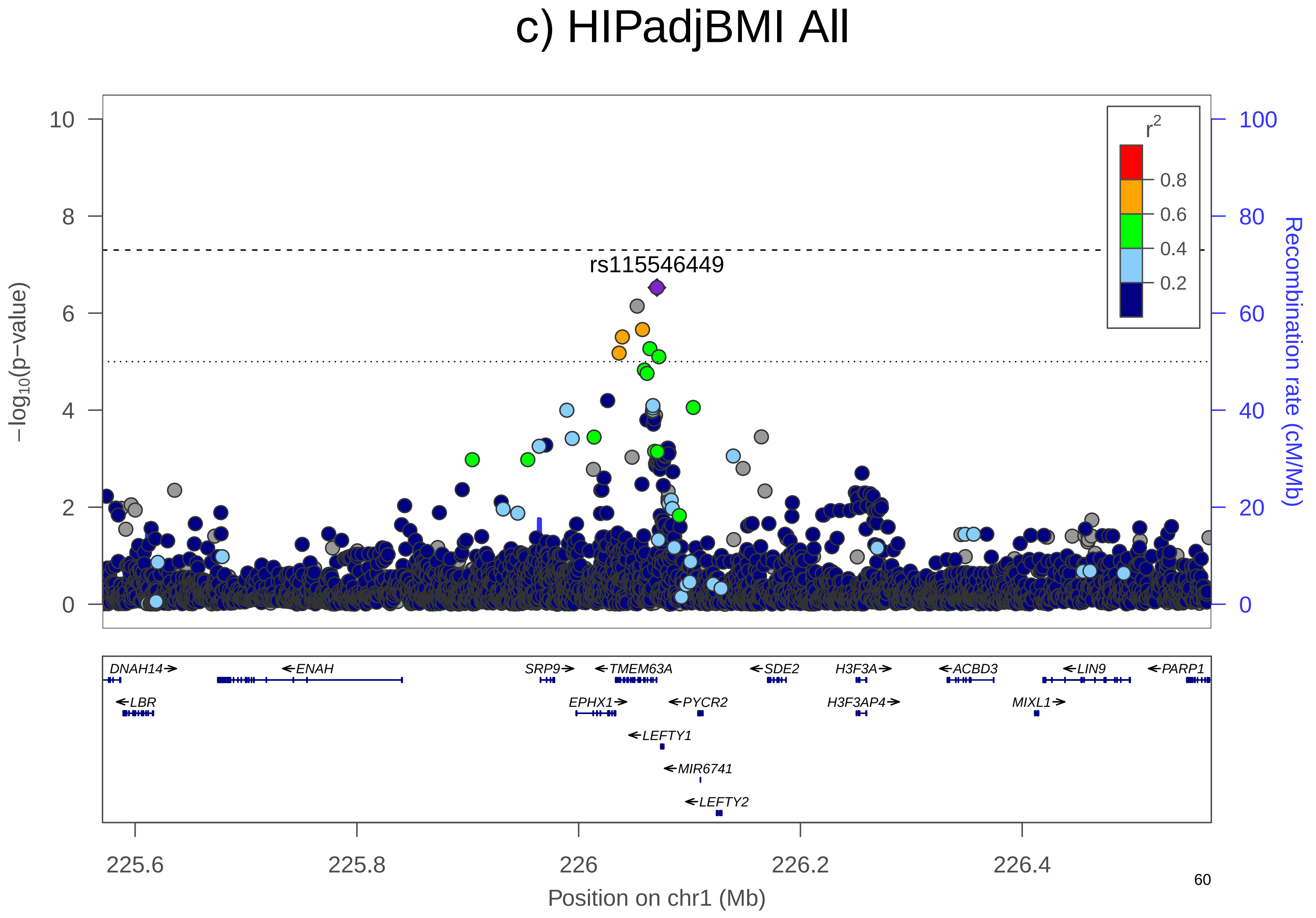
c) HIPadjBMI All
rs115546449
r2
0.8
0.6
0.4
0.2
−log10(p−value)
Recombination rate (cM/Mb)
10
8
6
4
2
0
100
80
60
40
20
0
DNAH14
ENAH
SRP9
TMEM63A
SDE2
H3F3A
ACBD3
LIN9
PARP1
LBR
EPHX1
PYCR2
H3F3AP4
MIXL1
LEFTY1
MIR6741
LEFTY2
225.6
225.8
226
226.2
226.4
Position on chr1 (Mb)

### d) HIPadjBMI All

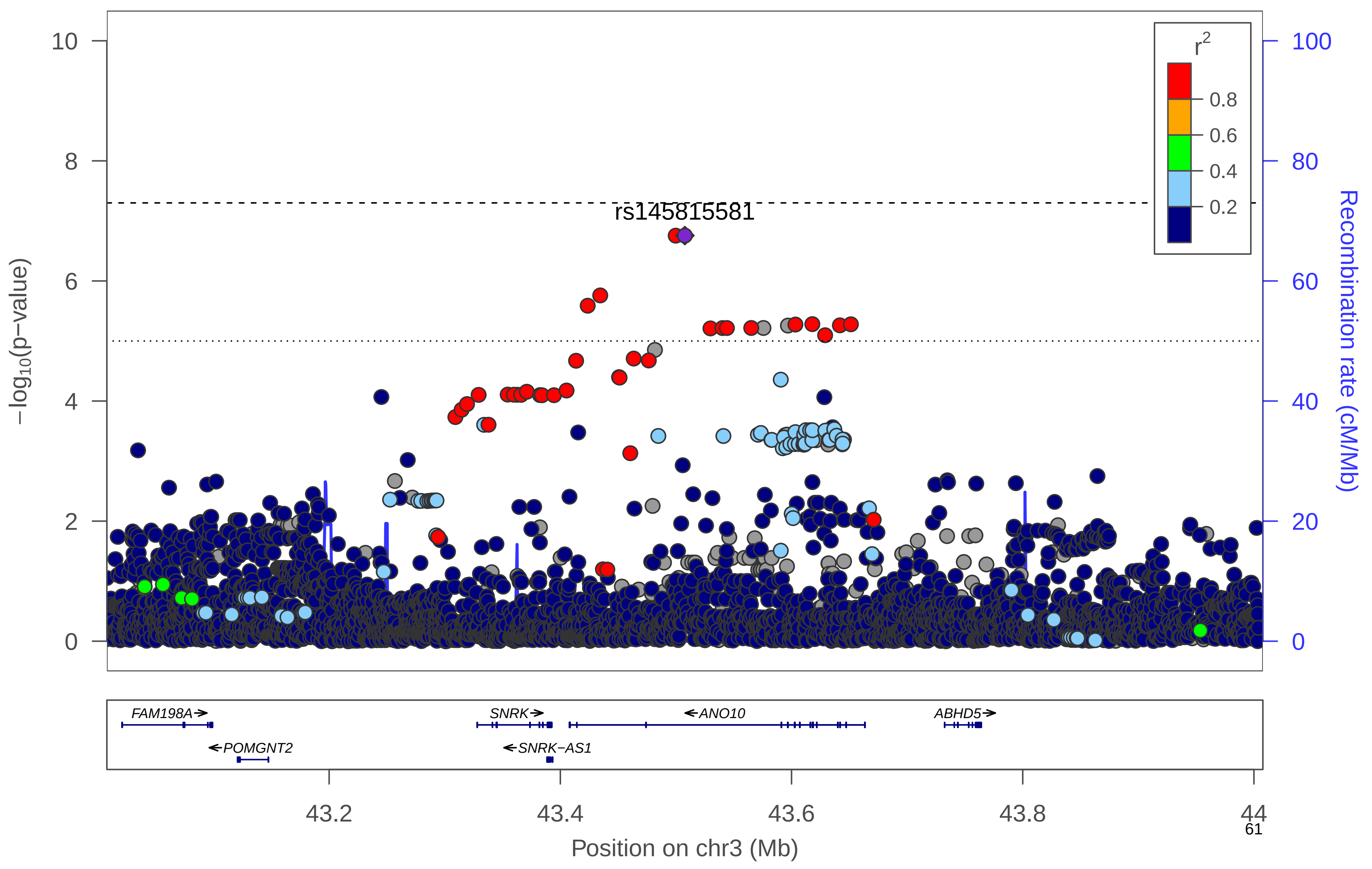

### e) HIPadjBMI All

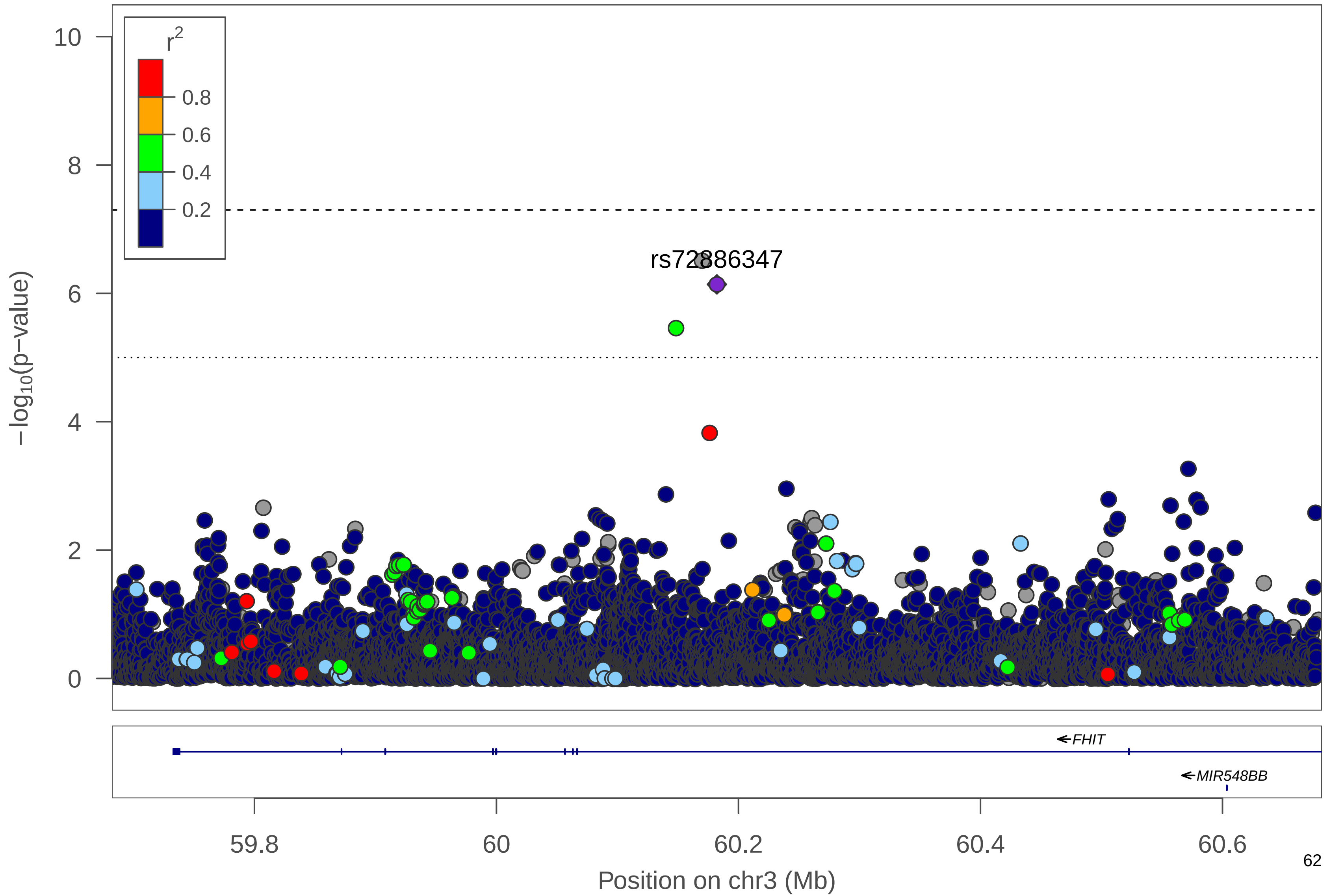

### f) HIPadjBMI All

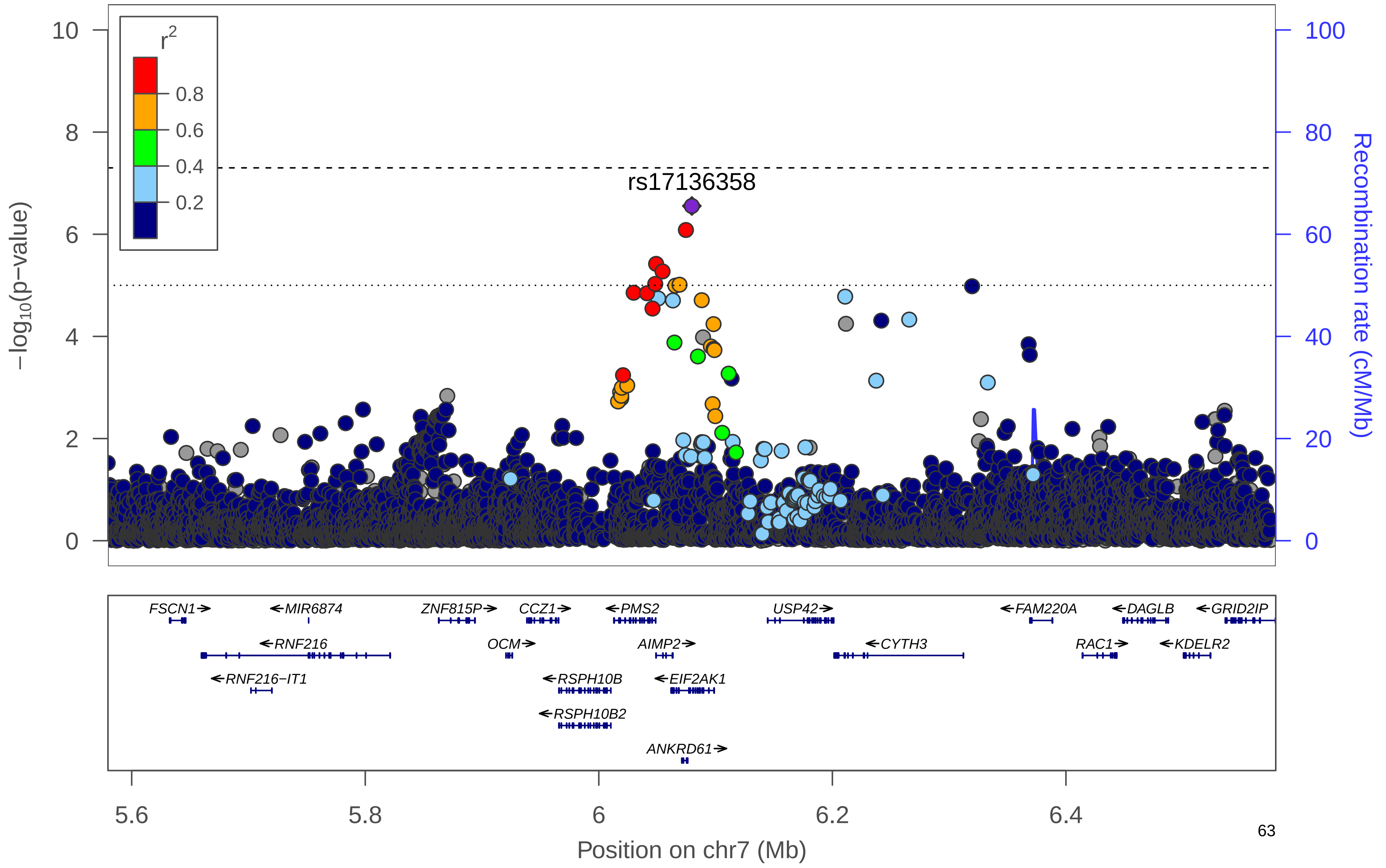

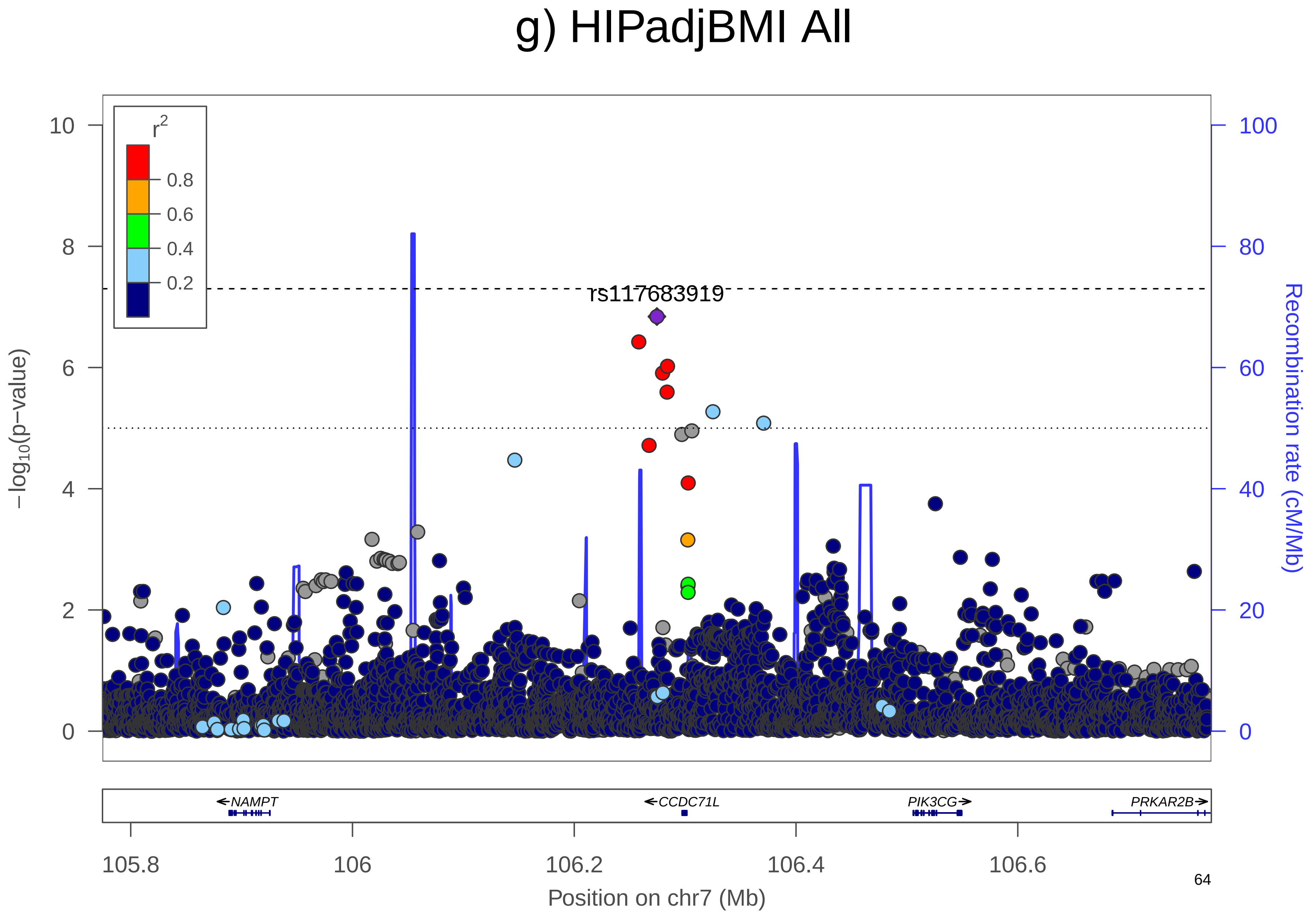

g) HIPadjBMI All
r2
0.8
0.6
0.4
0.2
rs117683919
10
8
6
4
2
0
100
80
60
40
20
0
−log10(p−value)
Recombination rate (cM/Mb)
NAMPT
CCDC71L
PIK3CG
PRKAR2B
105.8
106
106.2
106.4
106.6
Position on chr7 (Mb)

### h) HIPadjBMI All

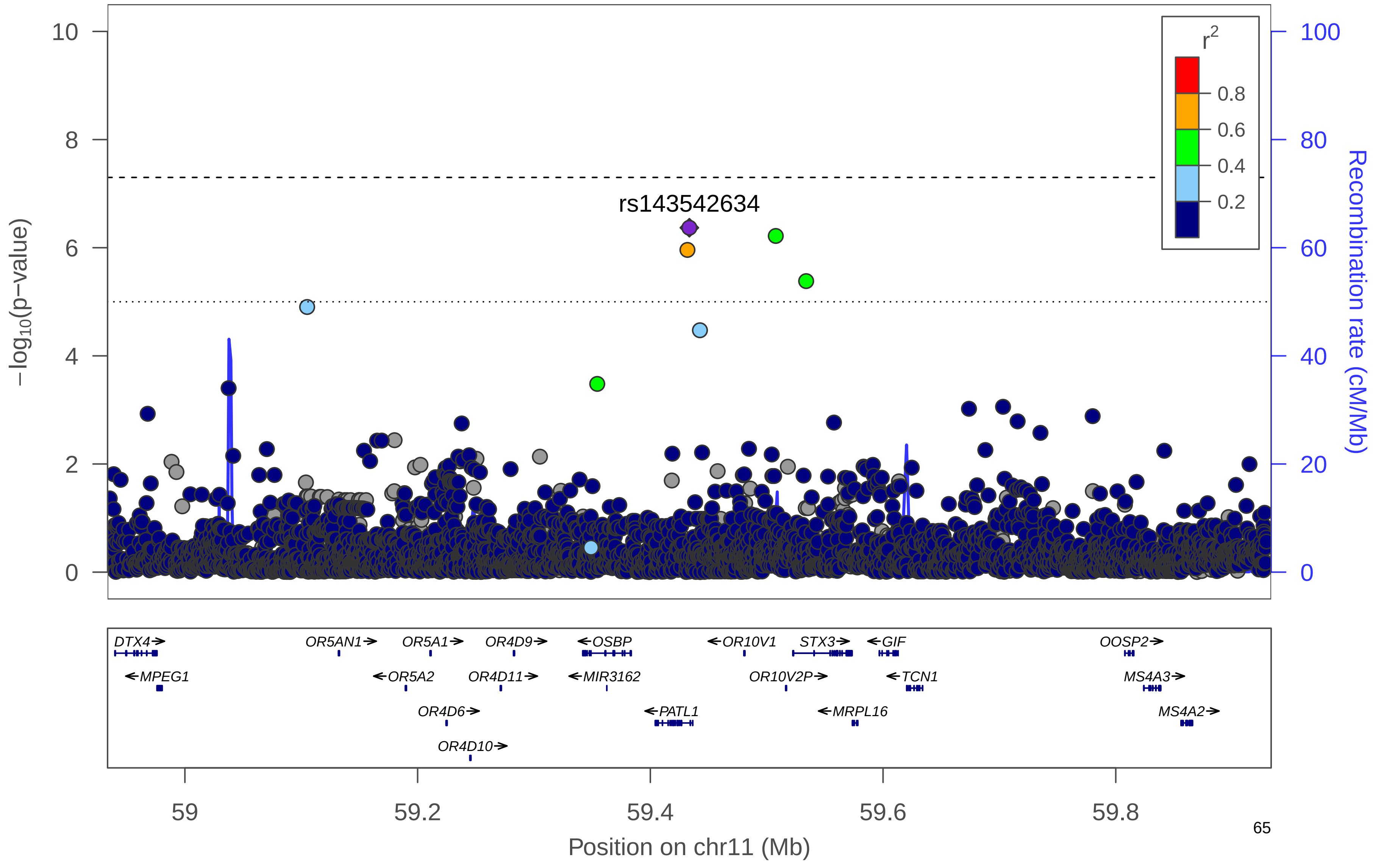

**Supplementary Figure 17. Manhattan plot.** Manhattan plot of the women-only analysis for HIPadjBMI. All suggestively significant ($P$<1x10$^{-6}$) variants are highlighted in orange if they are >500 Kb from any previously-reported HIPadjBMI associated variants. Previously reported loci (+/- 500 Kb) are highlighted in blue if any variant in the locus reached suggestive significance. All suggestively significant loci that meet our criteria for replication are annotated with the closest gene. †Replicated in African American meta-analysis. ‡ Replicated in Hispanic/Latino meta-analysis. ¥ Replicated in European American meta-analysis.

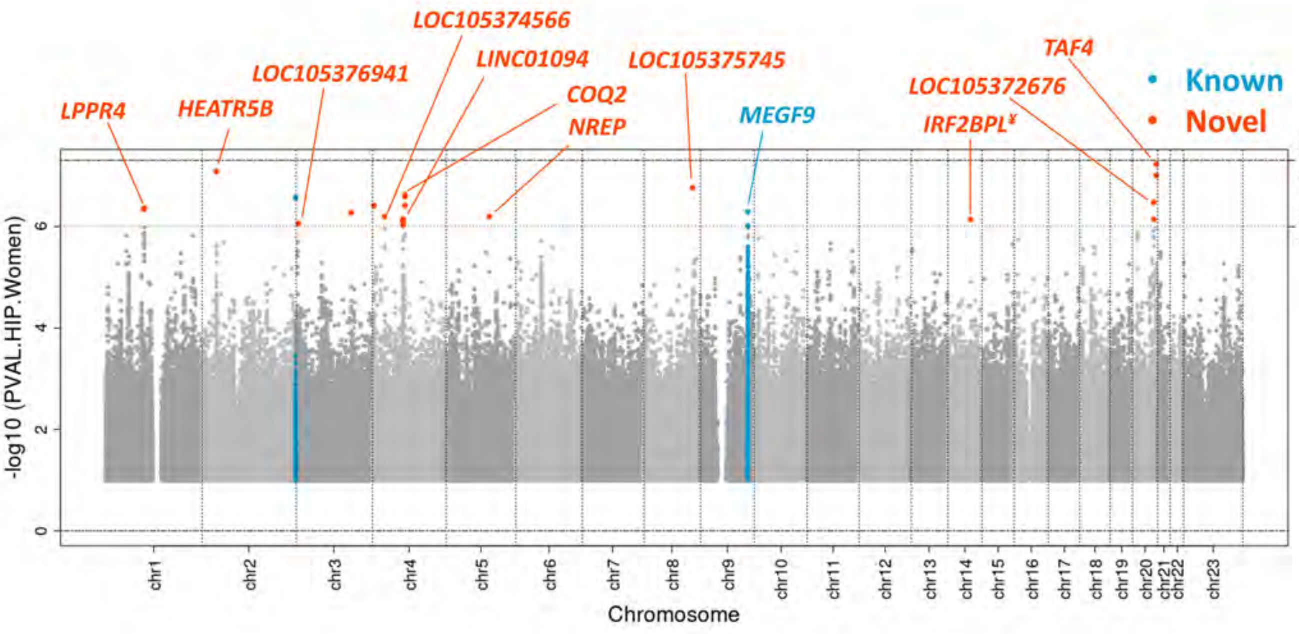

**Supplementary Figure 18. Locus Zoom Plots.** Regional association plots for suggestively significant loci in the HCHS/SOL HIPadjBMI women-only analysis. The plots appear in chromosome:position order. Dot color reflects $R^2$ calculated from the 1000 Genomes AMR reference dataset. Point symbols represent variant functional classifications: a) rs72978809, *LPPR4*; b) rs12478843, *HEATR5B*; c) rs115331260, *LOC105376941*; d) rs7662640, *LOC105374566*; e) rs6814739, *LINC01094*; f) rs11099588, *COQ2*; g) rs6860625, *NREP*; h) rs77186623, *LOC105375745*; i) rs10818474, *MEGF9*; j) rs28692724, *IRF2BPL*; k) rs6092086, *LOC105372676*; l) rs9631175, *TAF4*.

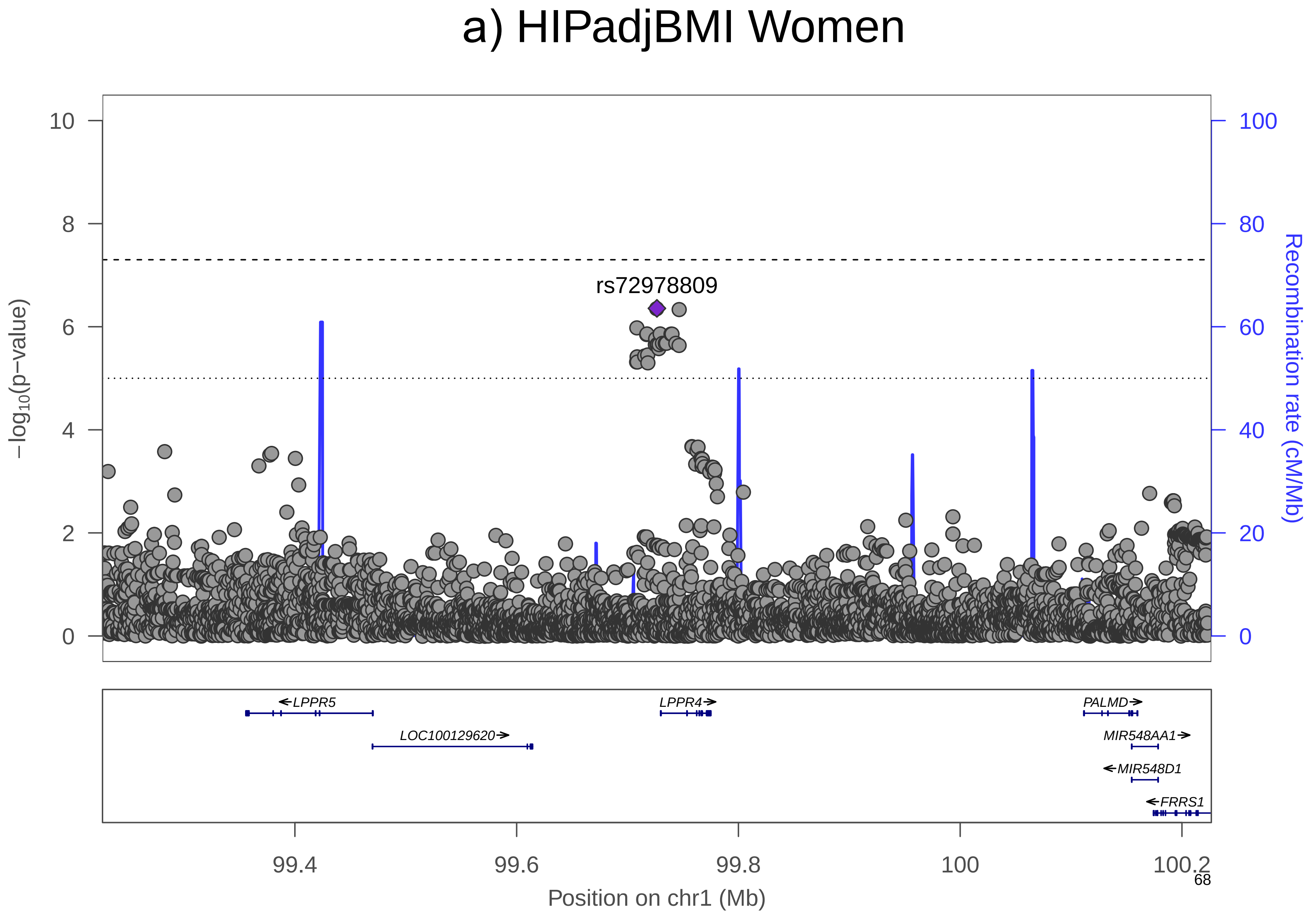
a) HIPadjBMI Women
rs72978809
−log10(p−value)
Recombination rate (cM/Mb)
LPPR5
LOC100129620
LPPR4
PALMD
MIR548AA1
MIR548D1
FRRS1
99.4
99.6
99.8
100
100.2
Position on chr1 (Mb)

### b) HIPadjBMI Women

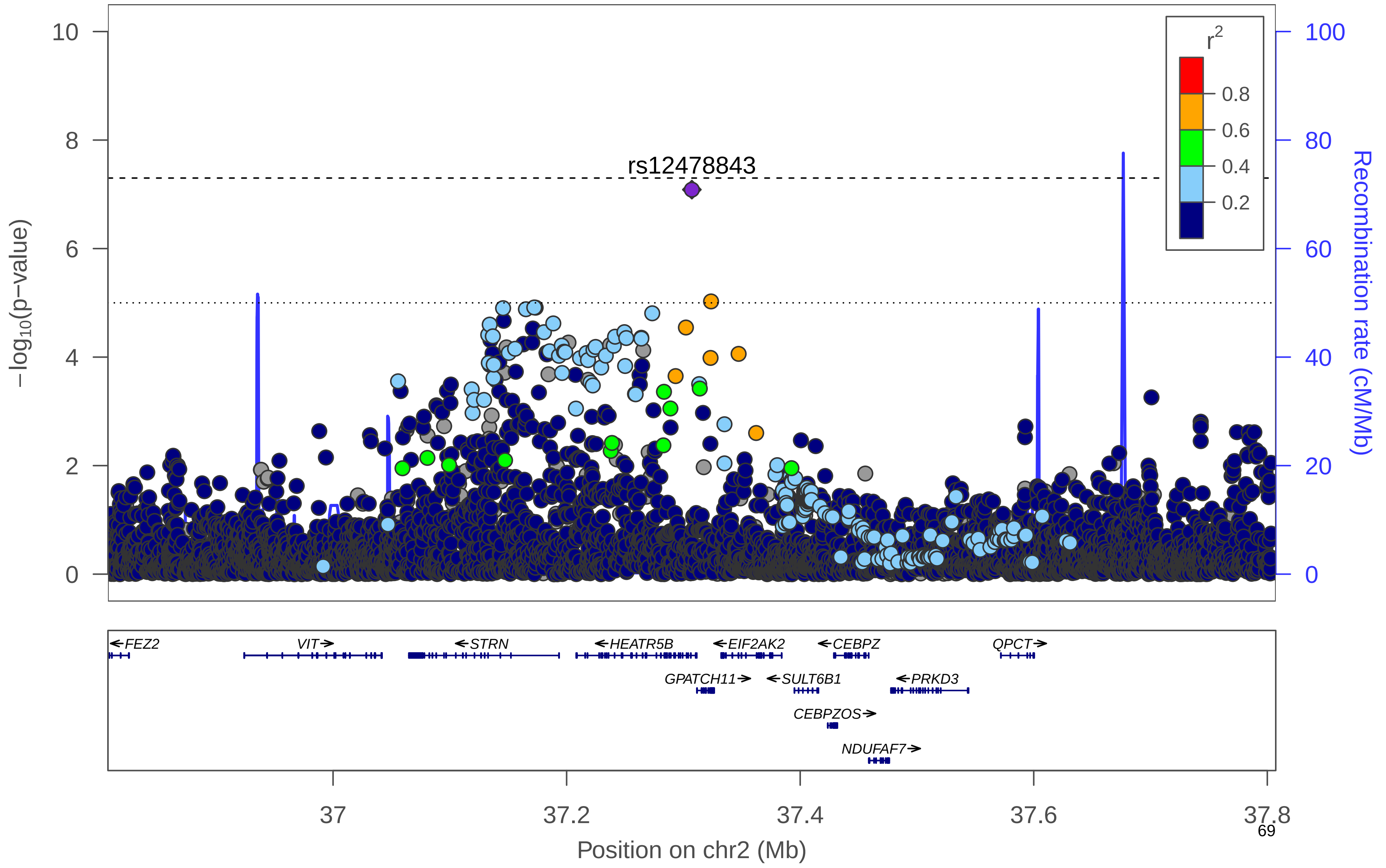

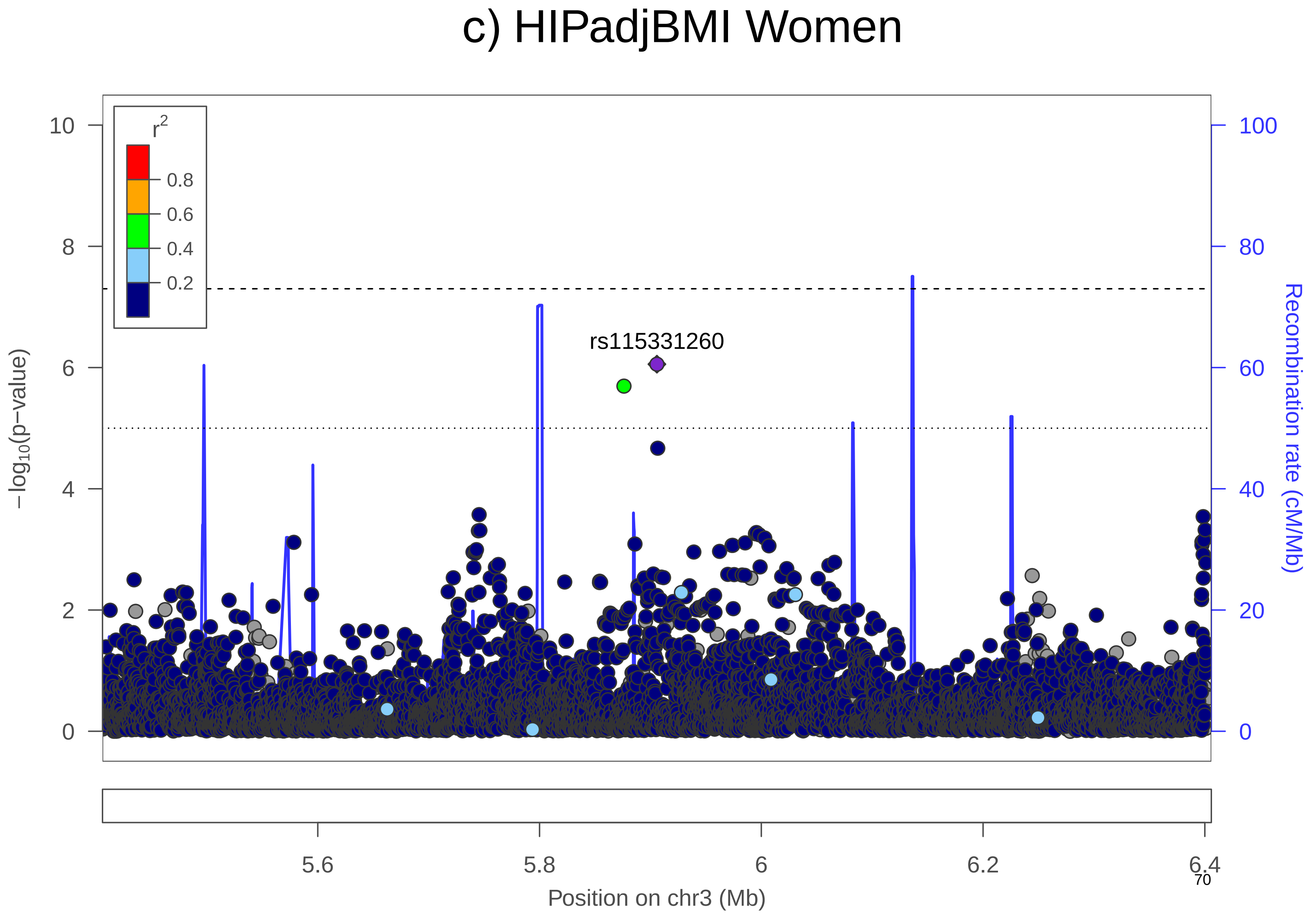
c) HIPadjBMI Women
r2
0.8
0.6
0.4
0.2
rs115331260
−log10(p−value)
Recombination rate (cM/Mb)
10
8
6
4
2
0
100
80
60
40
20
0
5.6
5.8
6
6.2
6.4
Position on chr3 (Mb)

### d) HIPadjBMI Women

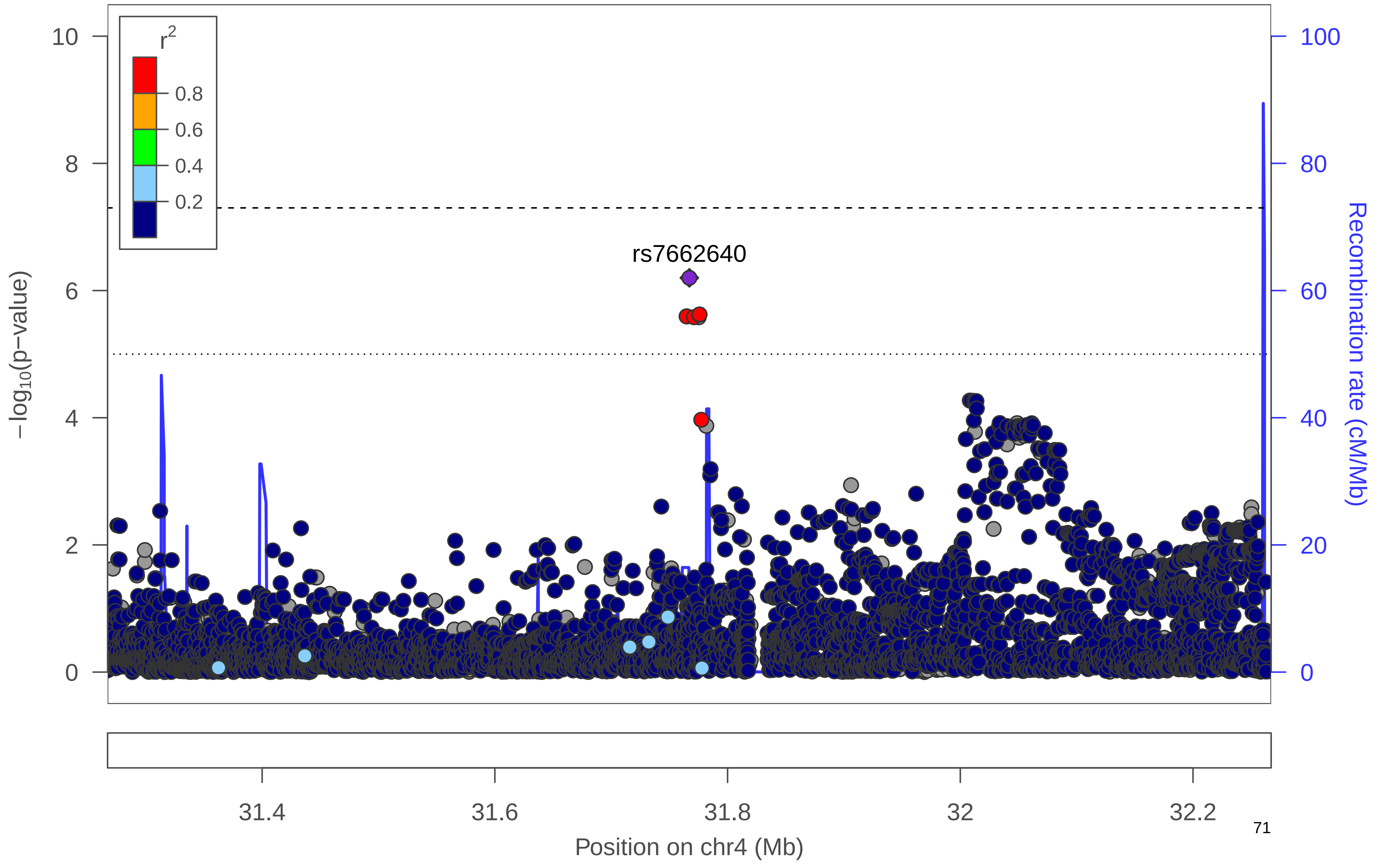

### e) HIPadjBMI Women

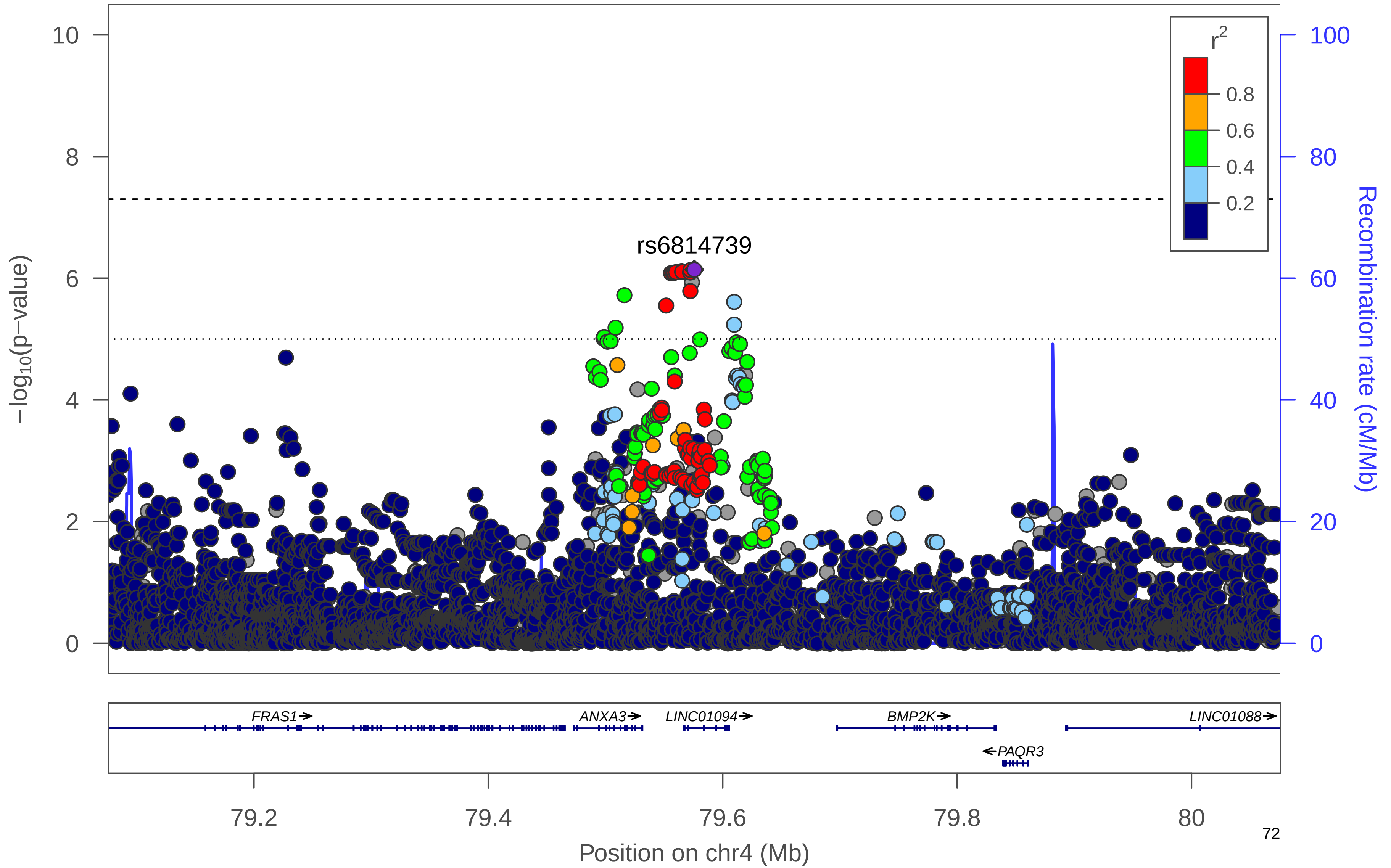

### f) HIPadjBMI Women

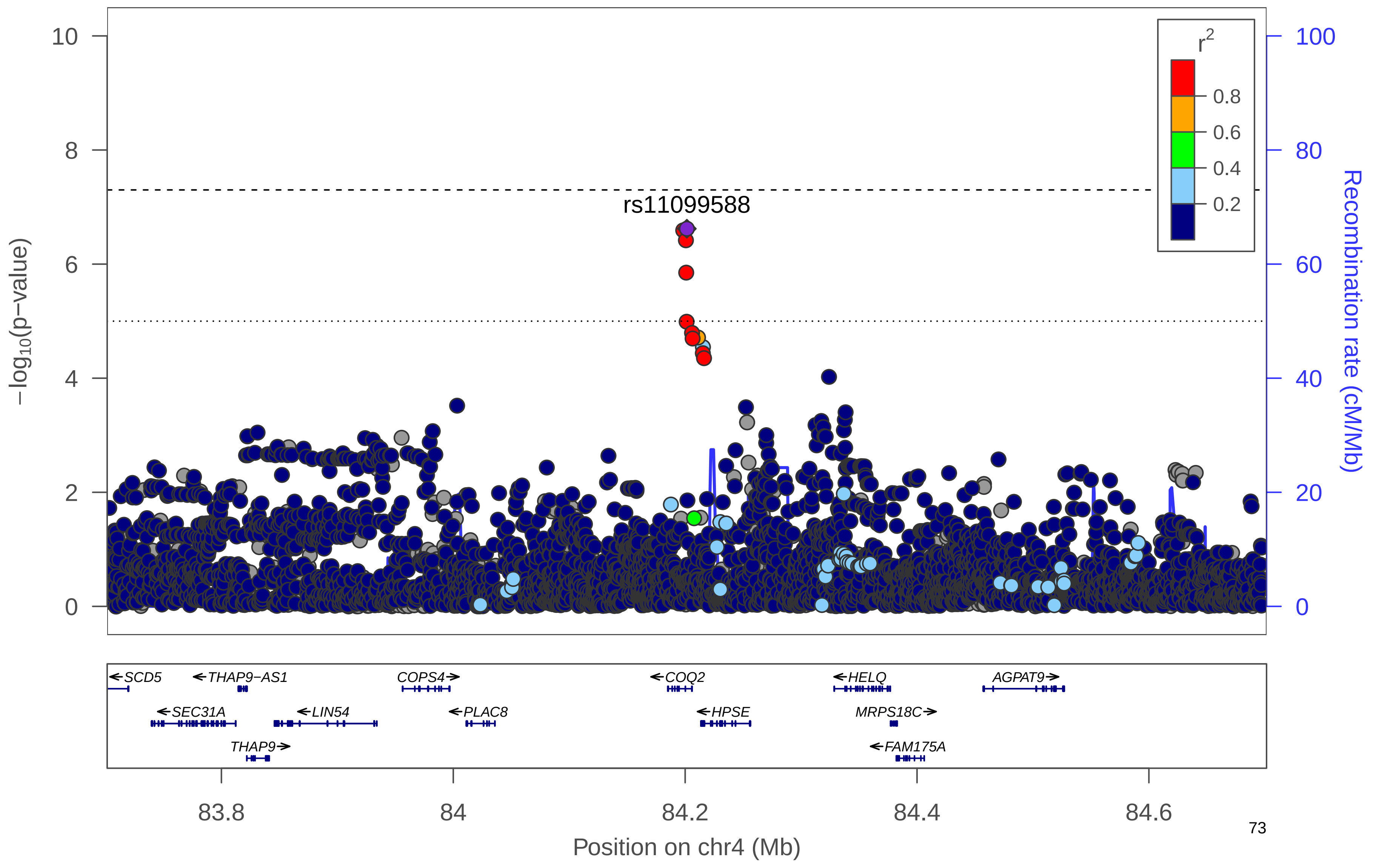

### g) HIPadjBMI Women

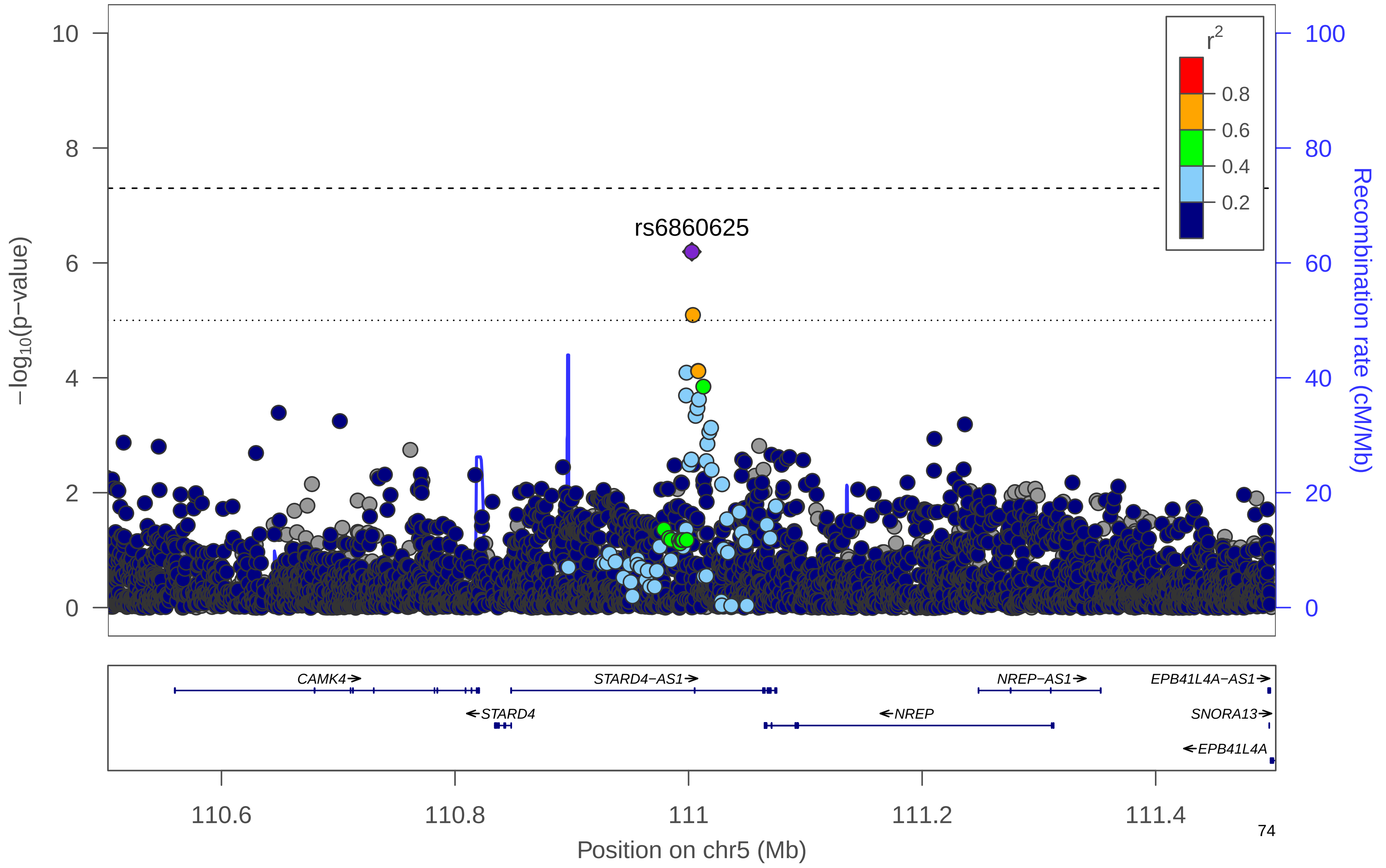

### h) HIPadjBMI Women

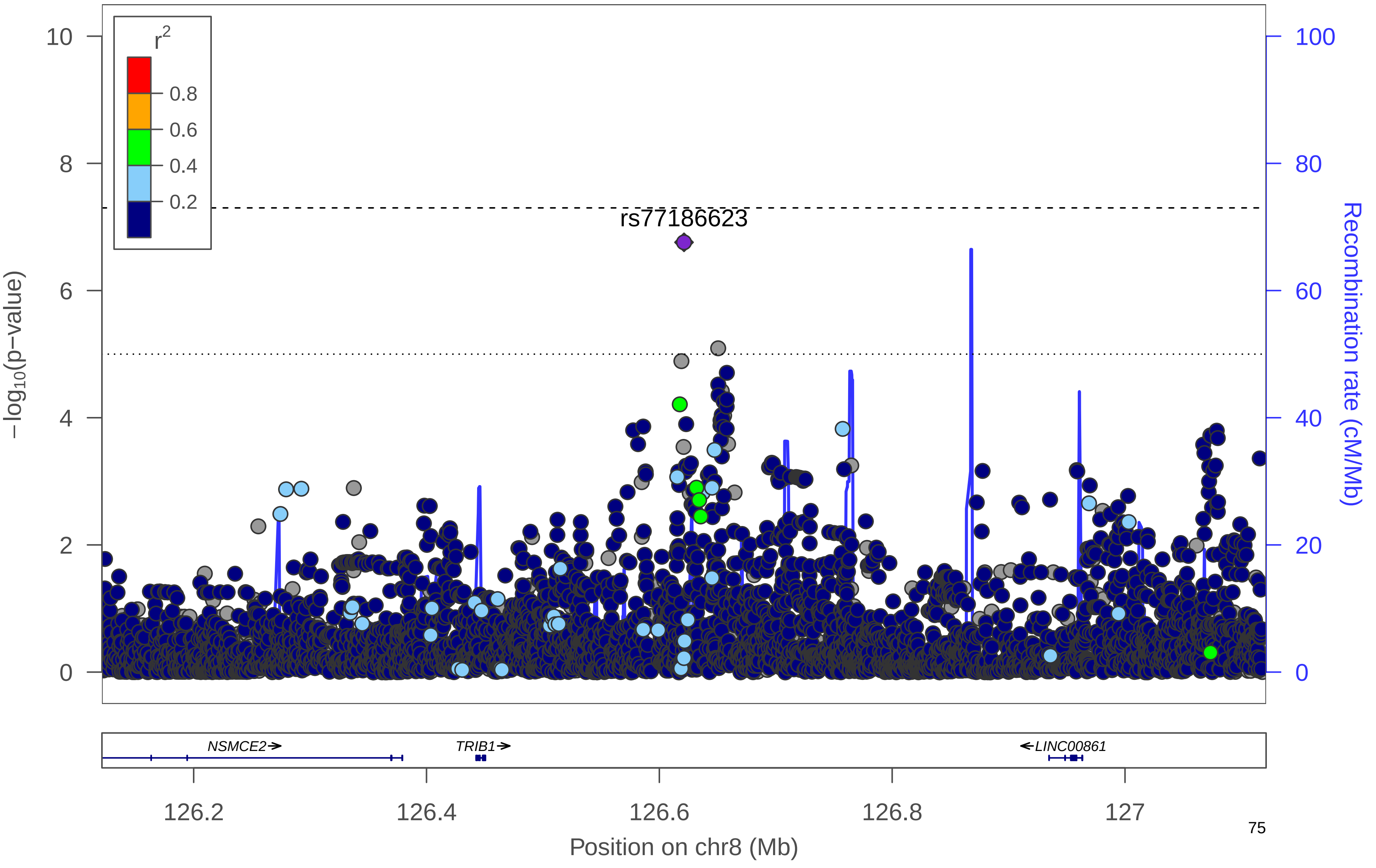

### i) HIPadjBMI Women

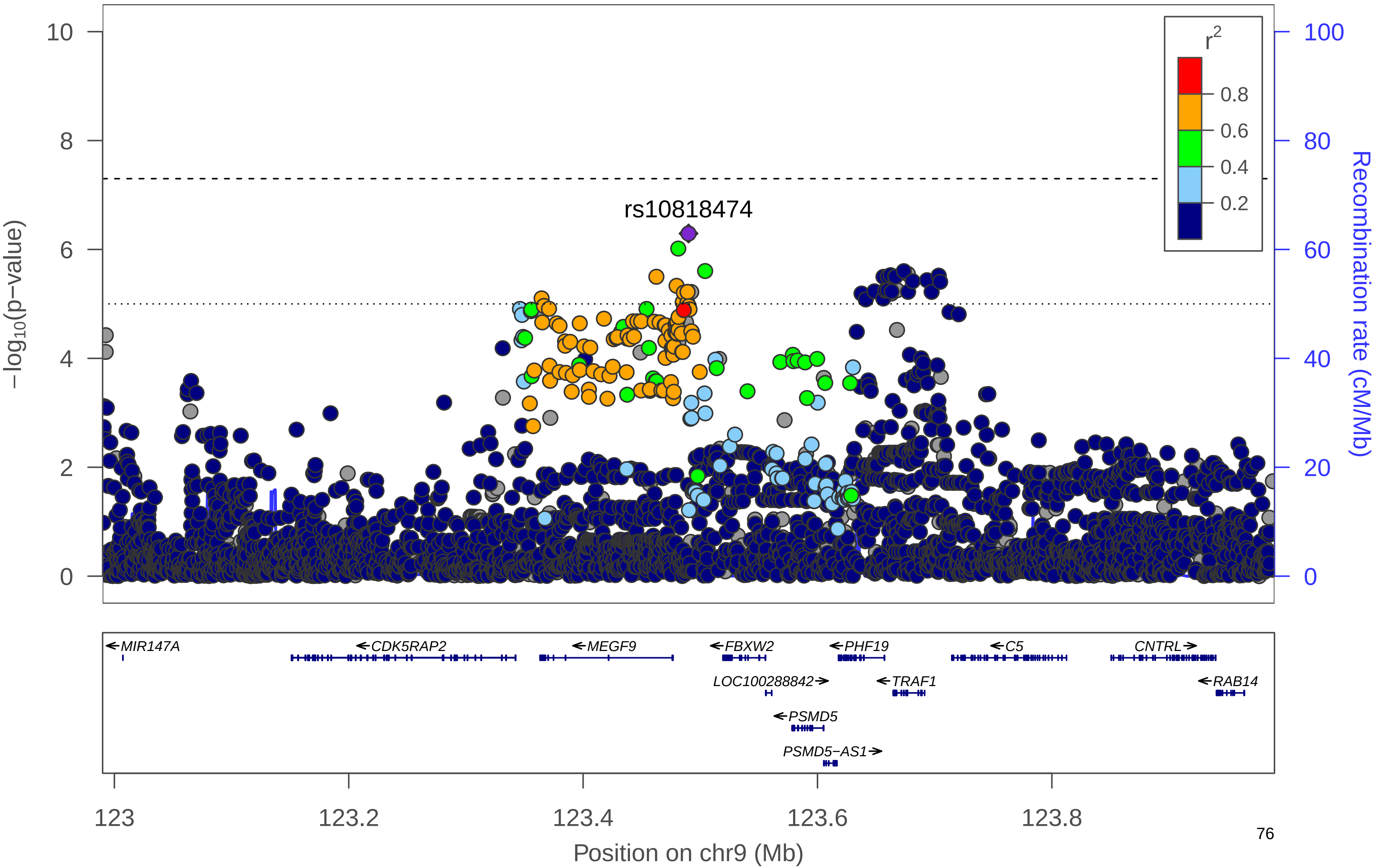

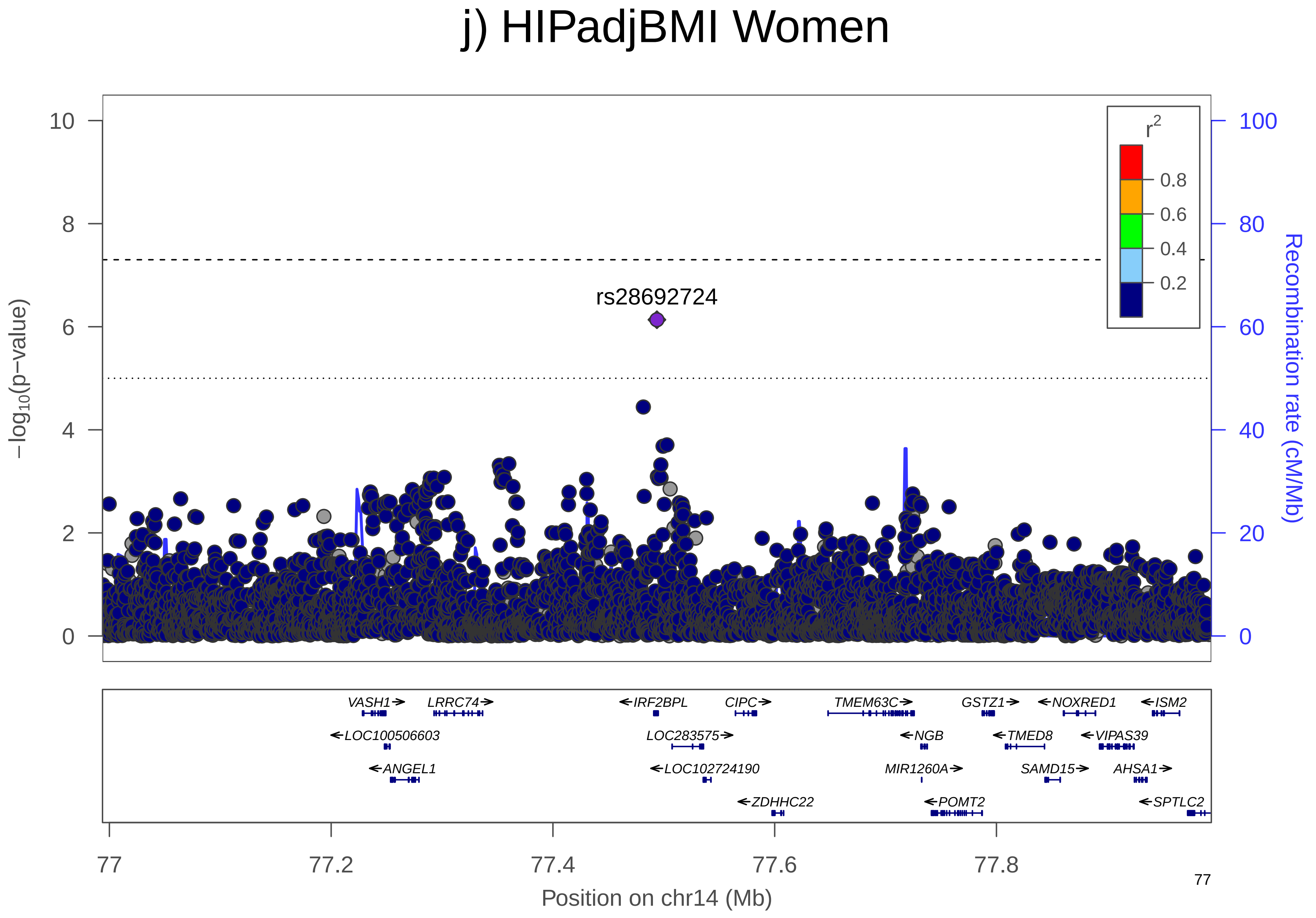

j) HIPadjBMI Women
rs28692724
r2
0.8
0.6
0.4
0.2
−log10(p−value)
Recombination rate (cM/Mb)
0
2
4
6
8
10
0
20
40
60
80
100
VASH1
LRRC74
IRF2BPL
CIPC
TMEM63C
GSTZ1
NOXRED1
ISM2
LOC100506603
LOC283575
NGB
TMED8
VIPAS39
ANGEL1
LOC102724190
MIR1260A
SAMD15
AHSA1
ZDHHC22
POMT2
SPTLC2
77
77.2
77.4
77.6
77.8
Position on chr14 (Mb)

k) HIPadjBMI Women

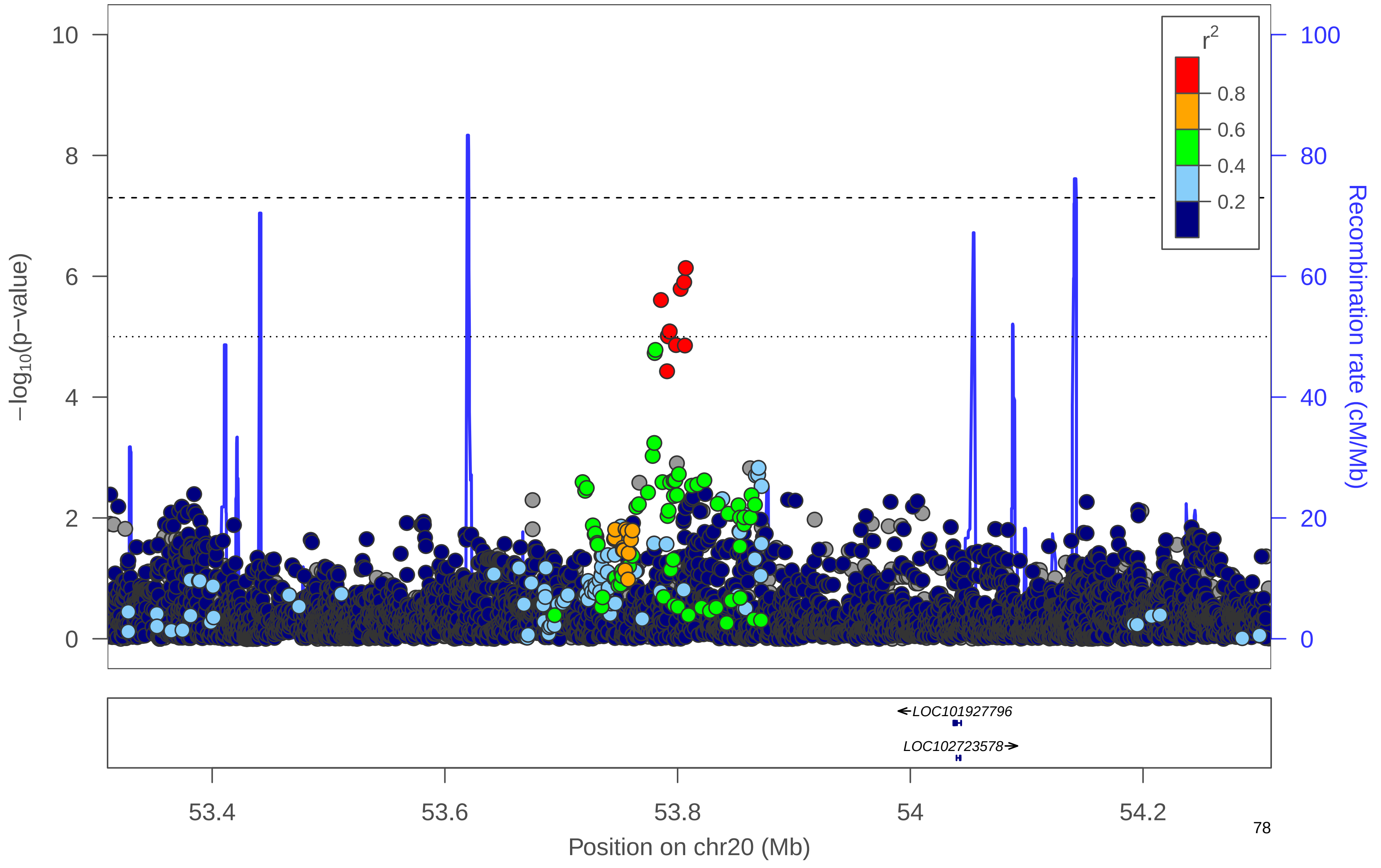

### l) HIPadjBMI Women

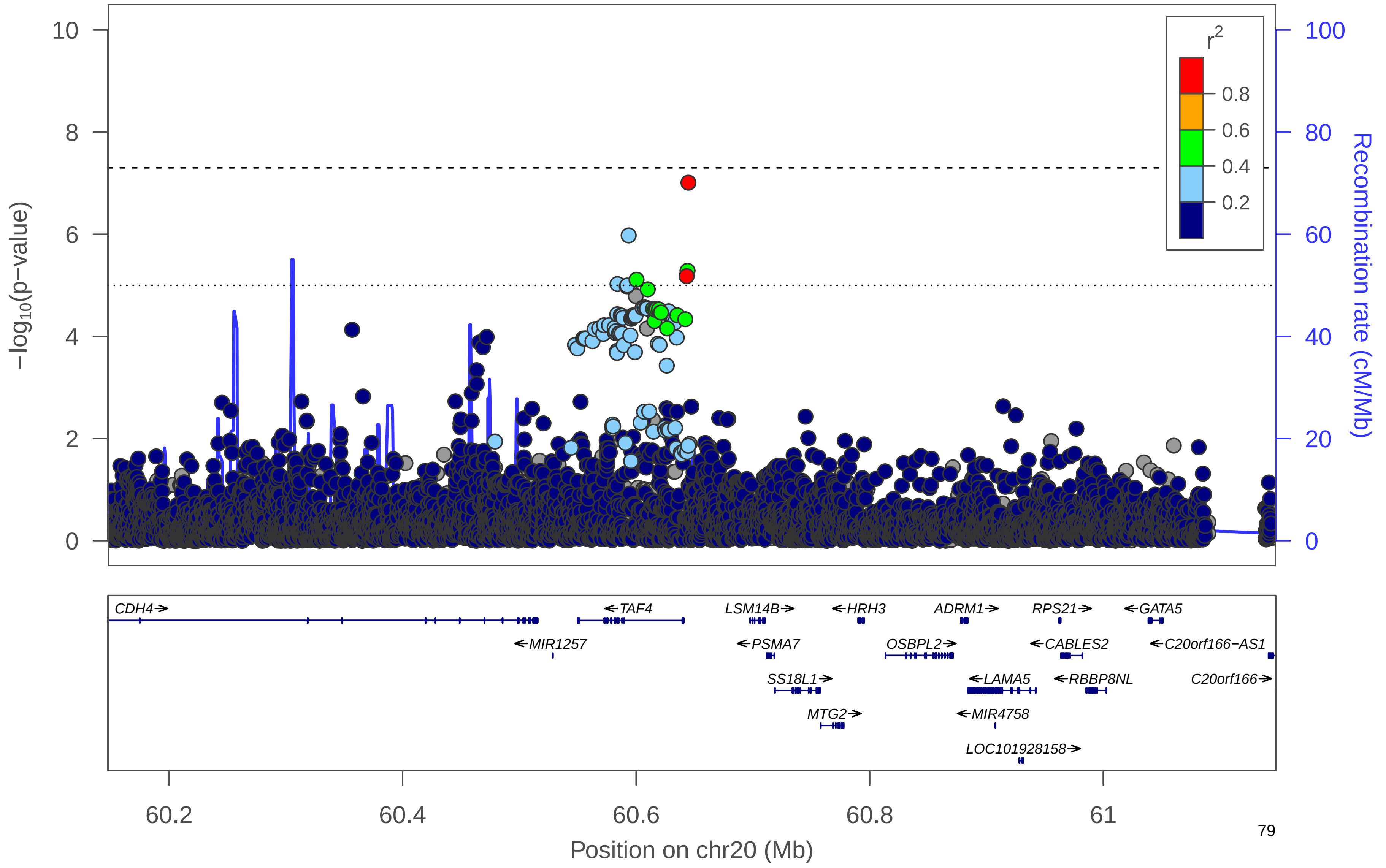

**Supplementary Figure 19. Manhattan plot.** Manhattan plot of the men-only analysis for HIPadjBMI. All suggestively significant (*P*<1x10$^{-6}$) variants are highlighted in orange if they are >500 Kb from any previously-reported HIPadjBMI associated variants. Previously reported loci (+/- 500 Kb) are highlighted in blue if any variant in the locus reached suggestive significance. All suggestively significant loci that meet our criteria for replication are annotated with the closest gene. †Replicated in African American meta-analysis. ‡ Replicated in Hispanic/Latino meta-analysis. ¥ Replicated in European American meta-analysis.

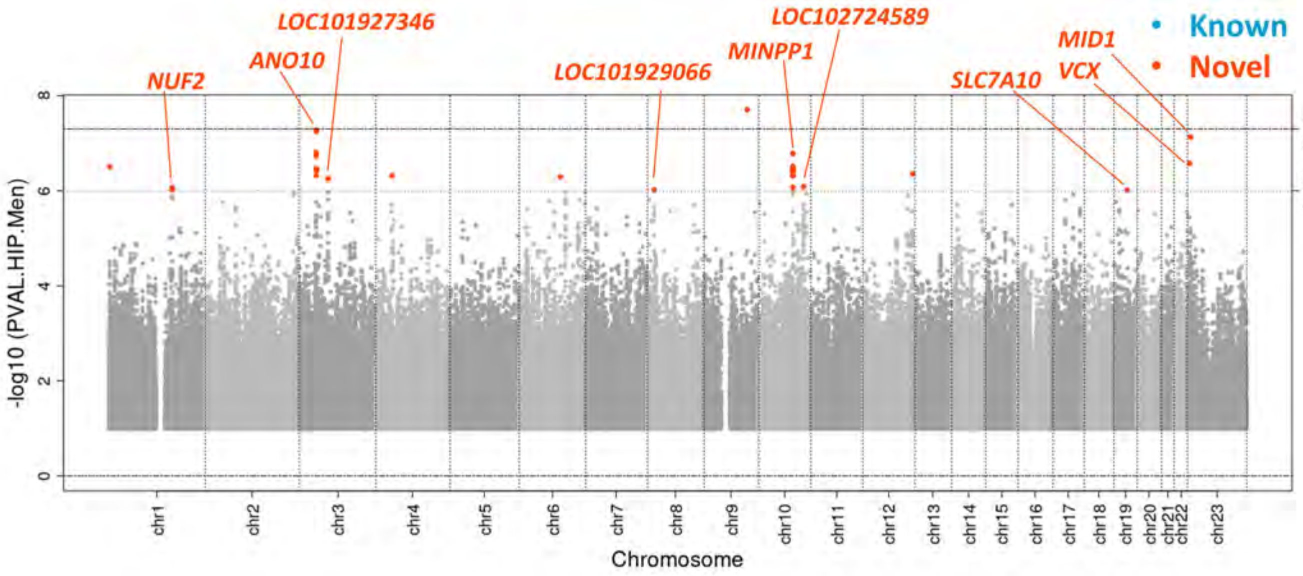

**Supplementary Figure 20. Locus Zoom Plots.** Regional association plots for suggestively significant loci in the HCHS/SOL HIPadjBMI men-only analysis. The plots appear in chromosome:position order. Dot color reflects $R^2$ calculated from the 1000 Genomes AMR reference dataset. Point symbols represent variant functional classifications: a) rs114865909, *NUF2*; b) rs149681500, *ANO10*; c) rs3915213, *LOC101927346*; d) rs12677587, *LOC101929066*; e) rs56405004, *MINPP1*; f) rs968849, *LOC102724589*; g) rs76469489, *SLC7A10*; h) rs7063750, *VCX*; i) rs112519383, *MID1*.

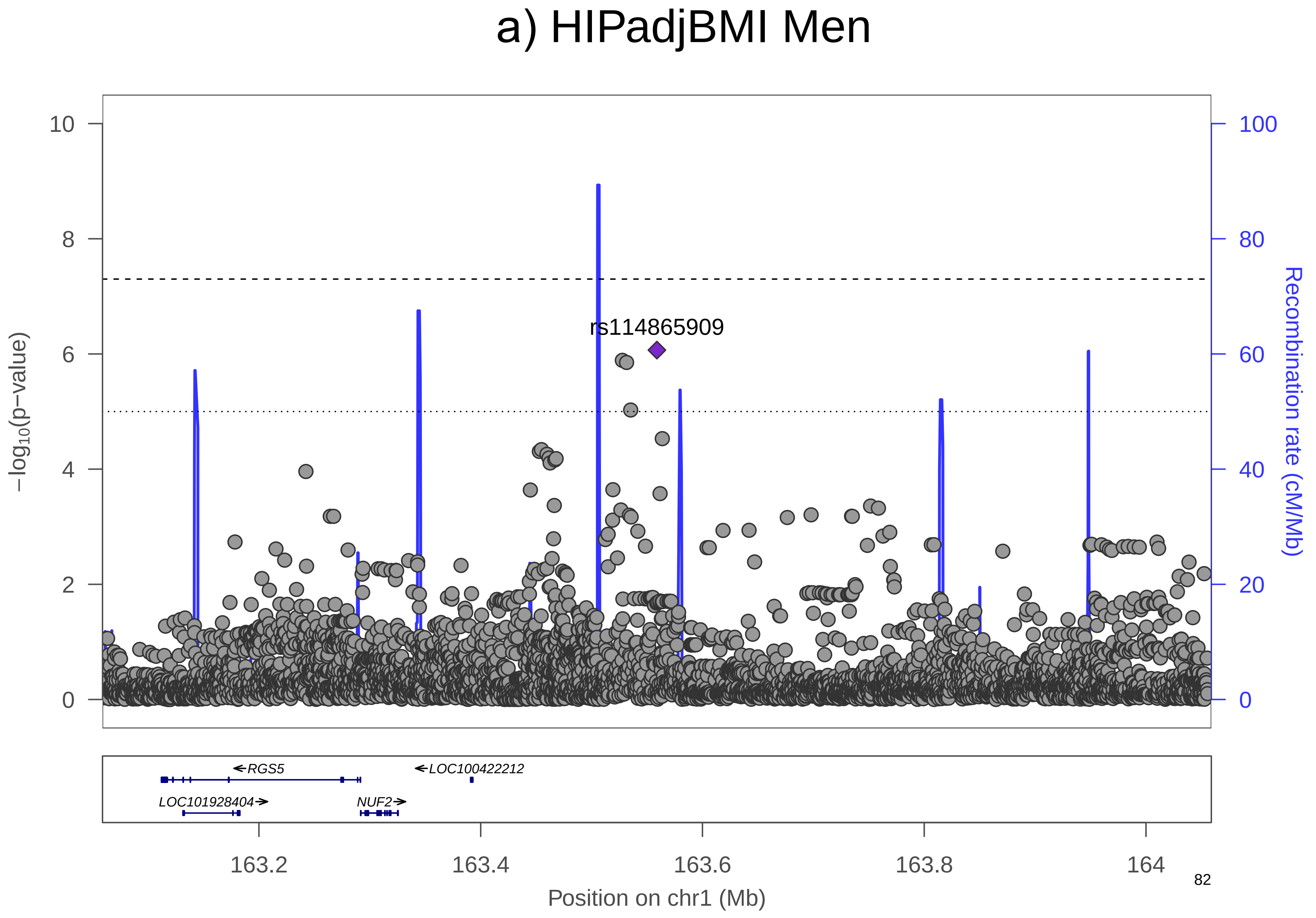

a) HIPadjBMI Men
rs114865909
−log10(p−value)
Recombination rate (cM/Mb)
RGS5
LOC100422212
LOC101928404
NUF2
163.2
163.4
163.6
163.8
164
Position on chr1 (Mb)

### b) HIPadjBMI Men

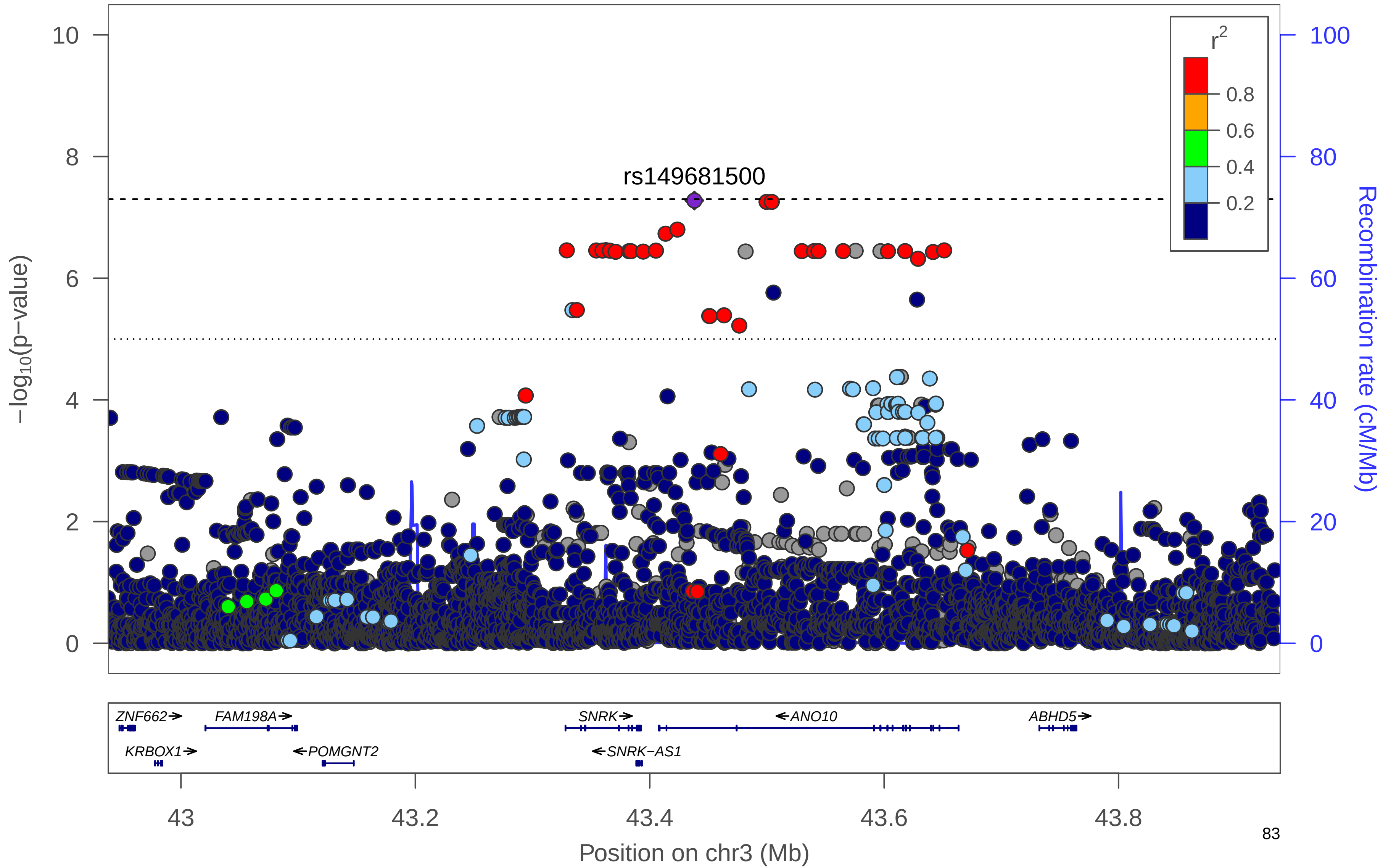

### c) HIPadjBMI Men

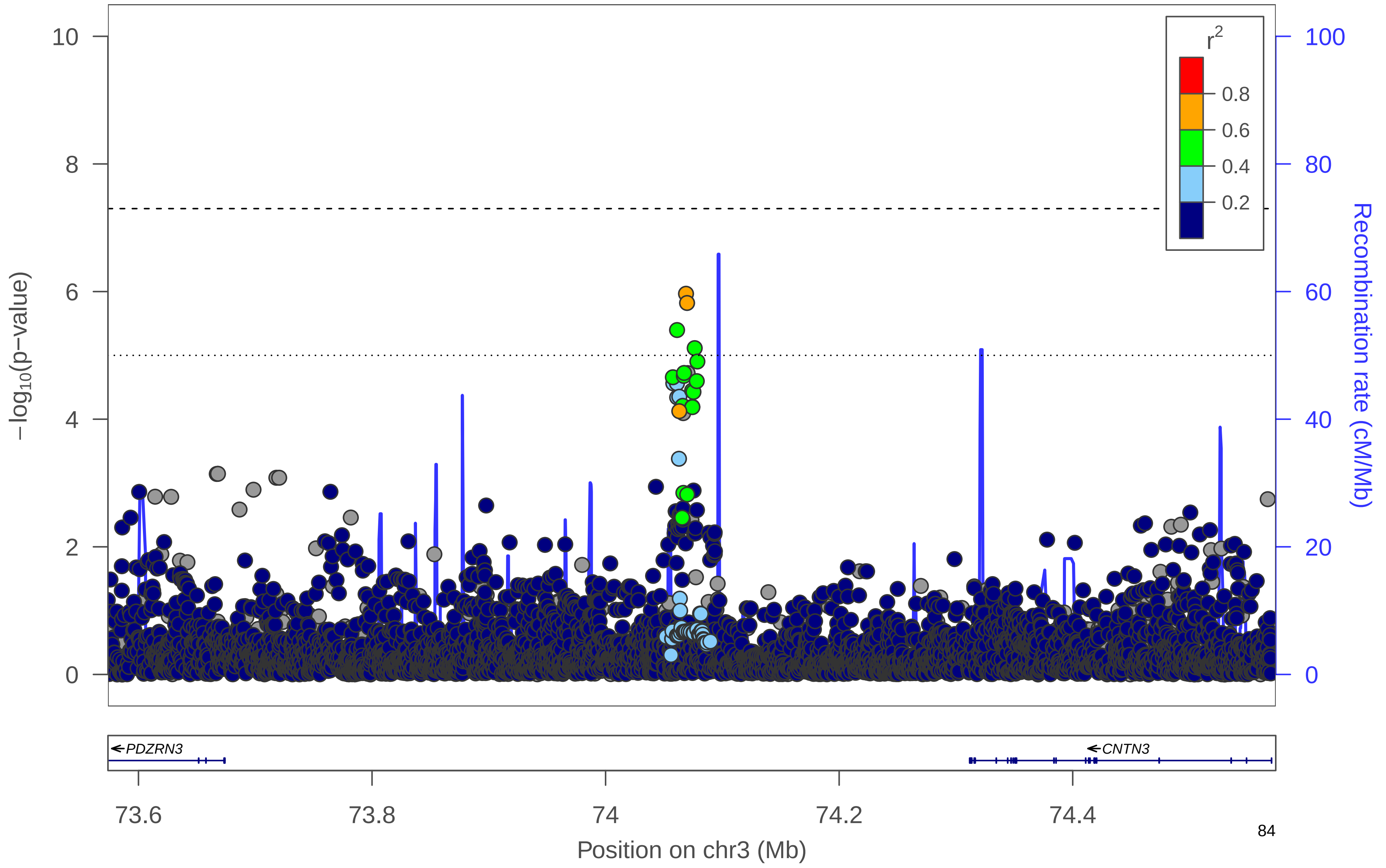

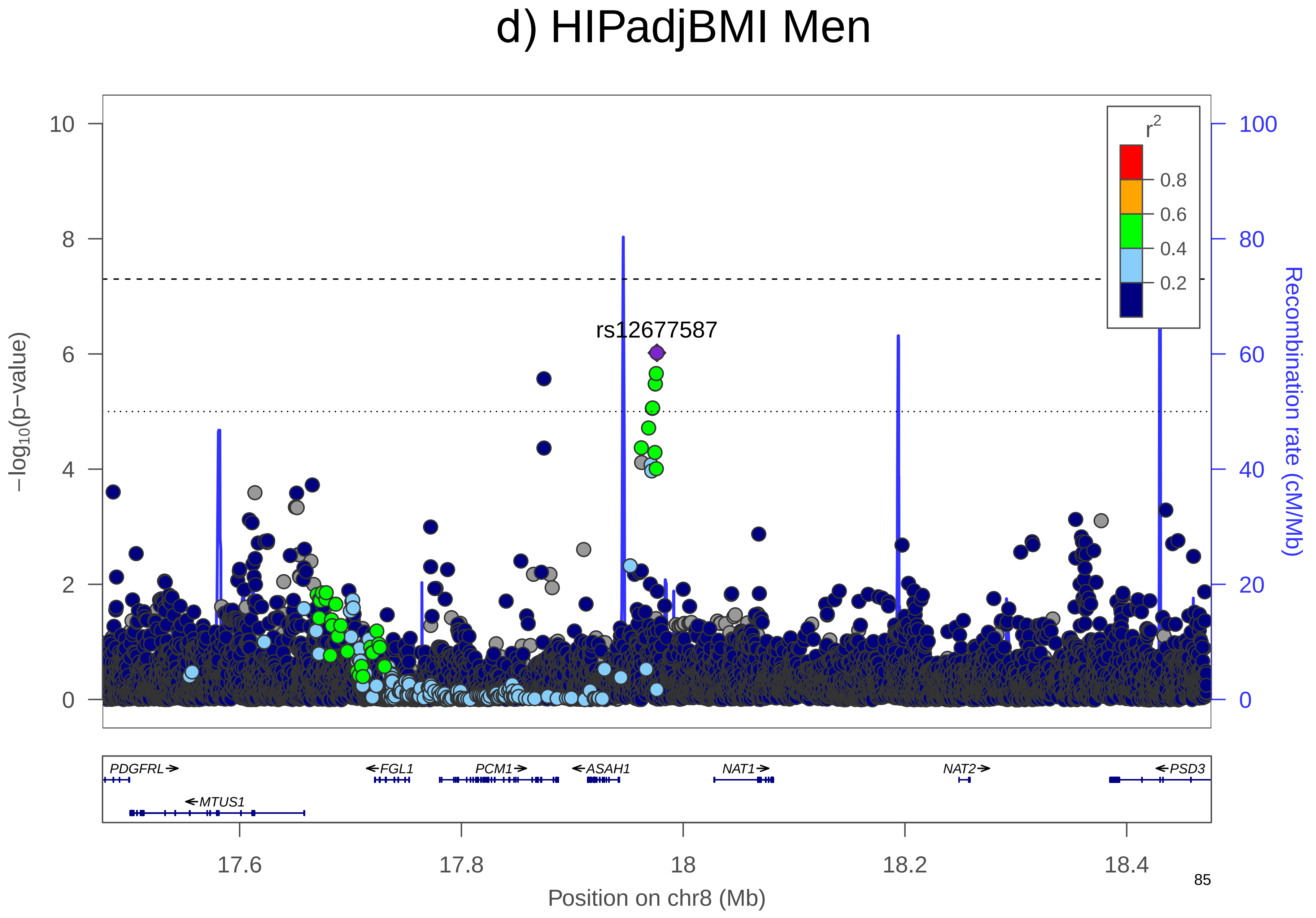
d) HIPadjBMI Men
−log10(p−value)
Recombination rate (cM/Mb)
r2
0.8
0.6
0.4
0.2
rs12677587
PDGFRL
MTUS1
FGL1
PCM1
ASAH1
NAT1
NAT2
PSD3
17.6
17.8
18
18.2
18.4
Position on chr8 (Mb)
0
2
4
6
8
10
20
40
60
80
100

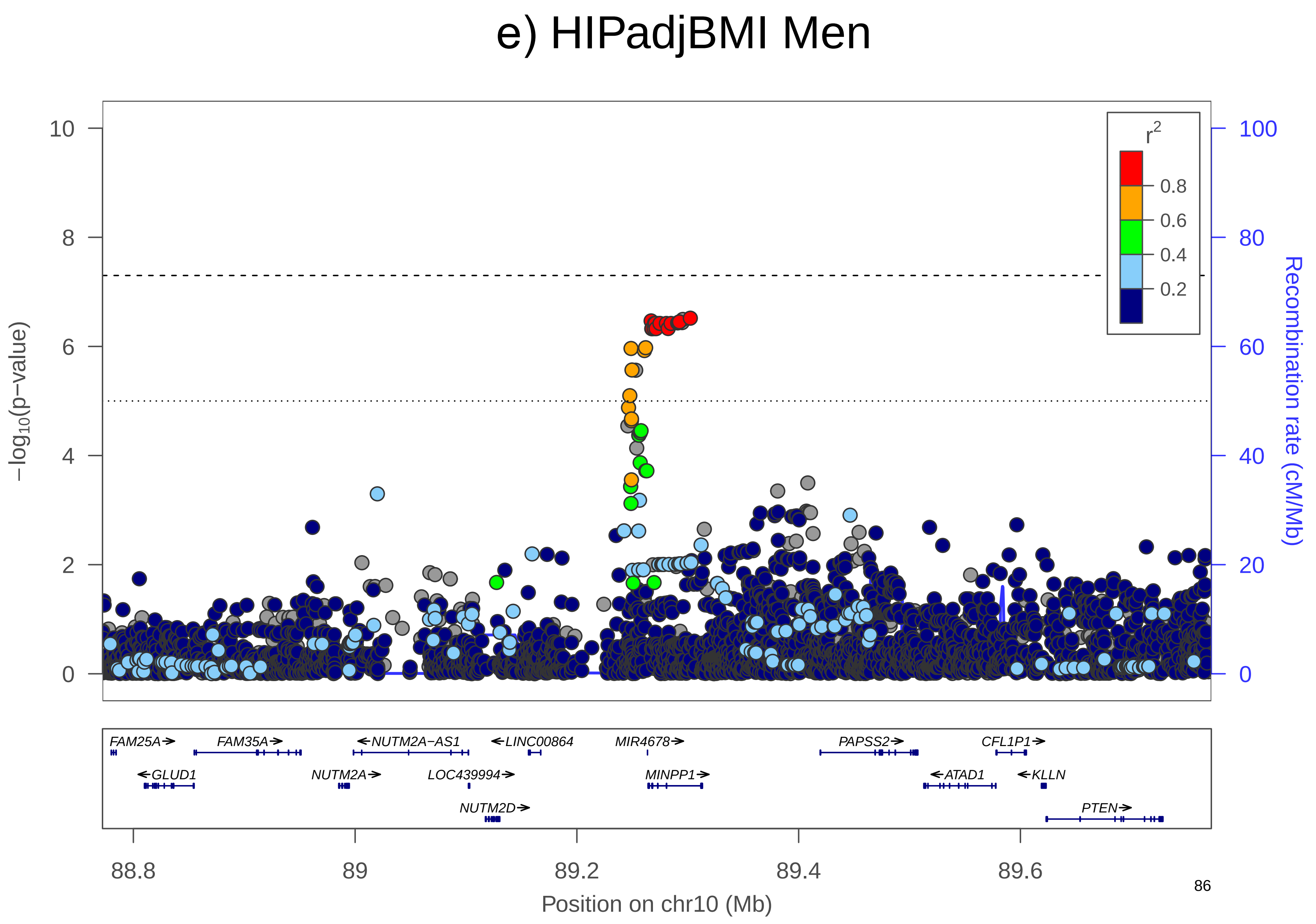

e) HIPadjBMI Men
−log10(p−value)
Recombination rate (cM/Mb)
r2
0.8
0.6
0.4
0.2
FAM25A
FAM35A
NUTM2A−AS1
LINC00864
MIR4678
PAPSS2
CFL1P1
GLUD1
NUTM2A
LOC439994
MINPP1
ATAD1
KLLN
NUTM2D
PTEN
88.8
89
89.2
89.4
89.6
Position on chr10 (Mb)

f) HIPadjBMI Men

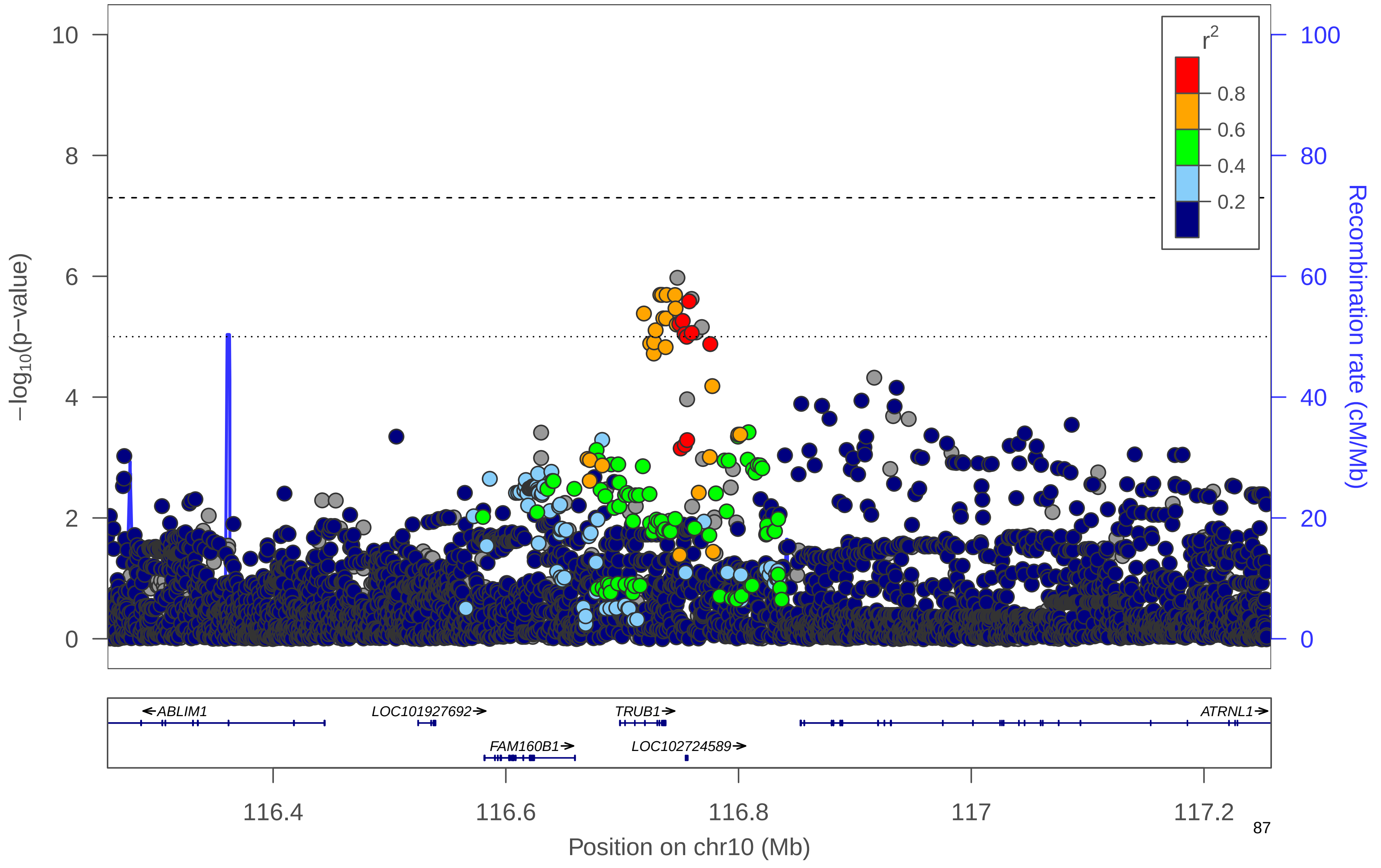

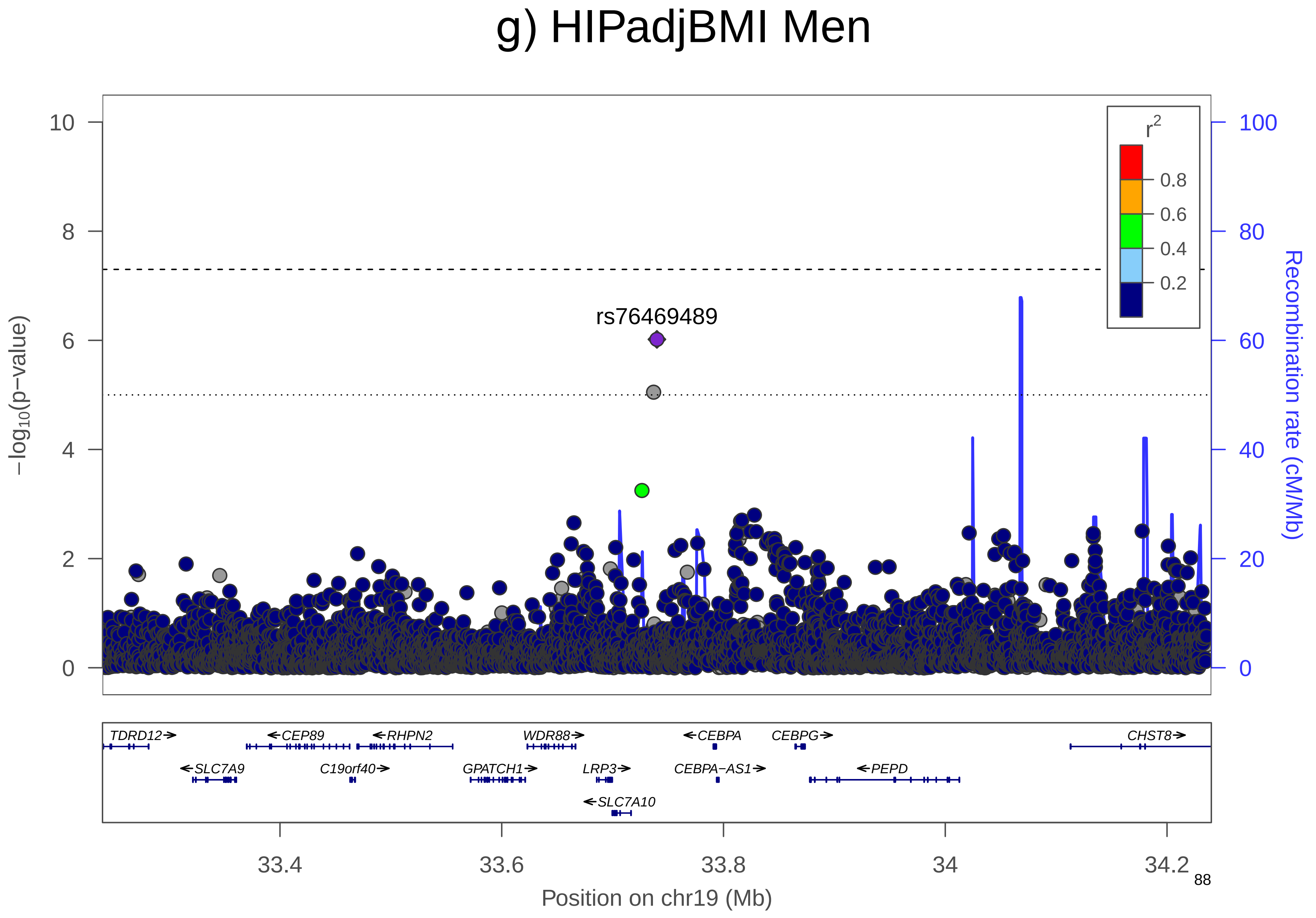

g) HIPadjBMI Men
rs76469489
r2
0.8
0.6
0.4
0.2
−log10(p−value)
Recombination rate (cM/Mb)
TDRD12
CEP89
RHPN2
WDR88
CEBPA
CEBPG
CHST8
SLC7A9
C19orf40
GPATCH1
LRP3
CEBPA−AS1
PEPD
SLC7A10
33.4
33.6
33.8
34
34.2
Position on chr19 (Mb)

### h) HIPadjBMI Men

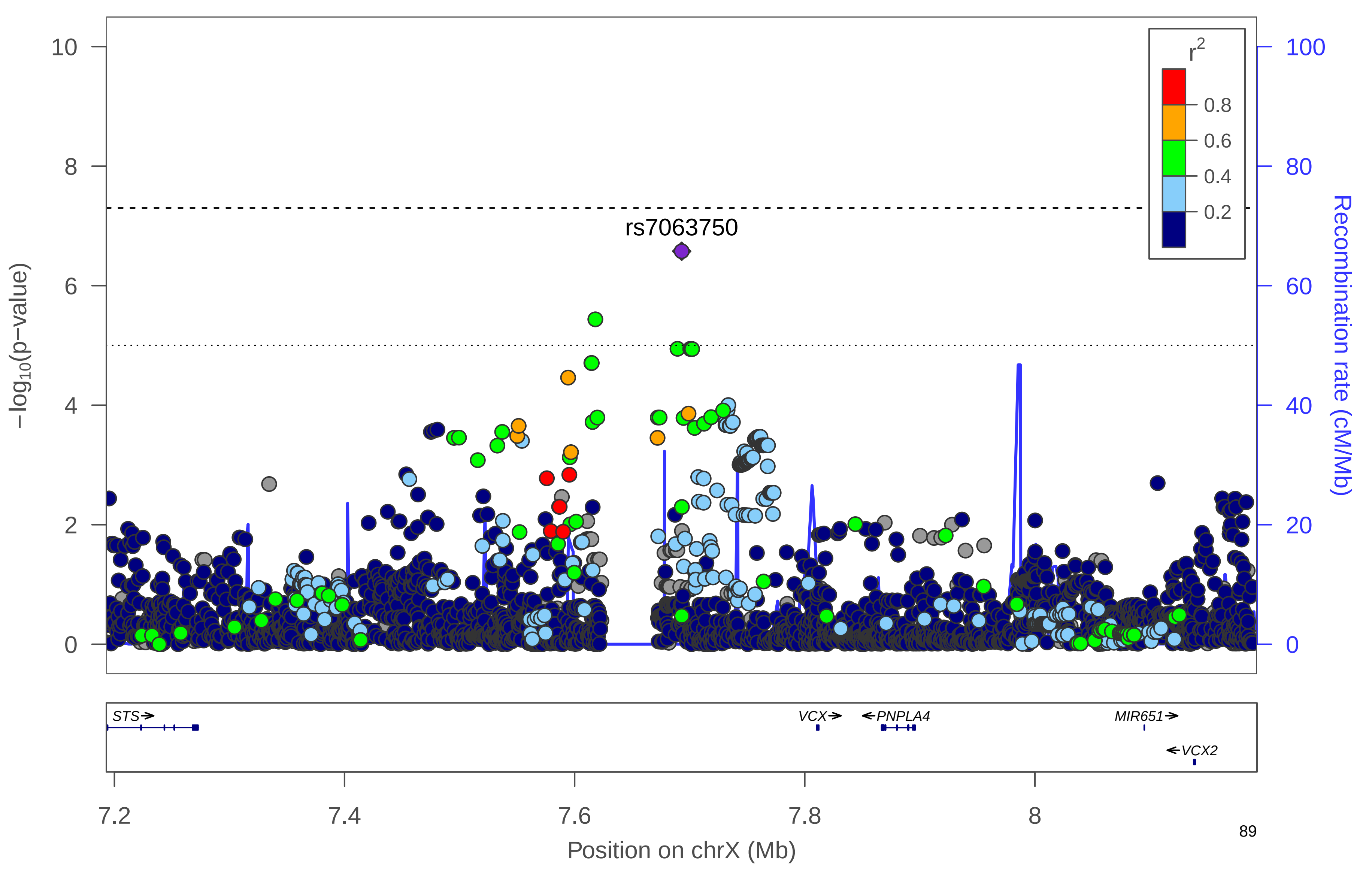

### i) HIPadjBMI Men

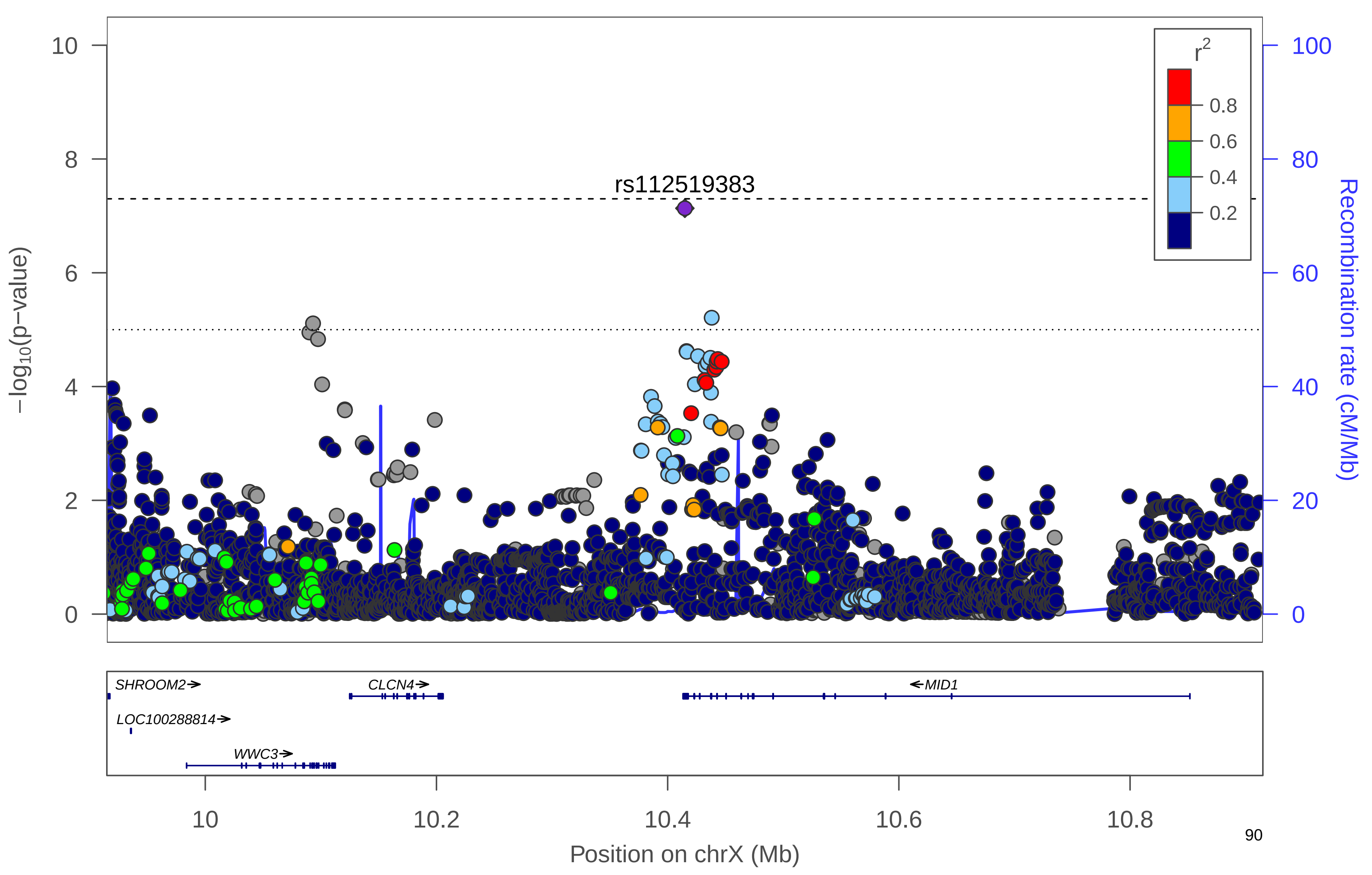

**Supplementary Figure 21. QQ Plots**. QQ Plots for HIPadjBMI, including sexes-combined (black), women-only (orange), men-only (blue).

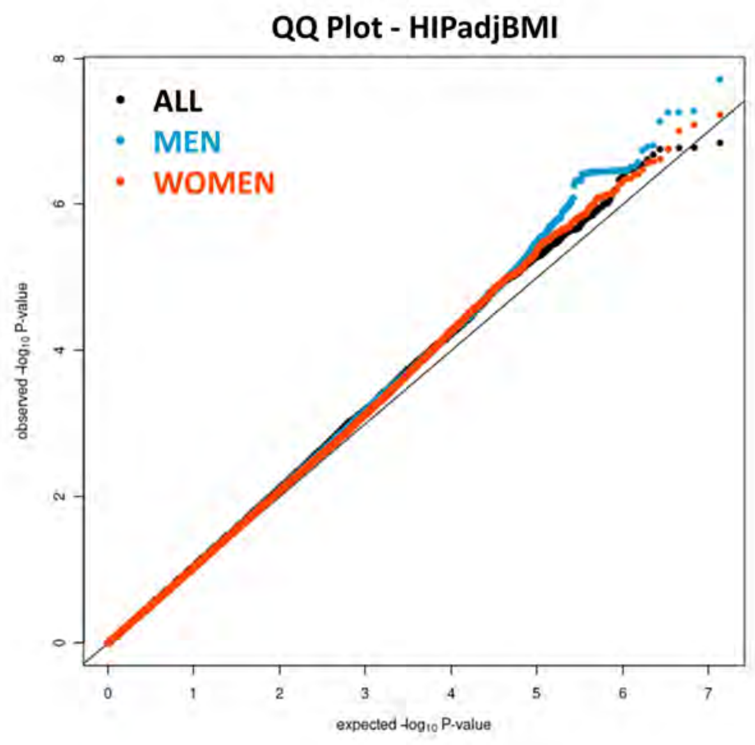

**Supplementary Figure 22. Generalization of Known WHRadjBMI Loci.** Comparison of the effect size estimates of known loci associated with WHRadjBMI in the sexes-combined analysis for GIANT (Shungin et al. 2015), all-ancestry GWAS, and the HCHS/SOL. Estimates are shown for all lead-gen SNPS with r-value < 0.1. Lead-gen SNPs are the SNPs with smallest r-value in a known locus. Loci were defined as regions of 1MB. Generalized SNPs (r-value<0.05) are highlighted in blue and non-generalized SNPs are highlighted in red.

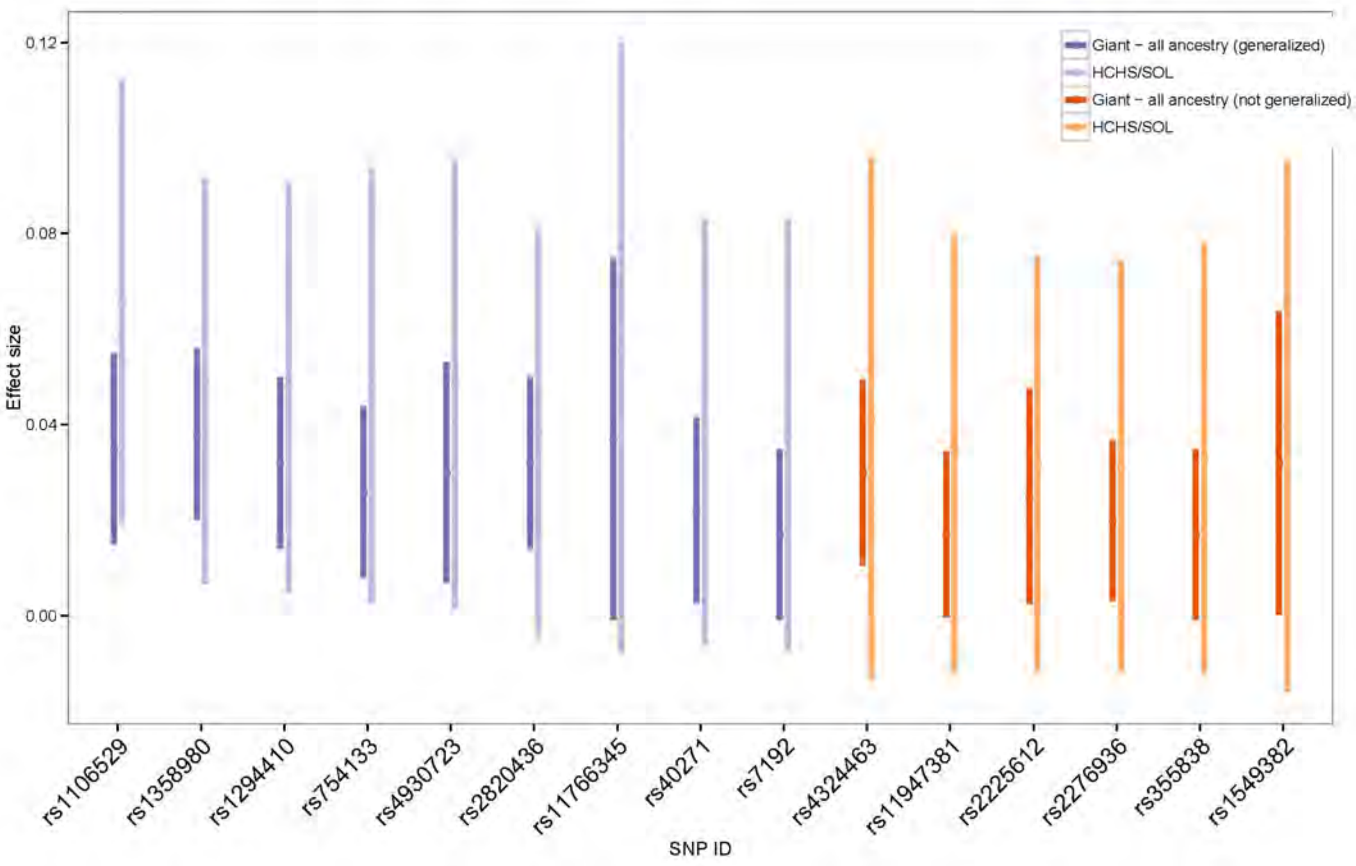

**Supplementary Figure 23. Generalization of Known WHRadjBMI Loci.** Comparison of the effect size estimates of known loci associated with WHRadjBMI in the women-only analysis for GIANT (Shungin et al. 2015), all-ancestry GWAS, and the HCHS/SOL women-only analysis. Estimates are shown for all lead-gen SNPS with r-value < 0.1. Lead-gen SNPs are the SNPs with smallest r-value in a known locus. Loci were defined as regions of 1MB. Generalized SNPs (r-value<0.05) are highlighted in blue and non-generalized SNPs are highlighted in red.

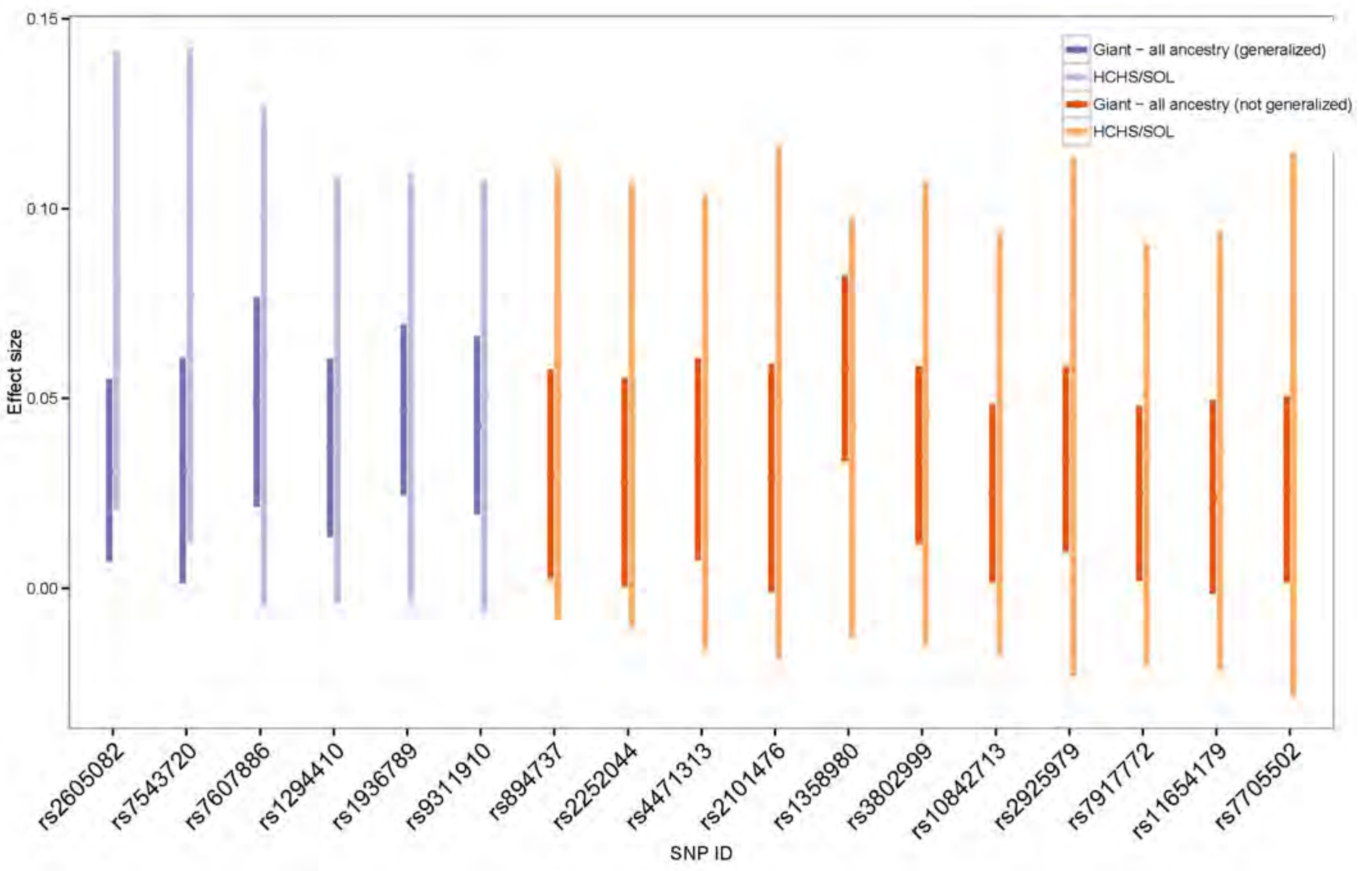

**Supplementary Figure 24. Generalization of Known WHRadjBMI Loci.** Comparison of the effect size estimates of known loci associated with WHRadjBMI in the men-only analysis for GIANT (Shungin et al. 2015), all-ancestry GWAS, and the HCHS/SOL men-only analysis. Estimates are shown for all lead-gen SNPS with r-value < 0.1. Lead-gen SNPs are the SNPs with smallest r-value in a known locus. Loci were defined as regions of 1MB. Generalized SNPs (r-value<0.05) are highlighted in blue and non-generalized SNPs are highlighted in red.

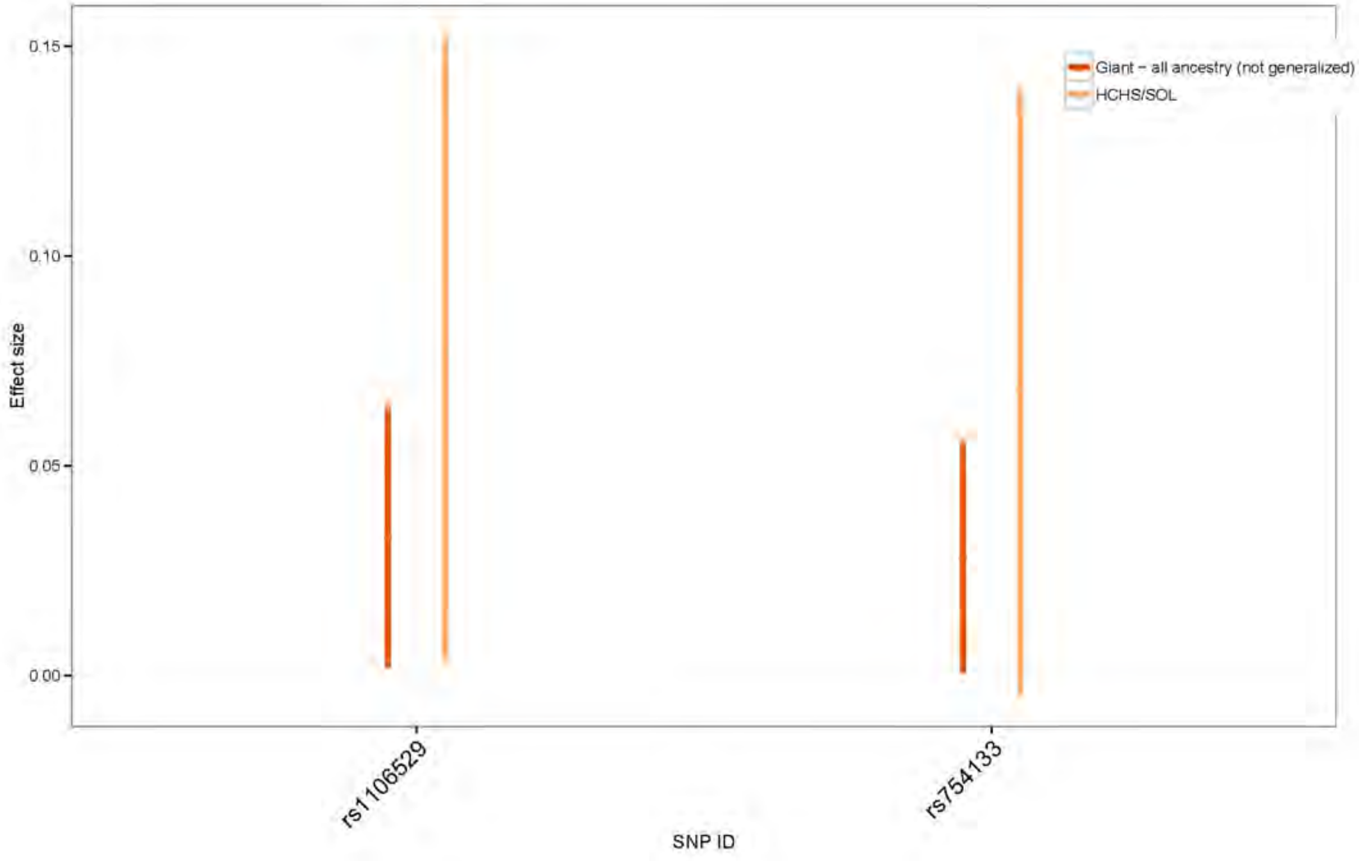

**Supplementary Figure 25. Generalization of Known WCadjBMI Loci.** Comparison of the effect size estimates of known loci associated with WCadjBMI in the sexes-combined analysis for GIANT (Shungin et al. 2015), all-ancestry GWAS, and the HCHS/SOL sexes-combined analysis. Estimates are shown for all lead-gen SNPS with r-value < 0.1. Lead-gen SNPs are the SNPs with smallest r-value in a known locus. Loci were defined as regions of 1MB. Generalized SNPs (r-value<0.05) are highlighted in blue and non-generalized SNPs are highlighted in red.

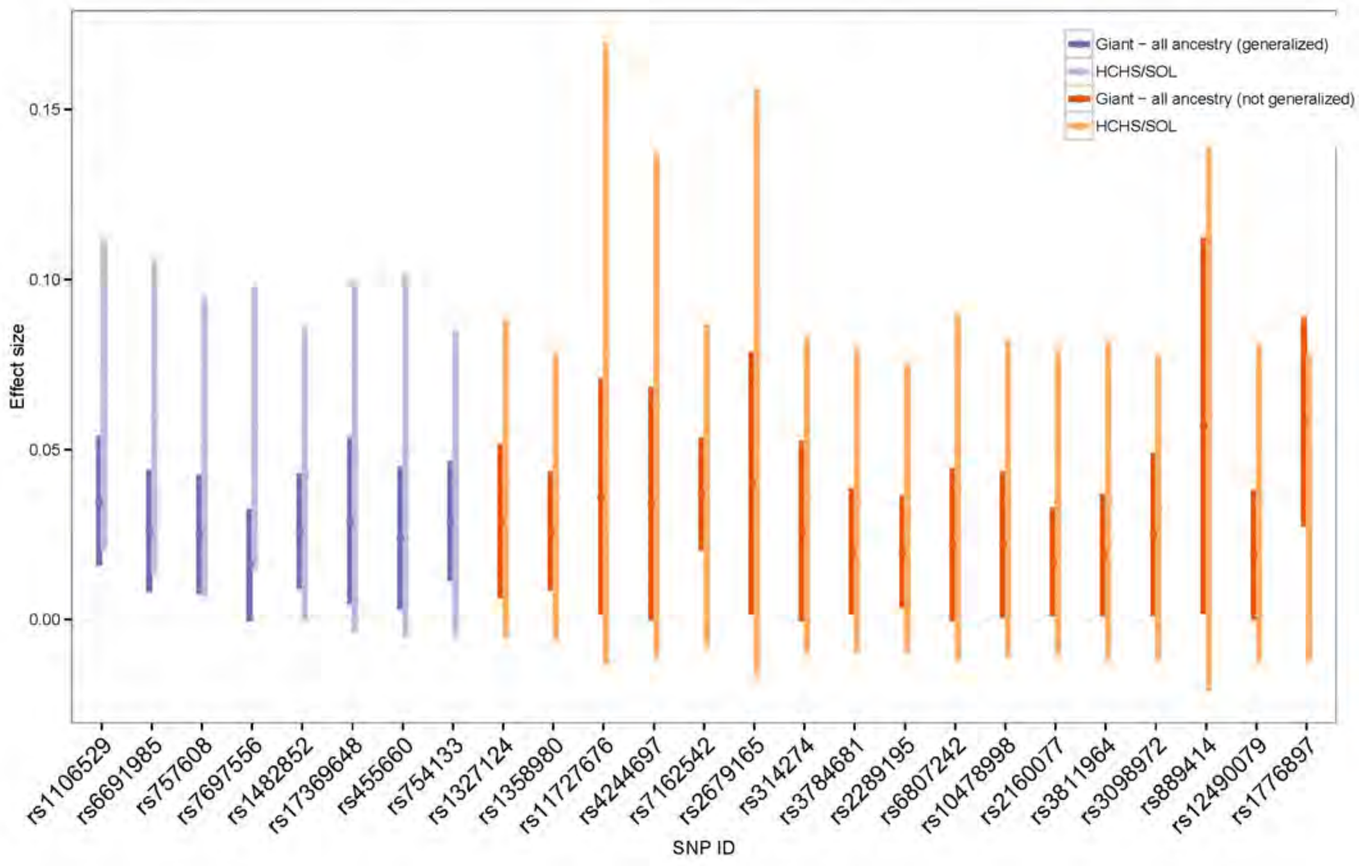

**Supplementary Figure 26. Generalization of Known WCadjBMI Loci.** Comparison of the effect size estimates of known loci associated with WCadjBMI in the women-only analysis for GIANT (Shungin et al. 2015), all-ancestry GWAS, and the HCHS/SOL women-only analysis. Estimates are shown for all lead-gen SNPS with r-value < 0.1. Lead-gen SNPs are the SNPs with smallest r-value in a known locus. Loci were defined as regions of 1MB. Generalized SNPs (r-value<0.05) are highlighted in blue and non-generalized SNPs are highlighted in red.

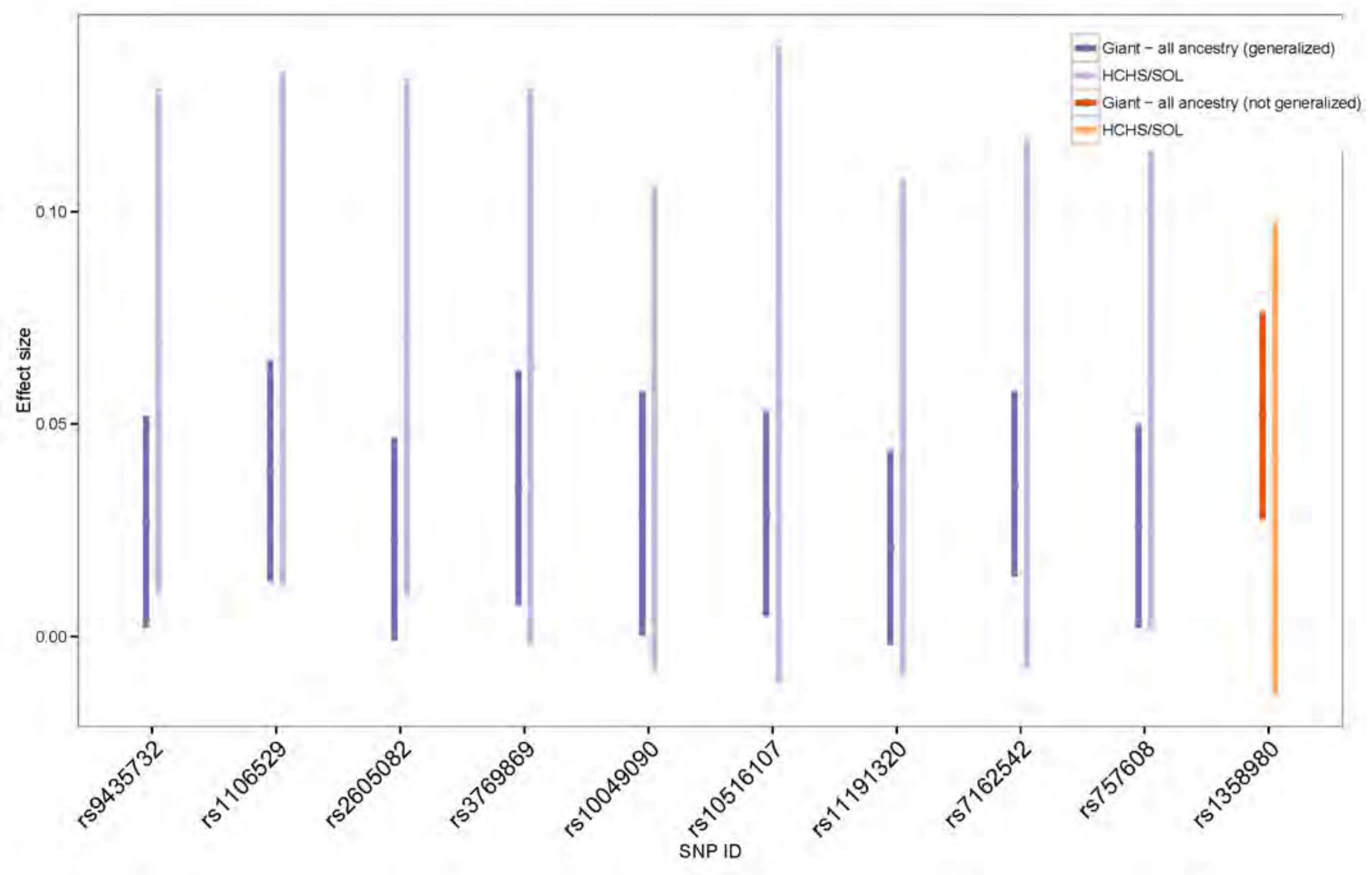

**Supplementary Figure 27. Generalization of Known WCadjBMI Loci.** Comparison of the effect size estimates of known loci associated with WCadjBMI in the men-only analysis for GIANT (Shungin et al. 2015), all-ancestry GWAS, and the HCHS/SOL men-only analysis. Estimates are shown for all lead-gen SNPS with r-value < 0.1. Lead-gen SNPs are the SNPs with smallest r-value in a known locus. Loci were defined as regions of 1MB. Generalized SNPs (r-value<0.05) are highlighted in blue and non-generalized SNPs are highlighted in red.

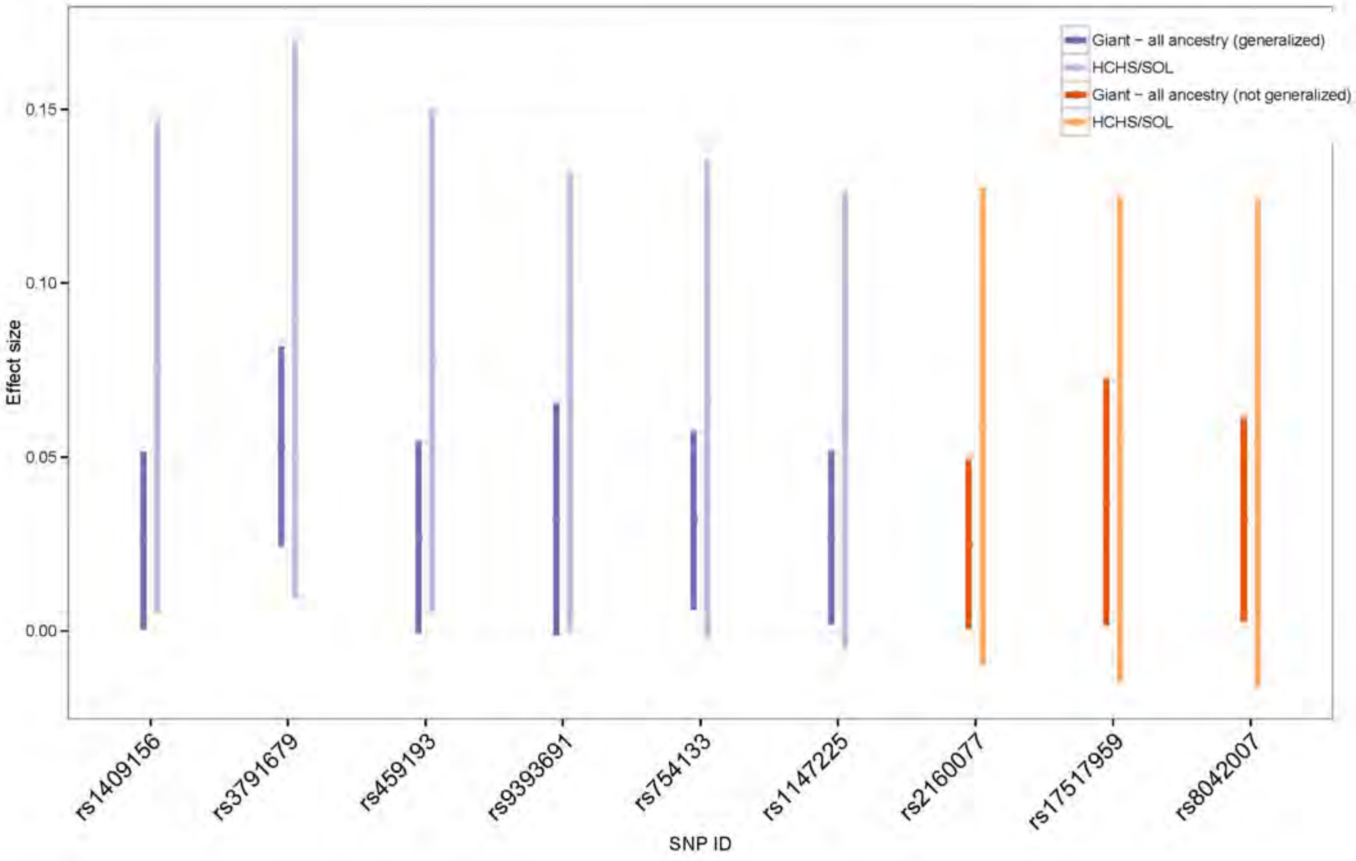

**Supplementary Figure 28. Generalization of Known HIPadjBMI Loci.** Comparison of the effect size estimates of known loci associated with HIPadjBMI in the sexes-combined analysis for GIANT (Shungin et al. 2015), all-ancestry GWAS, and the HCHS/SOL sexes-combined analysis. Estimates are shown for all lead-gen SNPS with r-value < 0.1. Lead-gen SNPs are the SNPs with smallest r-value in a known locus. Loci were defined as regions of 1MB. Generalized SNPs (r-value<0.05) are highlighted in blue and non-generalized SNPs are highlighted in red.

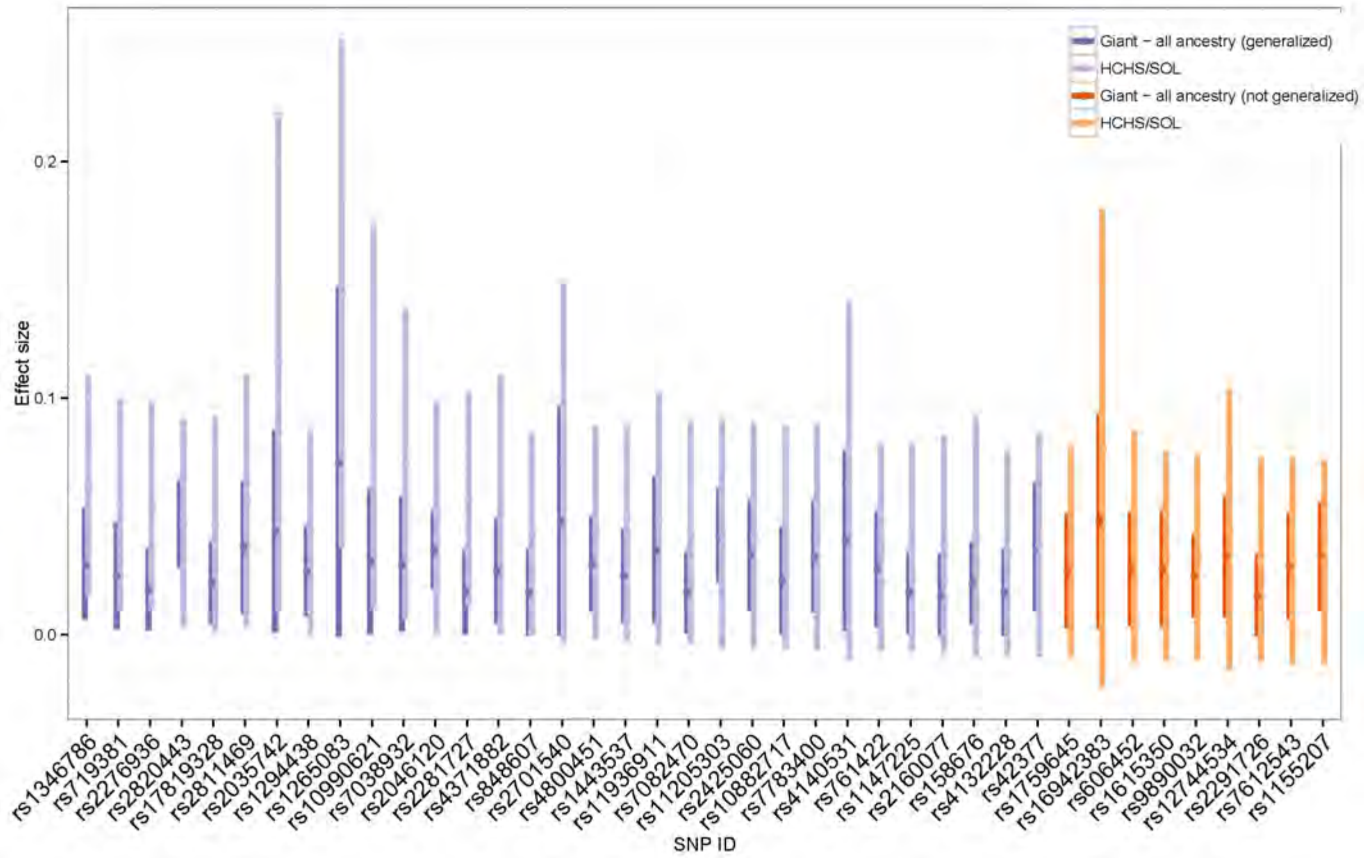

**Supplementary Figure 29. Generalization of Known HIPadjBMI Loci.** Comparison of the effect size estimates of known loci associated with HIPadjBMI in the women-only analysis for GIANT (Shungin et al. 2015), all-ancestry GWAS, and the HCHS/SOL women-only analysis. Estimates are shown for all lead-gen SNPS with r-value < 0.1. Lead-gen SNPs are the SNPs with smallest r-value in a known locus. Loci were defined as regions of 1MB. Generalized SNPs (r-value<0.05) are highlighted in blue and non-generalized SNPs are highlighted in red.

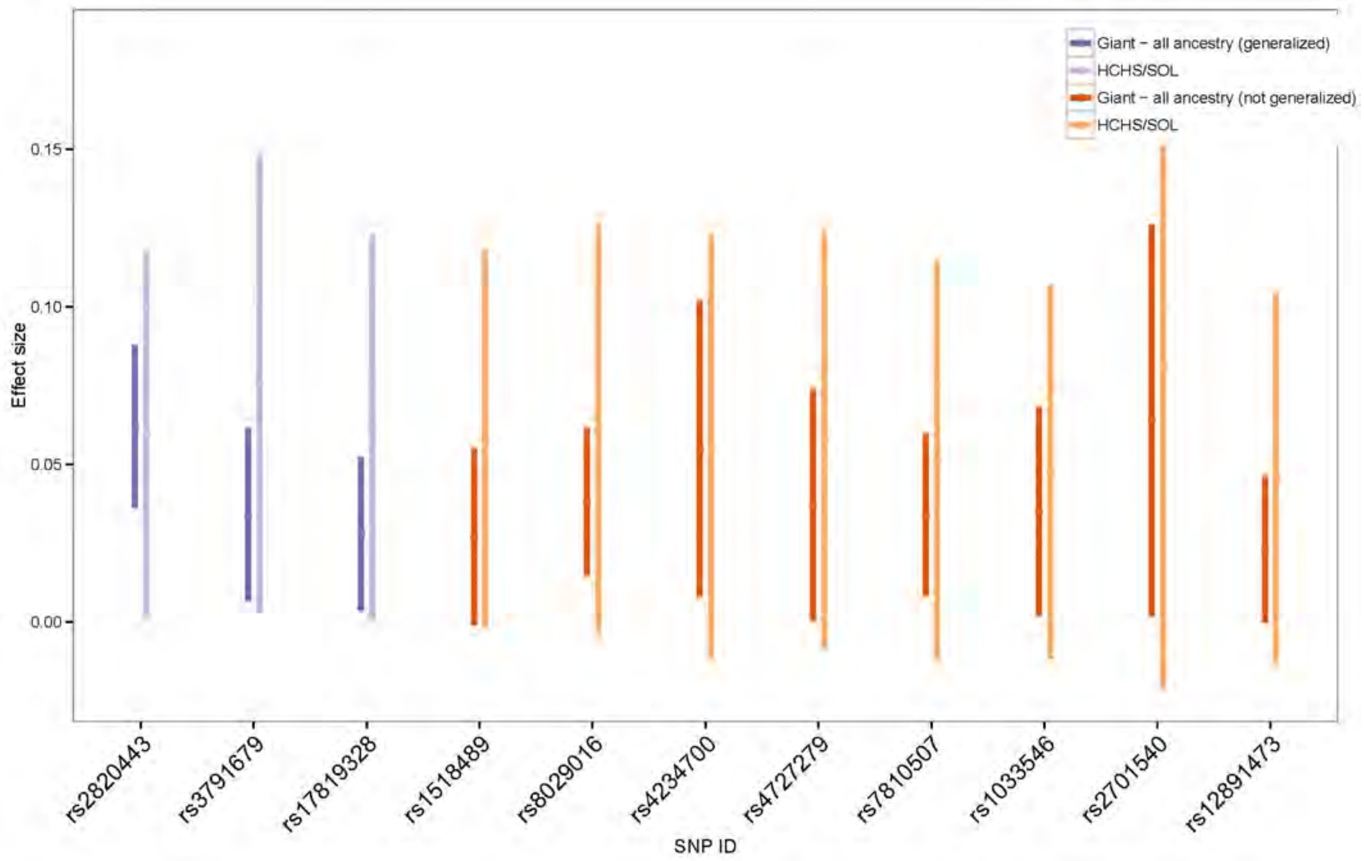

**Supplementary Figure 30. Generalization of Known HIPadjBMI Loci.** Comparison of the effect size estimates of known loci associated with HIPadjBMI in the men-only analysis for GIANT (Shungin et al. 2015), all-ancestry GWAS, and the HCHS/SOL men-only analysis. Estimates are shown for all lead-gen SNPS with r-value < 0.1. Lead-gen SNPs are the SNPs with smallest r-value in a known locus. Loci were defined as regions of 1MB. Generalized SNPs (r-value<0.05) are highlighted in blue and non-generalized SNPs are highlighted in red.

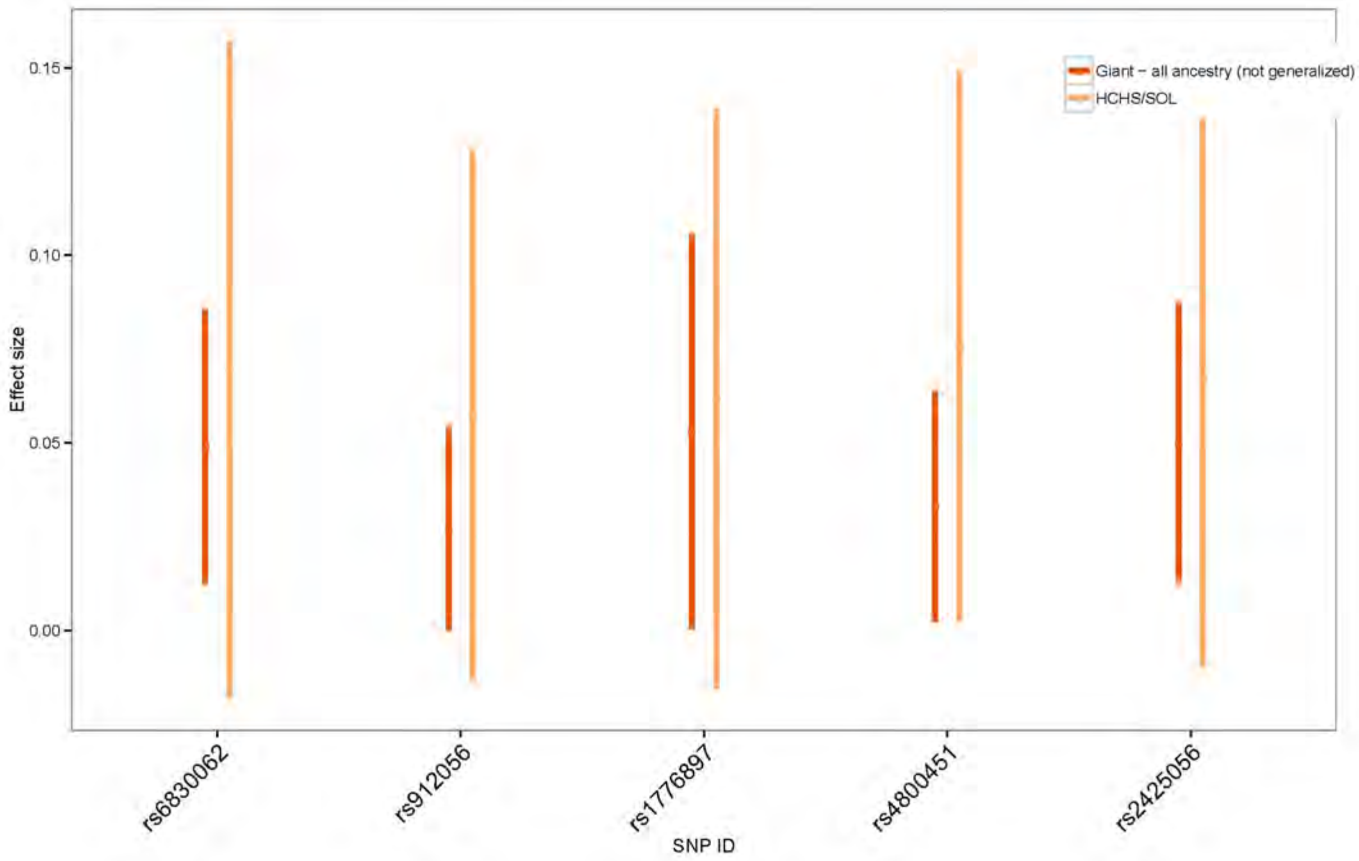

**Supplementary Table 1.** Study-specifc descriptive statistics for discovery and replication cohorts. Abbreviations: WC - waist circumference; HIP - hip circumference; WHR - waist-to-hip ratio; BMI - body mass index.

| Study name | Study Abbreviation | Study design | Call rate | Study-specific exclusions | For HL, % ancestry background for study sample (Mainland, Caribbean) | Measured/ Self-reported | MEN | | | | | | | | | | | | WOMEN | | | | | | | | | | | | Analysis software | | | | |
|---|---|---|---|---|---|---|---|---|---|---|---|---|---|---|---|---|---|---|---|---|---|---|---|---|---|---|---|---|---|---|---|---|---|---|---|
| | | | | | | | WC | | | HIP | | | WHR | | | BMI | | | WC | | | HIP | | | WHR | | | BMI | | | Analysis software - phenotype | Analysis software – SNP associations | Genotyping platform | Imputation software | References for Study Design |
| | | | | | | | N | Mean | SD | N | Mean | SD | N | Mean | SD | N | Mean | SD | N | Mean | SD | N | Mean | SD | N | Mean | SD | N | Mean | SD | | | | | |
| **Hispanic/Latino** | | | | | | | | | | | | | | | | | | | | | | | | | | | | | | | | | | | |
| Genetics of Latinos Diabetic Retinopathy | GOLDR | families recruited through diabete patients with eye disease | >=90% | Family QC, Gender mismatch | 98.13% Mainland, 1.87% Caribbean | measured | 219 | 103.90 | 14.58 | 54 | 103.06 | 13.44 | 54 | 0.98 | 0.06 | 223 | 31.03 | 6.25 | 371 | 107.32 | 14.32 | 84 | 110.38 | 15.72 | 84 | 0.95 | 0.05 | 374 | 32.63 | 7.02 | SAS 9.4 | R (GWAF) | Omni Express | SHAPEIT, Minimac3 | PMCID: 3343221 |
| Hispanic Community Health Study / Study of Latinos | HCHS/SOL | Population based cohort | >=98% | Gender mismatch; identity issues; PCA outliers | 55.51% Mainland, 44.49% Caribbean | measured | 5,200 | 99.10 | 13.41 | 5,200 | 103.58 | 10.08 | 5,200 | 0.96 | 0.07 | 5,200 | 29.07 | 5.32 | 7,272 | 97.76 | 14.36 | 7,272 | 108.42 | 13.14 | 7,272 | 0.90 | 0.07 | 7,272 | 30.34 | 6.48 | R 3.2.0 | R (GENESIS) | HumanOmni2.5-8v1-1 + custom | IMPUTE2 | PMIDs: 20609343, |
| Mexican–American Hypertension Study | HTN | families recruited through hypertensive probands. | >=95% | Family QC, Gender mismatch, Bad Concordance OMNI1 and 1S | 90.43% Mainland, 9.57% Caribbean | measured | 253 | 95.04 | 14.73 | 248 | 101.71 | 9.76 | 248 | 0.94 | 0.07 | 312 | 28.91 | 5.24 | 362 | 89.70 | 14.48 | 358 | 106.21 | 11.77 | 357 | 0.85 | 0.07 | 451 | 29.52 | 6.06 | SAS 9.4 | R (GWAF) | Omni Express + 1s | SHAPEIT, IMPUTEv2.3.0 | PMID: 11136689 |
| Mexican-American Coronary Artery Disease | MACAD | CAD proband, spouse of proband, and adult offspring and spouses of offspring | >=95% | Family QC, Gender mismatch, Bad Concordance OMNI1 and 1S | 94.84 Mainland, 5.16% Caribbean | measured | 311 | 96.53 | 11.40 | 311 | 103.41 | 8.92 | 311 | 0.93 | 0.06 | 346 | 28.92 | 4.52 | 417 | 90.46 | 13.19 | 416 | 105.69 | 11.89 | 416 | 0.86 | 0.07 | 457 | 29.00 | 5.58 | SAS 9.4 | R (GWAF) | Omni Express + 1s | SHAPEIT, IMPUTEv2.3.0 | PMID: 14693718 |
| Multi-Ethnic Study of Atherosclerosis | MESA | Population based | >=95% | Gender mismatch; PCA outliers | %67.8 Mexico ; 32.2% Caribbean | measured | 720 | 100.92 | 11.43 | 720 | 102.67 | 8.83 | 720 | 0.98 | 0.05 | 720 | 28.80 | 4.35 | 777 | 100.87 | 14.72 | 777 | 108.13 | 11.93 | 777 | 0.93 | 0.07 | 777 | 30.17 | 5.78 | SAS | SNPTEST2 | AFFY6.0 | IMPUTE2 | PMID: 12397006; Am J Epidemiol. 2002 Nov |
| Mexico-City Diabetes study | Mexico-City Cases | Case/Control | >=99% | Related individuals; PCA outliers; Gender misn | 100% Mexico | measured | 360 | 99.03 | 12.42 | 360 | 101.51 | 10.02 | 360 | 0.97 | 0.07 | 360 | 28.59 | 5.15 | 534 | 95.82 | 12.14 | 534 | 106.83 | 12.07 | 534 | 0.90 | 0.06 | 535 | 30.40 | 5.48 | SPSS | SNPTEST | Affymetrix Axiom LAT | SHAPEIT; IMPUTE2 | PMID: 26780889 |
| 1982 Pelotas Birth Cohort | PELOTAS | Prospective, population-ba | ≥95% | Samples excluded if there were sex mismatches (heterozygosity threshold: 0.02), heterozygosity rate outside the range of median ± 1.5 x IQR, missingness >3% and cryptic relatedness (kinship>0.1, as described in PMID: ). | NA | measured | 1,362 | 89.42 | 11.87 | 1,362 | 102.76 | 9.28 | 1,362 | 0.87 | 0.06 | 1,351 | 27.10 | 5.08 | 1,436 | 80.87 | 12.17 | 1,435 | 105.95 | 10.88 | 1,435 | 0.76 | 0.07 | 1,432 | 26.82 | 6.10 | R 3.2.0 | R 3.2.0 | HumanOmni2.5-8v1 (Illumina) | IMPUTE2 | PMID: 16373375; PMID: 25733577 |
| Starr County Health Studies | STARR - Cases | Case/Control | 0.9995 | None | 31% Native American | measured | 274 | 106.43 | 13.22 | 273 | 105.54 | 10.29 | 273 | 1.01 | 0.06 | 328 | 30.17 | 5.39 | 413 | 112.14 | 15.57 | 413 | 114.09 | 14.26 | 413 | 0.98 | 0.07 | 489 | 32.84 | 6.80 | Excel | SNPTEST | Affymetrix Genome-Wide Human SNP | IMPUTE2 | PMIDs: 8640221, 21647700 |
| Starr County Health Studies | STARR - Contr | Case/Control | 0.9995 | None | 31% Native American | measured | 223 | 100.86 | 12.88 | 223 | 106.11 | 9.30 | 223 | 0.95 | 0.06 | 223 | 29.37 | 5.27 | 556 | 94.79 | 15.08 | 555 | 110.13 | 13.26 | 553 | 0.86 | 0.07 | 558 | 30.45 | 6.58 | Excel | SNPTEST | Affymetrix Genome-Wide Human SNP | IMPUTE2 | PMIDs: 8640221, 21647700 |
| Women's Health Initiative | WHI | Prospective cohort study | >=95% | first-degree relatives | Unknown | measured | NA | NA | NA | NA | NA | NA | NA | NA | NA | NA | NA | NA | 3,420 | 86.46 | 12.21 | 3,420 | 105.75 | 11.01 | 3,420 | 0.82 | 0.07 | 3,420 | 28.85 | 5.68 | STATA (v14) | PLINK/ Probable | AFFY6.0 | MACH v1.0.16 | PMID: 9492970 |
| **African American** | | | | | | | | | | | | | | | | | | | | | | | | | | | | | | | | | | | |
| Atherosclerosis Risk in Communities Study | ARIC | Population based | >=90% | first-degree relatives ; ancestry outliers ; gender mismatch; identity issues; excessive heterozygosity; missing height or weight | N/A | measured | 1,056 | 97.45 | 11.88 | 1,056 | 103.25 | 8.70 | 1,056 | 0.94 | 0.05 | 1,056 | 27.91 | 4.59 | 1,773 | 99.64 | 15.22 | 1,773 | 109.84 | 11.31 | 1,773 | 0.91 | 0.08 | 1,773 | 30.40 | 6.00 | STATA (v14) | PLINK/FAST | Affymetrix 6.0 | MACH v1.0.16 | PMID: 2646917 |
| Multi-Ethnic Study of Atherosclerosis | MESA | Population based | >=95% | Gender mismatch; PCA outliers | N/A | measured | 758 | 100.79 | 12.82 | 758 | 106.03 | 9.76 | 758 | 0.95 | 0.06 | 758 | 28.76 | 4.74 | 891 | 101.63 | 16.10 | 891 | 112.94 | 13.07 | 891 | 0.90 | 0.08 | 891 | 31.32 | 6.41 | SAS | SNPTEST2 | AFFY6.0 | IMPUTE2 | PMID: 12397006; Am J Epidemiol. 2002 Nov |
| Women's Health Initiative | WHI | Prospective cohort study | >=95% | first-degree relatives | N/A | mreasured | NA | NA | NA | NA | NA | NA | NA | NA | NA | NA | NA | NA | 8,022 | 91.56 | 13.56 | 8,022 | 111.26 | 13.05 | 8,022 | 0.82 | 0.07 | 8,022 | 31.03 | 6.53 | STATA (v14) | PLINK/Probable | AFFY6.0 | MACH v1.0.16 | PMID: 9492970 |
| **European American** | | | | | | | | | | | | | | | | | | | | | | | | | | | | | | | | | | | |
| Atherosclerosis Risk in Communities Study | ARIC | Population based | >=90% | first-degree relatives ; ancestry outliers ; gender mismatch; identity issues; excessive heterozygosity; missing height or weight | N/A | measured | 4,281 | 99.62 | 10.21 | 4,281 | 102.71 | 7.25 | 4,281 | 0.97 | 0.05 | 4,281 | 27.42 | 3.89 | 4,795 | 92.75 | 14.34 | 4,795 | 103.92 | 10.31 | 4,795 | 0.89 | 0.08 | 4,795 | 26.47 | 5.27 | STATA (v14) | PLINK/FAST | Affymetrix 6.0 | MACH v1.0.16 | PMID: 2646917 |

**Supplementary Table 2**. Association and replication results for all loci that reached suggestive significance (*P* <1x10$^{-6}$) in the discovery phase for WHRadjBMI. Abbreviations: CHR- chromosome number; POS- position (build GRCh38); EAF- effect allele frequency; SE- standard error.

| | | | | | | | | | | | | WITHOUT SOL | | | | | | | | | | | | | | | | | | | | | | | |
|---|---|---|---|---|---|---|---|---|---|---|---|---|---|---|---|---|---|---|---|---|---|---|---|---|---|---|---|---|---|---|---|---|---|---|---|
| | | | | Known Locus | | | SOL ONLY | | | | | EUR¥ | | | | | | AAǂ | | | | | | HL‡ | | | | | | ALLMETA | | | | | |
| dnSNPID | Nearest Gene | CHR | POS | (PMID) | Effect Allele | Other Allele | EAF | Beta | SE | P | N | EAF | Beta | SE | P | ISQ | N | EAF | Beta | SE | P | ISQ | N | EAF | Beta | SE | P | ISQ | N | EAF | Beta | SE | P | ISQ | N |
| **MEN** | | | | | | | | | | | | | | | | | | | | | | | | | | | | | | | | | | | |
| rs12032174 | *RYR2* | 1 | 237692868 | - | T | C | 0.648 | 0.005 | 0.001 | 4.5E-07 | 5,200 | 0.627 | -0.002 | 0.001 | 1.2E-01 | - | 4,165 | 0.606 | -0.001 | 0.002 | 5.5E-01 | 0 | 1,813 | 0.617 | 0.001 | 0.001 | 4.6E-01 | 0 | 4,030 | 0.619 | -0.001 | 0.001 | 4.6E-01 | 0 | 10,008 |
| rs10475310 | *PLEKHG4B* | 5 | 23024 | - | C | G | 0.748 | -0.006 | 0.001 | **2.0E-08** | 5,200 | - | - | - | - | - | - | 0.754 | -4.E-04 | 0.003 | 8.9E-01 | 0 | 758 | 0.749 | 0.001 | 0.001 | 4.4E-01 | 32.3 | 4,030 | 0.750 | 0.001 | 0.001 | 5.0E-01 | 24.9 | 4,788 |
| rs16977373 | *RIT2* | 18 | 43189620 | - | A | C | 0.962 | -0.013 | 0.003 | 6.6E-07 | 5,200 | 0.997 | 0.007 | 0.010 | 4.8E-01 | - | 4,165 | 0.793 | 0.001 | 0.002 | 5.5E-01 | 5.5 | 1,812 | 0.959 | 1.E-04 | 0.004 | 9.8E-01 | 0 | 2,735 | 0.839 | 0.001 | 0.002 | 5.1E-01 | 0 | 8,712 |
| rs721424 | *CFAP61* | 20 | 20358666 | - | T | G | 0.484 | -0.005 | 0.001 | 8.2E-07 | 5,200 | 0.618 | -0.001 | 0.001 | 5.4E-01 | - | 4,165 | 0.256 | 1.E-04 | 0.002 | 9.8E-01 | 50.1 | 1,813 | 0.493 | 0.002 | 0.001 | 6.0E-02 | 29.8 | 4,030 | 0.517 | 0.001 | 0.001 | 3.7E-01 | 33.5 | 10,008 |
| rs148213302 | *STS* | 23 | 7416464 | - | T | C | 0.995 | 0.025 | 0.005 | 3.7E-07 | 5,189 | - | - | - | - | - | - | 0.987 | -0.012 | 0.007 | 1.2E-01 | 0 | 758 | - | - | - | - | - | - | 0.987 | -0.012 | 0.007 | 1.2E-01 | 0 | 758 |
| **WOMEN** | | | | | | | | | | | | | | | | | | | | | | | | | | | | | | | | | | | |
| rs77377042 | *MARCKSL1* | 1 | 32343334 | - | T | C | 0.024 | 0.018 | 0.004 | 8.0E-07 | 7,472 | - | - | - | - | - | - | 0.127 | 0.002 | 0.002 | 1.7E-01 | 72.5 | 10,684 | 0.021 | 0.006 | 0.004 | 1.8E-01 | 0 | 6,998 | 0.113 | 0.003 | 0.002 | 7.7E-02 | 35 | 17,682 |
| rs75120960 | *EPHA5* | 4 | 66367753 | - | A | G | 0.98 | -0.02 | 0.004 | 1.9E-07 | 7,472 | - | - | - | - | - | - | 0.882 | -0.002 | 0.002 | 2.9E-01 | 49 | 10,683 | 0.983 | -0.001 | 0.005 | 9.0E-01 | 69.6 | 5,616 | 0.891 | -0.002 | 0.002 | 2.9E-01 | 52.6 | 16,299 |
| rs16922424 | *FAM110B* | 8 | 57618850 | - | T | C | 0.982 | 0.021 | 0.004 | 4.9E-07 | 7471 | - | - | - | - | - | - | 0.892 | -0.002 | 0.002 | 2.2E-01 | 0 | 10,684 | 0.984 | 0.004 | 0.005 | 4.4E-01 | 26.6 | 5,616 | 0.900 | -0.002 | 0.002 | 3.4E-01 | 0 | 16,300 |
| rs79478137 | *SLC22A18AS* | 11 | 2891739 | rs116895106 (28552196)* | T | C | 0.015 | -0.023 | 0.004 | 2.0E-07 | 7,472 | 0.009 | 4.E-04 | 0.011 | 9.7E-01 | - | 4,678 | 0.084 | 0.001 | 0.002 | 7.7E-01 | 9.7 | 10,684 | 0.017 | -0.012 | 0.005 | 3.1E-02 | 0 | 6,582 | 0.074 | -0.001 | 0.002 | 6.2E-01 | 10.5 | 21,944 |
| rs113818604 | *NTM* | 11 | 131960980 | - | A | G | 0.013 | -0.027 | 0.005 | 5.5E-08 | 7,471 | 0.023 | 0.001 | 0.006 | 9.0E-01 | - | 4,678 | 0.004 | 0.003 | 0.009 | 7.6E-01 | 0 | 9,792 | 0.016 | -0.003 | 0.005 | 4.6E-01 | 0 | 6,786 | 0.017 | -0.001 | 0.003 | 7.3E-01 | 0 | 21,256 |
| rs115981023 | *TAOK3* | 12 | 118313300 | - | A | G | 0.009 | 0.029 | 0.006 | 8.9E-07 | 7,472 | 0.003 | -0.005 | 0.017 | 8.0E-01 | - | 4,678 | 0.049 | 0.002 | 0.002 | 3.9E-01 | 0 | 10,684 | 0.011 | 0.014 | 0.007 | 3.6E-02 | 71.5 | 5,616 | 0.044 | 0.003 | 0.002 | 1.4E-01 | 41.4 | 20,978 |
| rs146900844 | *ZNF207* | 17 | 32378136 | - | A | G | 0.005 | -0.041 | 0.008 | 6.3E-07 | 7,472 | 0.008 | 0.015 | 0.012 | 1.9E-01 | - | 4,678 | - | - | - | - | - | - | 0.007 | -0.001 | 0.013 | 9.6E-01 | 0 | 1,419 | 0.008 | 0.008 | 0.009 | 3.5E-01 | 0 | 6,097 |
| rs61305557 | *C19orf67* | 19 | 14081134 | - | A | G | 0.029 | -0.018 | 0.004 | 4.8E-07 | 7,472 | - | - | - | - | - | - | 0.121 | 0.008 | 0.005 | 1.5E-01 | 0 | 1,771 | 0.030 | 0.002 | 0.006 | 7.6E-01 | 37.7 | 2,612 | 0.083 | 0.005 | 0.004 | 1.9E-01 | 19.1 | 4,383 |
| **ALL** | | | | | | | | | | | | | | | | | | | | | | | | | | | | | | | | | | | |
| rs13301996 | *CDK5RAP2* | 9 | 120570806 | - | T | G | 0.808 | 0.005 | 0.001 | 5.7E-07 | 12,672 | 0.794 | 5.E-04 | 0.001 | 7.6E-01 | - | 8,845 | 0.872 | 0.004 | 0.001 | 1.1E-02 | 0 | 12,496 | 0.811 | 0.001 | 0.001 | 1.9E-01 | 0 | 12,341 | 0.823 | 0.002 | 0.001 | 1.7E-02 | 0 | 33,682 |
| rs115981023 | *TAOK3* | 12 | 118313300 | - | A | G | 0.008 | 0.023 | 0.004 | 2.1E-07 | 12,672 | 0.003 | 0.002 | 0.012 | 8.7E-01 | - | 8,845 | 0.049 | 0.001 | 0.002 | 6.3E-01 | 0 | 12,496 | 0.009 | 0.010 | 0.005 | 5.8E-02 | 64.5 | 8,456 | 0.043 | 0.002 | 0.002 | 2.5E-01 | 38.4 | 29,797 |
| rs185566196 | *KIAA0391* | 14 | 35152593 | rs12435790 (30239722) | T | C | 0.011 | 0.019 | 0.004 | 2.6E-07 | 12,672 | 0.003 | 0.003 | 0.017 | 8.8E-01 | - | 8,844 | 0.090 | 0.003 | 0.002 | 1.2E-01 | 0 | 12,496 | 0.017 | -0.004 | 0.004 | 3.3E-01 | 44.3 | 9,142 | 0.078 | 0.002 | 0.002 | 2.9E-01 | 22.3 | 30,482 |
| rs116612483 | *CDH4* | 20 | 60751325 | - | A | G | 0.006 | 0.027 | 0.005 | 6.0E-07 | 12,672 | - | - | - | - | - | - | 0.029 | -0.007 | 0.003 | 1.5E-02 | 0 | 12,496 | 0.005 | -0.007 | 0.007 | 3.2E-01 | 63.6 | 7,680 | 0.025 | -0.007 | 0.003 | 8.9E-03 | 17.4 | 20,176 |

| | | | | | | | | | | | | WITH SOL | | | | | | | | | | | | | | | | | | | | | | | |
|---|---|---|---|---|---|---|---|---|---|---|---|---|---|---|---|---|---|---|---|---|---|---|---|---|---|---|---|---|---|---|---|---|---|---|---|
| | | | | Known Locus | | | SOL ONLY | | | | | EUR¥ | | | | | | AAǂ | | | | | | HL‡ | | | | | | ALLMETA | | | | | |
| dnSNPID | Nearest Gene | CHR | POS | (PMID) | Effect Allele | Other Allele | EAF | Beta | SE | P | N | EAF | Beta | SE | P | ISQ | N | EAF | Beta | SE | P | ISQ | N | EAF | Beta | SE | P | ISQ | N | EAF | Beta | SE | P | ISQ | N |
| **MEN** | | | | | | | | | | | | | | | | | | | | | | | | | | | | | | | | | | | |
| rs12032174 | *RYR2* | 1 | 237692868 | - | T | C | 0.648 | 0.005 | 0.001 | 4.5E-07 | 5,200 | 0.639 | 0.002 | 0.001 | 5.1E-03 | 94.9 | 9,365 | 0.638 | 0.004 | 0.001 | 4.6E-05 | 78.7 | 7,013 | 0.634 | 0.003 | 0.001 | 2.3E-05 | 33.6 | 9,230 | 0.629 | 0.001 | 0.001 | 1.7E-02 | 59 | 15,208 |
| rs10475310 | *PLEKHG4B* | 5 | 23024 | - | C | G | 0.748 | -0.006 | 0.001 | **2.0E-08** | 5,200 | - | - | - | - | - | - | 0.749 | -0.006 | 0.001 | **8.5E-09** | 65 | 5,958 | 0.748 | -0.003 | 0.001 | 2.5E-05 | 70.6 | 9,230 | 0.749 | -0.003 | 0.001 | 3.8E-05 | 68.2 | 9,988 |
| rs16977373 | *RIT2* | 18 | 43189620 | - | A | C | 0.962 | -0.013 | 0.003 | 6.6E-07 | 5,200 | 0.965 | -0.011 | 0.003 | 8.1E-05 | 73.3 | 9,365 | 0.847 | -0.003 | 0.002 | 5.0E-02 | 87.8 | 7,012 | 0.961 | -0.008 | 0.002 | 9.1E-04 | 60.9 | 7,935 | 0.870 | -0.003 | 0.002 | 1.0E-01 | 65.8 | 13,912 |
| rs721424 | *CFAP61* | 20 | 20358666 | - | T | G | 0.484 | -0.005 | 0.001 | 8.2E-07 | 5,200 | 0.541 | -0.003 | 0.001 | 2.8E-05 | 87.3 | 9,365 | 0.443 | -0.004 | 0.001 | 6.2E-06 | 69.6 | 7,013 | 0.488 | -0.002 | 0.001 | 1.5E-02 | 73.6 | 9,230 | 0.505 | -0.001 | 0.001 | 2.3E-02 | 67.7 | 15,208 |
| rs148213302 | *STS* | 23 | 7416464 | - | T | C | 0.995 | 0.025 | 0.005 | 3.7E-07 | 5,189 | - | - | - | - | - | - | 0.993 | 0.014 | 0.004 | 1.1E-03 | 94 | 5,947 | - | - | - | - | - | - | 0.993 | 0.014 | 0.004 | 1.1E-03 | 94 | 5,947 |
| **WOMEN** | | | | | | | | | | | | | | | | | | | | | | | | | | | | | | | | | | | |
| rs77377042 | *MARCKSL1* | 1 | 32343334 | - | T | C | 0.024 | 0.018 | 0.004 | 8.0E-07 | 7,472 | - | - | - | - | - | - | 0.112 | 0.005 | 0.002 | 3.0E-03 | 85.4 | 18,156 | 0.023 | 0.012 | 0.003 | 2.9E-05 | 33.6 | 14,470 | 0.101 | 0.005 | 0.001 | 1.1E-03 | 64.1 | 25,154 |
| rs75120960 | *EPHA5* | 4 | 66367753 | - | A | G | 0.98 | -0.02 | 0.004 | 1.9E-07 | 7,472 | - | - | - | - | - | - | 0.895 | -0.004 | 0.002 | 4.9E-03 | 86.4 | 18,155 | 0.981 | -0.012 | 0.003 | 7.4E-05 | 81.1 | 13,088 | 0.902 | -0.004 | 0.001 | 6.2E-03 | 79.4 | 23,771 |
| rs16922424 | *FAM110B* | 8 | 57618850 | - | T | C | 0.982 | 0.021 | 0.004 | 4.9E-07 | 7471 | - | - | - | - | - | - | 0.905 | 0.001 | 0.002 | 4.0E-01 | 89.7 | 18,155 | 0.983 | 0.015 | 0.003 | 3.4E-06 | 67.8 | 13,087 | 0.911 | 0.002 | 0.002 | 3.1E-01 | 81.3 | 23,771 |
| rs79478137 | *SLC22A18AS* | 11 | 2891739 | rs116895106 (28552196)* | T | C | 0.015 | -0.023 | 0.004 | 2.0E-07 | 7,472 | 0.014 | -0.020 | 0.004 | 6.7E-08 | 74.5 | 12,150 | 0.070 | -0.005 | 0.002 | 1.6E-02 | 89.9 | 18,156 | 0.016 | -0.019 | 0.003 | **3.6E-09** | 2.8 | 14,054 | 0.063 | -0.005 | 0.002 | 3.4E-03 | 73.3 | 29,416 |
| rs113818604 | *NTM* | 11 | 131960980 | - | A | G | 0.013 | -0.027 | 0.005 | 5.5E-08 | 7,471 | 0.018 | -0.015 | 0.004 | 7.6E-05 | 92.6 | 12,149 | 0.011 | -0.020 | 0.004 | 5.1E-06 | 76.7 | 17,263 | 0.015 | -0.014 | 0.003 | 2.7E-05 | 63.8 | 14,257 | 0.016 | -0.009 | 0.003 | 1.1E-03 | 62.1 | 28,727 |
| rs115981023 | *TAOK3* | 12 | 118313300 | - | A | G | 0.009 | 0.029 | 0.006 | 8.9E-07 | 7,472 | 0.008 | 0.025 | 0.006 | 7.3E-06 | 69.9 | 12,150 | 0.044 | 0.006 | 0.002 | 1.0E-02 | 83 | 18,156 | 0.010 | 0.022 | 0.005 | 5.7E-07 | 69.4 | 13,088 | 0.040 | 0.006 | 0.002 | 2.4E-03 | 73.6 | 28,450 |
| rs146900844 | *ZNF207* | 17 | 32378136 | - | A | G | 0.005 | -0.041 | 0.008 | 6.3E-07 | 7,472 | 0.006 | -0.023 | 0.007 | 4.3E-04 | 93.6 | 12,150 | - | - | - | - | - | - | 0.006 | -0.030 | 0.007 | 1.0E-05 | 85.2 | 8,891 | 0.006 | -0.019 | 0.006 | 1.5E-03 | 88.8 | 13,569 |
| rs61305557 | *C19orf67* | 19 | 14081134 | - | A | G | 0.029 | -0.018 | 0.004 | 4.8E-07 | 7,472 | - | - | - | - | - | - | 0.062 | -0.009 | 0.003 | 6.3E-03 | 93.3 | 9,243 | 0.029 | -0.012 | 0.003 | 2.7E-04 | 70.9 | 10,084 | 0.055 | -0.007 | 0.003 | 2.2E-02 | 80.4 | 11,855 |
| **ALL** | | | | | | | | | | | | | | | | | | | | | | | | | | | | | | | | | | | |
| rs13301996 | *CDK5RAP2* | 9 | 120570806 | - | T | G | 0.808 | 0.005 | 0.001 | 5.7E-07 | 12,672 | 0.804 | 0.004 | 0.001 | 1.6E-05 | 84.5 | 21,517 | 0.830 | 0.005 | 0.001 | **2.9E-08** | 0 | 25,168 | 0.809 | 0.003 | 0.001 | 6.1E-06 | 19.5 | 25,013 | 0.818 | 0.003 | 0.001 | 1.1E-06 | 13.1 | 46,354 |
| rs115981023 | *TAOK3* | 12 | 118313300 | - | A | G | 0.008 | 0.023 | 0.004 | 2.1E-07 | 12,672 | 0.008 | 0.021 | 0.004 | **3.6E-08** | 63.8 | 21,517 | 0.040 | 0.006 | 0.002 | 2.0E-03 | 87.6 | 25,168 | 0.009 | 0.018 | 0.003 | **1.0E-08** | 67.4 | 21,128 | 0.036 | 0.006 | 0.002 | 4.2E-04 | 76 | 42,469 |
| rs185566196 | *KIAA0391* | 14 | 35152593 | rs12435790 (30239722) | T | C | 0.011 | 0.019 | 0.004 | 2.6E-07 | 12,672 | 0.011 | 0.018 | 0.004 | 3.2E-06 | 0 | 21,516 | 0.077 | 0.005 | 0.002 | 9.1E-04 | 79.8 | 25,168 | 0.014 | 0.008 | 0.003 | 6.9E-03 | 78.6 | 21,814 | 0.069 | 0.004 | 0.002 | 6.2E-03 | 66 | 43,154 |
| rs116612483 | *CDH4* | 20 | 60751325 | - | A | G | 0.006 | 0.027 | 0.005 | 6.0E-07 | 12,672 | - | - | - | - | - | - | 0.023 | 0.002 | 0.003 | 4.7E-01 | 91.4 | 25,168 | 0.006 | 0.015 | 0.004 | 1.6E-04 | 85.9 | 20,352 | 0.021 | 0.001 | 0.002 | 7.5E-01 | 85.7 | 32,848 |

ǂAfrican American (AA) replication samples included : Atherosclerosis Risk in Communities(ARIC), Multi-Ethnic Study of Atherosclerosis(MESA), Women's Health Initiative(WHI)

‡Hispanic Latino (HL) replication samples included : Genetics of Latinos Diabetic Retinopathy(GOLDR), Hispanic Community Health Study / Study of Latinos(HCHS/SOL), Mexican–American Hypertension Study(HTN), Mexican-American Coronary Artery Disease(MACAD), Multi-Ethnic Study of Atherosclerosis(MESA), Mexico-City, 1982 Pelotas Birth Cohort(PELOTAS), Starr County Health Studies(STARR), Women's Health Initiative(WHI)

¥European American (EUR) replication samples included : Atherosclerosis Risk in Communities(ARIC)

* Not genome-wide significant in original publication

**Supplementary Table 3**. Association and replication results for all loci that reached suggestive significance ($P < 1x10^{-6}$) in the discovery phase for WCadjBMI. Abbreviations: CHR- chromosome number; POS- position (build GRCh38); EAF- effect allele frequency; SE- standard error.

| | | | | | | | SOL ONLY | | | | | WITHOUT SOL | | | | | | | | | | | | | | | | | | | | | | | |
|---|---|---|---|---|---|---|---|---|---|---|---|---|---|---|---|---|---|---|---|---|---|---|---|---|---|---|---|---|---|---|---|---|---|---|---|
| | | | | | | | | | | | | EUR¥ | | | | | | AA† | | | | | | HL‡ | | | | | | ALLMETA | | | | | |
| dnSNPID | Nearest Gene | CHR | POS | Known Locus (PMID) | Effect Allele | Other Allele | EAF | Beta | SE | P | N | EAF | Beta | SE | P | ISQ | N | EAF | Beta | SE | P | ISQ | N | EAF | Beta | SE | P | ISQ | N | EAF | Beta | SE | P | ISQ | N |
| MEN | | | | | | | | | | | | | | | | | | | | | | | | | | | | | | | | | | | |
| rs74346221 | *GABRD* | 1 | 2029024 | - | T | G | 0.019 | -2.023 | 0.410 | 8.1E-07 | 5,202 | 0.002 | 2.847 | 4.337 | 5.1E-01 | - | 4,165 | 0.083 | -2.883 | 1.361 | **3.4E-02** | 0 | 1,054 | 0.017 | 0.240 | 0.663 | 7.2E-01 | 0 | 1,344 | 0.029 | -0.360 | 0.596 | 5.5E-01 | 76.5 | 2,398 |
| rs72693785 | *ATP1A1* | 1 | 116334965 | - | T | C | 0.006 | -4.088 | 0.734 | **2.5E-08** | 5,203 | 0.008 | -0.806 | 1.660 | 6.3E-01 | - | 4,165 | - | - | - | - | - | - | - | - | - | - | - | - | 0.008 | -0.806 | 1.660 | 6.3E-01 | 0 | 4,165 |
| rs11583298 | *ESRRG* | 1 | 217171261 | - | T | C | 0.822 | -0.711 | 0.140 | 4.0E-07 | 5,203 | 0.899 | -0.477 | 0.378 | 2.1E-01 | - | 4,166 | 0.977 | -1.496 | 0.820 | 6.8E-02 | 0 | 1,813 | 0.816 | 0.076 | 0.155 | 6.2E-01 | 0 | 4,206 | 0.833 | -0.048 | 0.142 | 7.3E-01 | 8.7 | 10,185 |
| rs76842062 | *MAP4K4* | 2 | 101847192 | - | T | G | 0.995 | 3.799 | 0.720 | 1.3E-07 | 5,203 | - | - | - | - | - | - | 0.946 | 0.452 | 0.531 | 3.9E-01 | 0 | 1,813 | - | - | - | - | - | - | 0.984 | 1.393 | 1.088 | 2.0E-01 | 0 | 1,812 |
| rs141365360 | *LOC102723448* | 3 | 21826 | - | A | G | 0.007 | 3.154 | 0.629 | 5.3E-07 | 5,203 | - | - | - | - | - | - | 0.012 | -1.676 | 1.672 | 3.2E-01 | 0 | 1,813 | 0.012 | 0.749 | 0.743 | 3.1E-01 | 0 | 1,344 | 0.035 | 0.382 | 0.488 | 4.3E-01 | 0 | 2,102 |
| rs6809759 | *PROK2* | 3 | 71937742 | rs7628338, rs6549455 (28448500); rs12330322 (25673412, 28443625, 28448500) | A | G | 0.497 | -0.586 | 0.102 | **9.4E-09** | 5,203 | 0.637 | 0.117 | 0.235 | 6.2E-01 | - | 4,166 | 0.394 | -0.014 | 0.239 | 9.6E-01 | 0 | 1,812 | 0.507 | 0.033 | 0.111 | 7.7E-01 | 0 | 4,206 | 0.510 | 0.039 | 0.092 | 6.8E-01 | 0 | 10,184 |
| rs76941364 | *COBL* | 7 | 52064843 | - | A | G | 0.989 | 2.443 | 0.498 | 9.4E-07 | 5,203 | 0.999 | 7.885 | 3.328 | **1.8E-02** | - | 4,165 | 0.984 | 1.393 | 1.088 | 2.0E-01 | 0 | 1,812 | 0.990 | 0.059 | 0.679 | 9.3E-01 | 0 | 2,064 | 0.963 | 0.421 | 0.415 | 3.1E-01 | 31.6 | 8,042 |
| rs139139519 | *SULF1* | 8 | 69625608 | - | A | G | 0.012 | -2.365 | 0.458 | 2.4E-07 | 5,203 | 0.012 | -0.941 | 1.113 | 4.0E-01 | - | 4,166 | 0.052 | 0.105 | 0.647 | 8.7E-01 | 0 | 758 | 0.016 | 0.132 | 0.550 | 8.1E-01 | 0 | 2,375 | 0.015 | -0.207 | 0.473 | 6.6E-01 | 0 | 8,354 |
| rs35569658 | *RBFOX1* | 16 | 5754433 | - | C | G | 0.313 | -0.562 | 0.112 | 5.3E-07 | 5,203 | 0.234 | -0.167 | 0.272 | 5.4E-01 | - | 4,166 | 0.148 | 0.025 | 0.344 | 9.4E-01 | 3.8 | 1,813 | 0.341 | -0.123 | 0.167 | 4.6E-01 | 32.2 | 2,862 | 0.288 | -0.111 | 0.132 | 4.0E-01 | 13.5 | 8,841 |
| rs148280037 | *SHISA9* | 16 | 12886370 | - | T | C | 0.984 | 2.081 | 0.412 | 4.5E-07 | 5,203 | 0.989 | -0.648 | 1.114 | 5.6E-01 | - | 4,166 | 0.984 | 0.809 | 1.117 | 4.7E-01 | 0 | 1,813 | 0.986 | 0.053 | 0.581 | 9.3E-01 | 52.6 | 2,338 | 0.986 | 0.062 | 0.468 | 8.9E-01 | 10.2 | 8,317 |
| rs143565319 | *PIK3C3* | 18 | 40962346 | - | T | C | 0.989 | 2.669 | 0.497 | 7.8E-08 | 5,203 | - | - | - | - | - | - | 0.932 | -0.614 | 0.475 | 2.0E-01 | 18.3 | 1,813 | 0.987 | 0.391 | 0.637 | 5.4E-01 | 0 | 2,064 | 0.951 | -0.255 | 0.381 | 5.0E-01 | 0 | 3,877 |
| WOMEN | | | | | | | | | | | | | | | | | | | | | | | | | | | | | | | | | | | |
| rs112469617 | *FZD7* | 2 | 201982887 | - | T | C | 0.009 | -2.977 | 0.594 | 5.5E-07 | 7,471 | - | - | - | - | - | - | 0.034 | 0.085 | 0.397 | 8.3E-01 | 69.8 | 10,683 | 0.005 | -1.154 | 1.295 | 3.7E-01 | 0 | 1,420 | 0.031 | -0.022 | 0.380 | 9.5E-01 | 59.7 | 12,103 |
| rs17385466 | *SOX5* | 12 | 23639004 | - | T | C | 0.923 | 1.089 | 0.209 | 1.8E-07 | 7,471 | 0.883 | -0.945 | 0.466 | **4.3E-02** | - | 4,678 | 0.974 | 0.507 | 0.366 | 1.7E-01 | 0 | 10,684 | 0.901 | 0.087 | 0.170 | 6.1E-01 | 57.7 | 8,613 | 0.911 | 0.053 | 0.146 | 7.2E-01 | 53.1 | 23,975 |
| ALL | | | | | | | | | | | | | | | | | | | | | | | | | | | | | | | | | | | |
| rs77993329 | *ACTRT2* | 1 | 3001535 | - | A | G | 0.020 | -1.457 | 0.288 | 4.1E-07 | 12,673 | 0.005 | -0.540 | 1.349 | 6.9E-01 | - | 8,845 | 0.095 | -0.144 | 0.191 | 4.5E-01 | 66.9 | 12,497 | 0.013 | -0.140 | 0.093 | 1.3E-01 | 66.2 | 9,803 | 0.029 | -0.142 | 0.084 | 8.9E-02 | 57 | 31,145 |
| rs138819965 | *LINC00299* | 2 | 8304328 | - | A | G | 0.994 | 2.706 | 0.525 | 2.5E-07 | 12,674 | - | - | - | - | - | - | 0.964 | -0.095 | 0.274 | 7.3E-01 | 0 | 12,497 | 0.995 | -0.042 | 0.632 | 9.5E-01 | 0 | 7,681 | 0.969 | -0.087 | 0.251 | 7.3E-01 | 0 | 20,178 |
| rs77319470 | *ADAMTS3* | 4 | 72638360 | rs7697556 (28443625); rs10518107 (26785701); rs16848284 (28448500) | T | G | 0.371 | 0.406 | 0.082 | 8.6E-07 | 12,674 | 0.417 | 0.146 | 0.204 | 4.7E-01 | - | 8,844 | 0.504 | -0.198 | 0.110 | 7.3E-02 | 45.3 | 12,496 | 0.344 | 0.012 | 0.017 | 4.8E-01 | 56.5 | 12,819 | 0.348 | 0.008 | 0.016 | 6.2E-01 | 54.1 | 34,159 |
| rs921999 | *FGF5* | 4 | 80296998 | - | A | C | 0.965 | -1.104 | 0.222 | 6.7E-07 | 12,674 | 0.999 | 1.464 | 3.837 | 7.0E-01 | - | 8,844 | 0.829 | 0.054 | 0.142 | 7.0E-01 | 80.5 | 12,496 | 0.989 | 0.056 | 0.084 | 5.0E-01 | 0 | 12,819 | 0.947 | 0.056 | 0.072 | 4.3E-01 | 6.5 | 34,159 |
| rs112404395 | *AP3B1* | 5 | 77964563 | - | C | G | 0.991 | -2.220 | 0.412 | 7.1E-08 | 12,674 | 0.985 | 0.412 | 0.991 | 6.8E-01 | - | 8,844 | 0.997 | -3.848 | 4.427 | 3.8E-01 | 0 | 2,825 | 0.992 | 0.025 | 0.142 | 8.6E-01 | 0 | 4,390 | 0.992 | 0.029 | 0.140 | 8.4E-01 | 0 | 16,059 |
| rs77264633 | *CCDC171* | 9 | 15823920 | - | A | G | 0.962 | -1.118 | 0.205 | **4.6E-08** | 12,674 | 0.928 | 0.161 | 0.385 | 6.8E-01 | - | 8,844 | 0.973 | 0.620 | 0.386 | 1.1E-01 | 0 | 12,497 | 0.975 | -0.006 | 0.049 | 9.0E-01 | 44.3 | 12,819 | 0.974 | 0.006 | 0.048 | 9.0E-01 | 35.9 | 34,160 |
| rs184067184 | *PHF21A* | 11 | 46189280 | - | A | C | 0.006 | -3.612 | 0.702 | 2.7E-07 | 12,674 | - | - | - | - | - | - | - | - | - | - | - | - | 0.010 | 0.282 | 1.287 | 8.3E-01 | 0 | 1,497 | 0.010 | 0.282 | 1.287 | 8.3E-01 | 0 | 1,497 |
| rs3168072 | *FADS2* | 11 | 61864038 | - | A | T | 0.725 | 0.514 | 0.102 | 5.3E-07 | 12,674 | 0.975 | 2.013 | 0.632 | **1.5E-03** | - | 8,845 | 0.977 | -0.233 | 0.363 | 5.2E-01 | 0 | 12,497 | 0.711 | 0.001 | 0.017 | 9.6E-01 | 0 | 12,819 | 0.711 | 0.002 | 0.017 | 9.2E-01 | 22.1 | 34,161 |
| rs60260780 | *WSB2* | 12 | 118041899 | - | T | C | 0.013 | 1.780 | 0.350 | 3.7E-07 | 12,674 | 0.025 | 0.676 | 0.779 | 3.9E-01 | - | 8,844 | 0.103 | -0.062 | 0.178 | 7.3E-01 | 0 | 12,496 | 0.021 | -0.037 | 0.065 | 5.7E-01 | 9.3 | 10,490 | 0.031 | -0.036 | 0.061 | 5.6E-01 | 0 | 31,830 |

| | | | | | | | SOL ONLY | | | | | WITH SOL | | | | | | | | | | | | | | | | | | | | | | | |
|---|---|---|---|---|---|---|---|---|---|---|---|---|---|---|---|---|---|---|---|---|---|---|---|---|---|---|---|---|---|---|---|---|---|---|---|
| | | | | | | | | | | | | EUR¥ | | | | | | AA† | | | | | | HL‡ | | | | | | ALLMETA | | | | | |
| dnSNPID | Nearest Gene | CHR | POS | Known Locus (PMID) | Effect Allele | Other Allele | EAF | Beta | SE | P | N | EAF | Beta | SE | P | ISQ | N | EAF | Beta | SE | P | ISQ | N | EAF | Beta | SE | P | ISQ | N | EAF | Beta | SE | P | ISQ | N |
| MEN | | | | | | | | | | | | | | | | | | | | | | | | | | | | | | | | | | | |
| rs74346221 | *GABRD* | 1 | 2029024 | - | T | G | 0.019 | -2.023 | 0.410 | 8.14E-07 | 5,202 | 0.024 | -2.095 | 0.393 | 9.5E-08 | 0 | 6,256 | 0.024 | -2.095 | 0.393 | 9.5E-08 | 0 | 6,256 | 0.018 | -1.398 | 0.349 | 6.2E-05 | 88.1 | 6,546 | 0.022 | -1.489 | 0.338 | 1.1E-05 | 79 | 7,600 |
| rs72693785 | *ATP1A1* | 1 | 116334965 | - | T | C | 0.006 | -4.088 | 0.734 | **2.5E-08** | 5,203 | 0.006 | -3.551 | 0.671 | 1.2E-07 | 69.4 | 9,368 | - | - | - | - | - | - | - | - | - | - | - | - | 0.006 | -3.551 | 0.671 | 1.2E-07 | 69.4 | 9,368 |
| rs11583298 | *ESRRG* | 1 | 217171261 | - | T | C | 0.822 | -0.711 | 0.140 | 4.0E-07 | 5,203 | 0.831 | -0.683 | 0.131 | 2.0E-07 | 0 | 9,369 | 0.826 | -0.733 | 0.138 | 1.1E-07 | 0 | 7,016 | 0.819 | -0.358 | 0.104 | 5.7E-04 | 57.4 | 9,409 | 0.827 | -0.383 | 0.100 | 1.2E-04 | 48.1 | 15,388 |
| rs76842062 | *MAP4K4* | 2 | 101847192 | - | T | G | 0.995 | 3.799 | 0.720 | 1.3E-07 | 5,203 | - | - | - | - | - | - | 0.969 | 1.511 | 0.363 | 3.2E-05 | 73.9 | 7,016 | - | - | - | - | - | - | 0.992 | 3.066 | 0.601 | 3.3E-07 | 44 | 7,015 |
| rs141365360 | *LOC102723448* | 3 | 21826 | - | A | G | 0.007 | 3.154 | 0.629 | 5.3E-07 | 5,203 | - | - | - | - | - | - | 0.012 | -2.317 | 0.442 | 1.6E-07 | 0 | 7,016 | 0.009 | 2.151 | 0.480 | 7.5E-06 | 83.6 | 6,547 | 0.024 | 1.423 | 0.386 | 2.2E-04 | 84.1 | 7,305 |
| rs6809759 | *PROK2* | 3 | 71937742 | rs7628338, rs6549455 (28448500); rs12330322 (25673412, 28443625, 28448500) | A | G | 0.497 | -0.586 | 0.102 | **9.4E-09** | 5,203 | 0.519 | -0.474 | 0.094 | 4.0E-07 | 86.7 | 9,369 | 0.481 | -0.498 | 0.094 | 1.1E-07 | 59.2 | 7,015 | 0.502 | -0.302 | 0.075 | 5.7E-05 | 54.4 | 9,409 | 0.504 | -0.243 | 0.069 | 3.9E-04 | 49.3 | 15,387 |
| rs76941364 | *COBL* | 7 | 52064843 | - | A | G | 0.989 | 2.443 | 0.498 | 9.4E-07 | 5,203 | 0.989 | 2.562 | 0.493 | 2.0E-07 | 61.8 | 9,368 | 0.992 | 3.066 | 0.601 | 3.3E-07 | 44 | 7,015 | 0.989 | 1.609 | 0.402 | 6.1E-05 | 76.1 | 7,267 | 0.974 | 1.249 | 0.319 | 8.9E-05 | 67.9 | 13,245 |
| rs139139519 | *SULF1* | 8 | 69625608 | - | A | G | 0.012 | -2.365 | 0.458 | 2.4E-07 | 5,203 | 0.012 | -2.159 | 0.424 | 3.5E-07 | 28.6 | 9,369 | 0.029 | 1.671 | 0.451 | 2.1E-04 | 91.3 | 5,961 | 0.014 | -1.343 | 0.352 | 1.4E-04 | 77.2 | 7,578 | 0.013 | -1.321 | 0.329 | 6.0E-05 | 55 | 13,557 |
| rs35569658 | *RBFOX1* | 16 | 5754433 | - | C | G | 0.313 | -0.562 | 0.112 | 5.3E-07 | 5,203 | 0.302 | -0.505 | 0.104 | 1.1E-06 | 44.6 | 9,369 | 0.297 | -0.506 | 0.107 | 2.0E-06 | 45.5 | 7,016 | 0.322 | -0.426 | 0.093 | 4.7E-06 | 47 | 8,065 | 0.302 | -0.373 | 0.085 | 1.2E-05 | 40.1 | 14,044 |
| rs148280037 | *SHISA9* | 16 | 12886370 | - | T | C | 0.984 | 2.081 | 0.412 | 4.5E-07 | 5,203 | 0.985 | 1.753 | 0.386 | 5.7E-06 | 81.1 | 9,369 | 0.984 | 1.929 | 0.387 | 6.1E-07 | 0 | 7,016 | 0.985 | 1.402 | 0.336 | 3.0E-05 | 75.7 | 7,541 | 0.985 | 1.199 | 0.309 | 1.1E-04 | 62.6 | 13,520 |
| rs143565319 | *PIK3C3* | 18 | 40962346 | - | T | C | 0.989 | 2.669 | 0.497 | 7.8E-08 | 5,203 | - | - | - | - | - | - | 0.959 | 0.953 | 0.343 | 5.5E-03 | 91.7 | 7,016 | 0.988 | 1.807 | 0.392 | 4.0E-06 | 74.8 | 7,267 | 0.965 | 0.826 | 0.302 | 6.3E-03 | 83.8 | 9,080 |
| WOMEN | | | | | | | | | | | | | | | | | | | | | | | | | | | | | | | | | | | |
| rs112469617 | *FZD7* | 2 | 201982887 | - | T | C | 0.009 | -2.977 | 0.594 | 5.5E-07 | 7,471 | - | - | - | - | - | - | 0.026 | -0.862 | 0.330 | 9.0E-03 | 24.961 | 18,154 | 0.008 | -2.660 | 0.540 | 8.4E-07 | 38.9 | 8,891 | 0.025 | -0.880 | 0.320 | 6.0E-03 | 84 | 19,574 |
| rs17385466 | *SOX5* | 12 | 23639004 | - | T | C | 0.923 | 1.089 | 0.209 | 1.8E-07 | 7,471 | 0.916 | 0.749 | 0.191 | 8.6E-05 | 93.7 | 12,149 | 0.936 | 0.946 | 0.181 | 1.9E-07 | 2.216 | 18,155 | 0.910 | 0.485 | 0.132 | 2.3E-04 | 71.5 | 16,084 | 0.915 | 0.393 | 0.120 | 1.0E-03 | 68.4 | 31,446 |

| ALL | | | | | | | | | | | | | | | | | | | | | | | | | | | | | | | | | | | |
|---|---|---|---|---|---|---|---|---|---|---|---|---|---|---|---|---|---|---|---|---|---|---|---|---|---|---|---|---|---|---|---|---|---|---|---|
| rs77993329 | *ACTRT2* | 1 | 3001535 | - | A | G | 0.020 | -1.457 | 0.288 | 4.1E-07 | 12,673 | 0.019 | -1.417 | 0.282 | 4.9E-07 | 0 | 21,518 | 0.072 | -0.546 | 0.159 | 6.1E-04 | 85.3 | 25,170 | 0.013 | -0.265 | 0.089 | 2.8E-03 | 82.2 | 22,476 | 0.028 | -0.245 | 0.080 | 2.3E-03 | 75.1 | 43,818 |
| rs138819965 | *LINC00299* | 2 | 8304328 | - | A | G | 0.994 | 2.706 | 0.525 | 2.5E-07 | 12,674 | - | - | - | - | - | - | 0.970 | 0.504 | 0.243 | 3.8E-02 | 87 | 25,171 | 0.994 | 1.585 | 0.404 | 8.7E-05 | 74.3 | 20,355 | 0.974 | 0.434 | 0.227 | 5.6E-02 | 75.2 | 32,852 |
| rs77319470 | *ADAMTS3* | 4 | 72638360 | rs7697556 (28443625); rs10518107 (26785701); rs16848284 (28448500) | T | G | 0.371 | 0.406 | 0.082 | 8.6E-07 | 12,674 | 0.377 | 0.370 | 0.076 | 1.2E-06 | 28.9 | 21,518 | 0.418 | 0.191 | 0.066 | 3.7E-03 | 86.9 | 25,170 | 0.345 | 0.027 | 0.016 | 9.5E-02 | 76.7 | 25,493 | 0.349 | 0.023 | 0.016 | 1.5E-01 | 72.6 | 46,833 |
| rs921999 | *FGF5* | 4 | 80296998 | - | A | C | 0.965 | -1.104 | 0.222 | 6.7E-07 | 12,674 | 0.965 | -1.095 | 0.222 | 7.7E-07 | 0 | 21,518 | 0.868 | -0.280 | 0.119 | 1.9E-02 | 89.9 | 25,170 | 0.986 | -0.088 | 0.078 | 2.6E-01 | 63.6 | 25,493 | 0.949 | -0.054 | 0.069 | 4.3E-01 | 63.7 | 46,833 |
| rs112404395 | *AP3B1* | 5 | 77964563 | - | C | G | 0.991 | -2.220 | 0.412 | 7.1E-08 | 12,674 | 0.990 | -1.832 | 0.380 | 1.5E-06 | 83.4 | 21,518 | 0.991 | -2.234 | 0.410 | 5.2E-08 | 0 | 15,499 | 0.992 | -0.213 | 0.134 | 1.1E-01 | 89.4 | 17,064 | 0.992 | -0.205 | 0.133 | 1.2E-01 | 83 | 28,733 |
| rs77264633 | *CCDC171* | 9 | 15823920 | - | A | G | 0.962 | -1.118 | 0.205 | **4.6E-08** | 12,674 | 0.954 | -0.836 | 0.181 | 3.9E-06 | 88.4 | 21,518 | 0.964 | -0.736 | 0.181 | 4.8E-05 | 82.5 | 25,171 | 0.974 | -0.067 | 0.048 | 1.6E-01 | 77.3 | 25,493 | 0.973 | -0.053 | 0.047 | 2.6E-01 | 71.3 | 46,834 |
| rs184067184 | *PHF21A* | 11 | 46189280 | - | A | C | 0.006 | -3.612 | 0.702 | 2.7E-07 | 12,674 | - | - | - | - | - | - | - | - | - | - | - | - | 0.007 | -2.719 | 0.616 | 1.0E-05 | 85.8 | 14,171 | 0.007 | -2.719 | 0.616 | 1.0E-05 | 85.8 | 14,171 |
| rs3168072 | *FADS2* | 11 | 61864038 | - | A | T | 0.725 | 0.514 | 0.102 | 5.3E-07 | 12,674 | 0.731 | 0.552 | 0.101 | **4.2E-08** | 81.7 | 21,519 | 0.744 | 0.459 | 0.098 | 2.9E-06 | 33 | 25,171 | 0.711 | 0.014 | 0.016 | 3.9E-01 | 67 | 25,493 | 0.712 | 0.015 | 0.016 | 3.6E-01 | 66.1 | 46,835 |
| rs60260780 | *WSB2* | 12 | 118041899 | - | T | C | 0.013 | 1.780 | 0.350 | 3.7E-07 | 12,674 | 0.015 | 1.595 | 0.319 | 5.9E-07 | 40.1 | 21,518 | 0.085 | 0.318 | 0.159 | 4.5E-02 | 87.4 | 25,170 | 0.021 | 0.023 | 0.064 | 7.2E-01 | 78.6 | 23,164 | 0.030 | 0.018 | 0.060 | 7.7E-01 | 68.9 | 44,504 |

ǂAfrican American (AA) replication samples included : Atherosclerosis Risk in Communities(ARIC), Multi-Ethnic Study of Atherosclerosis(MESA), Women's Health Initiative(WHI)

‡Hispanic Latino (HL) replication samples included : Genetics of Latinos Diabetic Retinopathy(GOLDR), Hispanic Community Health Study / Study of Latinos(HCHS/SOL), Mexican–American Hypertension Study(HTN), Mexican-American Coronary Artery Disease(MACAD), Multi-Ethnic Study of Atherosclerosis(MESA), Mexico-City, 1982 Pelotas Birth Cohort(PELOTAS), Starr County Health Studies(STARR), Women's Health Initiative(WHI)

¥European American (EUR) replication samples included : Atherosclerosis Risk in Communities(ARIC)

**Supplementary Table 4**. Association and replication results for all loci that reached suggestive significance ( $P<1\times10^{-6}$) in the discovery phase for HIPadjBMI. Abbreviations: CHR- chromosome number; POS- position (build GRCh38); EAF- effect allele frequency; SE- standard error.

| | | | | | | | SOL ONLY (Log10) | | | | | WITHOUT SOL | | | | | | | | | | | | | | | | | | | |
|---|---|---|---|---|---|---|---|---|---|---|---|---|---|---|---|---|---|---|---|---|---|---|---|---|---|---|---|---|---|---|---|
| | | | | Known Locus | | | | | | | | EUR¥ | | | | | AAǂ | | | | | HL‡ | | | | | ALLMETA | | | | |
| dbSNPID | Nearest Gene | CHR | POS | (PMID) | Effect Allele | Other Allele | EAF | Beta | SE | P | N | EAF | Beta | SE | P | N | EAF | Z | P | ISQ | N | EAF | Z | P | ISQ | N | EAF | Z | P | ISQ | N |
| **MEN** | | | | | | | | | | | | | | | | | | | | | | | | | | | | | | | |
| rs114865909 | *NUF2* | 1 | 163589134 | - | T | C | 0.994 | -0.012 | 0.002 | 8.6E-07 | 5,197 | - | - | - | - | - | 0.953 | -0.974 | 3.3E-01 | 78 | 1,812 | 0.993 | -0.579 | 5.6E-01 | 0 | 1,344 | 0.970 | -1.116 | 2.7E-01 | 56.5 | 3,156 |
| rs149681500 | *ANO10* | 3 | 43396572 | - | T | C | 0.007 | -0.012 | 0.002 | 5.3E-08 | 5,197 | - | - | - | - | - | 0.036 | -0.273 | 7.9E-01 | 0 | 1,812 | 0.006 | -1.529 | 1.3E-01 | 0 | 1,344 | 0.023 | -1.204 | 2.3E-01 | 0 | 3,156 |
| rs3915213 | *LOC101927346* | 3 | 74024720 | - | T | C | 0.779 | -0.002 | 0.000 | 5.6E-07 | 5,197 | 0.704 | 0.028 | 0.175 | 8.7E-01 | 4,165 | 0.683 | 0.128 | 9.0E-01 | 24.2 | 1,812 | 0.772 | 0.464 | 6.4E-01 | 14.3 | 4,027 | 0.728 | 0.453 | 6.5E-01 | 0 | 10,004 |
| rs12677587 | *LOC101929066* | 8 | 18118812 | - | C | G | 0.012 | -0.009 | 0.002 | 9.6E-07 | 5,197 | 0.002 | 0.009 | 0.964 | 9.9E-01 | 4,280 | 0.068 | -0.053 | 9.6E-01 | 23.4 | 1,814 | 0.015 | 0.082 | 9.3E-01 | 0 | 2,064 | 0.020 | 0.023 | 9.8E-01 | 0 | 8,158 |
| rs56405004 | *MINPP1* | 10 | 87512407 | - | T | C | 0.018 | 0.008 | 0.001 | 1.7E-07 | 5,197 | - | - | - | - | - | 0.092 | 0.036 | 9.7E-01 | 0 | 1,812 | 0.012 | -2.292 | 2.2E-02 | 67.1 | 2,064 | 0.050 | -1.648 | 9.9E-02 | 53.9 | 3,876 |
| rs968849 | *LOC102724589* | 10 | 114997972 | - | A | G | 0.195 | -0.002 | 0.000 | 8.2E-07 | 5,197 | 0.345 | 0.081 | 0.166 | 6.3E-01 | 4,166 | 0.195 | -0.138 | 8.9E-01 | 0 | 1,813 | 0.205 | 0.627 | 5.3E-01 | 11 | 4,027 | 0.261 | 0.654 | 5.1E-01 | 0 | 10,006 |
| rs76469489 | *SLC7A10* | 19 | 33249049 | - | C | G | 0.013 | -0.009 | 0.002 | 9.6E-07 | 5,197 | 0.036 | 0.052 | 0.488 | 9.2E-01 | 4,166 | 0.008 | -0.482 | 6.3E-01 | 0 | 1,054 | 0.018 | 0.068 | 9.5E-01 | 0 | 2,873 | 0.026 | -0.057 | 9.5E-01 | 0 | 8,093 |
| rs7063750 | *VCX* | 23 | 7724974 | - | T | G | 0.039 | 0.004 | 0.001 | 2.7E-07 | 5,186 | - | - | - | - | - | 0.160 | -1.128 | 2.6E-01 | 0 | 758 | 0.027 | -1.770 | 7.7E-02 | 0 | 720 | 0.095 | -2.043 | 4.1E-02 | 0 | 1,478 |
| rs112519383 | *MID1* | 23 | 10446834 | - | A | G | 0.988 | -0.007 | 0.001 | 7.4E-08 | 5,186 | - | - | - | - | - | 0.914 | -0.135 | 8.9E-01 | 0 | 758 | 0.987 | 0.546 | 5.9E-01 | 0 | 720 | 0.950 | 0.284 | 7.8E-01 | 0 | 1,478 |
| **WOMEN** | | | | | | | | | | | | | | | | | | | | | | | | | | | | | | | |
| rs72978809 | *LPPR4* | 1 | 99260939 | - | A | G | 0.994 | 0.011 | 0.002 | 4.4E-07 | 7,462 | - | - | - | - | - | 0.959 | -1.051 | 2.9E-01 | 18.6 | 10,683 | 0.996 | 0.323 | 7.5E-01 | 70.4 | 4,839 | 0.971 | -0.691 | 4.9E-01 | 39 | 15,522 |
| rs12478843 | *HEATR5B* | 2 | 37080089 | - | A | G | 0.248 | -0.002 | 0.000 | 8.2E-08 | 7,462 | 0.150 | -0.288 | 0.303 | 3.4E-01 | 4,679 | 0.056 | 0.898 | 3.7E-01 | 0 | 10,683 | 0.256 | 0.476 | 6.3E-01 | 2.9 | 8,315 | 0.145 | 0.464 | 6.4E-01 | 0 | 23,677 |
| rs115331260 | *LOC105376941* | 3 | 5864170 | - | A | G | 0.985 | 0.008 | 0.002 | 8.8E-07 | 7,462 | 0.942 | 0.006 | 0.517 | 9.9E-01 | 4,678 | 0.990 | -0.133 | 8.9E-01 | 0 | 10,683 | 0.979 | -0.525 | 6.0E-01 | 58.1 | 7,120 | 0.976 | -0.382 | 7.0E-01 | 28.3 | 22,481 |
| rs7662640 | *LOC105374566* | 4 | 31765535 | - | T | C | 0.012 | 0.008 | 0.002 | 6.3E-07 | 7,462 | 0.021 | 0.642 | 0.837 | 4.4E-01 | 4,678 | 0.009 | -0.673 | 5.0E-01 | 0 | 1,771 | 0.014 | -1.391 | 1.6E-01 | 42.3 | 7,873 | 0.016 | -0.829 | 4.1E-01 | 37.6 | 14,322 |
| rs6814739 | *LINC01094* | 4 | 78654647 | - | T | C | 0.569 | -0.002 | 0.000 | 7.2E-07 | 7,462 | 0.750 | -0.062 | 0.247 | 8.0E-01 | 4,679 | 0.258 | -0.776 | 4.4E-01 | 0 | 10,684 | 0.577 | -1.507 | 1.3E-01 | 27.3 | 8,315 | 0.467 | -1.526 | 1.3E-01 | 6.9 | 23,678 |
| rs11099588 | *COQ2* | 4 | 83280326 | - | T | C | 0.057 | 0.004 | 0.001 | 2.4E-07 | 7,462 | 0.032 | 0.560 | 0.644 | 3.8E-01 | 4,678 | 0.232 | 0.768 | 4.4E-01 | 0 | 10,683 | 0.162 | -0.621 | 5.4E-01 | 58.8 | 8,231 | 0.168 | 0.537 | 5.9E-01 | 44.8 | 23,592 |
| rs6860625 | *NREP* | 5 | 111667124 | - | A | G | 0.813 | -0.002 | 0.000 | 6.4E-07 | 7,462 | 0.848 | -0.172 | 0.303 | 5.7E-01 | 4,679 | 0.633 | -0.188 | 8.5E-01 | 59.5 | 10,683 | 0.831 | -1.523 | 1.3E-01 | 0 | 8,315 | 0.745 | -1.281 | 2.0E-01 | 0 | 23,677 |
| rs77186623 | *LOC105375745* | 8 | 125608952 | - | A | C | 0.022 | -0.006 | 0.001 | 1.7E-07 | 7,461 | - | - | - | - | - | 0.082 | -0.320 | 7.5E-01 | 38.4 | 10,683 | 0.013 | -0.972 | 3.3E-01 | 0 | 5,616 | 0.058 | -0.830 | 4.1E-01 | 0 | 16,299 |
| rs10818474 | *MEGF9* | 9 | 120727686 | rs7044106 (25673412) | T | C | 0.809 | -0.002 | 0.000 | 5.1E-07 | 7,462 | 0.753 | 0.014 | 0.100 | 8.9E-01 | 4,773 | 0.571 | -1.176 | 2.4E-01 | 0 | 10,682 | 0.742 | 0.479 | 6.3E-01 | 0 | 7,345 | 0.664 | -0.468 | 6.4E-01 | 0 | 22,800 |
| rs28692724 | *IRF2BPL* | 14 | 77027445 | - | T | C | 0.425 | 0.002 | 0.000 | 7.3E-07 | 7,462 | 0.303 | 0.789 | 0.305 | **9.6E-03** | 4,678 | 0.260 | -0.286 | 7.8E-01 | 68.9 | 10,683 | 0.391 | -0.143 | 8.9E-01 | 0 | 3,260 | 0.294 | 1.021 | 3.1E-01 | 39.9 | 18,621 |
| rs6092086 | *LOC105372676* | 20 | 55193568 | - | A | G | 0.922 | -0.003 | 0.001 | 3.4E-07 | 7,462 | 0.989 | 1.144 | 1.042 | 2.7E-01 | 4,679 | 0.653 | -0.031 | 9.8E-01 | 0 | 10,684 | 0.941 | 0.642 | 5.2E-01 | 21.2 | 8,231 | 0.820 | 0.848 | 4.0E-01 | 0.1 | 23,594 |
| rs9631175 | *TAF4* | 20 | 62072648 | - | A | C | 0.793 | 0.002 | 0.000 | 6.0E-08 | 7462 | 0.881 | -0.161 | 0.370 | 6.6E-01 | 4,679 | 0.755 | 1.676 | 9.4E-02 | 0 | 10,683 | 0.807 | 0.957 | 3.4E-01 | 26.8 | 8,315 | 0.798 | 1.499 | 1.3E-01 | 13.1 | 23,677 |
| **ALL** | | | | | | | | | | | | | | | | | | | | | | | | | | | | | | | |
| rs144655586 | *CLSPN* | 1 | 35725381 | - | T | C | 0.012 | 0.007 | 0.001 | 1.7E-07 | 12,658 | - | - | - | - | - | - | - | - | - | - | 0.009 | 0.738 | 4.6E-01 | 29.2 | 4,490 | 0.009 | 0.738 | 4.6E-01 | 29.2 | 4,490 |
| rs712900 | *LPPR4* | 1 | 99261313 | - | T | C | 0.014 | -0.006 | 0.001 | 3.4E-07 | 12,659 | 0.003 | 1.482 | 1.534 | 3.3E-01 | 8,845 | 0.096 | 2.159 | 3.1E-02 | 0 | 12,497 | 0.012 | -0.447 | 6.6E-01 | 0 | 7,680 | 0.045 | 1.720 | 8.5E-02 | 0 | 29,022 |
| rs115546449 | *TMEM63A* | 1 | 225882871 | - | A | G | 0.026 | -0.004 | 0.001 | 3.0E-07 | 12,659 | 0.004 | -0.040 | 1.365 | 9.8E-01 | 8,844 | 0.122 | -0.428 | 6.7E-01 | 0 | 12,497 | 0.021 | 0.225 | 8.2E-01 | 14.3 | 9,791 | 0.057 | -0.160 | 8.7E-01 | 0 | 31,132 |
| rs145815581 | *ANO10* | 3 | 43466242 | - | A | G | 0.007 | -0.008 | 0.002 | 1.8E-07 | 12,659 | - | - | - | - | - | 0.041 | 0.615 | 5.4E-01 | 32.4 | 12,497 | 0.005 | -0.057 | 9.5E-01 | 0 | 7,680 | 0.027 | 0.448 | 6.5E-01 | 0 | 20,177 |
| rs72886347 | *FHIT* | 3 | 60196461 | - | T | C | 0.993 | 0.008 | 0.002 | 7.2E-07 | 12,659 | - | - | - | - | - | 0.964 | -1.609 | 1.1E-01 | 74.9 | 12,497 | 0.994 | -1.119 | 2.6E-01 | 23.8 | 7,680 | 0.975 | -1.957 | 5.0E-02 | 52.9 | 20,177 |
| rs17136358 | *EIF2AK1* | 7 | 6040031 | - | T | C | 0.963 | -0.003 | 0.001 | 2.8E-07 | 12,659 | 0.939 | -0.369 | 0.286 | 2.0E-01 | 8,844 | 0.942 | 0.281 | 7.8E-01 | 0 | 12,497 | 0.966 | -0.118 | 9.1E-01 | 17.1 | 12,342 | 0.950 | -0.561 | 5.8E-01 | 1.1 | 33,683 |
| rs117683919 | *LOC105375440* | 7 | 106634111 | - | A | G | 0.006 | -0.009 | 0.002 | 1.4E-07 | 12,659 | 0.010 | -0.020 | 0.747 | 9.8E-01 | 8,844 | 0.004 | -0.852 | 4.0E-01 | 0 | 4,474 | 0.009 | 1.630 | 1.0E-01 | 23.7 | 10,871 | 0.008 | 0.710 | 4.8E-01 | 19.5 | 24,189 |
| rs143542634 | *PATL1* | 11 | 59666070 | - | A | G | 0.006 | 0.009 | 0.002 | 4.3E-07 | 12,659 | 0.009 | 0.999 | 0.863 | 2.5E-01 | 8,845 | 0.004 | 1.031 | 3.0E-01 | 0 | 4,474 | 0.011 | 0.300 | 7.6E-01 | 0 | 3,541 | 0.008 | 1.507 | 1.3E-01 | 0 | 16,860 |

| | | | | | | | SOL ONLY (Log10) | | | | | WITH SOL | | | | | | | | | | | | | | | | | | | |
|---|---|---|---|---|---|---|---|---|---|---|---|---|---|---|---|---|---|---|---|---|---|---|---|---|---|---|---|---|---|---|---|
| | | | | Known Locus | | | | | | | | EUR¥ | | | | | AAǂ | | | | | HL‡ | | | | | ALLMETA | | | | |
| dbSNPID | Nearest Gene | CHR | POS | (PMID) | Effect Allele | Other Allele | EAF | Beta | SE | P | N | EAF | Z | P | ISQ | N | EAF | Z | P | ISQ | N | EAF | Z | P | ISQ | N | EAF | Z | P | ISQ | N |
| **MEN** | | | | | | | | | | | | | | | | | | | | | | | | | | | | | | | |
| rs114865909 | *NUF2* | 1 | 163589134 | - | T | C | 0.994 | -0.012 | 0.002 | 8.6E-07 | 5,197 | - | - | - | - | - | 0.983 | -4.733 | 2.2E-06 | 72.7 | 7,009 | 0.994 | -4.650 | 3.3E-06 | 66 | 6,541 | 0.985 | -4.568 | 4.9E-06 | 67.4 | 8,353 |
| rs149681500 | *ANO10* | 3 | 43396572 | - | T | C | 0.007 | -0.012 | 0.002 | 5.3E-08 | 5,197 | - | - | - | - | - | 0.014 | -4.824 | 1.4E-06 | 68.9 | 7,009 | 0.007 | -5.543 | **3.0E-08** | 17.9 | 6,541 | 0.013 | -5.032 | 4.9E-07 | 55.4 | 8,353 |
| rs3915213 | *LOC101927346* | 3 | 74024720 | - | T | C | 0.779 | -0.002 | 0.000 | 5.6E-07 | 5,197 | 0.746 | -3.622 | 2.9E-04 | 91.6 | 9,362 | 0.754 | -4.246 | 2.2E-05 | 76.1 | 7,009 | 0.776 | -3.451 | 5.6E-04 | 60.4 | 9,224 | 0.745 | -2.560 | 1.0E-02 | 59.2 | 15,201 |
| rs12677587 | *LOC101929066* | 8 | 18118812 | - | C | G | 0.012 | -0.009 | 0.002 | 9.6E-07 | 5,197 | 0.007 | -3.623 | 2.9E-04 | 90.8 | 9,477 | 0.026 | -4.246 | 2.2E-05 | 72.6 | 7,011 | 0.013 | -4.102 | 4.1E-05 | 72.6 | 7,261 | 0.017 | -3.039 | 2.4E-03 | 69.1 | 13,355 |
| rs56405004 | *MINPP1* | 10 | 87512407 | - | T | C | 0.018 | 0.008 | 0.001 | 1.7E-07 | 5,197 | - | - | - | - | - | 0.037 | 4.526 | 6.0E-06 | 74.5 | 7,009 | 0.016 | 3.207 | 1.3E-03 | 92.1 | 7,261 | 0.032 | 2.885 | 3.9E-03 | 85.9 | 9,073 |
| rs968849 | *LOC102724589* | 10 | 114997972 | - | A | G | 0.195 | -0.002 | 0.000 | 8.2E-07 | 5,197 | 0.262 | -3.348 | 8.1E-04 | 92.5 | 9,363 | 0.195 | -4.316 | 1.6E-05 | 67.7 | 7,010 | 0.199 | -3.287 | 1.0E-03 | 60.7 | 9,224 | 0.239 | -2.353 | 1.9E-02 | 58.5 | 15,203 |
| rs76469489 | *SLC7A10* | 19 | 33249049 | - | C | G | 0.013 | -0.009 | 0.002 | 9.6E-07 | 5,197 | 0.023 | -3.579 | 3.4E-04 | 91.1 | 9,363 | 0.012 | -4.666 | 3.1E-06 | 59.5 | 6,251 | 0.015 | -3.892 | 1.0E-04 | 58.6 | 8,070 | 0.021 | -3.109 | 1.9E-03 | 61 | 13,290 |
| rs7063750 | *VCX* | 23 | 7724974 | - | T | G | 0.039 | 0.004 | 0.001 | 2.7E-07 | 5,186 | - | - | - | - | - | 0.054 | 4.404 | 1.1E-05 | 88 | 5,944 | 0.038 | 4.204 | 2.6E-05 | 91.6 | 5,906 | 0.051 | 3.577 | 3.5E-04 | 88.9 | 6,664 |
| rs112519383 | *MID1* | 23 | 10446834 | - | A | G | 0.988 | -0.007 | 0.001 | 7.4E-08 | 5,186 | - | - | - | - | - | 0.979 | -5.076 | 3.9E-07 | 69 | 5,944 | 0.988 | -4.853 | 1.2E-06 | 82.5 | 5,906 | 0.980 | -4.614 | 3.9E-06 | 75 | 6,664 |
| **WOMEN** | | | | | | | | | | | | | | | | | | | | | | | | | | | | | | | |
| rs72978809 | *LPPR4* | 1 | 99260939 | - | A | G | 0.994 | 0.011 | 0.002 | 4.4E-07 | 7,462 | - | - | - | - | - | 0.974 | 2.433 | 1.5E-02 | 87 | 18,145 | 0.995 | 4.137 | 3.5E-05 | 83.2 | 12,301 | 0.978 | 2.310 | 2.1E-02 | 81.6 | 22,984 |
| rs12478843 | *HEATR5B* | 2 | 37080089 | - | A | G | 0.248 | -0.002 | 0.000 | 8.2E-08 | 7,462 | 0.210 | -4.793 | 1.6E-06 | 85 | 12,141 | 0.135 | -2.750 | 6.0E-03 | 87.4 | 18,145 | 0.252 | -3.343 | 8.3E-04 | 63.1 | 15,777 | 0.169 | -2.221 | 2.6E-02 | 62.1 | 31,139 |
| rs115331260 | *LOC105376941* | 3 | 5864170 | - | A | G | 0.985 | 0.008 | 0.002 | 8.8E-07 | 7,462 | 0.968 | 3.862 | 1.1E-04 | 89.2 | 12,140 | 0.988 | 3.051 | 2.3E-03 | 80.5 | 18,145 | 0.982 | 3.150 | 1.6E-03 | 77.3 | 14,582 | 0.979 | 2.123 | 3.4E-02 | 69.1 | 29,943 |
| rs7662640 | *LOC105374566* | 4 | 31765535 | - | T | C | 0.012 | 0.008 | 0.002 | 6.3E-07 | 7,462 | 0.016 | 4.382 | 1.2E-05 | 83.9 | 12,140 | 0.011 | 4.184 | 2.9E-05 | 87.1 | 9,233 | 0.013 | 2.478 | 1.3E-02 | 75.6 | 15,335 | 0.014 | 2.243 | 2.5E-02 | 71.3 | 21,784 |
| rs6814739 | *LINC01094* | 4 | 78654647 | - | T | C | 0.569 | -0.002 | 0.000 | 7.2E-07 | 7,462 | 0.639 | -4.040 | 5.3E-05 | 87.9 | 12,141 | 0.386 | -3.773 | 1.6E-04 | 74.8 | 18,146 | 0.573 | -4.502 | 6.7E-06 | 47.2 | 15,777 | 0.492 | -3.756 | 1.7E-04 | 47.6 | 31,140 |
| rs11099588 | *COQ2* | 4 | 83280326 | - | T | C | 0.057 | 0.004 | 0.001 | 2.4E-07 | 7,462 | 0.047 | 4.589 | 4.5E-06 | 84.3 | 12,140 | 0.160 | 3.902 | 9.5E-05 | 76.8 | 18,145 | 0.112 | 3.112 | 1.9E-03 | 75.5 | 15,693 | 0.141 | 3.000 | 2.7E-03 | 67.3 | 31,054 |
| rs6860625 | *NREP* | 5 | 111667124 | - | A | G | 0.813 | -0.002 | 0.000 | 6.4E-07 | 7,462 | 0.827 | -4.255 | 2.1E-05 | 85.7 | 12,141 | 0.707 | -3.337 | 8.5E-04 | 83.9 | 18,145 | 0.823 | -4.530 | 5.9E-06 | 22.7 | 15,777 | 0.761 | -3.554 | 3.8E-04 | 46.4 | 31,139 |
| rs77186623 | *LOC105375745* | 8 | 125608952 | - | A | C | 0.022 | -0.006 | 0.001 | 1.7E-07 | 7,461 | - | - | - | - | - | 0.058 | -3.596 | 3.2E-04 | 83.1 | 18,144 | 0.018 | -4.584 | 4.6E-06 | 59.6 | 13,077 | 0.047 | -3.615 | 3.0E-04 | 67.9 | 23,760 |

| | | | | | | | | | | | | | | | | | | | | | | | | | | | | | | |
|---|---|---|---|---|---|---|---|---|---|---|---|---|---|---|---|---|---|---|---|---|---|---|---|---|---|---|---|---|---|---|
| rs10818474 | *MEGF9* | 9 | 120727686 | rs7044106 (25673412) | T | C | 0.809 | -0.002 | 0.000 | 5.1E-07 | 7,462 | 0.787 | -3.834 | 1.3E-04 | 90.5 | 12,235 | 0.669 | -4.123 | 3.7E-05 | 71.3 | 18,144 | 0.776 | -3.228 | 1.3E-03 | 62 | 14,807 | 0.700 | -2.900 | 3.7E-03 | 52.6 | 30,262 |
| rs28692724 | *IRF2BPL* | 14 | 77027445 | - | T | C | 0.425 | 0.002 | 0.000 | 7.3E-07 | 7,462 | 0.378 | 5.490 | **4.0E-08** | 8.3 | 12,140 | 0.328 | 2.956 | 3.1E-03 | 86.6 | 18,145 | 0.415 | 4.053 | 5.1E-05 | 45.8 | 10,722 | 0.331 | 3.512 | 4.5E-04 | 66.1 | 26,083 |
| rs6092086 | *LOC105372676* | 20 | 55193568 | - | A | G | 0.922 | -0.003 | 0.001 | 3.4E-07 | 7,462 | 0.948 | -3.318 | 9.1E-04 | 93.8 | 12,141 | 0.764 | -3.296 | 9.8E-04 | 81.4 | 18,146 | 0.932 | -3.053 | 2.3E-03 | 67 | 15,693 | 0.845 | -1.762 | 7.8E-02 | 63.5 | 31,056 |
| rs9631175 | *TAF4* | 20 | 62072648 | - | A | C | 0.793 | 0.002 | 0.000 | 6.0E-08 | 7462 | 0.827 | 3.978 | 7.0E-05 | 92.7 | 12,141 | 0.770 | 4.762 | 1.9E-06 | 71.4 | 18,145 | 0.800 | 4.422 | 9.8E-06 | 56.6 | 15,777 | 0.797 | 3.960 | 7.5E-05 | 54.7 | 31,139 |
| **ALL** | | | | | | | | | | | | | | | | | | | | | | | | | | | | | | | |
| rs144655586 | *CLSPN* | 1 | 35725381 | - | T | C | 0.012 | 0.007 | 0.001 | 1.7E-07 | 12,658 | - | - | - | - | - | - | - | - | - | - | 0.011 | 4.873 | 1.1E-06 | 57.1 | 17,148 | 0.011 | 4.873 | 1.1E-06 | 57.1 | 17,148 |
| rs712900 | *LPPR4* | 1 | 99261313 | - | T | C | 0.014 | -0.006 | 0.001 | 3.4E-07 | 12,659 | 0.009 | -3.293 | 9.9E-04 | 93.8 | 21,504 | 0.055 | -2.096 | 3.6E-02 | 88.9 | 25,156 | 0.013 | -4.298 | 1.7E-05 | 66 | 20,339 | 0.036 | -1.375 | 1.7E-01 | 78 | 41,681 |
| rs115546449 | *TMEM63A* | 1 | 225882871 | - | A | G | 0.026 | -0.004 | 0.001 | 3.0E-07 | 12,659 | 0.017 | -3.951 | 7.8E-05 | 90.6 | 21,503 | 0.074 | -3.937 | 8.3E-05 | 73.9 | 25,156 | 0.024 | -3.700 | 2.2E-04 | 67.5 | 22,450 | 0.048 | -2.891 | 3.8E-03 | 59.2 | 43,791 |
| rs145815581 | *ANO10* | 3 | 43466242 | - | A | G | 0.007 | -0.008 | 0.002 | 1.8E-07 | 12,659 | - | - | - | - | - | 0.024 | -3.273 | 1.1E-03 | 84.9 | 25,156 | 0.006 | -4.157 | 3.2E-05 | 71.1 | 20,339 | 0.020 | -2.892 | 3.8E-03 | 73.5 | 32,836 |
| rs72886347 | *FHIT* | 3 | 60196461 | - | T | C | 0.993 | 0.008 | 0.002 | 7.2E-07 | 12,659 | - | - | - | - | - | 0.979 | 2.381 | 1.7E-02 | 89.8 | 25,156 | 0.993 | 3.221 | 1.3E-03 | 83.4 | 20,339 | 0.982 | 1.542 | 1.2E-01 | 83.6 | 32,836 |
| rs17136358 | *EIF2AK1* | 7 | 6040031 | - | T | C | 0.963 | -0.003 | 0.001 | 2.8E-07 | 12,659 | 0.953 | -4.768 | 1.9E-06 | 81.2 | 21,503 | 0.953 | -3.446 | 5.7E-04 | 80.6 | 25,156 | 0.964 | -3.738 | 1.9E-04 | 57 | 25,001 | 0.954 | -3.163 | 1.6E-03 | 53.1 | 46,342 |
| rs117683919 | *LOC105375440* | 7 | 106634111 | - | A | G | 0.006 | -0.009 | 0.002 | 1.4E-07 | 12,659 | 0.008 | -4.053 | 5.1E-05 | 91.1 | 21,503 | 0.005 | -4.957 | 7.2E-07 | 53.1 | 17,133 | 0.007 | -2.750 | 6.0E-03 | 77.1 | 23,530 | 0.008 | -2.508 | 1.2E-02 | 69.8 | 36,848 |
| rs143542634 | *PATL1* | 11 | 59666070 | - | A | G | 0.006 | 0.009 | 0.002 | 4.3E-07 | 12,659 | 0.007 | 4.622 | 3.8E-06 | 82 | 21,504 | 0.005 | 4.873 | 1.1E-06 | 37.4 | 17,133 | 0.007 | 4.610 | 4.0E-06 | 54.7 | 16,200 | 0.007 | 4.450 | 8.6E-06 | 41.7 | 29,519 |

ƗAfrican American (AA) replication samples included : Atherosclerosis Risk in Communities(ARIC), Multi-Ethnic Study of Atherosclerosis(MESA), Women's Health Initiative(WHI)

‡Hispanic Latino (HL) replication samples included : Genetics of Latinos Diabetic Retinopathy(GOLDR), Hispanic Community Health Study / Study of Latinos(HCHS/SOL), Mexican–American Hypertension Study(HTN), Mexican-American Coronary Artery Disease(MACAD), Multi-Ethnic Study of Atherosclerosis(MESA), Mexico-City, 1982 Pelotas Birth Cohort(PELOTAS), Starr County Health Studies(STARR), Women's Health Initiative(WHI).

¥European American (EA) replication samples included : Atherosclerosis Risk in Communities(ARIC)

**Supplementary Table 5**. Summary of association results in SOL subgroup analyses for suggestively significant loci (P<1E-6) associated with WHRadjBMI. EAF-estimated allele frequency, CHR- chromosome, POS-position (build GRCh38), SE- standard error, ISQ- I squared heterogeneity. EAF for reference population obtained from 1000 Genomes Project Phase 3.

| | Nearest | | | Effect | Other | Combined | | | | | Caribbean | | | | | Mainland | | | | | | | EAF | | |
|---|---|---|---|---|---|---|---|---|---|---|---|---|---|---|---|---|---|---|---|---|---|---|---|---|---|
| **dbSNPID** | **Gene** | **CHR** | **POS** | **Allele** | **Allele** | **N** | **EAF** | **BETA** | **SE** | **P** | **N** | **EAF** | **BETA** | **SE** | **P** | **N** | **EAF** | **BETA** | **SE** | **P** | **ISQ** | $P_{diff}$ | **AFR** | **EUR** | **AMR** |
| **MEN** | | | | | | | | | | | | | | | | | | | | | | | | | |
| rs12032174 | *RYR2* | 1 | 237692868 | C | T | 5,200 | 0.352 | -0.005 | 0.001 | 4.5E-07 | 2,395 | 0.353 | -0.005 | 0.002 | 5.9E-04 | 2,793 | 0.352 | -0.005 | 0.001 | 4.5E-04 | 64.72 | 3.8E-01 | 0.401 | 0.383 | 0.370 |
| rs10475310 | *PLEKHG4B* | 5 | 23024 | C | G | 5,200 | 0.748 | -0.006 | 0.001 | **2.0E-08** | 2,395 | 0.747 | -0.006 | 0.002 | 1.8E-04 | 2,793 | 0.748 | -0.006 | 0.002 | 4.9E-05 | 0 | 3.9E-01 | 0.781 | 0.751 | 0.733 |
| rs16977373 | *RIT2* | 18 | 43189620 | A | C | 5,200 | 0.962 | -0.013 | 0.003 | 6.6E-07 | 2,395 | 0.939 | -0.015 | 0.003 | 9.7E-06 | 2,793 | 0.981 | -0.010 | 0.005 | 3.9E-02 | 83.56 | 2.7E-01 | 0.752 | 0.997 | 0.984 |
| rs721424 | *CFAP61* | 20 | 20358666 | G | T | 5,200 | 0.516 | 0.005 | 0.001 | 8.2E-07 | 2,395 | 0.520 | 0.005 | 0.002 | 2.4E-03 | 2,793 | 0.513 | 0.005 | 0.001 | 2.3E-04 | 0 | 4.0E-01 | 0.816 | 0.421 | 0.548 |
| rs148213302 | *STS* | X | 7416464 | T | C | 5,189 | 0.995 | 0.025 | 0.005 | 3.7E-07 | 2,391 | 0.992 | 0.030 | 0.006 | 6.5E-07 | 2,786 | 0.997 | 0.009 | 0.009 | 3.0E-01 | 0 | 5.5E-02 | 0.981 | 1.000 | 1.000 |
| **WOMEN** | | | | | | | | | | | | | | | | | | | | | | | | | |
| rs77377042 | *MARCKSL1* | 1 | 32343334 | C | T | 7,472 | 0.976 | -0.018 | 0.004 | 8.0E-07 | 3,238 | 0.961 | -0.021 | 0.005 | 3.7E-06 | 4,220 | 0.987 | -0.011 | 0.006 | 8.2E-02 | 81.79 | 1.7E-01 | 0.831 | 1.000 | 0.988 |
| rs75120960 | *EPHA5* | 4 | 66367753 | A | G | 7,472 | 0.980 | -0.020 | 0.004 | 1.9E-07 | 3,238 | 0.966 | -0.018 | 0.005 | 1.6E-04 | 4,220 | 0.991 | -0.027 | 0.008 | 3.9E-04 | 0 | 2.5E-01 | 0.860 | 1.000 | 0.987 |
| rs16922424 | *FAM110B* | 8 | 57618850 | T | C | 7,471 | 0.982 | 0.021 | 0.004 | 4.9E-07 | 3,237 | 0.969 | 0.017 | 0.005 | 1.1E-03 | 4,220 | 0.992 | 0.034 | 0.008 | 2.6E-05 | 24.99 | 8.8E-02 | 0.868 | 1.000 | 0.987 |
| rs79478137 | *SLC22A18AS* | 11 | 2891739 | C | T | 7,472 | 0.985 | 0.023 | 0.004 | 2.0E-07 | 3,238 | 0.975 | 0.025 | 0.006 | 8.9E-06 | 4,220 | 0.992 | 0.024 | 0.008 | 2.7E-03 | 0 | 4.0E-01 | 0.905 | 0.989 | 0.986 |
| rs113818604 | *NTM* | 11 | 131960980 | G | A | 7,471 | 0.987 | 0.027 | 0.005 | **5.5E-08** | 3,238 | 0.987 | 0.014 | 0.008 | 8.1E-02 | 4,219 | 0.986 | 0.034 | 0.006 | **1.6E-08** | 78.48 | 6.1E-02 | 0.998 | 0.977 | 0.991 |
| rs115981023 | *TAOK3* | 12 | 118313300 | G | A | 7,472 | 0.991 | -0.029 | 0.006 | 8.9E-07 | 3,238 | 0.984 | -0.030 | 0.007 | 2.7E-05 | 4,220 | 0.997 | -0.027 | 0.012 | 2.5E-02 | 0 | 3.9E-01 | 0.937 | 0.998 | 0.999 |
| rs146900844 | *ZNF207* | 17 | 32378136 | G | A | 7,472 | 0.995 | 0.041 | 0.008 | 6.3E-07 | 3,238 | 0.993 | 0.042 | 0.012 | 3.7E-04 | 4,220 | 0.996 | 0.040 | 0.012 | 6.6E-04 | 39.45 | 4.0E-01 | 1.000 | 0.994 | 0.996 |
| rs61305557 | *C19orf67* | 19 | 14081134 | G | A | 7,472 | 0.971 | 0.018 | 0.004 | 4.8E-07 | 3,238 | 0.946 | 0.021 | 0.004 | 1.2E-06 | 4,220 | 0.990 | 0.005 | 0.008 | 4.8E-01 | 72.14 | 8.6E-02 | 0.818 | 0.999 | 0.978 |
| **COMBINED** | | | | | | | | | | | | | | | | | | | | | | | | | |
| rs13301996 | *CDK5RAP2* | 9 | 120570806 | T | G | 12,672 | 0.808 | 0.005 | 0.001 | 5.7E-07 | 5,633 | 0.789 | 0.006 | 0.002 | 2.5E-04 | 7,013 | 0.822 | 0.003 | 0.001 | 2.0E-02 | 37.02 | 1.8E-01 | 0.886 | 0.809 | 0.794 |
| rs115981023 | *TAOK3* | 12 | 118313300 | G | A | 12,672 | 0.992 | -0.023 | 0.004 | 2.1E-07 | 5,633 | 0.985 | -0.019 | 0.006 | 8.9E-04 | 7,013 | 0.997 | -0.030 | 0.010 | 2.6E-03 | 0 | 2.6E-01 | 0.937 | 0.998 | 0.999 |
| rs185566196 | *KIAA0391* | 14 | 35152593 | C | T | 12,672 | 0.989 | -0.019 | 0.004 | 2.6E-07 | 5,633 | 0.979 | -0.025 | 0.005 | 5.3E-07 | 7,013 | 0.996 | -0.019 | 0.009 | 3.6E-02 | 0 | 3.5E-01 | 0.956 | 0.993 | 0.996 |
| rs116612483 | *CDH4* | 20 | 60751325 | G | A | 12,672 | 0.994 | -0.027 | 0.005 | 6.0E-07 | 5,633 | 0.989 | -0.025 | 0.007 | 2.6E-04 | 7,013 | 0.998 | -0.011 | 0.016 | 5.0E-01 | 0 | 2.9E-01 | 0.950 | 1.000 | 0.999 |

**Supplementary Table 6**. Summary of association results in SOL subgroup analyses for suggestively significant loci (P<1E-6) associated with WCadjBMI. EAF-estimated allele frequency, CHR- chromosome, POS- position (build GRCh38), SE- standard error, ISQ- I squared heterogeneity. EAF for reference population obtained from 1000 Genomes Project Phase 3.

| dbSNPID | Nearest Gene | CHR | POS | Effect Allele | Other Allele | Combined | | | | | Caribbean | | | | | Mainland | | | | | ISQ | $P_{diff}$ | EAF | | |
|---|---|---|---|---|---|---|---|---|---|---|---|---|---|---|---|---|---|---|---|---|---|---|---|---|---|
| | | | | | | N | EAF | BETA | SE | PVAL | N | EAF | BETA | SE | PVAL | N | EAF | BETA | SE | PVAL | | | AFR | EUR | AMR |
| **MEN** | | | | | | | | | | | | | | | | | | | | | | | | | |
| rs74346221 | *GABRD* | 1 | 2029024 | T | G | 5,202 | 0.019 | -2.023 | 0.41 | 8.1E-07 | 2,397 | 0.031 | -1.811 | 0.490 | 2.2E-04 | 2,793 | 0.009 | -2.565 | 0.779 | 9.9E-04 | 0 | 2.9E-01 | 0.121 | 0.000 | 0.009 |
| rs72693785 | *ATP1A1* | 1 | 116334965 | T | C | 5,203 | 0.006 | -4.088 | 0.734 | **2.5E-08** | 2,398 | 0.009 | -4.557 | 0.858 | 1.1E-07 | 2,793 | 0.003 | -3.336 | 1.516 | 2.8E-02 | 0 | 3.1E-01 | 0.002 | 0.009 | 0.001 |
| rs11583298 | *ESRRG* | 1 | 217171261 | T | C | 5,203 | 0.822 | -0.711 | 0.14 | 4.0E-07 | 2,398 | 0.910 | -0.644 | 0.270 | 1.7E-02 | 2,793 | 0.747 | -0.711 | 0.161 | 1.0E-05 | 0 | 3.9E-01 | 0.998 | 0.920 | 0.790 |
| rs76842062 | *MAP4K4* | 2 | 101847192 | T | G | 5,203 | 0.995 | 3.799 | 0.72 | 1.3E-07 | 2,398 | 0.990 | 3.052 | 0.805 | 1.5E-04 | 2,793 | 0.998 | 6.881 | 1.688 | 4.6E-05 | 0 | **4.9E-02** | 0.962 | 0.999 | 0.996 |
| rs141365360 | *LOC102723448* | 3 | 21826 | A | G | 5,203 | 0.007 | 3.154 | 0.629 | 5.3E-07 | 2,398 | 0.011 | 3.437 | 0.756 | 5.6E-06 | 2,793 | 0.004 | 2.159 | 1.153 | 6.1E-02 | 0 | 2.6E-01 | 0.056 | 0.000 | 0.004 |
| rs6809759 | *PROK2* | 3 | 71937742 | A | G | 5,203 | 0.497 | -0.586 | 0.102 | **9.4E-09** | 2,398 | 0.514 | -0.461 | 0.155 | 2.9E-03 | 2,793 | 0.482 | -0.673 | 0.135 | 6.5E-07 | 0 | 2.3E-01 | 0.358 | 0.601 | 0.504 |
| rs76941364 | *COBL* | 7 | 52064843 | A | G | 5,203 | 0.989 | 2.443 | 0.498 | 9.4E-07 | 2,398 | 0.983 | 1.753 | 0.597 | 3.3E-03 | 2,793 | 0.995 | 4.303 | 0.922 | 3.0E-06 | 0 | **2.7E-02** | 0.950 | 0.999 | 0.994 |
| rs139139519 | *SULF1* | 8 | 69625608 | A | G | 5,203 | 0.012 | -2.365 | 0.458 | 2.4E-07 | 2,398 | 0.018 | -2.668 | 0.573 | 3.2E-06 | 2,793 | 0.008 | -1.747 | 0.775 | 2.4E-02 | 0 | 2.5E-01 | 0.018 | 0.011 | 0.009 |
| rs35569658 | *RBFOX1* | 16 | 5754433 | C | G | 5,203 | 0.313 | -0.562 | 0.112 | 5.3E-07 | 2,398 | 0.250 | -0.560 | 0.180 | 1.8E-03 | 2,793 | 0.367 | -0.536 | 0.142 | 1.6E-04 | 0 | 4.0E-01 | 0.138 | 0.249 | 0.353 |
| rs148280037 | *SHISA9* | 16 | 12886370 | T | C | 5,203 | 0.984 | 2.081 | 0.412 | 4.5E-07 | 2,398 | 0.978 | 2.358 | 0.527 | 7.5E-06 | 2,793 | 0.989 | 1.638 | 0.674 | 1.5E-02 | 0 | 2.8E-01 | 0.979 | 0.987 | 0.994 |
| rs143565319 | *PIK3C3* | 18 | 40962346 | T | C | 5,203 | 0.989 | 2.669 | 0.497 | 7.8E-08 | 2,398 | 0.983 | 2.656 | 0.583 | 5.3E-06 | 2,793 | 0.995 | 2.111 | 1.001 | 3.5E-02 | 0 | 3.6E-01 | 0.925 | 1.000 | 0.996 |
| **WOMEN** | | | | | | | | | | | | | | | | | | | | | | | | | |
| rs112469617 | *FZD7* | 2 | 201982887 | T | C | 7,471 | 0.009 | -2.977 | 0.594 | 5.5E-07 | 3,237 | 0.014 | -3.047 | 0.744 | 4.3E-05 | 4,220 | 0.006 | -2.895 | 0.984 | 3.3E-03 | 0 | 4.0E-01 | 0.064 | 0.000 | 0.007 |
| rs17385466 | *SOX5* | 12 | 23639004 | T | C | 7,471 | 0.923 | 1.089 | 0.209 | 1.8E-07 | 3,237 | 0.899 | 1.286 | 0.283 | 5.6E-06 | 4,220 | 0.941 | 0.869 | 0.308 | 4.8E-03 | 0 | 2.4E-01 | 0.995 | 0.886 | 0.916 |
| **ALL** | | | | | | | | | | | | | | | | | | | | | | | | | |
| rs77993329 | *ACTRT2* | 1 | 3001535 | A | G | 12,673 | 0.02 | -1.457 | 0.288 | 4.1E-07 | 5,634 | 0.031 | -1.610 | 0.371 | 1.4E-05 | 7,013 | 0.011 | -1.523 | 0.528 | 3.9E-03 | 0 | 4.0E-01 | 0.091 | 0.000 | 0.019 |
| rs138819965 | *LINC00299* | 2 | 8304328 | A | G | 12,674 | 0.994 | 2.706 | 0.525 | 2.5E-07 | 5,635 | 0.989 | 2.781 | 0.624 | 8.2E-06 | 7,013 | 0.998 | 1.670 | 1.230 | 1.7E-01 | 0 | 2.9E-01 | 0.953 | 1.000 | 0.999 |
| rs77319470 | *ADAMTS3* | 4 | 72638360 | T | G | 12,674 | 0.371 | 0.406 | 0.082 | 8.6E-07 | 5,635 | 0.409 | 0.434 | 0.130 | 8.3E-04 | 7,013 | 0.341 | 0.329 | 0.115 | 4.1E-03 | 0 | 3.3E-01 | 0.580 | 0.391 | 0.359 |
| rs921999 | *FGF5* | 4 | 80296998 | A | C | 12,674 | 0.965 | -1.104 | 0.222 | 6.7E-07 | 5,635 | 0.939 | -0.850 | 0.275 | 2.0E-03 | 7,013 | 0.986 | -1.160 | 0.456 | 1.1E-02 | 0 | 3.4E-01 | 0.791 | 1.000 | 0.980 |
| rs112404395 | *AP3B1* | 5 | 77964563 | C | G | 12,674 | 0.991 | -2.22 | 0.412 | 7.1E-08 | 5,635 | 0.989 | -2.344 | 0.602 | 9.9E-05 | 7,013 | 0.992 | -2.164 | 0.627 | 5.6E-04 | 0 | 3.9E-01 | 0.999 | 0.979 | 0.993 |
| rs77264633 | *CCDC171* | 9 | 15823920 | A | G | 12,674 | 0.962 | -1.118 | 0.205 | **4.6E-08** | 5,635 | 0.955 | -1.132 | 0.302 | 1.8E-04 | 7,013 | 0.967 | -1.242 | 0.304 | 4.4E-05 | 0 | 3.9E-01 | 0.995 | 0.931 | 0.963 |
| rs184067184 | *PHF21A* | 11 | 46189280 | A | C | 12,674 | 0.006 | -3.612 | 0.702 | 2.7E-07 | 5,635 | 0.008 | -3.988 | 0.833 | 1.7E-06 | 7,013 | 0.004 | -2.871 | 1.720 | 9.5E-02 | 0 | 3.4E-01 | 0.000 | 0.000 | 0.004 |
| rs3168072 | *FADS2* | 11 | 61864038 | A | T | 12,674 | 0.725 | 0.514 | 0.102 | 5.3E-07 | 5,635 | 0.885 | 0.417 | 0.207 | 4.4E-02 | 7,013 | 0.597 | 0.455 | 0.120 | 1.6E-04 | 0 | 3.9E-01 | 0.990 | 0.967 | 0.627 |
| rs60260780 | *WSB2* | 12 | 118041899 | T | C | 12,674 | 0.013 | 1.78 | 0.35 | 3.7E-07 | 5,635 | 0.023 | 2.070 | 0.427 | 1.2E-06 | 7,013 | 0.005 | 0.620 | 0.759 | 4.1E-01 | 63.92 | 1.0E-01 | 0.117 | 0.001 | 0.006 |

**Supplementary Table 7**. Summary of association results in SOL subgroup analyses for suggestively significant loci (P<1E-6) associated with HIPadjBMI. EAF-estimated allele frequency, CHR- chromosome, POS-position (build GRCh38), SE- standard error, ISQ- I squared heterogeneity. EAF for reference population obtained from 1000 Genomes Project Phase 3.

| | | | | | | Combined | | | | | Caribbean | | | | | Mainland | | | | | | | EAF | | |
|---|---|---|---|---|---|---|---|---|---|---|---|---|---|---|---|---|---|---|---|---|---|---|---|---|---|
| dbSNPID | Nearest Gene | CHR | POS | Effect Allele | Other Allele | N | EAF | BETA | SE | PVAL | N | EAF | BETA | SE | PVAL | N | EAF | BETA | SE | PVAL | ISQ | $P_{diff}$ | AFR | EUR | AMR |
| **MEN** | | | | | | | | | | | | | | | | | | | | | | | | | |
| rs114865909 | *NUF2* | 1 | 163589134 | T | C | 5,197 | 0.994 | -0.012 | 0.002 | 8.6E-07 | 2,394 | 0.989 | -0.012 | 0.003 | 1.5E-05 | 2,791 | 0.998 | -0.010 | 0.006 | 9.8E-02 | 7.27 | 3.7E-01 | 0.950 | 0.999 | 0.999 |
| rs149681500 | *ANO10* | 3 | 43396572 | T | C | 5,197 | 0.007 | -0.012 | 0.002 | 5.3E-08 | 2,394 | 0.013 | -0.014 | 0.003 | 9.6E-08 | 2,791 | 0.002 | -0.002 | 0.005 | 7.7E-01 | 80.06 | **4.4E-02** | 0.051 | 0.000 | 0.004 |
| rs3915213 | *LOC101927346* | 3 | 74024720 | T | C | 5,197 | 0.779 | -0.002 | 0.000 | 5.6E-07 | 2,394 | 0.712 | -0.002 | 0.001 | 2.0E-04 | 2,791 | 0.836 | -0.002 | 0.001 | 1.5E-03 | 0.00 | 3.8E-01 | 0.665 | 0.698 | 0.831 |
| rs12677587 | *LOC101929066* | 8 | 18118812 | C | G | 5,197 | 0.012 | -0.009 | 0.002 | 9.6E-07 | 2,394 | 0.020 | -0.009 | 0.002 | 2.7E-05 | 2,791 | 0.005 | -0.008 | 0.004 | 3.2E-02 | 0.00 | 3.7E-01 | 0.058 | 0.002 | 0.010 |
| rs56405004 | *MINPP1* | 10 | 87512407 | T | C | 5,197 | 0.018 | 0.008 | 0.001 | 1.7E-07 | 2,394 | 0.027 | 0.007 | 0.002 | 5.1E-05 | 2,791 | 0.010 | 0.008 | 0.002 | 7.5E-04 | 0.00 | 3.9E-01 | 0.108 | 0.002 | 0.010 |
| rs968849 | *LOC102724589* | 10 | 114997972 | A | G | 5,197 | 0.195 | -0.002 | 0.000 | 8.2E-07 | 2,394 | 0.237 | -0.002 | 0.001 | 1.2E-03 | 2,791 | 0.158 | -0.002 | 0.001 | 4.0E-04 | 0.00 | 3.9E-01 | 0.161 | 0.313 | 0.154 |
| rs76469489 | *SLC7A10* | 19 | 33249049 | C | G | 5,197 | 0.013 | -0.009 | 0.002 | 9.6E-07 | 2,394 | 0.015 | -0.009 | 0.002 | 2.7E-04 | 2,791 | 0.010 | -0.008 | 0.003 | 1.5E-03 | 0.00 | 3.9E-01 | 0.002 | 0.035 | 0.016 |
| rs7063750 | *VCX* | X | 7724974 | T | G | 5,186 | 0.039 | 0.004 | 0.001 | 2.7E-07 | 2,390 | 0.068 | 0.004 | 0.001 | 2.1E-05 | 2,784 | 0.014 | 0.004 | 0.002 | 4.6E-03 | 0.00 | 3.7E-01 | 0.628 | 0.086 | 0.120 |
| rs112519383 | *MID1* | X | 10446834 | A | G | 5,186 | 0.988 | -0.007 | 0.001 | 7.4E-08 | 2,390 | 0.980 | -0.007 | 0.001 | 2.5E-06 | 2,784 | 0.995 | -0.006 | 0.003 | 3.0E-02 | 0.00 | 3.5E-01 | 0.883 | 0.999 | 0.990 |
| **WOMEN** | | | | | | | | | | | | | | | | | | | | | | | | | |
| rs72978809 | *LPPR4* | 1 | 99260939 | A | G | 7,462 | 0.994 | 0.011 | 0.002 | 4.4E-07 | 3,232 | 0.990 | 0.012 | 0.003 | 1.4E-05 | 4,216 | 0.997 | 0.010 | 0.004 | 1.5E-02 | 0.00 | 3.8E-01 | 0.956 | 1.000 | 0.996 |
| rs12478843 | *HEATR5B* | 2 | 37080089 | A | G | 7,462 | 0.248 | -0.002 | 0.000 | 8.2E-08 | 3,232 | 0.154 | -0.002 | 0.001 | 6.0E-03 | 4,216 | 0.320 | -0.002 | 0.000 | 6.5E-06 | 1.68 | 3.9E-01 | 0.023 | 0.137 | 0.268 |
| rs115331260 | *LOC105376941* | 3 | 5864170 | A | G | 7,462 | 0.985 | 0.008 | 0.002 | 8.8E-07 | 3,232 | 0.982 | 0.009 | 0.002 | 4.0E-05 | 4,216 | 0.987 | 0.005 | 0.002 | 1.9E-02 | 0.00 | 1.9E-01 | 0.003 | 0.037 | 0.004 |
| rs7662640 | *LOC105374566* | 4 | 31765535 | T | C | 7,462 | 0.012 | 0.008 | 0.002 | 6.3E-07 | 3,232 | 0.012 | 0.009 | 0.003 | 5.8E-04 | 4,216 | 0.012 | 0.007 | 0.002 | 4.4E-04 | 0.00 | 3.7E-01 | 0.011 | 0.024 | 0.012 |
| rs6814739 | *LINC01094* | 4 | 78654647 | T | C | 7,462 | 0.569 | -0.002 | 0.000 | 7.2E-07 | 3,232 | 0.594 | -0.002 | 0.001 | 2.8E-04 | 4,216 | 0.550 | -0.002 | 0.000 | 1.7E-04 | 0.00 | 3.5E-01 | 0.154 | 0.766 | 0.566 |
| rs11099588 | *COQ2* | 4 | 83280326 | T | C | 7,462 | 0.057 | 0.004 | 0.001 | 2.4E-07 | 3,232 | 0.090 | 0.005 | 0.001 | 1.9E-06 | 4,216 | 0.032 | 0.003 | 0.001 | 3.8E-02 | 64.14 | 2.0E-01 | 0.281 | 0.033 | 0.036 |
| rs6860625 | *NREP* | 5 | 111667124 | A | G | 7,462 | 0.813 | -0.002 | 0.000 | 6.4E-07 | 3,232 | 0.789 | -0.004 | 0.001 | 1.3E-07 | 4,216 | 0.831 | -0.001 | 0.001 | 6.4E-02 | 28.03 | **9.5E-03** | 0.593 | 0.869 | 0.839 |
| rs77186623 | *LOC105375745* | 8 | 125608952 | A | C | 7,461 | 0.022 | -0.006 | 0.001 | 1.7E-07 | 3,231 | 0.041 | -0.008 | 0.001 | **3.1E-08** | 4,216 | 0.008 | -0.001 | 0.003 | 5.7E-01 | 55.31 | **4.2E-02** | 0.115 | 0.000 | 0.017 |
| rs10818474 | *MEGF9* | 9 | 120727686 | T | C | 7,462 | 0.809 | -0.002 | 0.000 | 5.1E-07 | 3,232 | 0.743 | -0.002 | 0.001 | 3.1E-03 | 4,216 | 0.860 | -0.003 | 0.001 | 4.2E-06 | 0.00 | 1.6E-01 | 0.548 | 0.764 | 0.817 |
| rs28692724* | *IRF2BPL* | 14 | 77027445 | T | C | 7,462 | 0.425 | 0.002 | 0.000 | 7.3E-07 | 3,232 | 0.377 | 0.002 | 0.001 | 4.0E-05 | 4,216 | 0.462 | 0.002 | 0.001 | 8.7E-04 | 0.00 | 2.3E-01 | 0.160 | 0.384 | 0.310 |
| rs6092086 | *LOC105372676* | 20 | 55193568 | A | G | 7,462 | 0.922 | -0.003 | 0.001 | 3.4E-07 | 3,232 | 0.871 | -0.003 | 0.001 | 4.0E-05 | 4,216 | 0.962 | -0.003 | 0.001 | 6.4E-03 | 0.00 | 4.0E-01 | 0.562 | 0.989 | 0.944 |
| rs9631175 | *TAF4* | 20 | 62072648 | A | C | 7,462 | 0.793 | 0.002 | 0.000 | 6.0E-08 | 3,232 | 0.834 | 0.002 | 0.001 | 1.3E-02 | 4,216 | 0.762 | 0.003 | 0.001 | 2.1E-06 | 38.00 | 2.8E-01 | 0.701 | 0.888 | 0.785 |
| **ALL** | | | | | | | | | | | | | | | | | | | | | | | | | |
| rs144655586 | *CLSPN* | 1 | 35725381 | T | C | 12,658 | 0.012 | 0.007 | 0.001 | 1.7E-07 | 5,625 | 0.012 | 0.010 | 0.002 | 9.1E-06 | 7,007 | 0.011 | 0.006 | 0.002 | 3.5E-03 | 49.55 | 1.5E-01 | 0.001 | 0.014 | 0.006 |
| rs712900 | *LPPR4* | 1 | 99261313 | T | C | 12,659 | 0.014 | -0.006 | 0.001 | 3.4E-07 | 5,626 | 0.022 | -0.005 | 0.001 | 1.3E-04 | 7,007 | 0.007 | -0.006 | 0.002 | 3.2E-03 | 0.00 | 3.8E-01 | 0.098 | 0.001 | 0.007 |
| rs115546449 | *TMEM63A* | 1 | 225882871 | A | G | 12,659 | 0.026 | -0.004 | 0.001 | 3.0E-07 | 5,626 | 0.047 | -0.004 | 0.001 | 1.4E-04 | 7,007 | 0.010 | -0.005 | 0.002 | 4.0E-03 | 0.00 | 3.2E-01 | 0.136 | 0.001 | 0.014 |
| rs145815581 | *ANO10* | 3 | 43466242 | A | G | 12,659 | 0.007 | -0.008 | 0.002 | 1.8E-07 | 5,626 | 0.013 | -0.009 | 0.002 | 2.3E-07 | 7,007 | 0.003 | -0.002 | 0.003 | 5.9E-01 | 76.32 | **4.8E-02** | 0.051 | 0.000 | 0.004 |
| rs72886347 | *FHIT* | 3 | 60196461 | T | C | 12,659 | 0.993 | 0.008 | 0.002 | 7.2E-07 | 5,626 | 0.989 | 0.010 | 0.002 | 4.5E-07 | 7,007 | 0.997 | 0.006 | 0.003 | 6.0E-02 | 15.13 | 2.2E-01 | 0.940 | 1.000 | 0.994 |
| rs17136358 | *EIF2AK1* | 7 | 6040031 | T | C | 12,659 | 0.963 | -0.003 | 0.001 | 2.8E-07 | 5,626 | 0.951 | -0.003 | 0.001 | 2.6E-03 | 7,007 | 0.972 | -0.004 | 0.001 | 4.9E-04 | 0.00 | 3.4E-01 | 0.958 | 0.953 | 0.967 |
| rs117683919 | *LOC105375440* | 7 | 106634111 | A | G | 12,659 | 0.006 | -0.009 | 0.002 | 1.4E-07 | 5,626 | 0.009 | -0.011 | 0.002 | 4.0E-07 | 7,007 | 0.004 | -0.007 | 0.003 | 1.5E-02 | 0.00 | 1.9E-01 | 0.005 | 0.014 | 0.003 |
| rs143542634 | *PATL1* | 11 | 59666070 | A | G | 12,659 | 0.006 | 0.009 | 0.002 | 4.3E-07 | 5,626 | 0.006 | 0.011 | 0.003 | 6.7E-05 | 7,007 | 0.007 | 0.008 | 0.002 | 2.1E-04 | 0.00 | 2.9E-01 | 0.001 | 0.006 | 0.004 |

**Supplementary Table 8.** Local ancestry results for all suggestively significant loci across all traits.

| rsID | Nearest Gene | CHR | POS (GRCh38) | Sex | Effect Allele | Other Allele | EAF | | |
|---|---|---|---|---|---|---|---|---|---|
| | | | | | | | African | Native American | European |
| **WHR** | | | | | | | | | |
| rs13301996 | *CDK5RAP2* | 9 | 120570806 | All | T | G | 0.9029 | 0.8890 | 0.7414 |
| rs115981023 | *TAOK3* | 12 | 118313300 | All | G | A | 0.9431 | 1.0000 | 0.9996 |
| rs185566196 | *KIAA0391* | 14 | 35152593 | All | NA | NA | NA | NA | NA |
| rs116612483 | *CDH4* | 20 | 60751325 | All | G | A | 0.9631 | 1.0000 | 0.9999 |
| rs12032174 | *RYR2* | 1 | 237692868 | Men | C | T | 0.4060 | 0.3051 | 0.3693 |
| rs10475310 | *PLEKHG4B* | 5 | 23024 | Men | NA | NA | NA | NA | NA |
| rs16977373 | *RIT2* | 18 | 43189620 | Men | A | C | 0.7718 | 1.0000 | 0.9911 |
| rs721424 | *CFAP61* | 20 | 20358666 | Men | G | T | 0.8124 | 0.5079 | 0.4454 |
| rs148213302 | *STS* | 23 | 7416464 | Men | NA | NA | NA | NA | NA |
| rs77377042 | *MARCKSL1* | 1 | 32343334 | Women | C | T | 0.8421 | 1.0000 | 0.9994 |
| rs75120960 | *EPHA5* | 4 | 66367753 | Women | A | G | 0.8609 | 1.0000 | 0.9994 |
| rs16922424 | *FAM110B* | 8 | 57618850 | Women | T | C | 0.8762 | 1.0000 | 0.9994 |
| rs79478137 | *SLC22A18AS* | 11 | 2891739 | Women | C | T | 0.9232 | 0.9989 | 0.9946 |
| rs113818604 | *NTM* | 11 | 131960980 | Women | G | A | 1.0000 | 1.0000 | 0.9809 |
| rs115981023 | *TAOK3* | 12 | 118313300 | Women | G | A | 0.9431 | 1.0000 | 0.9996 |
| rs146900844 | *ZNF207* | 17 | 32378136 | Women | G | A | 1.0000 | 1.0000 | 0.9942 |
| rs61305557 | *C19orf67* | 19 | 14081134 | Women | NA | NA | NA | NA | NA |
| **WC** | | | | | | | | | |
| rs77993329 | *ACTRT2* | 1 | 3001535 | All | G | A | 0.9126 | 0.9996 | 0.9904 |
| rs138819965 | *LINC00299* | 2 | 8304328 | All | A | G | 0.9598 | 1.0000 | 1.0000 |
| rs77319470 | *ADAMTS3* | 4 | 72638360 | All | G | T | 0.5050 | 0.7252 | 0.6110 |
| rs921999 | *FGF5* | 4 | 80296998 | All | A | C | 0.7737 | 0.9998 | 0.9941 |
| rs112404395 | *AP3B1* | 5 | 77964563 | All | C | G | 1.0000 | 1.0000 | 0.9835 |
| rs77264633 | *CCDC171* | 9 | 15823920 | All | A | G | 1.0000 | 0.9985 | 0.9318 |
| rs184067184 | *PHF21A* | 11 | 46189280 | All | C | A | 1.0000 | 0.9946 | 1.0000 |
| rs3168072 | *FADS2* | 11 | 61864038 | All | A | T | 0.9884 | 0.1949 | 0.9755 |
| rs60260780 | *WSB2* | 12 | 118041899 | All | C | T | 0.9112 | 0.9999 | 0.9998 |
| rs74346221 | *GABRD* | 1 | 2029024 | Men | NA | NA | NA | NA | NA |
| rs72693785 | *ATP1A1* | 1 | 116334965 | Men | C | T | 1.0000 | 1.0000 | 0.9943 |
| rs11583298 | *ESRRG* | 1 | 217171261 | Men | T | C | 1.0000 | 0.5320 | 0.9309 |
| rs76842062 | *MAP4K4* | 2 | 101847192 | Men | T | G | 0.9775 | 1.0000 | 0.9997 |
| rs141365360 | *LOC102723448* | 3 | 21826 | Men | G | A | 0.9512 | 0.9999 | 0.9996 |
| rs6809759 | *PROK2* | 3 | 71937742 | Men | G | A | 0.6679 | 0.6342 | 0.3908 |
| rs76941364 | *COBL* | 7 | 52064843 | Men | A | G | 0.9472 | 1.0000 | 0.9971 |
| rs139139519 | *SULF1* | 8 | 69625608 | Men | G | A | 0.9803 | 1.0000 | 0.9820 |
| rs35569658 | *RBFOX1* | 16 | 5754433 | Men | G | C | 0.8982 | 0.5073 | 0.7564 |
| rs148280037 | *SHISA9* | 16 | 12886370 | Men | T | C | 0.9780 | 0.9997 | 0.9825 |
| rs143565319 | *PIK3C3* | 18 | 40962346 | Men | T | C | 0.9216 | 1.0000 | 0.9998 |
| rs112469617 | *FZD7* | 2 | 201982887 | Women | C | T | 0.9471 | 1.0000 | 1.0000 |
| rs17385466 | *SOX5* | 12 | 23639004 | Women | T | C | 0.9990 | 0.9998 | 0.8576 |
| **HIP** | | | | | | | | | |
| rs144655586 | *CLSPN* | 1 | 35725381 | All | C | T | 0.9976 | 1.0000 | 0.9914 |
| rs712900 | *LPPR4* | 1 | 99261313 | All | C | T | 0.8977 | 1.0000 | 0.9998 |
| rs115546449 | *TMEM63A* | 1 | 225882871 | All | G | A | 0.8206 | 1.0000 | 0.9983 |
| rs145815581 | *ANO10* | 3 | 43466242 | All | G | A | 0.9531 | 1.0000 | 1.0000 |

| | | | | | | | | | |
|---|---|---|---|---|---|---|---|---|---|
| rs72886347 | *FHIT* | 3 | 60184786 | All | T | C | 0.9608 | 1.0000 | 0.9998 |
| rs17136358 | *EIF2AK1* | 7 | 6040031 | All | T | C | 0.9423 | 0.9995 | 0.9486 |
| rs117683919 | *LOC105375440* | 7 | 106634111 | All | G | A | 0.9944 | 1.0000 | 0.9909 |
| rs143542634 | *PATL1* | 11 | 59666070 | All | G | A | 1.0000 | 1.0000 | 0.9905 |
| rs114865909 | *NUF2* | 1 | 163589134 | Men | T | C | 0.9550 | 1.0000 | 0.9996 |
| rs149681500 | *ANO10* | 3 | 43396572 | Men | C | T | 0.9518 | 1.0000 | 1.0000 |
| rs3915213 | *LOC101927346* | 3 | 74024720 | Men | T | C | 0.6733 | 0.9898 | 0.6870 |
| rs12677587 | *LOC101929066* | 8 | 18118812 | Men | G | C | 0.9245 | 0.9997 | 0.9972 |
| rs56405004 | *MINPP1* | 10 | 87512407 | Men | C | T | 0.8830 | 0.9997 | 0.9953 |
| rs968849 | *LOC102724589* | 10 | 114997972 | Men | G | A | 0.8553 | 0.9888 | 0.6894 |
| rs76469489 | *SLC7A10* | 19 | 33249049 | Men | G | C | 1.0000 | 0.9991 | 0.9775 |
| rs7063750 | *VCX* | 23 | 7724974 | Men | NA | NA | NA | NA | NA |
| rs112519383 | *MID1* | 23 | 10446834 | Men | NA | NA | NA | NA | NA |
| rs72978809 | *LPPR4* | 1 | 99260939 | Women | A | G | 0.9607 | 1.0000 | 0.9998 |
| rs12478843 | *HEATR5B* | 2 | 37080089 | Women | G | A | 0.9869 | 0.4895 | 0.8482 |
| rs115331260 | *LOC105376941* | 3 | 5864170 | Women | A | G | 1.0000 | 1.0000 | 0.9843 |
| rs7662640 | *LOC105374566* | 4 | 31765535 | Women | C | T | 0.9930 | 0.9924 | 0.9864 |
| rs6814739 | *LINC01094* | 4 | 78654647 | Women | C | T | 0.8707 | 0.6437 | 0.1988 |
| rs11099588 | *COQ2* | 4 | 83280326 | Women | T | C | 0.2944 | 0.0006 | 0.0308 |
| rs6860625 | *NREP* | 5 | 111667124 | Women | A | G | 0.5807 | 0.8582 | 0.8526 |
| rs77186623 | *LOC105375745* | 8 | 125608952 | Women | C | A | 0.8587 | 1.0000 | 0.9965 |
| rs10818474 | *MEGF9* | 9 | 120727686 | Women | C | T | 0.4459 | 0.0034 | 0.2387 |
| rs28692724 | *IRF2BPL* | 14 | 77027445 | Women | C | T | 0.8296 | 0.4506 | 0.6436 |
| rs6092086 | *LOC105372676* | 20 | 55193568 | Women | A | G | 0.5493 | 0.9995 | 0.9692 |
| rs9631175 | *TAF4* | 20 | 62072648 | Women | A | C | 0.7289 | 0.6444 | 0.8909 |

**Supplemental Table 9. Top variants for each generalized locus for waist-to-hip ratio adjusted for body mass index in SOL across known loci found in GIANT GWAS**

| | | | | | | GIANT estimates | | | | HCHS/SOL estimates | | | | | Meta-Analysis estimates | | | |
|---|---|---|---|---|---|---|---|---|---|---|---|---|---|---|---|---|---|---|
| **dbSNPID** | **Region** | **Chr** | **Position (build GRCh38)** | **Effect Allele** | **Other Allele** | **Effect Allele Frequency** | **Beta** | **SE** | **P-value** | **Effect Allele Frequency** | **Beta** | **SE** | **P-value** | **Generalization r-value** | **Beta** | **SE** | **P-value** | **Locus #** |
| **WOMEN** | | | | | | | | | | | | | | | | | | |
| rs7543720 | *LOC101929147* | 1 | 119163343 | A | G | 0.325 | -0.031 | 0.006 | 1.00E-07 | 0.239 | -0.077 | 0.019 | 5.98E-05 | 0.035 | -0.035 | 0.006 | 3.48E-10 | **1** |
| rs2605082 | *LOC102723886* | 1 | 219446196 | G | A | 0.305 | 0.031 | 0.005 | 4.90E-11 | 0.291 | 0.081 | 0.018 | 5.97E-06 | 0.007 | 0.034 | 0.005 | 5.08E-14 | **2** |
| rs3769869 | *COBLL1* | 2 | 164686689 | A | G | 0.783 | 0.051 | 0.006 | 1.80E-20 | 0.769 | 0.072 | 0.019 | 1.71E-04 | 0.038 | 0.053 | 0.005 | 2.56E-23 | **3** |
| rs9311910 | *ADAMTS9-AS2* | 3 | 64721593 | G | A | 0.550 | 0.043 | 0.005 | 6.30E-21 | 0.417 | 0.051 | 0.017 | 2.42E-03 | 0.047 | 0.044 | 0.004 | 9.49E-23 | **4** |
| rs1294410 | *LOC101928004* | 6 | 6738519 | C | T | 0.625 | 0.037 | 0.005 | 6.00E-16 | 0.529 | 0.052 | 0.017 | 1.66E-03 | 0.038 | 0.038 | 0.004 | 8.58E-18 | **5** |
| rs2800703 | *LOC105377989* | 6 | 127105653 | G | T | 0.333 | 0.048 | 0.005 | 2.20E-22 | 0.419 | 0.054 | 0.017 | 1.36E-03 | 0.038 | 0.048 | 0.005 | 6.99E-25 | **6** |
| **ALL** | | | | | | | | | | | | | | | | | | |
| rs1106529 | *TBX15* | 1 | 118988874 | G | A | 0.275 | -0.035 | 0.004 | 1.80E-19 | 0.315 | -0.066 | 0.014 | 1.59E-06 | 0.002 | -0.037 | 0.004 | 2.64E-23 | **1** |
| rs2605095 | *LOC107985272* | 1 | 219468108 | T | A | 0.317 | -0.039 | 0.005 | 8.00E-18 | 0.442 | -0.038 | 0.013 | 2.47E-03 | 0.027 | -0.039 | 0.004 | 4.31E-20 | **2** |
| rs40271 | *intergenic* | 5 | 56500492 | C | T | 0.225 | 0.022 | 0.004 | 3.80E-09 | 0.362 | 0.038 | 0.013 | 3.51E-03 | 0.034 | 0.023 | 0.004 | 1.86E-10 | **7** |
| rs1294410 | *LOC101928004* | 6 | 6738519 | C | T | 0.625 | 0.032 | 0.004 | 7.70E-20 | 0.524 | 0.048 | 0.013 | 1.67E-04 | 0.010 | 0.033 | 0.003 | 9.69E-23 | **5** |
| rs7192 | *HLA-DRA* | 6 | 32443869 | G | T | 0.583 | 0.017 | 0.004 | 5.40E-07 | 0.670 | 0.038 | 0.013 | 4.62E-03 | 0.041 | 0.018 | 0.003 | 6.04E-08 | **8** |
| rs1358980 | *LOC105375070* | 6 | 43796814 | T | C | 0.450 | 0.038 | 0.004 | 1.30E-26 | 0.480 | 0.049 | 0.013 | 8.84E-05 | 0.008 | 0.039 | 0.003 | 1.15E-30 | **9** |
| rs11766345 | *LAMB1* | 7 | 107971205 | G | T | 0.875 | 0.037 | 0.007 | 7.10E-07 | 0.871 | 0.056 | 0.019 | 3.05E-03 | 0.031 | 0.040 | 0.007 | 9.64E-09 | **10** |
| rs894737 | *HOXC4* | 12 | 54024359 | C | A | 0.342 | 0.029 | 0.004 | 2.40E-11 | 0.323 | 0.048 | 0.013 | 3.44E-04 | 0.012 | 0.031 | 0.004 | 1.62E-13 | **11** |
| rs4930723 | *CCDC92* | 12 | 123939053 | G | C | 0.617 | 0.030 | 0.005 | 1.70E-11 | 0.702 | 0.048 | 0.014 | 4.74E-04 | 0.015 | 0.032 | 0.004 | 1.16E-13 | **12** |

Abbreviations: Chr, chromosome; SE, standard error

**Supplemental Table 10. Top variants for each generalized locus for waist circumference adjusted for body mass index in SOL across known loci found in GIANT GWAS**

| | | | | | | GIANT estimates | | | | HCHS/SOL estimates | | | | | Meta-Analysis estimates | | | |
|---|---|---|---|---|---|---|---|---|---|---|---|---|---|---|---|---|---|---|
| dbSNPID | Region | Chr | Position (build GRCh38) | Effect Allele | Other Allele | Effect Allele Frequency | Beta | SE | P-value | Effect Allele Frequency | Beta | SE | P-value | Generalization r-value | Beta | SE | P-value | Locus # |
| **MEN** | | | | | | | | | | | | | | | | | | |
| rs1409156 | *TBX15* | 1 | 118750684 | G | A | 0.342 | 0.026 | 0.005 | 1.30E-07 | 0.325 | 0.076 | 0.021 | 3.95E-04 | 0.034 | 0.028 | 0.005 | 2.50E-09 | 1 |
| rs3791679 | *EFEMP1* | 2 | 55723261 | A | G | 0.725 | 0.053 | 0.006 | 6.20E-22 | 0.796 | 0.090 | 0.024 | 2.40E-04 | 0.034 | 0.055 | 0.005 | 1.83E-24 | 2 |
| rs1147225 | *NPR3* | 5 | 32823835 | C | T | 0.642 | 0.027 | 0.005 | 1.50E-08 | 0.576 | 0.060 | 0.020 | 2.43E-03 | 0.050 | 0.029 | 0.005 | 6.48E-10 | 3 |
| rs459193 | *C5orf67* | 5 | 56546681 | A | G | 0.217 | 0.027 | 0.005 | 4.50E-07 | 0.277 | 0.078 | 0.022 | 3.88E-04 | 0.034 | 0.030 | 0.005 | 7.28E-09 | 4 |
| rs9358927 | *LOC10192874* | 6 | 26438243 | T | G | 0.583 | 0.027 | 0.005 | 1.50E-08 | 0.591 | 0.067 | 0.020 | 7.79E-04 | 0.034 | 0.029 | 0.005 | 4.05E-10 | 5 |
| rs754133 | *HOXC4* | 12 | 52311403 | A | G | 0.333 | 0.032 | 0.005 | 8.80E-11 | 0.326 | 0.067 | 0.021 | 1.36E-03 | 0.037 | 0.034 | 0.005 | 1.33E-12 | 6 |
| **WOMEN** | | | | | | | | | | | | | | | | | | |
| rs6691985 | *CROCC* | 1 | 16963416 | T | C | 0.225 | -0.024 | 0.005 | 2.80E-07 | 0.339 | -0.074 | 0.018 | 3.54E-05 | 0.025 | -0.027 | 0.005 | 2.15E-09 | 7 |
| rs1106529 | *TBX15* | 1 | 118790397 | G | A | 0.275 | -0.039 | 0.005 | 1.00E-14 | 0.312 | -0.073 | 0.018 | 4.11E-05 | 0.025 | -0.041 | 0.005 | 6.62E-18 | 1 |
| rs2605082 | *LOC10272388* | 1 | 217512819 | G | A | 0.305 | 0.023 | 0.005 | 7.00E-07 | 0.292 | 0.070 | 0.018 | 8.32E-05 | 0.027 | 0.026 | 0.004 | 5.78E-09 | 8 |
| rs3769869 | *COBLL1* | 2 | 164394935 | A | G | 0.783 | 0.035 | 0.005 | 2.90E-11 | 0.769 | 0.063 | 0.019 | 9.55E-04 | 0.027 | 0.037 | 0.005 | 4.34E-13 | 9 |
| rs10049090 | *LINC02029* | 3 | 158562607 | G | A | 0.600 | 0.029 | 0.006 | 1.00E-07 | 0.531 | 0.049 | 0.017 | 3.20E-03 | 0.027 | 0.031 | 0.005 | 3.00E-09 | 10 |
| rs7705502 | *CPEB4* | 5 | 173826418 | A | G | 0.292 | 0.029 | 0.005 | 4.40E-10 | 0.167 | 0.064 | 0.022 | 3.46E-03 | 0.027 | 0.030 | 0.005 | 1.29E-11 | 11 |
| rs10748827 | *SUFU* | 10 | 102598569 | G | T | 0.700 | 0.022 | 0.004 | 5.80E-07 | 0.652 | 0.050 | 0.017 | 3.55E-03 | 0.027 | 0.024 | 0.004 | 2.57E-08 | 12 |
| rs11853983 | *ADAMTSL3* | 15 | 81961473 | A | G | 0.725 | 0.031 | 0.006 | 2.30E-08 | 0.782 | 0.064 | 0.020 | 1.28E-03 | 0.027 | 0.033 | 0.005 | 5.60E-10 | 13 |
| rs757608 | Intergenic | 17 | 58774698 | A | G | 0.300 | 0.026 | 0.005 | 1.40E-08 | 0.364 | 0.060 | 0.017 | 4.20E-04 | 0.027 | 0.028 | 0.004 | 1.83E-10 | 14 |
| **ALL** | | | | | | | | | | | | | | | | | | |
| rs6691985 | *CROCC* | 1 | 16963416 | T | C | 0.225 | -0.026 | 0.004 | 3.90E-13 | 0.337 | -0.060 | 0.014 | 1.71E-05 | 0.008 | -0.028 | 0.003 | 6.92E-16 | 7 |
| rs1106529 | *TBX15* | 1 | 118790397 | G | A | 0.275 | -0.035 | 0.004 | 1.00E-20 | 0.315 | -0.066 | 0.014 | 1.58E-06 | 0.002 | -0.037 | 0.004 | 3.08E-24 | 1 |
| rs17369648 | Intergenic | 2 | 55708944 | C | T | 0.7 | 0.029 | 0.005 | 4.20E-09 | 0.788 | 0.048 | 0.016 | 1.97E-03 | 0.042 | 0.031 | 0.005 | 4.91E-11 | 2 |
| rs10049090 | *LINC02029* | 3 | 158562607 | G | A | 0.6 | 0.030 | 0.005 | 2.60E-11 | 0.532 | 0.043 | 0.013 | 8.81E-04 | 0.033 | 0.031 | 0.004 | 1.46E-13 | 10 |
| rs7697556 | Intergenic | 4 | 72868460 | C | T | 0.4917 | -0.016 | 0.003 | 9.00E-07 | 0.557 | -0.057 | 0.013 | 8.30E-06 | 0.026 | -0.019 | 0.003 | 6.08E-09 | 15 |
| rs455660 | *C5orf67* | 5 | 56556818 | T | C | 0.1333 | 0.024 | 0.004 | 3.70E-09 | 0.202 | 0.049 | 0.016 | 2.46E-03 | 0.046 | 0.026 | 0.004 | 3.07E-10 | 4 |
| rs754133 | *HOXC4* | 12 | 52311403 | A | G | 0.3333 | 0.029 | 0.004 | 4.00E-17 | 0.322 | 0.040 | 0.013 | 2.84E-03 | 0.048 | 0.030 | 0.003 | 1.79E-18 | 6 |
| rs7214743 | Intergenic | 17 | 58775473 | A | G | 0.2917 | 0.024 | 0.005 | 1.30E-07 | 0.363 | 0.051 | 0.013 | 1.12E-04 | 0.022 | 0.027 | 0.004 | 5.79E-10 | 14 |

Abbreviations: Chr, chromosome; SE, standard error

**Supplemental Table 11. Top variants for each generalized locus for hip ratio adjusted for body mass index in SOL across known loci found in GIANT GWAS**

| | | | | | | GIANT estimates | | | | HCHS/SOL estimates | | | | | Meta-Analysis estimates | | | |
|---|---|---|---|---|---|---|---|---|---|---|---|---|---|---|---|---|---|---|
| dbSNPID | Region | Chr | Position (build GRCh38) | Effect Allele | Other Allele | Effect Allele Frequency | Beta | SE | P-value | Effect Allele Frequency | Beta | SE | P-value | Generalization r-value | Beta | SE | P-value | Locus # |
| **WOMEN** | | | | | | | | | | | | | | | | | | |
| rs2820443 | Regulatory Region | 1 | 217646790 | C | T | 0.300 | 0.062 | 0.005 | 4.30E-35 | 0.402 | 0.060 | 0.017 | 3.175E-04 | 0.047 | 0.062 | 0.005 | 3.88E-38 | 1 |
| rs1346786 | *EFEMP1* | 2 | 55734702 | C | T | 0.667 | 0.030 | 0.006 | 2.60E-07 | 0.691 | 0.081 | 0.018 | 6.000E-06 | 0.047 | 0.035 | 0.006 | 4.12E-10 | 2 |
| rs17819328 | *PPARG* | 3 | 12422843 | G | T | 0.450 | -0.028 | 0.005 | 1.60E-09 | 0.375 | -0.062 | 0.017 | 3.839E-04 | 0.047 | -0.030 | 0.005 | 2.44E-11 | 3 |
| **ALL** | | | | | | | | | | | | | | | | | | |
| rs761422 | *MFAP2* | 1 | 16975285 | A | G | 0.440 | -0.028 | 0.005 | 1.40E-08 | 0.523 | -0.037 | 0.013 | 4.161E-03 | 0.040 | -0.029 | 0.005 | 3.90E-10 | 4 |
| rs11205303 | *MTMR11* | 1 | 149934520 | C | T | 0.358 | 0.042 | 0.004 | 6.40E-26 | 0.271 | 0.042 | 0.014 | 2.792E-03 | 0.037 | 0.042 | 0.004 | 9.52E-28 | 5 |
| rs2494196 | Intergenic | 1 | 217655862 | A | C | 0.250 | 0.051 | 0.005 | 7.60E-23 | 0.422 | 0.049 | 0.013 | 1.533E-04 | 0.018 | 0.051 | 0.005 | 1.12E-26 | 1 |
| rs848607 | *CRIM1* | 2 | 36396553 | A | G | 0.367 | -0.018 | 0.004 | 9.60E-07 | 0.455 | -0.043 | 0.013 | 7.675E-04 | 0.021 | -0.020 | 0.004 | 1.99E-08 | 6 |
| rs1346786 | *EFEMP1* | 2 | 55734702 | C | T | 0.667 | 0.030 | 0.005 | 5.40E-10 | 0.690 | 0.063 | 0.014 | 4.500E-06 | 0.013 | 0.034 | 0.005 | 1.24E-13 | 2 |
| rs9872031 | *PPARG* | 3 | 12429962 | A | G | 0.442 | -0.024 | 0.005 | 1.70E-07 | 0.377 | -0.047 | 0.013 | 3.930E-04 | 0.018 | -0.026 | 0.004 | 1.14E-09 | 3 |
| rs2811469 | *LXND1* | 3 | 131045304 | G | A | 0.708 | -0.037 | 0.006 | 5.30E-11 | 0.788 | -0.057 | 0.016 | 2.904E-04 | 0.018 | -0.039 | 0.005 | 2.15E-13 | 7 |
| rs11936911 | *LCORL* | 4 | 17556143 | G | A | 0.158 | -0.036 | 0.006 | 1.40E-08 | 0.209 | -0.049 | 0.016 | 1.688E-03 | 0.032 | -0.038 | 0.006 | 9.57E-11 | 8 |
| rs7670141 | | 4 | 72863854 | A | G | 0.517 | -0.026 | 0.005 | 7.10E-09 | 0.604 | -0.056 | 0.013 | 1.740E-05 | 0.015 | -0.029 | 0.004 | 6.67E-12 | 9 |
| rs1443537 | | 4 | 81448750 | A | C | 0.242 | 0.025 | 0.004 | 4.70E-10 | 0.332 | 0.042 | 0.013 | 1.539E-03 | 0.030 | 0.026 | 0.004 | 5.37E-12 | 10 |
| rs3775380 | *FAM13A* | 4 | 89037680 | G | A | 0.492 | -0.018 | 0.004 | 2.50E-07 | 0.491 | -0.057 | 0.013 | 6.970E-06 | 0.018 | -0.021 | 0.003 | 7.72E-10 | 11 |
| rs2035742 | *HHIP* | 4 | 144875668 | A | G | 0.958 | 0.044 | 0.009 | 3.40E-07 | 0.957 | 0.115 | 0.031 | 2.055E-04 | 0.018 | 0.049 | 0.008 | 4.32E-09 | 12 |
| rs1147225 | *NPR3* | 5 | 32823835 | C | T | 0.642 | 0.018 | 0.004 | 3.50E-07 | 0.577 | 0.037 | 0.013 | 4.326E-03 | 0.041 | 0.019 | 0.003 | 1.13E-08 | 13 |
| rs1294438 | *LOC101928004* | 6 | 6696825 | C | T | 0.667 | -0.027 | 0.004 | 4.70E-12 | 0.512 | -0.044 | 0.013 | 5.751E-04 | 0.018 | -0.028 | 0.004 | 2.37E-14 | 14 |
| rs4371882 | *BMP6* | 6 | 7737935 | A | G | 0.833 | -0.027 | 0.005 | 3.30E-09 | 0.809 | -0.055 | 0.016 | 6.764E-04 | 0.020 | -0.029 | 0.004 | 2.20E-11 | 15 |
| rs1265083 | *CCHCR1* | 6 | 31251549 | T | G | 0.008 | -0.073 | 0.015 | 9.20E-07 | 0.042 | -0.144 | 0.032 | 5.100E-06 | 0.018 | -0.086 | 0.014 | 2.13E-10 | 16 |
| rs4140531 | *ADGRG6* | 6 | 142405651 | G | A | 0.933 | 0.040 | 0.008 | 3.50E-07 | 0.910 | 0.065 | 0.022 | 3.673E-03 | 0.040 | 0.043 | 0.007 | 6.63E-09 | 17 |
| rs7783400 | *GNA12* | 7 | 2821326 | A | C | 0.275 | -0.033 | 0.005 | 8.30E-12 | 0.280 | -0.041 | 0.014 | 3.479E-03 | 0.040 | -0.034 | 0.005 | 9.34E-14 | 18 |
| rs42377 | *CDK6* | 7 | 92452294 | A | G | 0.371 | 0.037 | 0.006 | 1.40E-11 | 0.297 | 0.038 | 0.014 | 6.272E-03 | 0.050 | 0.037 | 0.005 | 3.84E-13 | 19 |
| rs10990768 | *SLC35D2* | 9 | 95413062 | C | T | 0.858 | -0.031 | 0.006 | 5.50E-07 | 0.924 | -0.096 | 0.024 | 5.700E-05 | 0.018 | -0.035 | 0.006 | 4.84E-09 | 20 |
| rs7082470 | *JMJD1C* | 10 | 63187272 | G | A | 0.517 | -0.018 | 0.004 | 4.90E-07 | 0.679 | -0.043 | 0.014 | 1.742E-03 | 0.032 | -0.020 | 0.003 | 8.66E-09 | 21 |
| rs10882717 | *ENTPD1-AS1* | 10 | 96081177 | G | C | 0.692 | 0.023 | 0.005 | 6.10E-07 | 0.699 | 0.041 | 0.014 | 3.133E-03 | 0.038 | 0.025 | 0.004 | 1.38E-08 | 22 |
| rs2701540 | *MACROD1* | 11 | 63858211 | T | A | 0.042 | -0.048 | 0.010 | 9.90E-07 | 0.086 | -0.073 | 0.023 | 1.169E-03 | 0.028 | -0.052 | 0.009 | 7.24E-09 | 23 |
| rs2160077 | Regulatory Region | 14 | 91031819 | G | A | 0.608 | 0.017 | 0.004 | 8.90E-07 | 0.677 | 0.038 | 0.014 | 4.611E-03 | 0.042 | 0.018 | 0.003 | 6.26E-08 | 24 |
| rs8029016 | *ADAMTSL3* | 15 | 82085127 | C | T | 0.475 | 0.036 | 0.003 | 1.80E-25 | 0.664 | 0.052 | 0.014 | 2.424E-04 | 0.018 | 0.037 | 0.003 | 7.14E-29 | 25 |
| rs2281727 | *SMG6* | 17 | 2161401 | G | A | 0.333 | -0.018 | 0.004 | 5.30E-07 | 0.361 | -0.057 | 0.013 | 1.830E-05 | 0.018 | -0.021 | 0.003 | 2.78E-09 | 26 |
| rs143499 | *SMG6* | 17 | 2175780 | T | C | 0.325 | -0.018 | 0.004 | 9.10E-07 | 0.365 | -0.057 | 0.013 | 1.750E-05 | 0.018 | -0.021 | 0.003 | 2.75E-09 | 27 |
| rs4800451 | *CABLES1* | 18 | 21390842 | T | C | 0.683 | 0.030 | 0.004 | 9.70E-14 | 0.634 | 0.043 | 0.013 | 1.178E-03 | 0.028 | 0.031 | 0.004 | 1.88E-15 | 28 |
| rs158676 | *CDK5RAP1* | 20 | 32850250 | G | A | 0.283 | -0.022 | 0.004 | 2.10E-09 | 0.246 | -0.042 | 0.015 | 4.904E-03 | 0.044 | -0.023 | 0.003 | 4.04E-11 | 29 |
| rs2425060 | *UQCC1* | 20 | 34790312 | T | C | 0.675 | -0.034 | 0.005 | 5.50E-13 | 0.710 | -0.042 | 0.014 | 2.659E-03 | 0.037 | -0.035 | 0.004 | 5.54E-15 | 30 |

Abbreviations: Chr, chromosome; SE, standard error

**ST12. P-values from testing genetic scores for each of the traits and populations**

| Strata | p-value<1e-8 | 1e-8<p-value<1e-7 | 1e-7<p-value<1e-6 |
|---|---|---|---|
| **WHRadjBMI** | | | |
| MEN | 9.00E-03 | 4.55E-02 | 3.02E-01 |
| WOMEN | 5.95E-06 | 3.85E-03 | 8.36E-03 |
| ALL | 2.00E-06 | 5.78E-03 | 2.87E-01 |
| **HIPadjBMI** | | | |
| MEN | 1.46E-10 | 2.14E-03 | 9.17E-02 |
| WOMEN | 1.89E-11 | 2.36E-05 | 2.95E-02 |
| ALL | 3.81E-07 | 6.46E-03 | 4.45E-05 |
| **WCadjBMI** | | | |
| MEN | 1.17E-02 | 8.20E-05 | 4.33E-06 |
| WOMEN | 2.74E-04 | 5.46E-01 | 1.62E-02 |
| ALL | 9.41E-12 | 1.24E-03 | 9.68E-03 |

**Supplementary Table 13.** GWAS lookups of replication variants from current study in Phenoscanner. Only significant SNP-phenotype associations shown ($P<0.05/7631=6.55x10^{-5}$).

| Discovery Trait | dbSNPID | CHR | POS (GRCh38) | Effect Allele | Other Allele | GWAS Trait | PMID/ Source | Ancestry | Beta | SE | P | N | N (cases) | N (controls) |
|---|---|---|---|---|---|---|---|---|---|---|---|---|---|---|
| WHRadjBMI | rs79478137 | 11 | 2891739 | C | T | Cause of death: multisystem degeneration | UKBB | European | -0.024 | 0.004 | **4.8E-11** | 7637 | 8 | 7629 |
| WHRadjBMI | rs79478137 | 11 | 2891739 | C | T | Cause of death: tongue, unspecified | UKBB | European | -0.023 | 0.004 | **4.8E-08** | 7637 | 11 | 7626 |
| WHRadjBMI | rs79478137 | 11 | 2891739 | C | T | Home area population density: postcode not linkable | UKBB | European | -0.001 | 0.000 | **1.4E-07** | 333997 | 12 | 333985 |
| WCadjBMI | rs3168072 | 11 | 61864038 | A | T | Cause of death: other specified respiratory disorders | UKBB | European | -0.006 | 0.001 | **1.7E-06** | 7637 | 4 | 7633 |

**Supplementary Table 14. eQTL** lookups of replicated variants from current study in Phenoscanner. Only significant SNP-Gene expression associations shown ($P<0.05/88=5.68x10^{-4}$ ).

| Discovery Trait | dbSNPID | CHR | POS (GRCh38) | Nearest Gene | Effect Allele | Other Allele | PMID/ Source | Ancestry | Tissue | Gene | Beta | SE | P | N | Dataset |
|---|---|---|---|---|---|---|---|---|---|---|---|---|---|---|---|
| WHRadjBMI | rs13301996 | 9 | 120570806 | *CDK5RAP2* | G | T | 25954001 | Unspecified | Adipose subcutaneous | *MEGF9* | 0.293 | 0.055 | **1.9E-07** | 385 | GTEx-V7_eQTL_EUR_2017 |
| WHRadjBMI | rs13301996 | 9 | 120570806 | *CDK5RAP2* | G | T | 25954001 | Unspecified | Adipose subcutaneous | *AHCYP2* | 0.351 | 0.084 | **4.1E-05** | 385 | GTEx-V7_eQTL_EUR_2017 |
| WHRadjBMI | rs13301996 | 9 | 120570806 | *CDK5RAP2* | G | T | 25954001 | Unspecified | Adipose subcutaneous | *MEGF9* | 0.254 | 0.067 | **1.7E-04** | 298 | GTEx-V6p_eQTL_EUR_2016 |
| WHRadjBMI | rs13301996 | 9 | 120570806 | *CDK5RAP2* | G | T | 25954001 | Unspecified | Adipose subcutaneous | *PSMD5-AS1* | 0.289 | 0.082 | **5.0E-04** | 385 | GTEx-V7_eQTL_EUR_2017 |
| WHRadjBMI | rs13301996 | 9 | 120570806 | *CDK5RAP2* | G | T | 25954001 | Unspecified | Artery aorta | *PSMD5-AS1* | 0.363 | 0.103 | **5.1E-04** | 267 | GTEx-V7_eQTL_EUR_2017 |
| WHRadjBMI | rs13301996 | 9 | 120570806 | *CDK5RAP2* | G | T | 25954001 | Unspecified | Artery coronary | - | 0.593 | 0.150 | **1.5E-04** | 118 | GTEx-V6p_eQTL_EUR_2016 |
| WHRadjBMI | rs13301996 | 9 | 120570806 | *CDK5RAP2* | G | T | 25954001 | Unspecified | Artery tibial | *PSMD5-AS1* | 0.458 | 0.087 | **2.5E-07** | 388 | GTEx-V7_eQTL_EUR_2017 |
| WHRadjBMI | rs13301996 | 9 | 120570806 | *CDK5RAP2* | G | T | 25954001 | Unspecified | Artery tibial | *PSMD5-AS1* | 0.436 | 0.102 | **3.0E-05** | 285 | GTEx-V6p_eQTL_EUR_2016 |
| WHRadjBMI | rs13301996 | 9 | 120570806 | *CDK5RAP2* | G | T | 25954001 | Unspecified | Brain cerebellum | *PSMD5-AS1* | 0.564 | 0.152 | **3.9E-04** | 103 | GTEx-V6p_eQTL_EUR_2016 |
| WHRadjBMI | rs13301996 | 9 | 120570806 | *CDK5RAP2* | G | T | 25954001 | Unspecified | Colon sigmoid | *PSMD5-AS1* | 0.404 | 0.110 | **3.1E-04** | 203 | GTEx-V7_eQTL_EUR_2017 |
| WHRadjBMI | rs13301996 | 9 | 120570806 | *CDK5RAP2* | G | T | 25954001 | Unspecified | Colon sigmoid | *PSMD5-AS1* | 0.511 | 0.142 | **5.1E-04** | 124 | GTEx-V6p_eQTL_EUR_2016 |
| WHRadjBMI | rs13301996 | 9 | 120570806 | *CDK5RAP2* | G | T | 25954001 | Unspecified | Colon sigmoid | *AHCYP2* | 0.346 | 0.098 | **5.5E-04** | 203 | GTEx-V7_eQTL_EUR_2017 |
| WHRadjBMI | rs13301996 | 9 | 120570806 | *CDK5RAP2* | G | T | 25954001 | Unspecified | Esophagus muscularis | *PSMD5-AS1* | 0.407 | 0.091 | **1.2E-05** | 335 | GTEx-V7_eQTL_EUR_2017 |
| WHRadjBMI | rs13301996 | 9 | 120570806 | *CDK5RAP2* | G | T | 25954001 | Unspecified | Esophagus muscularis | *PSMD5-AS1* | 0.451 | 0.112 | **7.8E-05** | 218 | GTEx-V6p_eQTL_EUR_2016 |
| WHRadjBMI | rs13301996 | 9 | 120570806 | *CDK5RAP2* | G | T | 25954001 | Unspecified | Esophagus muscularis | *AHCYP2* | 0.309 | 0.088 | **5.3E-04** | 335 | GTEx-V7_eQTL_EUR_2017 |
| WHRadjBMI | rs13301996 | 9 | 120570806 | *CDK5RAP2* | G | T | 25954001 | Unspecified | Liver | *PSMD5* | 0.354 | 0.093 | **2.2E-04** | 153 | GTEx-V7_eQTL_EUR_2017 |
| WHRadjBMI | rs13301996 | 9 | 120570806 | *CDK5RAP2* | G | T | 25954001 | Unspecified | Liver | *PSMD5-AS1* | 0.361 | 0.100 | **4.6E-04** | 153 | GTEx-V7_eQTL_EUR_2017 |
| WHRadjBMI | rs13301996 | 9 | 120570806 | *CDK5RAP2* | G | T | 25954001 | Unspecified | Lung | *PSMD5-AS1* | 0.349 | 0.076 | **6.7E-06** | 383 | GTEx-V7_eQTL_EUR_2017 |
| WHRadjBMI | rs13301996 | 9 | 120570806 | *CDK5RAP2* | G | T | 25954001 | Unspecified | Muscle skeletal | *PSMD5* | 0.261 | 0.056 | **4.1E-06** | 491 | GTEx-V7_eQTL_EUR_2017 |
| WHRadjBMI | rs13301996 | 9 | 120570806 | *CDK5RAP2* | G | T | 25954001 | Unspecified | Muscle skeletal | *PSMD5* | 0.327 | 0.070 | **4.5E-06** | 361 | GTEx-V6p_eQTL_EUR_2016 |
| WHRadjBMI | rs13301996 | 9 | 120570806 | *CDK5RAP2* | G | T | 25954001 | Unspecified | Muscle skeletal | *PSMD5-AS1* | 0.302 | 0.072 | **3.7E-05** | 491 | GTEx-V7_eQTL_EUR_2017 |
| WHRadjBMI | rs13301996 | 9 | 120570806 | *CDK5RAP2* | G | T | 25954001 | Unspecified | Muscle skeletal | *PSMD5-AS1* | 0.320 | 0.090 | **4.2E-04** | 361 | GTEx-V6p_eQTL_EUR_2016 |
| WHRadjBMI | rs13301996 | 9 | 120570806 | *CDK5RAP2* | G | T | 25954001 | Unspecified | Nerve tibial | *PSMD5-AS1* | 0.461 | 0.091 | **6.6E-07** | 361 | GTEx-V7_eQTL_EUR_2017 |
| WHRadjBMI | rs13301996 | 9 | 120570806 | *CDK5RAP2* | G | T | 25954001 | Unspecified | Nerve tibial | *PSMD5-AS1* | 0.428 | 0.105 | **6.2E-05** | 256 | GTEx-V6p_eQTL_EUR_2016 |
| WHRadjBMI | rs13301996 | 9 | 120570806 | *CDK5RAP2* | G | T | 25954001 | Unspecified | Skin not sun exposed suprapubic | *PSMD5-AS1* | 0.333 | 0.083 | **8.3E-05** | 335 | GTEx-V7_eQTL_EUR_2017 |
| WHRadjBMI | rs13301996 | 9 | 120570806 | *CDK5RAP2* | G | T | 25954001 | Unspecified | Skin sun exposed lower leg | *PSMD5-AS1* | 0.328 | 0.082 | **7.5E-05** | 414 | GTEx-V7_eQTL_EUR_2017 |
| WHRadjBMI | rs13301996 | 9 | 120570806 | *CDK5RAP2* | G | T | 25954001 | Unspecified | Skin sun exposed lower leg | *PSMD5-AS1* | 0.387 | 0.096 | **8.0E-05** | 302 | GTEx-V6p_eQTL_EUR_2016 |
| WHRadjBMI | rs13301996 | 9 | 120570806 | *CDK5RAP2* | G | T | 25954001 | Unspecified | Skin sun exposed lower leg | *MEGF9* | 0.123 | 0.033 | **2.8E-04** | 302 | GTEx-V6p_eQTL_EUR_2016 |
| WHRadjBMI | rs13301996 | 9 | 120570806 | *CDK5RAP2* | G | T | 27863251 | European | T cells | *MEGF9* | -0.437 | 0.126 | **5.5E-04** | 169 | BLUEPRINT_eQTL_EUR_2016 |
| WHRadjBMI | rs13301996 | 9 | 120570806 | *CDK5RAP2* | G | T | 25954001 | Unspecified | Testis | *CDK5RAP2* | 0.251 | 0.060 | **4.8E-05** | 225 | GTEx-V7_eQTL_EUR_2017 |
| WHRadjBMI | rs13301996 | 9 | 120570806 | *CDK5RAP2* | G | T | 25954001 | Unspecified | Thyroid | *PSMD5-AS1* | 0.395 | 0.082 | **2.4E-06** | 399 | GTEx-V7_eQTL_EUR_2017 |
| WHRadjBMI | rs13301996 | 9 | 120570806 | *CDK5RAP2* | G | T | 25954001 | Unspecified | Thyroid | *PSMD5* | 0.194 | 0.051 | **1.8E-04** | 399 | GTEx-V7_eQTL_EUR_2017 |
| WHRadjBMI | rs13301996 | 9 | 120570806 | *CDK5RAP2* | G | T | 25954001 | Unspecified | Thyroid | *PSMD5-AS1* | 0.365 | 0.101 | **3.6E-04** | 278 | GTEx-V6p_eQTL_EUR_2016 |
| WHRadjBMI | rs13301996 | 9 | 120570806 | *CDK5RAP2* | G | T | eQTLGen | European | Whole blood | *MEGF9* | NA | NA | **1.8E-149** | 30523 | eQTLGen_eQTL_EUR_2018 |
| WHRadjBMI | rs13301996 | 9 | 120570806 | *CDK5RAP2* | G | T | eQTLGen | European | Whole blood | *RP11-27I1.2* | NA | NA | **8.5E-107** | 16488 | eQTLGen_eQTL_EUR_2018 |
| WHRadjBMI | rs13301996 | 9 | 120570806 | *CDK5RAP2* | G | T | eQTLGen | European | Whole blood | *CDK5RAP2* | NA | NA | **1.7E-61** | 30737 | eQTLGen_eQTL_EUR_2018 |
| WHRadjBMI | rs13301996 | 9 | 120570806 | *CDK5RAP2* | G | T | eQTLGen | European | Whole blood | *PSMD5* | NA | NA | **1.6E-10** | 30737 | eQTLGen_eQTL_EUR_2018 |
| WHRadjBMI | rs13301996 | 9 | 120570806 | *CDK5RAP2* | G | T | eQTLGen | European | Whole blood | *PHF19* | NA | NA | **1.0E-08** | 29988 | eQTLGen_eQTL_EUR_2018 |
| WHRadjBMI | rs13301996 | 9 | 120570806 | *CDK5RAP2* | G | T | eQTLGen | European | Whole blood | *GGTA1P* | NA | NA | **1.5E-07** | 24529 | eQTLGen_eQTL_EUR_2018 |
| WHRadjBMI | rs13301996 | 9 | 120570806 | *CDK5RAP2* | G | T | 28122634 | Mixed | Whole blood | *MEGF9* | -0.018 | 0.003 | **2.6E-07** | 5257 | Joehanes-R_eQTL_EUR_2017 |
| WHRadjBMI | rs13301996 | 9 | 120570806 | *CDK5RAP2* | G | T | 28122634 | Mixed | Whole blood | *CYP4Z1;CYP4Z2P* | -0.030 | 0.007 | **7.7E-06** | 5257 | Joehanes-R_eQTL_EUR_2017 |
| WHRadjBMI | rs13301996 | 9 | 120570806 | *CDK5RAP2* | G | T | 28122634 | Mixed | Whole blood | *RAB14* | 0.015 | 0.003 | **7.8E-06** | 5257 | Joehanes-R_eQTL_EUR_2017 |
| WHRadjBMI | rs13301996 | 9 | 120570806 | *CDK5RAP2* | G | T | eQTLGen | European | Whole blood | *RAB14* | NA | NA | **8.0E-06** | 29988 | eQTLGen_eQTL_EUR_2018 |

| | | | | | | | | | | | | | | | |
|---|---|---|---|---|---|---|---|---|---|---|---|---|---|---|---|
| WHRadjBMI | rs13301996 | 9 | 120570806 | *CDK5RAP2* | G | T | 28122634 | Mixed | Whole blood | *BPIFB3* | 0.013 | 0.003 | **3.9E-05** | 5257 | Joehanes-R_eQTL_EUR_2017 |
| WHRadjBMI | rs13301996 | 9 | 120570806 | *CDK5RAP2* | G | T | 28122634 | Mixed | Whole blood | *PSMD5* | 0.018 | 0.004 | **4.4E-05** | 5257 | Joehanes-R_eQTL_EUR_2017 |
| WHRadjBMI | rs13301996 | 9 | 120570806 | *CDK5RAP2* | G | T | 28122634 | Mixed | Whole blood | *RBM3;SLC38A5* | 0.013 | 0.003 | **5.1E-05** | 5257 | Joehanes-R_eQTL_EUR_2017 |
| WHRadjBMI | rs13301996 | 9 | 120570806 | *CDK5RAP2* | G | T | 28122634 | Mixed | Whole blood | *SIN3A* | 0.010 | 0.002 | **7.7E-05** | 5257 | Joehanes-R_eQTL_EUR_2017 |
| WHRadjBMI | rs13301996 | 9 | 120570806 | *CDK5RAP2* | G | T | 25954001 | Unspecified | Whole blood | *PSMD5-AS1* | 0.270 | 0.068 | **9.4E-05** | 369 | GTEx-V7_eQTL_EUR_2017 |
| WHRadjBMI | rs13301996 | 9 | 120570806 | *CDK5RAP2* | G | T | 28122634 | Mixed | Whole blood | *MRPL14* | -0.019 | 0.005 | **9.9E-05** | 5257 | Joehanes-R_eQTL_EUR_2017 |
| HIPadjBMI | rs28692724 | 14 | 77027445 | *IRF2BPL* | C | T | eQTLGen | European | Whole blood | *IRF2BPL* | NA | NA | **8.6E-23** | 6544 | eQTLGen_eQTL_EUR_2018 |
| HIPadjBMI | rs28692724 | 14 | 77027445 | *IRF2BPL* | C | T | 27918533 | European | Whole blood | *RP11-7F17.5* | NA | NA | **2.1E-10** | 2116 | BIOSQTL_eQTL_EUR_2017 |
| HIPadjBMI | rs28692724 | 14 | 77027445 | *IRF2BPL* | C | T | 27918533 | European | Whole blood | *RP11-7F17.5;RP11-7F17.3* | NA | NA | **1.4E-09** | 2116 | BIOSQTL_eQTL_EUR_2017 |
| HIPadjBMI | rs28692724 | 14 | 77027445 | *IRF2BPL* | C | T | 27918533 | European | Whole blood | *RP11-7F17.3* | NA | NA | **1.6E-09** | 2116 | BIOSQTL_eQTL_EUR_2017 |
| HIPadjBMI | rs28692724 | 14 | 77027445 | *IRF2BPL* | C | T | 27918533 | European | Whole blood | *RP11-7F17.7* | NA | NA | **2.2E-08** | 2116 | BIOSQTL_eQTL_EUR_2017 |
| HIPadjBMI | rs28692724 | 14 | 77027445 | *IRF2BPL* | C | T | 27918533 | European | Whole blood | *IRF2BPL* | NA | NA | **2.3E-07** | 2116 | BIOSQTL_eQTL_EUR_2017 |
| HIPadjBMI | rs28692724 | 14 | 77027445 | *IRF2BPL* | C | T | 25954001 | Unspecified | Colon sigmoid | *ANGEL1* | 0.447 | 0.119 | **2.7E-04** | 124 | GTEx-V6p_eQTL_EUR_2016 |
| WCadjBMI | rs3168072 | 11 | 61864038 | *FADS2* | A | T | 25954001 | Unspecified | Artery tibial | *INCENP* | -0.623 | 0.129 | **2.5E-06** | 285 | GTEx-V6p_eQTL_EUR_2016 |
| WCadjBMI | rs3168072 | 11 | 61864038 | *FADS2* | A | T | 25954001 | Unspecified | Artery tibial | *INCENP* | -0.358 | 0.101 | **4.6E-04** | 388 | GTEx-V7_eQTL_EUR_2017 |
| WCadjBMI | rs3168072 | 11 | 61864038 | *FADS2* | A | T | 25954001 | Unspecified | Brain caudate basal ganglia | *LRRN4CL* | 1.155 | 0.325 | **5.4E-04** | 144 | GTEx-V7_eQTL_EUR_2017 |
| WCadjBMI | rs3168072 | 11 | 61864038 | *FADS2* | A | T | 25954001 | Unspecified | Cells EBV-transformed lymphocytes | *SDHAF2* | 0.865 | 0.229 | **2.8E-04** | 117 | GTEx-V7_eQTL_EUR_2017 |
| WCadjBMI | rs3168072 | 11 | 61864038 | *FADS2* | A | T | 25954001 | Unspecified | Nerve tibial | *SDHAF2* | 0.551 | 0.155 | **4.6E-04** | 256 | GTEx-V6p_eQTL_EUR_2016 |
| WCadjBMI | rs3168072 | 11 | 61864038 | *FADS2* | A | T | 25954001 | Unspecified | Pancreas | *UBXN1* | 0.411 | 0.110 | **2.5E-04** | 220 | GTEx-V7_eQTL_EUR_2017 |
| WCadjBMI | rs3168072 | 11 | 61864038 | *FADS2* | A | T | 25954001 | Unspecified | Skin not sun exposed suprapubic | *TAF6L* | 0.368 | 0.098 | **2.1E-04** | 335 | GTEx-V7_eQTL_EUR_2017 |
| WCadjBMI | rs3168072 | 11 | 61864038 | *FADS2* | A | T | 25954001 | Unspecified | Skin sun exposed lower leg | *TMEM258* | -0.409 | 0.117 | **5.6E-04** | 302 | GTEx-V6p_eQTL_EUR_2016 |
| WCadjBMI | rs3168072 | 11 | 61864038 | *FADS2* | A | T | 25954001 | Unspecified | Small intestine terminal ileum | *SDHAF2* | 0.813 | 0.210 | **1.9E-04** | 122 | GTEx-V7_eQTL_EUR_2017 |
| WCadjBMI | rs3168072 | 11 | 61864038 | *FADS2* | A | T | eQTLGen | European | Whole blood | *TMEM258* | NA | NA | **5.8E-11** | 25068 | eQTLGen_eQTL_EUR_2018 |
| WCadjBMI | rs3168072 | 11 | 61864038 | *FADS2* | A | T | 28122634 | Mixed | Whole blood | *C11orf10* | -0.053 | 0.009 | **1.3E-09** | 5257 | Joehanes-R_eQTL_EUR_2017 |
| WCadjBMI | rs3168072 | 11 | 61864038 | *FADS2* | A | T | 28122634 | Mixed | Whole blood | *XG;XGPY2* | 0.049 | 0.012 | **2.9E-05** | 5257 | Joehanes-R_eQTL_EUR_2017 |
| WCadjBMI | rs3168072 | 11 | 61864038 | *FADS2* | A | T | eQTLGen | European | Whole blood | *FTH1* | NA | NA | **3.3E-05** | 30638 | eQTLGen_eQTL_EUR_2018 |
| WCadjBMI | rs3168072 | 11 | 61864038 | *FADS2* | A | T | 28122634 | Mixed | Whole blood | *APOA4* | -0.056 | 0.014 | **4.7E-05** | 5257 | Joehanes-R_eQTL_EUR_2017 |
| WCadjBMI | rs3168072 | 11 | 61864038 | *FADS2* | A | T | eQTLGen | European | Whole blood | *MYRF* | NA | NA | **5.7E-05** | 25068 | eQTLGen_eQTL_EUR_2018 |
| WCadjBMI | rs3168072 | 11 | 61864038 | *FADS2* | A | T | 28122634 | Mixed | Whole blood | *MED21* | -0.046 | 0.011 | **6.0E-05** | 5257 | Joehanes-R_eQTL_EUR_2017 |
| WCadjBMI | rs3168072 | 11 | 61864038 | *FADS2* | A | T | 28122634 | Mixed | Whole blood | *POU4F2* | 0.040 | 0.010 | **6.4E-05** | 5257 | Joehanes-R_eQTL_EUR_2017 |
| WCadjBMI | rs3168072 | 11 | 61864038 | *FADS2* | A | T | eQTLGen | European | Whole blood | *FADS1* | NA | NA | **5.0E-04** | 31062 | eQTLGen_eQTL_EUR_2018 |

**Supplementary Table 15.** mQTL lookups of replicated variants from current study in Phenoscanner. Only significant SNP-metabolite associations shown ($P<0.05/488=P<1.02x10^{-4}$).

| Discovery Trait | dbSNPID | CHR | POS (GRCh38) | Effect Allele | Other Allele | Metabolite | PMID/ Source | Ancestry | Beta | SE | P | N |
|---|---|---|---|---|---|---|---|---|---|---|---|---|
| WCadjBMI | rs3168072 | 11 | 61864038 | A | T | Other polyunsaturated fatty acids than 18:2 | 27005778 | European | 0.235 | 0.034 | **7.4E-12** | 13549 |
| WCadjBMI | rs3168072 | 11 | 61864038 | A | T | CH2 groups in fatty acids | 27005778 | European | -0.169 | 0.032 | **1.3E-07** | 19021 |
| WCadjBMI | rs3168072 | 11 | 61864038 | A | T | Ratio of bis allylic bonds to double bonds in lipids | 27005778 | European | 0.180 | 0.034 | **1.8E-07** | 13524 |
| WCadjBMI | rs3168072 | 11 | 61864038 | A | T | CH2 groups to double bonds ratio | 27005778 | European | -0.171 | 0.034 | **5.7E-07** | 13532 |
| WCadjBMI | rs3168072 | 11 | 61864038 | A | T | Ratio of bis allylic bonds to total fatty acids in lipids | 27005778 | European | 0.171 | 0.035 | **9.3E-07** | 13171 |

**Supplementary Table 16.** Lookups of replicated variants and variants in high LD (R2>0.8) HaploReg v4.1.

| SOL SNP | Trait | CHR | POS(hg38) | $R^2$ | D' | rsID | REF | ALT | AFR | AMR | ASN | EUR | Chromatin States | Chromatin States Imputed |
|---|---|---|---|---|---|---|---|---|---|---|---|---|---|---|
| rs13301996 | WHRadjBMI (Combined) | 9 | 120569993 | 0.84 | 1 | rs13299371 | A | T | 0.13 | 0.19 | 0.09 | 0.17 | E017,6_EnhG;E049,6_EnhG;E068,6_EnhG;E06 | E017,11_TxEnh3;E026,11_TxEnh3; |
| **rs13301996** | WHRadjBMI (Combined) | 9 | 120570806 | 1 | 1 | **rs13301996** | T | G | 0.13 | 0.22 | 0.09 | 0.18 | E017,6_EnhG;E023,7_Enh;E026,7_Enh;E027,7 | E006,11_TxEnh3;E017,11_TxEnh3; |
| rs79478137 | WHRadjBMI (Women) | 11 | 2888469 | 1 | 1 | rs80153297 | C | T | 0.09 | 0.02 | 0.01 | 0.0026 | E001,12_EnhBiv;E003,12_EnhBiv;E005,7_Enh; | E005,16_EnhW1;E006,16_EnhW1; |
| rs79478137 | WHRadjBMI (Women) | 11 | 2888527 | 1 | 1 | rs76505120 | C | T | 0.11 | 0.02 | 0 | 0.0013 | E001,12_EnhBiv;E003,12_EnhBiv;E005,7_Enh; | E005,16_EnhW1;E006,16_EnhW1; |
| **rs79478137** | WHRadjBMI (Women) | 11 | 2891739 | 1 | 1 | **rs79478137** | C | T | 0.1 | 0.02 | 0.01 | 0.01 | E001,7_Enh;E002,7_Enh;E003,7_Enh;E004,7_ | E001,14_EnhA2;E002,17_EnhW2;E |
| rs79478137 | WHRadjBMI (Women) | 11 | 2892257 | 1 | 1 | rs114179983 | A | T | 0.09 | 0.02 | 0 | 0.0013 | E001,1_TssA;E003,7_Enh;E005,7_Enh;E006,2_ | E001,2_PromU;E002,2_PromU;E00 |
| rs79478137 | WHRadjBMI (Women) | 11 | 2894144 | 1 | 1 | rs12282506 | G | A | 0.11 | 0.02 | 0 | 0.0013 | E018,10_TssBiv | E007,19_DNase;E117,19_DNase |
| rs79478137 | WHRadjBMI (Women) | 11 | 2894333 | 0.8 | 1 | rs114826751 | G | C | 0.1 | 0.03 | 0 | 0.004 | E090,7_Enh | |
| rs79478137 | WHRadjBMI (Women) | 11 | 2894922 | 1 | 1 | rs12269740 | T | G | 0.09 | 0.02 | 0 | 0.0013 | E089,12_EnhBiv;E090,7_Enh | |
| rs79478137 | WHRadjBMI (Women) | 11 | 2896281 | 1 | 1 | rs12276467 | G | A | 0.09 | 0.02 | 0 | 0.0013 | E059,6_EnhG | E056,17_EnhW2;E059,17_EnhW2 |
| rs79478137 | WHRadjBMI (Women) | 11 | 2896432 | 1 | 1 | rs12290113 | C | T | 0.09 | 0.02 | 0 | 0.0013 | E056,12_EnhBiv;E059,7_Enh;E089,12_EnhBiv | E056,17_EnhW2;E059,17_EnhW2 |
| rs3168072 | WCadjBMI (Combined) | 11 | 61860488 | 0.92 | 0.96 | rs74771917 | C | T | 0.06 | 0.32 | 0.2 | 0.04 | E027,7_Enh;E056,3_TxFlnk;E057,6_EnhG;E06 | E027,11_TxEnh3;E055,11_TxEnh3; |
| rs3168072 | WCadjBMI (Combined) | 11 | 61862661 | 0.96 | 1 | rs11605884 | T | C | 0.08 | 0.33 | 0.24 | 0.04 | E022,6_EnhG;E055,6_EnhG;E056,3_TxFlnk;E0 | E055,11_TxEnh3;E056,11_TxEnh3; |
| **rs3168072** | WCadjBMI (Combined) | 11 | 61864038 | 1 | 1 | **rs3168072** | A | T | 0.01 | 0.32 | 0.21 | 0.04 | E022,6_EnhG | E056,11_TxEnh3 |
| rs3168072 | WCadjBMI (Combined) | 11 | 61864838 | 0.91 | 0.99 | rs12577276 | A | G | 0.2 | 0.33 | 0.21 | 0.04 | | |
| rs3168072 | WCadjBMI (Combined) | 11 | 61874245 | 0.9 | -0.97 | rs7115739 | T | G | 0.66 | 0.67 | 0.76 | 0.96 | E024,7_Enh;E052,6_EnhG;E055,6_EnhG;E056, | E005,17_EnhW2;E007,17_EnhW2; |
| **rs28692724** | HIPadjBMI (Women) | 14 | 77027445 | 1 | 1 | **rs28692724** | C | T | 0.25 | 0.32 | 0.29 | 0.29 | E001,1_TssA;E003,1_TssA;E004,1_TssA;E005, | E001,3_PromD1;E002,3_PromD1;E |

| Chromatin Marks | DNAse | Proteins | eQTL | Motifs | GENCODE | GENCODE | RefSeq ID | RefSeq Gene Na | dbSNP Fun | Enhancer Histone Marks |
|---|---|---|---|---|---|---|---|---|---|---|
| E007,H3K9ac_Pro;E017 | | . | GTEx2015_ | Foxc1_2;PT | ENSG00000 | CDK5RAP2 | NM_018249 | CDK5RAP2 | Intronic | |
| E006,H3K4me1_Enh;E0 | | . | GTEx2015_ | AhR_1;Maf | ENSG00000 | CDK5RAP2 | NM_018249 | CDK5RAP2 | Intronic | |
| E001,H3K4me1_Enh;E0 | E003;E004; | A549,CTCF, | . | BCL_disc9 | ENSG00000 | SLC22A18A | NM_007105 | SLC22A18AS | Nonsynony | SKIN, CRVX |
| E001,H3K4me1_Enh;E0 | E003;E004; | A549,CTCF, | . | Sin3Ak-20_ | ENSG00000 | SLC22A18A | NM_007105 | SLC22A18AS | Nonsynony | SKIN, CRVX |
| E001,H3K4me1_Enh;E0 | E007;E051; | . | . | . | ENSG00000 | SLC22A18A | NM_007105 | SLC22A18AS | Intronic | IPSC, BLD, GI, CRVX |
| E001,H3K4me1_Enh;E0 | E003;E004; | A549,CTCF, | . | HDAC2_dis | ENSG00000 | SLC22A18A | NM_007105 | SLC22A18AS | Intronic | ESC, ESDR, LNG, IPSC, FAT, ST |
| E006,H3K4me3_Pro;E0 | E007;E093; | . | . | . | ENSG00000 | SLC22A18A | NM_007105 | SLC22A18AS | Intronic | IPSC |
| E007,H3K4me3_Pro;E0 | | . | . | EBF_disc2; | ENSG00000 | SLC22A18A | NM_007105 | SLC22A18AS | Intronic | |
| E006,H3K4me1_Enh;E0 | E032;E034; | . | . | PLAG1 | ENSG00000 | SLC22A18A | NM_007105 | SLC22A18AS | Intronic | |
| E006,H3K4me1_Enh;E0 | E100 | . | . | THAP1_dis | ENSG00000 | SLC22A18A | NM_007105 | SLC22A18AS | Intronic | |
| E006,H3K4me1_Enh;E0 | E097;E100 | . | . | EWSR1-FLI | ENSG00000 | SLC22A18A | NM_007105 | SLC22A18AS | Intronic | |
| E005,H3K4me1_Enh;E0 | E004;E022; | K562,ELF1, | GTEx2015_ | BRCA1_kno | ENSG00000 | FADS2 | NM_004265 | FADS2 | Intronic | BLD |
| E005,H3K4me1_Enh;E0 | | . | . | CAC-bindin | ENSG00000 | FADS2 | NM_004265 | FADS2 | Intronic | |
| E005,H3K4me1_Enh;E0 | | . | GTEx2015_ | GCM;Maf_ | ENSG00000 | FADS2 | NM_004265 | FADS2 | Intronic | |
| E002,H3K9ac_Pro;E005 | | . | . | EWSR1-FLI | ENSG00000 | FADS2 | NM_004265 | FADS2 | Intronic | |
| E001,H3K4me1_Enh;E0 | E059;E081; | HSMM,CTC | . | ZID | ENSG00000 | FADS3 | NM_021727 | FADS3 | Intronic | |
| E001,H3K4me3_Pro;E0 | E004;E006; | Fibrobl,CTC | . | Myf_1;NRS | ENSG00000 | IRF2BPL | NM_024496 | IRF2BPL | Synonymou | ESC, ESDR, LNG, IPSC, FAT, ST |
